# Transmission and cost-effectiveness modelling to estimate the progress towards elimination and future strategy optimisation for *gambiense* human African trypanosomiasis in Uganda

**DOI:** 10.64898/2025.12.23.25342894

**Authors:** Ching-I Huang, Marina Antillon, Ronald E Crump, Samuel A Sutherland, Paul R Bessell, Albert Mugenyi, Richard Selby, Paul E Brown, Brady Hooley, Andrew Hope, Sophie Dunkley, Rob Sunnucks, Steve J Torr, Fabrizio Tediosi, Joseph Ndung’u, Emily H Crowley, Eric Kidega, Charles Wamboga, Kat S Rock

## Abstract

**Background:** *Gambiense* human African trypanosomiasis (gHAT) is a vectorborne disease with hundreds of thousands of people living at risk across Sub-Saharan Africa. Uganda, having reduced gHAT cases reported annually from 948 in 2000 to zero local cases since 2020, is a frontrunner to be verified by the World Health Organization (WHO) as having achieved elimination of transmission (EoT) of gHAT. It is now crucial to quantify the impact of the interventions deployed, quantify whether the last transmission event (LTE) and last case have already occurred, and determine the resources required to sustain the gains.

**Methods:** We employed a suite of mechanistic compartmental gHAT models, fitted to data from seven districts (Adjumani, Amuru, Arua, Koboko, Maracha, Moyo, and Yumbe) in Uganda that reported gHAT cases during 2000–2022, to address these questions. By combining and weighting the evidence from each model variant, we captured the uncertainty of achieving different elimination metrics across Uganda. Additionally, we utilised the dynamic transmission model outputs to perform a health economic analysis, identifying the most cost-effective strategies to avert disease burden.

**Results:** Predictions estimate that Uganda had the LTE in or before 2021 with high certainty (>95%). The model suggests that the local Ugandan case reported in 2019 may have been the last, but that there remains a 1.125% chance that cases could be reported after 2025. The cost-effectiveness analysis recommends that as transmission dynamics remain similarly low across all strategies, the *Stop 2026* strategy, in which there is only passive screening from 2026 and subsequent reactive screening and vector control are only implemented if cases are detected, has the highest probability of being cost-effective and is predicted to have an economic cost of $193K ($161K–$220K) between 2026–2040.

**Conclusions:** To sustain Uganda’s success whilst ensuring efficient resource use, this modelling analysis recommends implementing the *Stop 2026* strategy in all seven analysed districts, which focuses on maintaining strong passive screening post-2026. This recommendation aligns with the current gHAT strategy in Uganda.

## 1 Introduction

Human African trypanosomiasis (HAT) is a deadly vector-borne disease affecting sub-Saharan Africa. The disease is caused by a protozoan parasite, *Trypanosoma brucei gambiense* and *Trypanosoma brucei rhodesiense* for *gambiense* and *rhodesiense* HAT (gHAT and rHAT), respectively, and transmitted by tsetse (*Glossina*). The World Health Organization’s (WHO’s) Global Health Observatory shows an increase in HAT cases during the 1990s, peaking at 37, 991 new cases reported annually in 1998 in 22 countries [1]. The successful implementation of interventions from the late 1990s has meant that only 546 gHAT and 37 rHAT cases were reported globally in 2024 [1]. Uganda is the only country that has reported both forms of HAT in geographically separated regions—gHAT in Northern Uganda and rHAT in central and Southern Uganda [2]. Uganda was validated by the WHO as having eliminated the *gambiense* form of sleeping sickness as a public health problem (EPHP) in 2022 [3]. Among the seven countries that have received the official gHAT EPHP validation from the WHO, several (Benin, Ghana, Togo, Rwanda, and Uganda) have had zero cases for at least 5 consecutive years and appear close to achieving elimination of transmission (EoT) to humans [4]. Uganda provides a unique setting for examining elimination of gHAT; the very large decrease in case reporting from the 1990s (2066 in 1990, to 948 in 2000, 101 in 2010, and zero in 2020 [1]) is a success, but conversely risks from cross-border human migration from higher endemic countries (especially refugees from South Sudan [5, 6]) could add uncertainty to achieving and sustaining EoT.

Historically, there have been three key interventions available in Uganda, active screening (AS) combined with treatment, passive screening (PS) combined with treatment, and vector control (VC). AS is constituted by mobile teams performing screening in the centre of endemic villages. PS refers to medical diagnosis and treatment sought directly in a health facility by a patient with symptoms, which has improved since 2014 since the introduction of rapid diagnostic tests (RDTs). Treatment has evolved substantially since 2000, particularly with the introduction of nifurtimox–eflornithine combination therapy (NECT) for advanced disease in 2009, which replaced melarsoprol, a potentially toxic treatment. Finally, in addition to the medical interventions, VC has been used in Uganda since 2012 to protect people from becoming infected [7]. The reported effectiveness of tsetse control via the “Tiny Target” method in Uganda was found to produce a *>*80% reduction in tsetse density in intervention areas in four districts (larger rivers of Arua, Maracha, Koboko and Yumbe districts in North West Uganda) [7], which in line with the 60–99% tsetse reduction achieved through the Tiny Target approach in other geographies, thus substantially reducing the opportunity for new infections to humans [8–12].

Through a combination of the three interventions described above, Uganda is one of the countries leading the way to EoT among endemic countries in Africa. The goal of achieving EPHP by 2020 and EoT by 2030, first proposed by the WHO at the 2012 London Declaration, was recently reaffirmed in the WHO roadmap for neglected tropical diseases (NTDs) [13, 14]. Subsequently, the indicator for verification of EoT has been specified as five consecutive years of no case reporting whilst maintaining suitable surveillance activities [15]. In April 2022, Uganda was the fourth country to be validated as having reached EPHP [16]. The next stage is understanding when the country should expect verification for EoT.

Whilst much of the success of the Ugandan programme to combat gHAT is clear, there remain outstanding quantitative questions that we seek to address in the present study:

- What was the differential impact of different types of interventions on transmission and case reporting in the last two decades?
- Has Uganda already had its last transmission event? If so, when did it happen?
- What will case reporting look like in the next five to ten years in each of the seven districts?
- Should the districts continue to have no vertical interventions in place or should there be renewed, targeted activities to seek out and remove extant infections? Would the cost of renewed activities be an effective use of resources?
- What are the resource needs for the next decade?

To address these questions, we fitted a suite of mechanistic compartmental gHAT models to the historical data in the seven districts reporting cases of gHAT from 2000 to 2022 in Uganda. To capture uncertainty, we combined the estimates from each model, weighted by the strength of evidence for each model, to create a weighted ensemble estimate for each district that reflects our uncertainty about transmission. We used the underlying dynamics predicted by the ensemble model to estimate when the last transmission event (LTE) was achieved, and the year after which there was no remaining infection (NRI) in each of the seven districts. We then performed projections under five different strategies to investigate the expected pattern of case reporting and the threat of resurgence from the cessation of vertical interventions.

We coupled the dynamic transmission outputs with health economic analysis to determine the strategies with the highest probability of being cost-effective in each of the districts. We also looked at the minimum cost to conduct strategies with a high chance of reaching LTE by 2030 under different levels of cost constraints. Whilst variations of this model have been used to assess gHAT in other countries [17–20], this study is the first time that a transmission model of gHAT has been fitted to longitudinal data from Uganda to assess progress and prospects for elimination.

## 2 Methods

### Study area

Uganda is the only country that has reported both the *gambiense* (gHAT) and *rhodesiense* (rHAT) forms of HAT [2] with cases of the former reported in northern Uganda and cases of the latter reported in central and southern Uganda, but with no overlap in the geographical distributions of gHAT and rHAT in Uganda. The WHO Global Health Observatory shows that more than 97% of the globally reported cases of sleeping sickness in 2023 were *T.b.gambiense* and 3% were *T.b.rhodesiense*. In contrast to the global shares of each disease, there have been more cases of rHAT than gHAT in Uganda in the last decade (134 compared to 21) [1]. This study focuses on the *gambiense* form of HAT in seven former districts in northeast Uganda sharing the border with South Sudan.

A shapefile from the Database of Global Administrative Areas (GADM) which provides county borders was used to create a district-level shapefile which represented the district boundaries for the period 2006–2019 [21]; the boundaries changed after this period, with some districts splitting to create three additional districts (see Figure S1 in Supplement 1). We selected the geographic boundaries that were valid between 2006–2019 because (a) they cover most of our fitted data period (2000–2022) and (b) some of our data are not geolocated and are only available by district name, making mapping to contemporary district boundaries impossible.

Population numbers for each district by year were estimated from the 2014 census [22, 23] with constant annual population growth based on the change in population from 2002 to 2014 (1991 to 2014 for Moyo district) in each district [22, 23] (see Table S1 in Supplement 1).

The gHAT focus covered a large geographical area (around 16,000 km^2^) with a mixture of crop and cash-crop production and different levels of livestock production among the districts. The growing population of around 2.6 million has become increasingly urbanised and has welcomed large numbers of refugees, who were received at reception centres before being designated to a camp with the materials to build a house. Refugees either became settled or returned to South Sudan or DRC after a period, resulting in substantial bidirectional movement through the South Sudanese border.

### Historical interventions

During the period covered by this study, AS, PS, treatment, and VC were implemented to reduce the transmission and disease burden of gHAT in Uganda. Fig 1 shows both important time points in the history of the disease and public health activities on the ground, as well as the inputs of our analysis. During this period most of the activities were carried out by the national sleeping sickness control programme, but Médecins Sans Frontières (MSF) carried out AS up to 2002 and in 2010. The national programme has been supported by various other partners including WHO, the Gates Foundation, LSTM, and FIND.

**Fig. 1:**
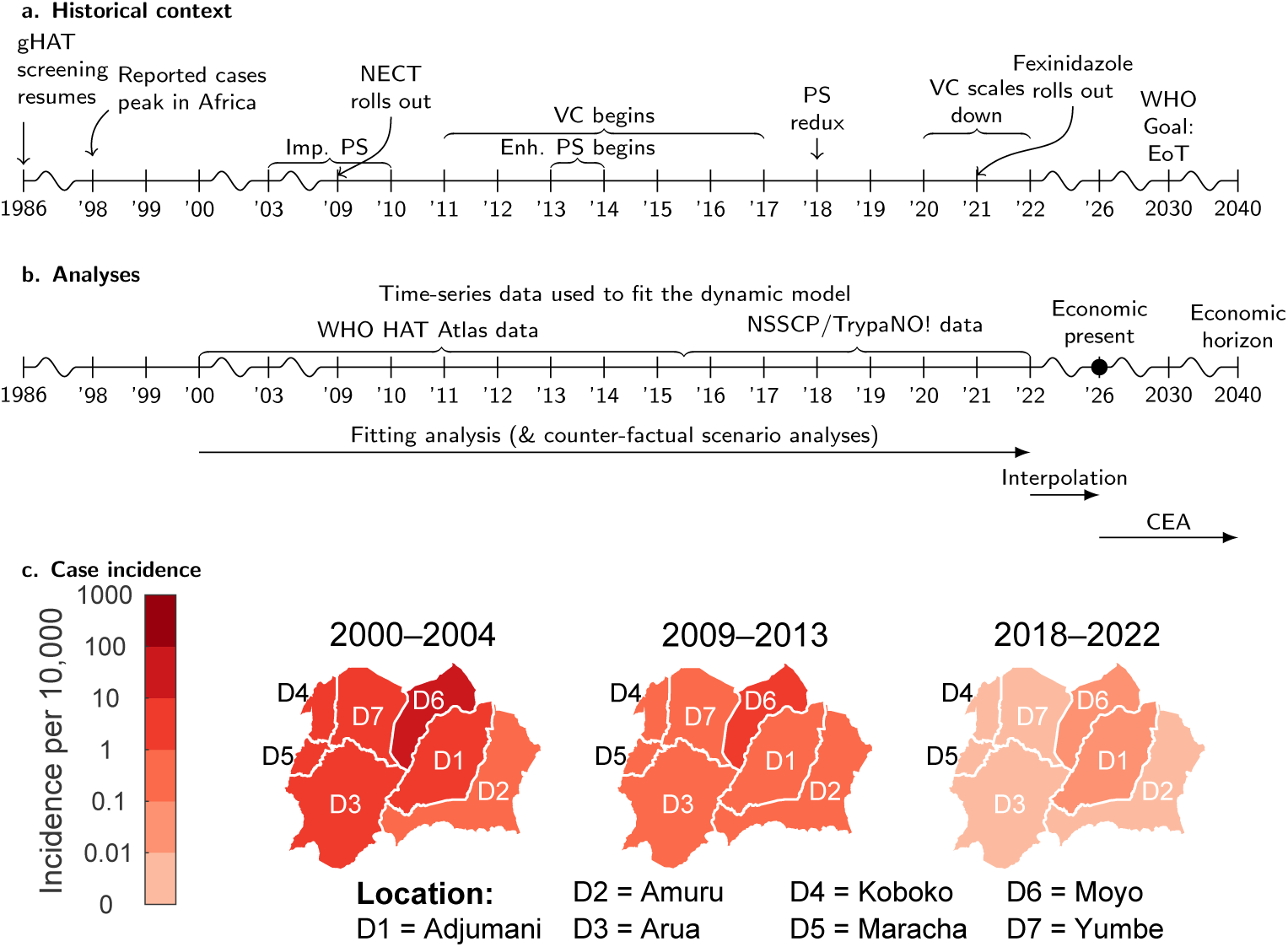
Timeline of major events in Ugandan gHAT history and our analysis and the evolution of case incidence. (a) Timeline of major events in Ugandan gHAT history. Vector control started in Arua and Maracha (December 2011), then Koboko (December 2012), Moyo (January 2014) and Yumbe (November 2014), and finally in Adjumani and Amuru (June 2017). Maracha stopped deploying Tiny Targets in mid-2020, and other districts ceased deployments in 2023. (b) Timeline of our analysis. (c) Case incidence per 10,000 inhabitants per year from the WHO HAT Atlas (2000–2015) and Trypa-NO! project (2016–2022). Shapefiles used to produce these maps are available under an academic publishing license allowing CC-BY publication at https://gadm.org/download_country_v3.html. Abbreviations: Enh. PS: enhanced passive screening that occurred due to expansion of RDT availability throughout districts, as described in [25]. Imp. PS: Improvement in PS that occurred due to better skills in diagnosis, as fitted by the parameter in the model. VC: vector control. EoT: elimination of transmission. CEA: cost-effectiveness analysis. NSSCP: National Sleeping Sickness Control Program. TrypaNO!: Gates-funded consortium to drive several gHAT-endemic countries to elimination.

AS, a medical intervention, is the mass screening of individuals living in “at-risk” villages. WHO has produced guidelines for selecting villages to be screened by mobile teams based on case reporting from previous years [24]; AS is recommended to continue until there are three years of zero cases reported, after which AS can be scaled back if no further cases are detected during a supplementary AS in the fifth year [24]. The mobile teams use the Card Agglutination Test for Trypanosomiasis (CATT) to detect trypanosome-specific antibodies byusing one drop of finger-prick blood per person [24]. Those positive on CATT at a 1/4 dilution have neck lymph glands examined and, if palpable, aspirated for microscopic examination. Otherwise, if negative, blood is examined for parasites (using capillary tube centrifugation (CTC)), and optionally by examination of cerebrospinal fluid (CSF) if there is a high index of suspicion. A confirmed sighting of the parasite in blood, lymph fluid, or CSF is considered a confirmed case.

PS is another medical intervention and refers to screening and care at local fixed healthcare facilities where there are staff trained to recognise gHAT symptoms and obtain a diagnosis via the appropriate algorithm. Historically, CATT tests have been used in PS when the prevalence of gHAT was high, but CATT tests has been replaced by rapid diagnostic tests (RDTs) since 2013 [25]. Similar to AS, visualisation of the parasite by microscopy, and therefore confirmation of the case, is required in PS before treatment.

In 2017 there were a total of 170 health facilities with HAT diagnostic capabilities. Due to very low case numbers, a careful scale-back operation was put in place making use of a statistical modelling approach to reduce resource use whilst maintaining high coverage for at-risk people [26]. Since 2018, there are a total of 52 facilities with the capacity for RDT screening across the 7 former districts, and 13 of these facilities have the ability to confirm cases, distributed so that at least one confirmation facility exists in each district. If facilities without confirmation find an RDT-positive person, the person is referred to the confirmation centre for follow-up testing before treatment can be administered [25, 26]. However, in Uganda, like in DRC, referral completion is low in part due to provider communication or costs [27, 28].

Following a case confirmation in either AS or PS, treatment must be administered, as the likelihood of death without treatment is high [29]. Before 2020, the cerebrospinal fluid (CSF) examination and white blood cell count would determine whether the trypanosomes had penetrated the blood-brain barrier, which typically occurs after a person has been infected for 1–2 years. Cases were staged based on the presence of trypanosomes in the cerebrospinal fluid (stage 1 if absent, stage 2 if present), which determined the appropriate treatment. Since the colonial period, stage 1 patients received pentamidine and stage 2 melarsoprol. In 2007, nifurtimox-eflornithine combination therapy (NECT) became available for stage 2 infections, which reduced the toxicity of treatment [24]. Treatments for both stages were given intravenously in the hospital over 10–14 days. In 2020, a new oral treatment, fexinidazole, was approved that could be used for both stage 1 and 2 cases but it has not been used in Uganda because the last cases were reported before approval of the drug.

VC using Tiny Targets was rolled out in all seven former districts during 2012– 2017. Initial VC deployment areas were followed by multiple scaleups and deployments took place twice per year [7]. Another study estimated that Tiny Targets reduced the incidence of newly reported cases of gHAT in Uganda by 25% between 2012– 2019 [30]. Similar to the scale-back of PS, due to low case reporting and low tsetse abundance found in routine entomological monitoring, VC has now stopped in all districts; Maracha stopped VC in mid-2020 and the rest of the districts stopped in 2023.

### Demography, screening and case data

Historical case data covering the period 2000–2015 were obtained from the WHO HAT Atlas [2, 31]. Using geolocation data, where available, we mapped the HAT Atlas entries into the seven districts of northern Uganda (see Figures S2–S3 and Table S2 in Supplement 1). If the geolocation was unknown, we used the listed district information. The WHO HAT Atlas data were then aggregated by year within allocated districts to provide annual numbers of reported cases and people actively screened. The data on reported cases further indicated the type of screening that detected the case (AS or PS), and the disease stage (1, 2 or unknown). Within these data, there were 44 district-year combinations for which the number of people actively screened was not available. The Ugandan National Sleeping Sickness Programme (NSSCP) were able to retrieve some missing information from local records, along with additional information on the stage of the disease, and these values were incorporated into the aggregated data set. Despite these efforts, there remained years in which the number of people actively screened was unknown (Yumbe has four years with unknown screening numbers; Adjumani, Koboko and Maracha have two each; and Arua has one). Lastly, aggregated data in the same format was available for 2016–2024 from the Trypa-NO! project [32] data repository, which were appended to create a complete 2000–2022 dataset.

There were several large refugee settlements in the gHAT endemic region of Uganda, and cases reported from these refugee populations after VC implementation started were excluded from the dataset under the assumption that VC targeted local transmission and that it is *>* 60% effective at killing the vector. Consequently, infections reported among refugee populations after VC began in Uganda are highly likely to have occurred in South Sudan, not in Uganda. Figure 1c shows the trend of case incidence across three different time periods in each district based on the aggregated data from these different sources.

### Transmission model

The model used in this study was previously developed by Rock et al. [17]. Several refinements have been made since the original publication [9, 19, 33–37] through discussions with the NSSCP and implementing partners (LSTM and FIND) and fitting to data from different countries to reflect new knowledge and requirements. A sensitivity analysis of the model was previously performed [38]. It is a compartmental transmission model incorporating disease dynamics in humans, tsetse and – in some model variants – non-human animals. Eight model variants, described in previous studies for Chad [9, 19] use alternative human-population stratifications to represent low-and high-risk exposure to tsetse, participation or not in AS, and the presence or absence of animal transmission (Model 6–8 in Table 2). See Supplement 1 for model equations, illustration (Figure S4), and parameter tables (Tables S3–S5).

As all districts had VC for a portion of the 2000–2024 period, we had to account for changes to the tsetse population in our model. We assumed that deployments always happened in the beginning and middle of the year (see Table S6 in Supplement 1) and VC coverage is a constant (see Table S7 in Supplement 1), although the actual deployment dates vary. In our tsetse model, the dynamics under VC (see Figures S5– S7 in Supplement 1) are determined by the tsetse birth rate (*B_V_*), the probability of hitting targets and dying (*p*_targetdie_), and other biological parameters (see Tables S3 and S8 in Supplement 1).

The team also provided case coverage for each district based on the method of assigning deployments to their watersheds [30]. Combining the assumed or estimated. VC effectiveness with the average case coverage data in each district from the Trypa-NO! project data repository, we could then model the tsetse population dynamics under VC in each district.

### Uganda-specific model assumptions

In addition to using case, screening, and VC data collected from the seven endemic districts in Uganda as direct inputs to our model, some specific model assumptions were made based on local contextual information. The timelines in Figure 1a–1b provide graphical summary for this information.

For AS, we assumed that the specificity of the AS algorithm improved to 100% in 2011 due to overall improvements in the programme and sudden drops in case reporting in all seven districts in Uganda.

For PS, with the introduction of the gHAT RDT in 2013–14 [25], the number of health facilities offering gHAT screening increased in all seven districts in Uganda. Coupled with staff training and community sensitisation, we believe this decreased the average time to detection. In each endemic district in Uganda, the same broad priors (one for stage 1 detections and one for stage 2) were used to fit the timing of overall PS improvement, as well as the amount of improvement in all model variants. Broad priors allow us to express our uncertainty as to when different PS improvements began, how long improvements took to reach their maximum potential, and the magnitude of this improvement. During the model fitting process, parameters are selected by matching to local case reporting data, and therefore return different sets of passive detection rates in different districts.

For VC, we used reported tsetse capture data [7] to estimate district-specific *B_V_* and *p*_targetdie_ in Arua, Koboko, and Maracha. For districts where tsetse capture data were unavailable, we assumed the effectiveness of VC (i.e. the reduction in tsetse numbers) to be 80% in Adjumani and Amuru and 90% in Moyo and Yumbe after one year of deployment. These assumptions were based on discussions with those conducting VC deployments on the ground in Uganda and were informed by observations that a higher proportion of flies (compared to the pre-intervention level) were present after VC deployment in Adjumani and Amuru compared with Moyo and Yumbe, in line with a tsetse habitat suitability analysis performed recently using a spatiotemporal model [39].

### Fitting to historical data

Deterministic model variants were fitted to historical data for each district using an adaptive Markov chain Monte Carlo (MCMC) algorithm which has previously been used to fit to gHAT data from DRC, Chad and Ĉote d’Ivoire [12, 19, 36, 37]. In summary, this statistical fitting algorithm tests at least 100,000 different possible parameter sets to find the district-specific parameters where the model looks most like the observed district case data. Details of how the likelihood function, which measures how close the model outputs are to the observed data, is computed can be found in the Section S7.1 in Supplement 1.

We assumed AS started two years before the data period in 1998, when the CATT test became available, thereby moving the system away from its endemic equilibrium. The annual number of people actively screened is pivotally important to our analysis, as it has a direct influence on case detections, enabling us to make inferences on unobserved transmission dynamics. However, the data was not always complete and although efforts made by the Ugandan national programme to digitise paper records reduced the amount of missing information, in six of the seven districts our datasets remained incomplete. For that reason, our MCMC method imputes the screening coverage in the years of missing data. Moreover, a second piece of pivotal information to study unobserved dynamics, is screening type and the disease stage data, which enabled the estimation of the time when detected cases were originally infected. To this data we had to calculate errors made due to the diagnostic properties of the algorithms, we assumed the diagnostic sensitivity of the AS algorithm was fixed at 0.91, while the specificity until 2011 was fitted with a prior with 95% of its mass between 0.9989 and 0.9999.

PS was also assumed to improve in 1998 with the introduction of the CATT test. To reflect this, our fitted parameters included a parameter for the rate of case detection from stage 2 by PS pre-1998, a parameter for the rate of case detection by PS from stage 1 from 1998, and a parameter for the rate of case detection by PS from stage 2 from 1998. We assumed there was no case detection by PS from stage 1 before 1998.

Each model variant and district combination was fitted separately. This procedure treats districts in isolation, taking no account of the movement of humans or tsetse between districts or migration from outside the country. Parameters that are not believed to vary geographically, for which estimates are available, were fixed in the model (see Table S3 in Supplement 1). The model fitting provides 2000 samples from joint district-specific posterior distributions for the fitted parameters of each model (see Table S4 in Supplement 1). The number of fitted model parameters ranges from 10 in the simplest model variant (Model 1 in Table 2) to 16 in the most complex (Model 8).

### Ensemble model

Once we had fitted the eight models, we combined the posterior estimates of each model into an “ensemble” model, which is a weighted combination of all the models. Weightings were computed for each district separately by assessing the support for each model by the data. We used our ensemble model to make future projections under a range of strategies. Technical details of this are given in Section S7.4 in Supplement 1.

### Filtering

To ensure that our modelling results aligned with the observed data, filtering was applied to the stochastic simulation results for the period with observations, retaining realisations that matched the observed presence/absence of reported cases in the last *n* + 1 years, where *n* is the number of consecutive years without case reporting. To incorporate structural and observational uncertainties, we first simulated 500,000 realisations for our projections by running 250 realisations from each of 2,000 ensemble model samples from the joint posterior of the fitted model parameters in each district. We then discarded realisations that contradicted the observed presence/absence of district-level reported cases in the last *n* + 1 years, which means we discarded realisations which had cases in the last *n* years when there were no reported cases, and had no cases in the last year with actual case reporting. The last reported cases in Adjumani, Amuru, Arua, Koboko, Maracha, Moyo, and Yumbe were in 2019, 2016, 2016, 2015, 2012, 2016, and 2018. Therefore, the *n* is equal to 6, 9, 9, 10, 13, 9, 7, respectively. N.B. We did not have case numbers in 2025 at the time of analysis. We aimed to present our projection results based on 20,000 realisations from each district. If there were fewer than 20,000 realisations left after this filtering process in any district, we reran simulations in these districts until we generated the targeted number of filtered realisations.

To be able to estimate the reduction in transmission and deaths due to VC, we ran a counterfactual scenario (CFS). The CFS simulated disease transmission in the absence of VC in each district while simulating all other interventions (PS and AS) in identical intensity to our main analysis. We computed the cumulative number of new infections and deaths under the CFS and the factual scenario, and we used the difference in infections and deaths from these two scenarios to compute the attributable impact of the VC component of the strategy. We used the reproducible method of simulation for stochastic models described in Sunnucks et al. [40] to ensure fair comparison between strategies whilst preserving the random nature of transmission dynamics near elimination. See the Section S9 in Supplement 1 for further details of the calculation.

### Projections

Because of the low number of cases being reported in conjunction with a range of different interventions (including robust surveillance) over the last 10 years, we believe that transmission has been very low in recent years in all endemic districts in Uganda. To better capture the dynamics around elimination, we used stochastic projections, which consider the movements between any two compartments as chance events instead of fixed rates as in the deterministic approach. In the stochastic simulations, infection transmission and progression events are drawn randomly with a mean equal to the rate used in the deterministic model. Previous modelling studies have demonstrated good agreement between stochastic and deterministic models of gHAT [41]. Thus, we performed stochastic projections within each district from 2026 for 30 years into the future based on the parameters from the deterministic model. There is a three-year gap between the end of the fitted period (2022) and the beginning of our projections (2026); to ensure that the projections start in 2026 before simulating the different future strategies, we used the AS coverage for 2023–2024 and assumed AS carried on with the mean coverage of 2020–2024 in 2025.

Our projections implement a range of possible future intervention strategies shown in Table 3. All strategies assume PS was maintained at the same level as at the end of the fitted period (i.e. the same as 2022) but with different combinations of vertical interventions (AS and VC). The specificity of the diagnostic algorithm is assumed to continue to be 100%. The *Stop 2026* strategy (i.e. PS only with no regular AS and VC) is the current strategy in Uganda due to low case reporting. Despite the absence of both AS and VC in all districts, we simulated the *Mean AS* and *Mean AS + VC* strategies to assess whether the current strategy is more cost-effective than reinstating any vertical interventions. In these strategies, we simulate at least one year of AS at the mean level of 2020–2024 starting in 2026 and subsequently use the cessation criteria described below to decide when to cease routine vertical intervention(s) and switch to a reactive strategy involving reactive screening (RS) and reactive VC (RVC). The *Max AS (d2d)* strategy was considered to investigate if higher coverage, but also proportionally higher coverage among high-risk people (e.g. by performing door-to-door screenings), can detect any remaining infections. We assumed that people who would not previously present for AS would have an equal probability of being screened compared to those who would have participated in the past; we expect that this would have a higher effectiveness of detection per person screened. Finally, the *Reduced AS* strategy was suggested by partners to explore if there would be value in the NSSCP spending a small amount of money to implement a low coverage of AS, equivalent to 50% of the coverage of the mean AS for 2020–2024. The mean, maximum, and reduced AS coverages vary from district to district (see Table 1 and mean and reduced coverage turned out to be zero in Amuru, making *Stop 2026*, *Mean AS*, and *Reduced AS* equivalent strategies.

**Table 1:** Screening coverage in the districts. Absolute numbers are shown together with the equivalent percentages, expressed as a proportion of the 2026 population (in brackets), for the mean annual of AS coverage in 2020–2024 and the maximum coverage historically between 2000–2024.

|  | District |  |  |  |  |  |  |
| --- | --- | --- | --- | --- | --- | --- | --- |
|  | Adjumani | Amuru | Arua | Koboko | Maracha | Moyo | Yumbe |
| Mean AS | 1,572<br>(0.6%) | 0<br>(0%) | 2,323<br>(0.2%) | 3,628<br>(1.1%) | 600<br>(0.3%) | 3,555<br>(1.9%) | 652<br>(0.1%) |
| Max AS | 5,751<br>(2.3%) | 1,305<br>(0.5%) | 26,118<br>(2.4%) | 30,442<br>(9.2%) | 5,680<br>(2.4%) | 34,755<br>(18.6%) | 22,188<br>(2.4%) |

**Table 2:** Differences between model variants considered in this study. We modelled the population’s participation in AS as one of two broad types. The “Random” column indicates individuals willing to participate in AS who may choose to attend the AS campaigns that occur in the centre of villages; the percentage of people attending AS is in accordance with observed AS levels. In contrast, the other group of people are assumed not to participate at all (column denoted as “Never”). We also included different risks of exposure to tsetse for different people in our model. High-risk humans receive *r >* 1 times more bites than low-risk humans and therefore are more likely to be infected. The possibility of animals contributing to transmission is considered in some model variants.

| Model | Participation in active screening: |  |  |  | Animal transmission |
| --- | --- | --- | --- | --- | --- |
|  | Random |  | Never |  |  |
|  | Low-risk | High-risk | Low-risk | High-risk |  |
| 1 | ✓ |  |  |  |  |
| 2 | ✓ | ✓ |  |  |  |
| 3 | ✓ |  | ✓ |  |  |
| 4 | ✓ |  |  | ✓ |  |
| 5 | ✓ | ✓ | ✓ | ✓ |  |
| 6 | ✓ |  |  |  | ✓ |
| 7 | ✓ |  |  | ✓ | ✓ |
| 8 | ✓ | ✓ | ✓ | ✓ | ✓ |

**Table 3:** Intervention strategies. Five future strategies were simulated from 2026. These include the current level of passive screening (PS), four levels of active screening (AS) (no AS, reduced AS, mean AS, maximum AS from low to high) before case reporting met cessation criterion, two different participation patterns (low-risk humans only or both low- and high-risk humans) and scale back or cessation criteria for AS, with vector control (VC) continuing until 5 years of no cases in the *MeanAS + VC* strategy.

| Strategy<sup>1</sup><br>(from 2026) | Passive<br>screening<br>(PS) | Active screening (AS) |  |  | Vector<br>control<br>(VC) |
| --- | --- | --- | --- | --- | --- |
|  |  | Coverage | Participant<sup>2</sup> | Scale back<br>criteria<sup>3</sup> |  |
| *Stop 2026* | Continue<br>at the<br>same<br>level<br>as in<br>2025 | None | NA | From 2026 | Stop from 2023 |
| *Max AS (d2d)* |  | Historical<br>maximum | All | 5 years of<br>no cases | Stop from 2023 |
| *Mean AS + VC* |  | Mean of<br>2018–2022 | Traditional AS<br>participants | WHO<br>algorithm<sup>4</sup> | Until 5 years of<br>no cases<sup>3</sup> |
| *Mean AS* |  | Mean of<br>2018–2022 | Traditional AS<br>participants | WHO<br>algorithm<sup>4</sup> | Stop from 2023 |
| *Reduced AS* |  | 50% of mean | Traditional AS<br>participants | WHO<br>algorithm<sup>4</sup> | Stop from 2023 |

<sup>1</sup>Reactive interventions are defined in Table 4.
<sup>2</sup>Randomly participating depending on the coverage of AS.
<sup>3</sup>Earliest possible year to scale back vertical interventions is 2026.
<sup>4</sup>5 years of no cases; no year 4 screening.

The cessation criteria we used in our modelled strategies with AS were based on WHO guidelines and involve annual AS continuing until there are three consecutive calendar years with no case reporting, then a further screening in the fifth year after the last reported case [24]. In the *Max AS (d2d)* strategy, we also simulate a screening in the fourth year, making five consecutive years of screening without cases before cessation in order to test whether RS is avoidable by implementing additional AS. If a strategy included VC, we assumed that VC continued until after the fifth year of no case reporting, so that VC cessation coincides with the cessation of AS. In any case, both AS and VC continue for at least one year in the strategies containing them, meaning that the earliest possible switch to RS occurs in 2027, which means all strategy components were implemented for at least one year in projections, even if there had already been five years of no cases in the district. This approach avoided simulating strategies which were expected to have AS and/or VC but were in districts that had met the cessation criteria. Once vertical interventions were scaled back, reactive interventions were simulated when cases were detected.

RS and RVC were triggered as a reaction to future case detection after all vertical interventions had met their cessation criterion and been scaled back. To reflect the RS plans designed by the NSSCP and Trypa-NO!, we considered the reactive interventions for each strategy (see Table 4) and had the following assumptions in our model: 1. The initial RS targets a coverage equal to the average village population and can scale back immediately if no new cases are found; 2. If more cases are detected during initial RS, indicating potential remaining transmission, regular RS with the same number of people screened each year will be scheduled until the WHO classic algorithm is met; 3. Each year with non-zero cases, two rounds of small-scale RVC (approximately 50 km^2^ with 100% case coverage) will be implemented the following year during dry seasons at the beginning and the middle of the year (same as historical VC); 4. We simulated the scaleback of PS following 10 years with no reported cases. Scaleback changed the passive detection rate parameters to their 1998 (pre-improvement) values, based to the assumption that the number of health facilities with HAT diagnostics and trained staff would be reduced after a decade without cases.

**Table 4:**
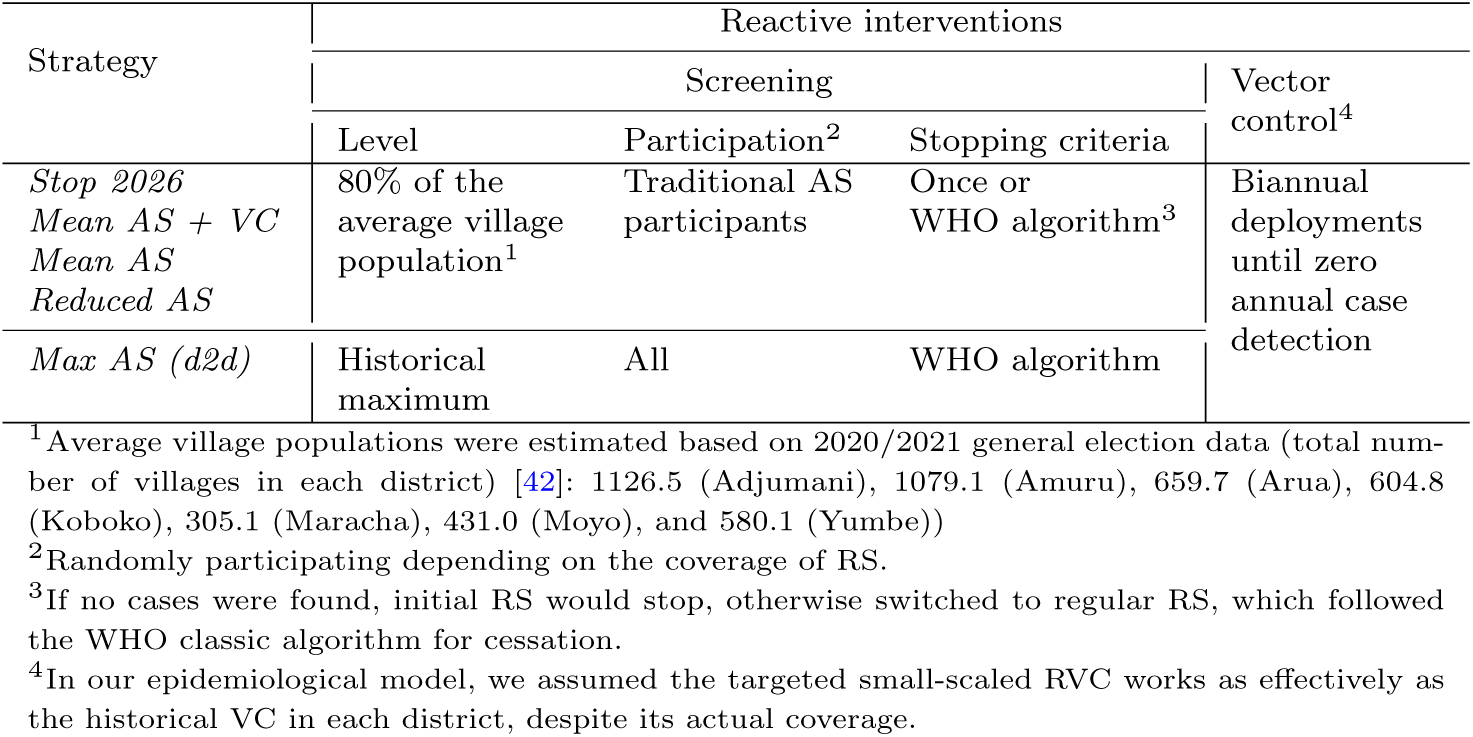
Reactive intervention for each strategy. Reactive interventions, which define the algorithms reacting to case findings after cessation, were considered as parts of the strategies.

| Strategy | Reactive interventions |  |  |  |
| --- | --- | --- | --- | --- |
|  | Screening |  |  | Vector control<sup>4</sup> |
|  | Level | Participation<sup>2</sup> | Stopping criteria |  |
| *Stop 2026*<br>*Mean AS + VC*<br>*Mean AS*<br>*Reduced AS* | 80% of the average village population<sup>1</sup> | Traditional AS participants | Once or WHO algorithm<sup>3</sup> | Biannual deployments until zero annual case detection |
| *Max AS (d2d)* | Historical maximum | All | WHO algorithm |  |

<sup>1</sup> Average village populations were estimated based on 2020/2021 general election data (total number of villages in each district) [42]: 1126.5 (Adjumani), 1079.1 (Amuru), 659.7 (Arua), 604.8 (Koboko), 305.1 (Maracha), 431.0 (Moyo), and 580.1 (Yumbe)
<sup>2</sup> Randomly participating depending on the coverage of RS.
<sup>3</sup> If no cases were found, initial RS would stop, otherwise switched to regular RS, which followed the WHO classic algorithm for cessation.
<sup>4</sup> In our epidemiological model, we assumed the targeted small-scaled RVC works as effectively as the historical VC in each district, despite its actual coverage.

### Elimination predictions

Whilst the underlying process is not observable in the real world, within our modelling framework we directly simulate transmission and infection and can therefore predict when we expect to have no further transmission events to human and animal hosts in each district (last transmission event; LTE). We can also measure when the last hosts cease to be infected to predict when there is no remaining infection (NRI). For each district we compute the probabilities that LTE and NRI by year have been achieved by taking the proportion of realisations meeting that target during or before that year out of the 20,000 realisations.

We can then compare our predictions of LTE and NRI. We note that in the Warwick group’s previous modelling work (e.g. [18]), what we called EoT was defined as the year after what we refer to here as LTE, as discussed elsewhere [43].

### Country-level aggregations

To scale from individual districts to the country level, we must aggregate our results. To aggregate the number of cases, deaths and other outputs and compute the 95% predictive intervals we assume perfect correlation (i.e. rank-correlation equal to 1) between all districts. Under this assumption, district-level outcomes are perfectly aligned, such that upper (or lower) bounds occur simultaneously across all districts. Before the assumption of rank-correlation=1 was made in other analyses, it was found that simply summing results across regions (total independence, zero correlation) gave implausibly narrow uncertainty when considering aggregates to the country level [18]. In contrast to the assumption of independence between districts, which minimises the reported uncertainty in aggregated country-level results, assuming perfect rank correlation between districts maximises the reported uncertainty in the aggregated country-level results. At low levels of transmission, as in Uganda, this is expected to have minimal impact on the presented results.

For aggregated estimates of the probability of elimination at the national level by year, we likewise assumed perfect correlation between districts, in line with case and infection aggregations. For each realization, the national year in which each of the goals is achieved (LTE, NRI or 5 years of no cases) was defined as the maximum year of achievement across all districts, after sorting district-level realizations by the year of target achievement. The probability of reaching a goal by a given year is then the proportion of realisations that have reached the goal before or during a given year. This approach yields a prediction interval for timing of the goals at the national level that is wider than under an assumption of independence between districts.

### Costs and Cost-effectiveness analysis

To assess the costs and cost-effectiveness for the elimination of gHAT in Uganda we used a cost model coupled with the transmission model to estimate economic costs of continuing strategies between 2026 and 2040, as shown in Figure S8 of Supplement 1. Parameters for the model are presented in Table S9–S10 in Supplement 1 and in more detail in Supplement 2: Parameter Glossary. The treatment model is shown in Figure S9 of Supplement 1, and the eligibility for fexinidazole, pentamidine, and NECT are calculated in Table S11, while the treatment outcomes are calculated in Table S12 of Supplement 1. This cost model is adapted from health economic evaluations of gHAT strategies in the DRC and Chad and shown with Uganda-specific parameters in Tables S13–S19 in Supplement 1 [18, 44, 45]. The dynamic model outputs yield inputs for the economic model in terms of cases detected, cases not detected (which accrue deaths and DALYs), years when AS and VC are operating, and occurrences of RS and RVC. In the economic model, we also account for PS scaleback and lower costs) after 10 years of zero cases, in line with the assumptions of the transmission model.

The model takes the Ministry of Health and donor perspective and includes all direct costs associated with AS, PS, treatment, VC, sensitisation, and management. The economic costs include donor costs which are not part of the NSSCP budget such as Tiny Targets (which are currently donated by Vestergaard) and drugs (which are donated by Bayer and Sanofi and shipped by WHO). Indirect costs such as the loss of income to HAT patients or out-of-pocket payments by patients for things like travel to health facilities, are not included in this analysis. All costs in this analysis are denoted in 2023 US$ and a default 3% discounting rate is used. As we have uncertainty in our costs, for our simulations we draw unit costs from a probability distribution; this generates total cost variation in our outcomes even for the same strategy.

Key differences between the cost model used in this study and the previous analyses for other countries are:

- The number of health facilities per district in Uganda and their RDT usage is parameterised using local data (see Table S10).
- Costs of management associated with PS, transport of RDT-positive samples for confirmatory testing, and allowances paid to LAMP centres are all taken from the NSSCP’s 2023 budget (see Section S10.4.1 in Supplement 1).
- Costs of case confirmation are fixed on a per-clinic basis, rather than on the basis of suspected gHAT cases.
- Costs of conducting a reactive screening activity are also taken from the NSSCP’s 2023 budget (see Tables S16).

Primarily, we have used NSSCP budgets and previous literature on health care in Uganda to parameterise our model. We have used literature costs derived from other places where no recent and/or Uganda-specific information was available (e.g. the cost of a lumbar puncture on the rare occasion when one is warranted). VC costs per km^2^ were estimated previously for Uganda but were adjusted to 2023 US$ values, and we have confirmed these estimates with NSSCP coauthors. More details including the cost functions and parameters used can be found in the Section S10 of Supplement 1.

For our cost-effectiveness analysis, we compare economic costs for the different strategies with predicted changes in disease burden. We use the disability-adjusted life year (DALY) as our metric for disease burden, which includes the number of years of life lost to mortality from gHAT and the number of years lived with disability, weighted by the severity of the disease during that time. To assess the probability that each strategy is cost-effective, we use the net monetary benefits (NMB) framework, which takes into account uncertainties in the transmission model forecast for deaths and disease as well as uncertainty in the cost inputs.

The willingness-to-pay (WTP) or how much money should be spent to avert one unit of disease burden (DALY) is a difficult question, as most countries do not have an agreed-upon threshold. However, former guidance by WHO-CHOICE suggested that strategies costing less than the per capita gross domestic product (GDP) of a country per DALY averted were very cost-effective, and those costing less than three times the GDP per DALY averted were cost-effective [46]. Based on Uganda’s GDP in 2023, the thresholds would be $1002 per DALY averted for very cost-effective strategies or $3006 for cost-effective strategies [47]. However, in Uganda, the WTP for health interventions to avert DALYs in the past has been estimated at only $12–313 and $141– 186 (inflated to 2023 US$) [48, 49]. Therefore, we present a range of plausible WTP values from $0–3000 to determine whether our recommended strategy would change across different thresholds. We have run the sensitivity analyses with alternative time horizons, with and without discounting, and we show these results in the companion GUI (https://hatmepp.warwick.ac.uk/uganda/v6/).

As with the country-level aggregations for case reporting and new infections described in the transmission model section, we included aggregations of costs in the same manner; i.e. we assumed rank-correlation=1 between districts. The cost-effectiveness analysis, however, is only performed at a district level as there is no meaningful way to aggregate ICERs.

### Scenario analysis on cross-boundary importation

The location of Uganda raises concerns about cross-boundary movement of both hosts and vectors, which could introduce infection risk to the local community and undermine elimination efforts. The primary foci in South Sudan (Mundri East and West) do not border Uganda, and therefore, we do not consider the importation of infected vectors due to the limited range of movement in tsetse. For the importation of infected hosts, we have no data to quantify the level of infected people and animals arriving from South Sudan, the prevalence in different areas in South Sudan, how many and where the infected hosts are coming from, nor the mixing with the local community in UGA for now and in the future. Therefore, we performed a scenario analysis where a single stage 1 infected human (randomly assigned to a risk/active screening participation group according to the proportion of each group at endemic equilibrium) arrives in the local community to explore the chance of secondary infections and delay on the LTE.

Because VC already scaled back in all seven districts, modelled tsetse populations experience different levels of rebound, depending on birth rates and the remaining tsetse at the time of cessation of VC. Therefore, we considered two specific cross-boundary human importation scenarios, one in 2026 and the other in 2035, to provide a comprehensive overview of the impact of cross-boundary importation.

### Reporting Guidelines

To improve the accessibility and interpretation of this study, we completed the Policy-Relevant Items for Reporting Models in Epidemiology of Neglected Tropical Diseases (PRIME-NTD) summary table [50] to cover five principles for presenting our transmission model results and we used the Consolidated Health Economic Evaluation Reporting Standards (CHEERS) checklist [51] to summarise information of interventions and findings in our cost-effectiveness analysis (see Supplement 3 and Supplement 4).

## 3 Results

### Fitting and the ensemble model

Table 5 shows which model variants are most supported by the data after model fitting (see Figure S12). In the table, the percentages are the weights we use to build our ensemble model for projections – e.g., in Adjumani, 17.2% of the simulations come from Model 1, 40.2% from Model 2, etc. The most favoured model in six of the seven districts is Model 2, which is also the second most favoured model in the seventh district (Amuru). Model 2 has groups of people at low and high risk of infection, but with members of those groups participating in AS at random. There is relatively little evidence for any model that includes animals contributing to transmission (Models 6–8); in all districts except Amuru, fewer than 6% of the ensemble simulations are from these model variants. In Amuru district, the difference between models is less pronounced, but its ensemble still only includes *<*15% from models with animal transmission.

**Table 5:** Relative model evidence (%) – weights used to make up the ensemble model.

| District | Model variant |  |  |  |  |  |  |  |
| --- | --- | --- | --- | --- | --- | --- | --- | --- |
|  | 1 | 2 | 3 | 4 | 5 | 6 | 7 | 8 |
| Adjumani | 17.2 | 40.2 | 15.6 | 2.9 | 19.3 | 1.6 | 0.4 | 2.7 |
| Amuru | 10.2 | 24.1 | 9.7 | 15.9 | 24.4 | 2.8 | 5.6 | 7.2 |
| Arua | 22.4 | 41.2 | 21.1 | 0.4 | 12.5 | 1.3 | <0.1 | 1.0 |
| Koboko | 23.0 | 44.2 | 19.2 | 0.5 | 11.3 | 1.2 | <0.1 | 0.7 |
| Maracha | 17.4 | 41.4 | 15.2 | 2.7 | 17.8 | 2.1 | 0.2 | 3.2 |
| Moyo | 14.4 | 46.2 | 9.8 | 2.6 | 23.7 | 1.4 | 0.2 | 1.7 |
| Yumbe | 29.2 | 39.6 | 28.7 | 1.3 | 0.1 | 1.0 | <0.1 | <0.1 |

Summaries of the fitted parameter estimates are given in Figures S13–S19 in Supplement 1.

Figure 2 shows the fitting results of our ensemble model aggregated at the country level (left) and in the example district of Moyo (right). These results show that, despite variable case reporting during 2000–2022, the model infers there has been a steady decline in new infections across all districts, with an extremely low number of transmission events happening from 2015. Ensemble fitting results for all districts are available in Figures S20–S26 in Supplement 1 and our graphical user interface (GUI) at: https://hatmepp.warwick.ac.uk/uganda/v6/.

**Fig. 2:**
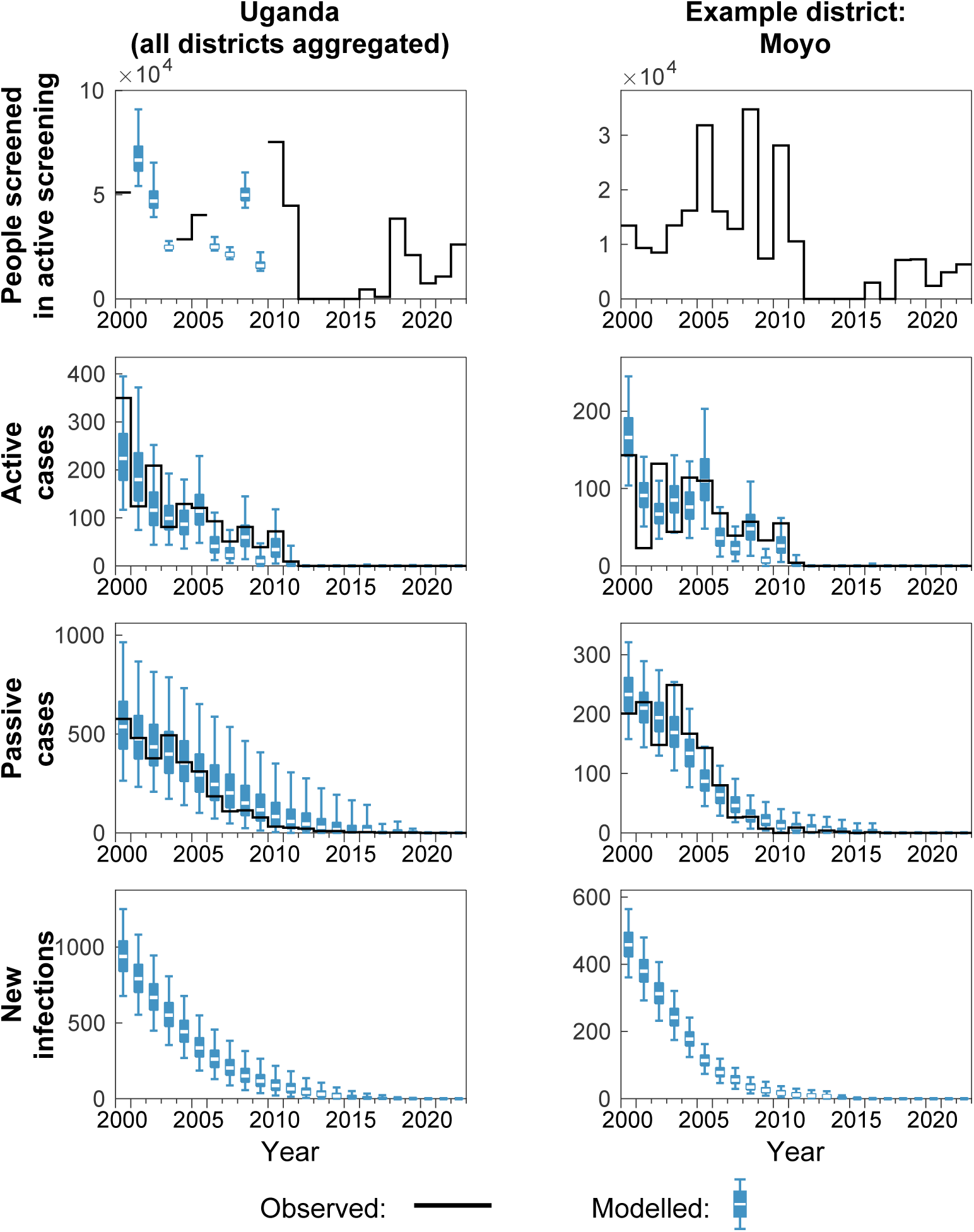
Results of model fitting aggregated at country level (left) and for the example district Moyo (right). Model fitting was independently performed at the district level and results were aggregated in all districts to produce the results for the whole country (Uganda panel, left column). Estimation of missing active screening (AS) information was performed in 2001 (Yumbe), 2002 (Arua and Koboko), 2003 (Koboko), 2006 (Maracha and Yumbe), 2007 (Maracha), 2008 (Adjumani and Yumbe) and 2009 (Adjumani and Yumbe). The right panel shows an example result of the model fitting for Moyo district which has no missing data. All data is shown as black lines, and model outputs as blue box and whisker plots (the middle is the median and the whiskers represent 95% credible intervals). The top row shows the coverage of AS each year, the second row shows the active case reporting, the third row shows passive case reporting and the bottom row shows the estimated number of new infections each year.

By running the CFS to compute the predicted impact of a strategy without VC in the past and comparing it to the actual strategy that occurred (with VC) we have been able to compute the reduction in new infections and deaths attributable to the introduction of VC for each district (see Table 6). The model suggests that, whilst there was high uncertainty in our estimates due to already low prevalence in each district when VC began, around 69–84% of the reduction in transmission in each district could be attributable to the VC intervention and around 22–44% of the reduction in deaths for each district could be attributable to VC. As VC is aimed at preventing people becoming newly infected, it is unsurprising that there was more impact of the VC intervention on transmission compared to deaths. However, it is also likely that some infections that were averted would have otherwise died, hence the model suggests a positive impact on deaths too.

**Table 6:** Reduction in transmission and deaths attributable to vector control (VC).

| Location | % reduction in cumulative new infections due to VC |  |  | % reduction in cumulative deaths due to VC |  |  |
| --- | --- | --- | --- | --- | --- | --- |
|  | Mean | Median | 95% PI | Mean | Median | 95% PI |
| Adjumani | 69.7 | 64.6 | [0, 100] | 38.6 | 28.6 | [0, 100] |
| Amuru | 74.9 | 69.6 | [0, 100] | 33.6 | 0 | [0, 100] |
| Arua | 74.2 | 72.3 | [46, 86.4] | 40 | 35.7 | [0, 61.5] |
| Koboko | 77.3 | 85.4 | [0, 100] | 24.1 | 0 | [0, 100] |
| Maracha | 73.8 | 80.6 | [0, 100] | 22.6 | 0 | [0, 100] |
| Moyo | 83.3 | 82 | [25, 100] | 43.5 | 36.4 | [0, 92.9] |
| Yumbe | 77.8 | 76 | [0, 100] | 42.5 | 34.6 | [0, 100] |
| Entire country | 75.5 | 74.1 | [37.3, 90.7] | 40.3 | 33.8 | [0, 70.7] |

### Projections

Our transmission model indicates that dynamics were very similar among all strategies within the same district. There were likely very few remaining infections at the beginning of our projections (2026) and, even with the cessation of VC, there was no resurgence of gHAT in humans in any of our simulations. Consequently, we only show projection results here from the *Stop 2026* strategy in the main text. Results from other strategies are available in Figures S27–S33 in Supplement 1 or in our GUI at: https://hatmepp.warwick.ac.uk/uganda/v6/.

Figure 3 shows the probabilities and median years of achieving LTE and NRI in each district and at the country level (see Table S20 in Supplement 1 for 95% PIs). We note that, as it is harder to achieve any of these metrics across the country than in a single district, the country-level predictions are always later than any single district. Therefore, very high probabilities are needed in each district to have high confidence that elimination is achieved everywhere.

**Fig. 3:**
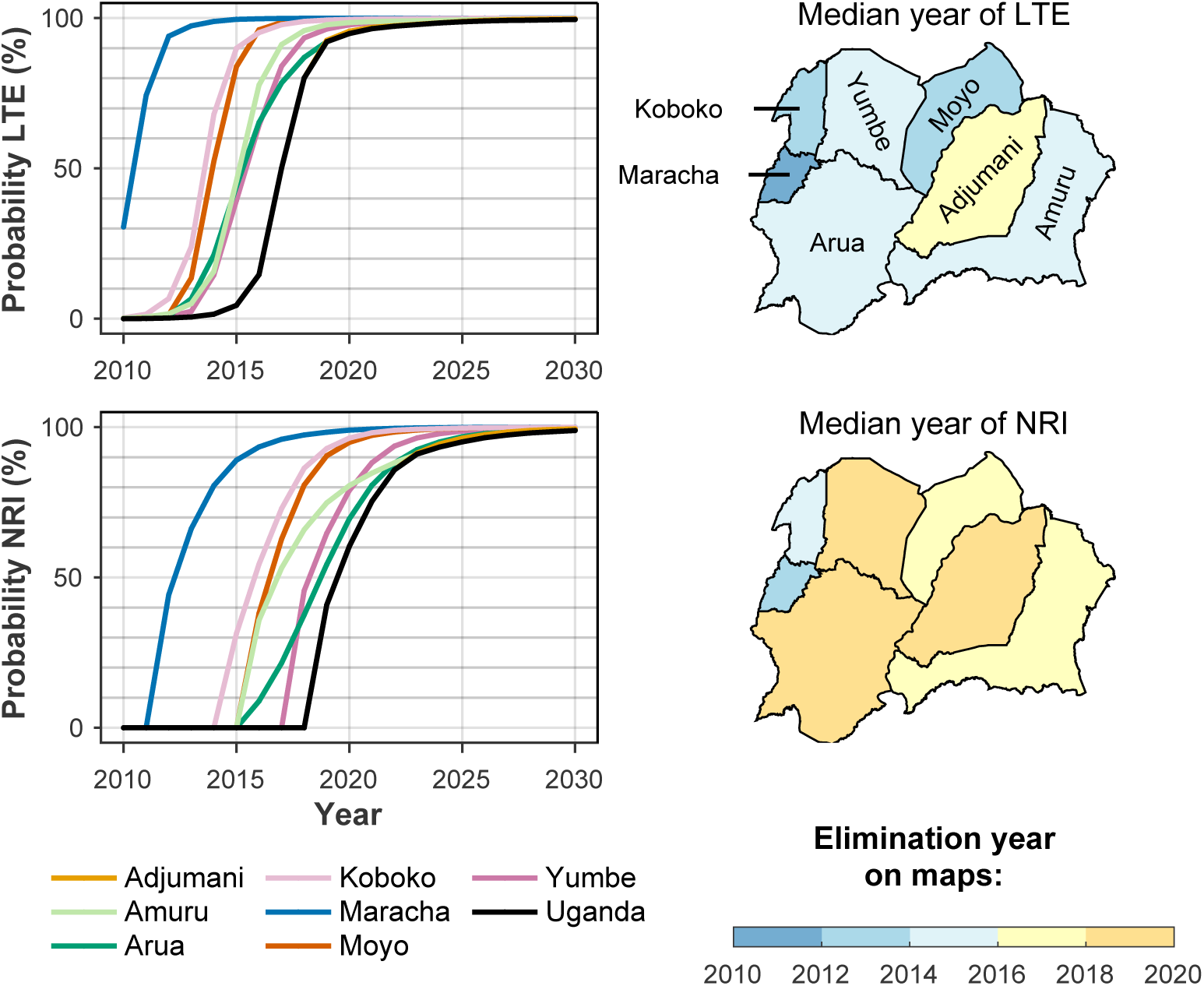
Probabilities and median years of achieving different elimination metrics. Probability curves (left) show the percentage of modelling realisations that match the criteria described below by year in each district (coloured lines) and at the country level (black line) under the *Stop 2026* strategy. Maps show the median year of reaching the criteria in each district under the *Stop 2026* strategy. The definitions of the two elimination metrics presented are: 1. Last transmission event (LTE) – the year in which the final transmission event to humans takes place in the model; 2. No remaining infection (NRI) – the first year in which there are no infected hosts remaining in the model (including both humans and animals capable of acquiring and transmitting infection). Our aggregated results estimate the median years of LTE and NRI for Uganda are 2017 and 2020 respectively. Shapefiles used to produce these maps are available under an academic publishing permitting licence (allowing CC-BY publication) at https://gadm.org/download_country_v3.html.

The probability curves (left panels) present the change in certainty of elimination by year at district and country levels. The probability of LTE (upper left panel) shows that our model predicted Uganda has achieved LTE with high certainty (*>* 95%) in all districts (the earliest in Maracha by 2013 and the latest in Arua by 2021) as well as at the country level (by 2021).

The last remaining infection is necessarily in the same year or after the last transmission event, therefore NRI curves are always after the LTE curves. Adjumani and Amuru are predicted to be the last district to clear infections (in 2025 with *>* 95% certainty) and therefore heavily influence the shape of the whole-country NRI probability curve. The results indicate that we have high confidence (*>*95%) that Uganda will have achieved NRI (no remaining infections) by 2025.

The median year maps (right panels of Figure 3) summarise the average predictions of the year in which the three elimination metrics are met in each district. Our predictions of the median year at the district level vary from 2011 (in Maracha) to 2017 (in Adjumani) for LTE and from 2013 (in Maracha) to 2020 (in Adjumani) for NRI. Our aggregated results estimated the median year of LTE and NRI for Uganda are 2017 and 2020 respectively.

Figure 4 shows when we would expect to have case reporting at the country level under the *Stop 2026* strategy, if there are any after 2025. We see that, whilst it is very unlikely to have case reporting from 2026 onwards, cases would be more likely to be found before 2030 if they do happen. Only 1.125% of our simulations had country-level case reporting after 2025.

**Fig. 4:**
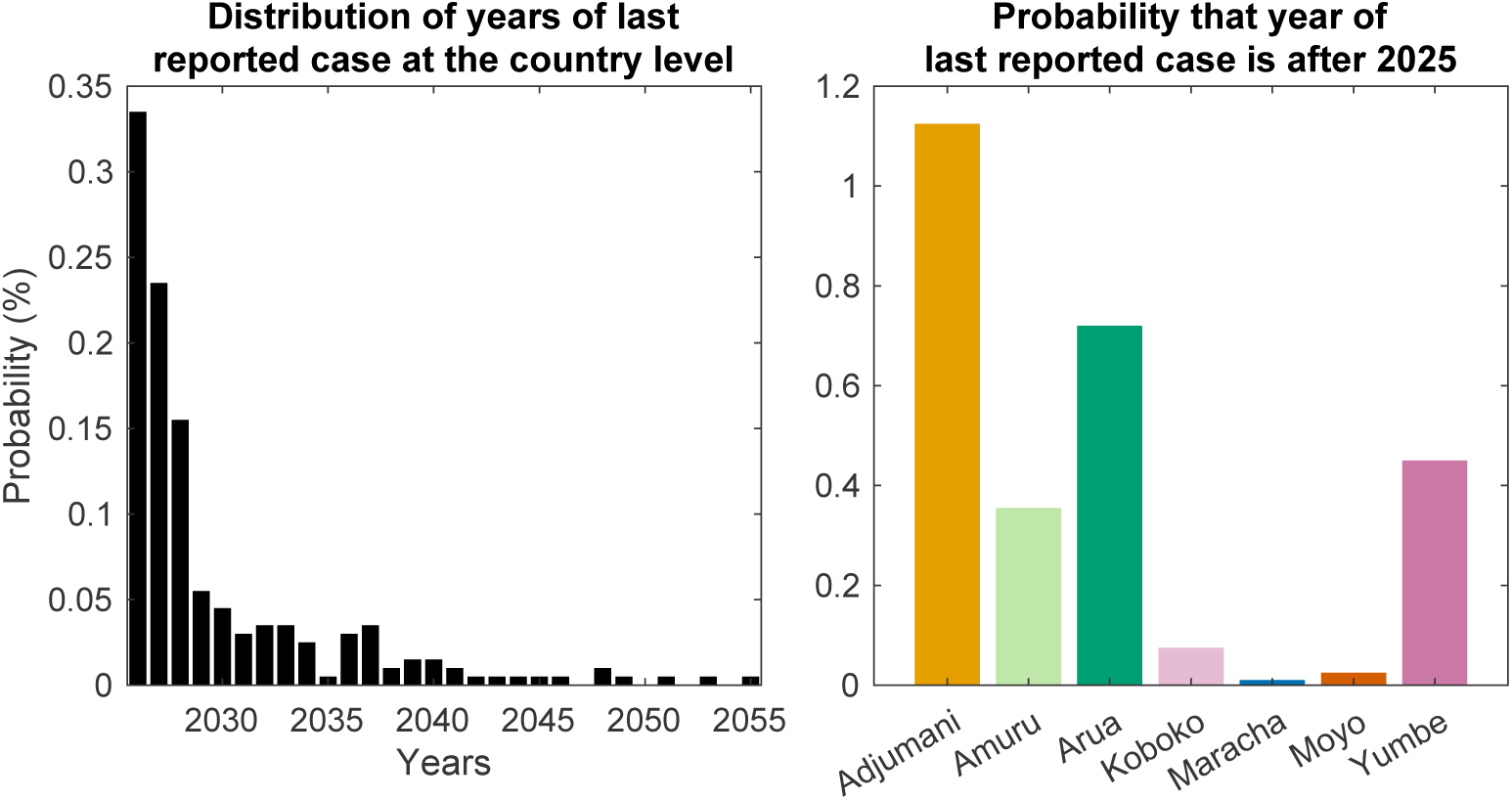
Model prediction for the last reported cases. The left plot shows model predictions of the country-level distribution of when we expect to see case reporting after 2025, e.g. 0.335% of the simulations have cases in 2026. We note that only 1.125% of our simulations have any case reporting for this period, with the majority of simulations finding that the last case has already happened (past years are not shown in the graph). The right plot shows the model predictions for the probability there will be cases reported after 2025 for each district. All simulations in these plots are generated under the *Stop 2026* strategy with filtering applied so that presence/absence of cases in the model realisations matches the observed presence/absence of reported cases for *n* + 1 years (see the ‘Filtering’ section of the Methods for more details).

The same figure also shows the probability of case reporting after 2025 in each district under the *Stop 2026* strategy. The model indicates that in all districts, we have a high level of certainty that the last case has been reported. Adjumani is most likely amongst them to potentially report a case in the future; however, this is still very unlikely—only 1.125% of simulations in Adjumani have reported cases after 2025. Maracha is the least likely district to report cases after 2025.

### Costs and cost-effectiveness analysis

Our cost-effectiveness results are summarised in Table 7 with more detailed information given in Supplementary Tables S21–S27. Table 7 gives the discounted cost and DALY differences between the *Stop 2026* strategy and the other strategies for each district. It provides the incremental cost-effectiveness ratios (ICERs) for each strategy, and also the probability that a particular strategy is cost-effective under four different WTP values. This information can also be viewed in the GUI, (https://hatmepp.warwick.ac.uk/uganda/v6/), which do not change the qualitative results in any substantive manner.

**Table 7:**
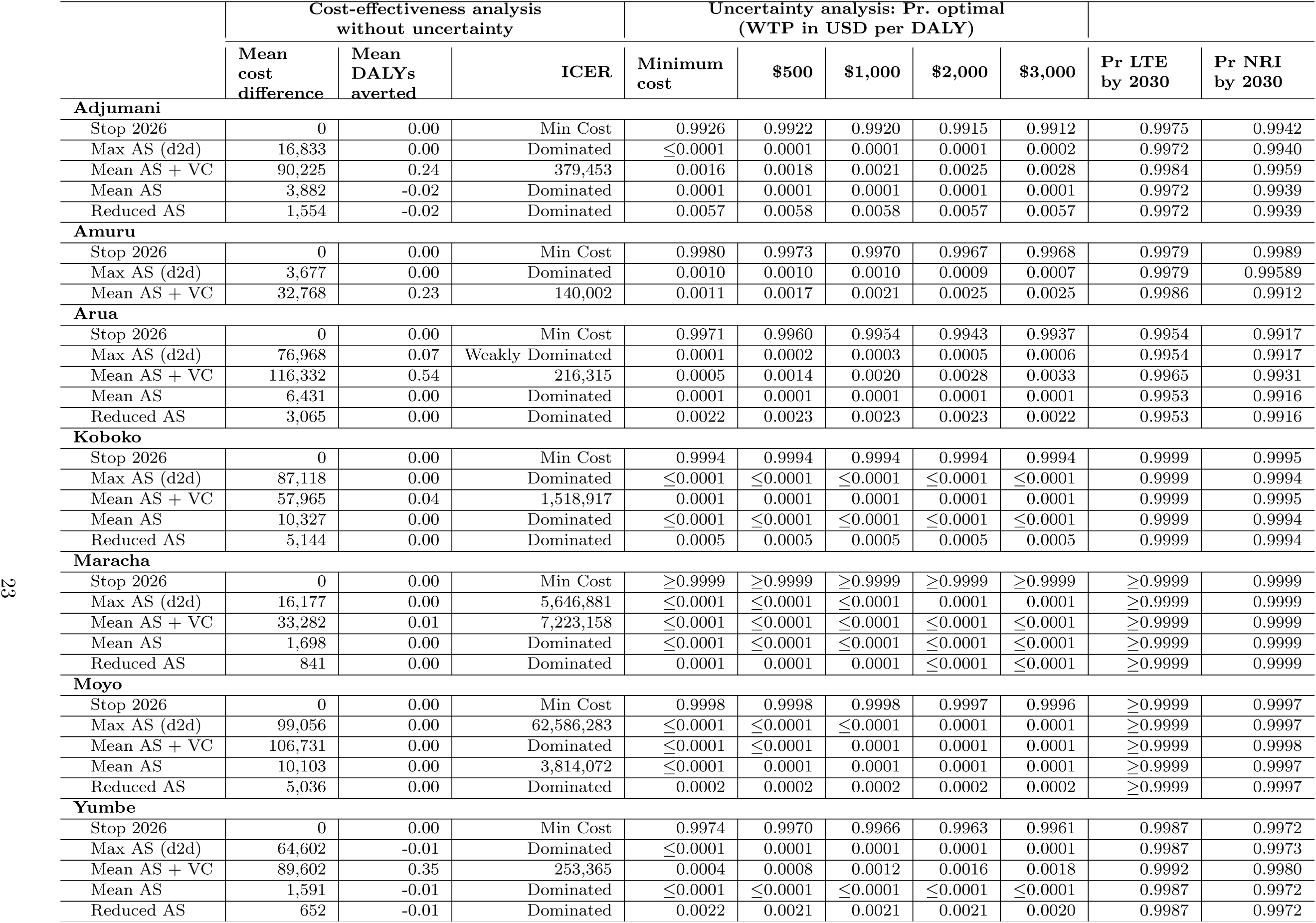
Summary of cost-effectiveness, assuming a time horizon of 2026–2040. Cost differences and DALYs averted are relative to the comparator, which is the first strategy listed for each location. DALYs averted and cost differences are discounted at 3 percent per year in accordance with guidelines for determining cost-effectiveness. In the uncertainty analysis (columns 5–9), the probability that a strategy is optimal is shown as a proportion of all simulations, which allows us to account of parameter uncertainty. The *Stop 2026* strategy is optimal strategy in all districts; it is the strategy for which the mean net monetary benefit (NMB) is highest, equivalent to the information found in cost-effectiveness acceptability curves (CEACs) as well as equivalent to the pie graphs found in the online GUI at https://hatmepp.warwick.ac.uk/uganda/v6/. More details are found in Supplementary Tables S21–S27. ICER: incremental cost-effectiveness ratio, DALY: disability adjusted life-years, Pr: probability, LTE: last transmission event, NRI: no remaining infections. Dominated refers to a strategy that averts fewer DALYs but costs more money than another strategy, while weakly dominated refers to a strategy that has a higher cost-per-DALY averted (ICER) than the next more expensive strategy.

As we have such a high probability that the LTE in Uganda has already occurred and that there are likely few or no remaining infections, most of the strategies lead to similar outcomes in terms of cases and DALYs. *Stop 2026* (the *Status Quo* objective) is always the lowest-cost strategy, and since more expensive strategies yield fewer than 1 DALY averted on average, ICERs are between approximately $191K to over $81M per DALY averted, and the optimal strategy according to the NMB approach to account for uncertainty is always *Stop 2026*. Pie charts showing similar results can be viewed in our GUI under alternative horizons and discounting choices (https://hatmepp.warwick.ac.uk/uganda/v6/). Across alternate assumptions of discounting and time horizons, the recommendation remains *Stop 2026* in all districts. Figure 5 shows that, when present, Max AS or VC would dominate costs in the first year, and for the period 2026-2040, but Mean or Reduced AS would correspond to a small portion of costs compared to PS.

**Fig. 5:**
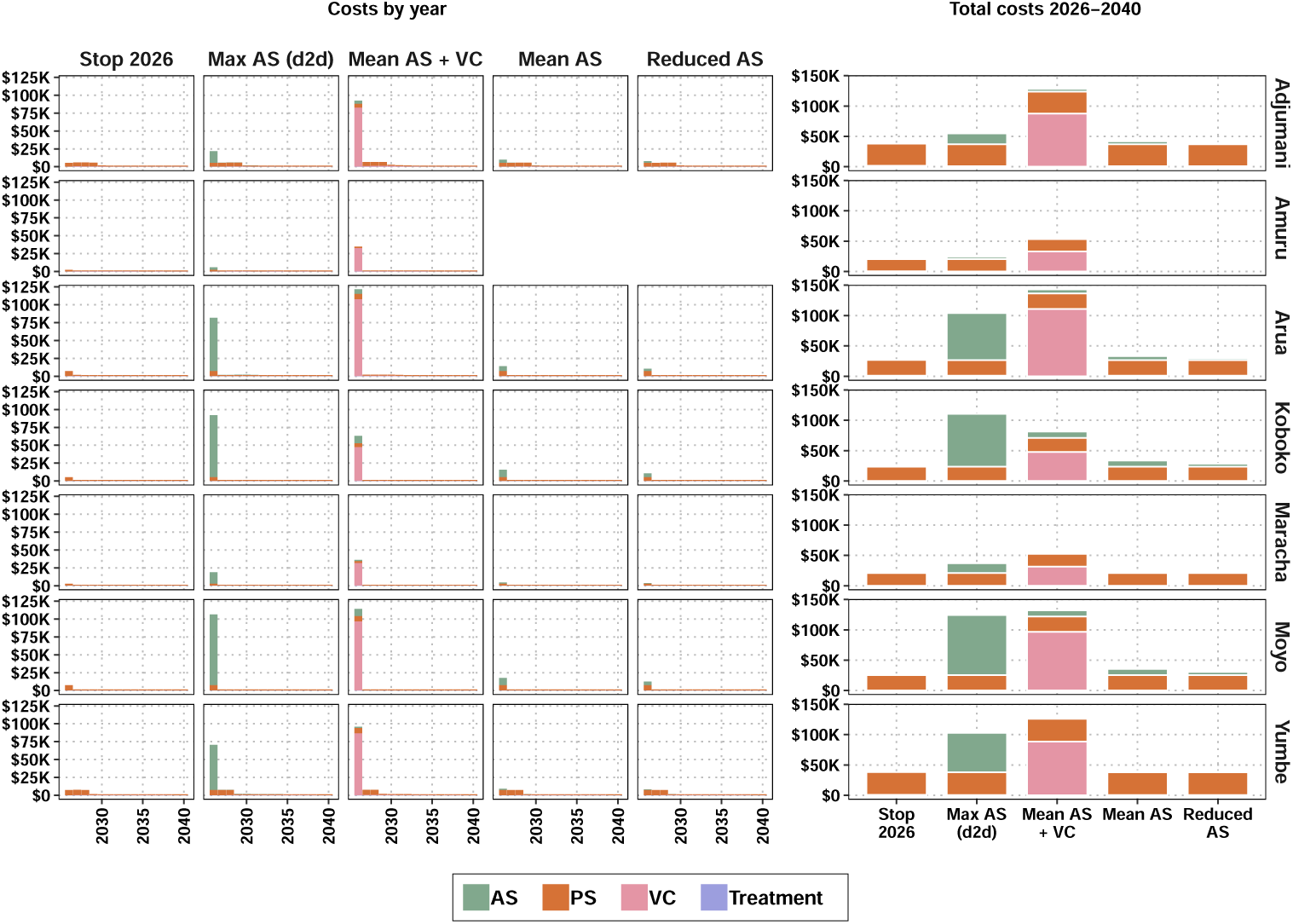
Cost per year and total cost breakdown across all seven former districts of Uganda. Costs shown are economic costs from the programme and donor perspective without discounting (to align more closely with planning). Amuru does not show the strategies *Mean AS* and *Reduced AS* as these are identical to *Stop 2026* because Amuru has had no AS during 2020–2024. AS: active screening, PS: passive screening, VC: vector control

We note that in a very low number of realisations (*<*2% in each district), due to the impact of reactive strategies, it is possible to have fewer DALYs averted by the *Reduced AS* or *Mean AS* strategy compared to the *Stop 2026* strategy (Table 7). The reason that *Stop 2026* strategy may avert more DALYs than more ambitious strategies is that RVC is triggered along with RAS, whereas in strategies without VC where AS has not ceased by 2026, RVC is never implemented. This can have a marked effect on the dynamics and allows extra infections to arise in the strategies without this RVC. This is to be expected since very few infections and cessation make the strategies very similar.

The aggregated results show that, under *Stop 2026* the highest costs will be incurred in 2026, which constitutes over one-fifth of costs over the 15-year time horizon (Table 8 and Figure 6). Costs nationwide would rapidly decrease to about $10,000 per year by 2030.

**Fig. 6:**
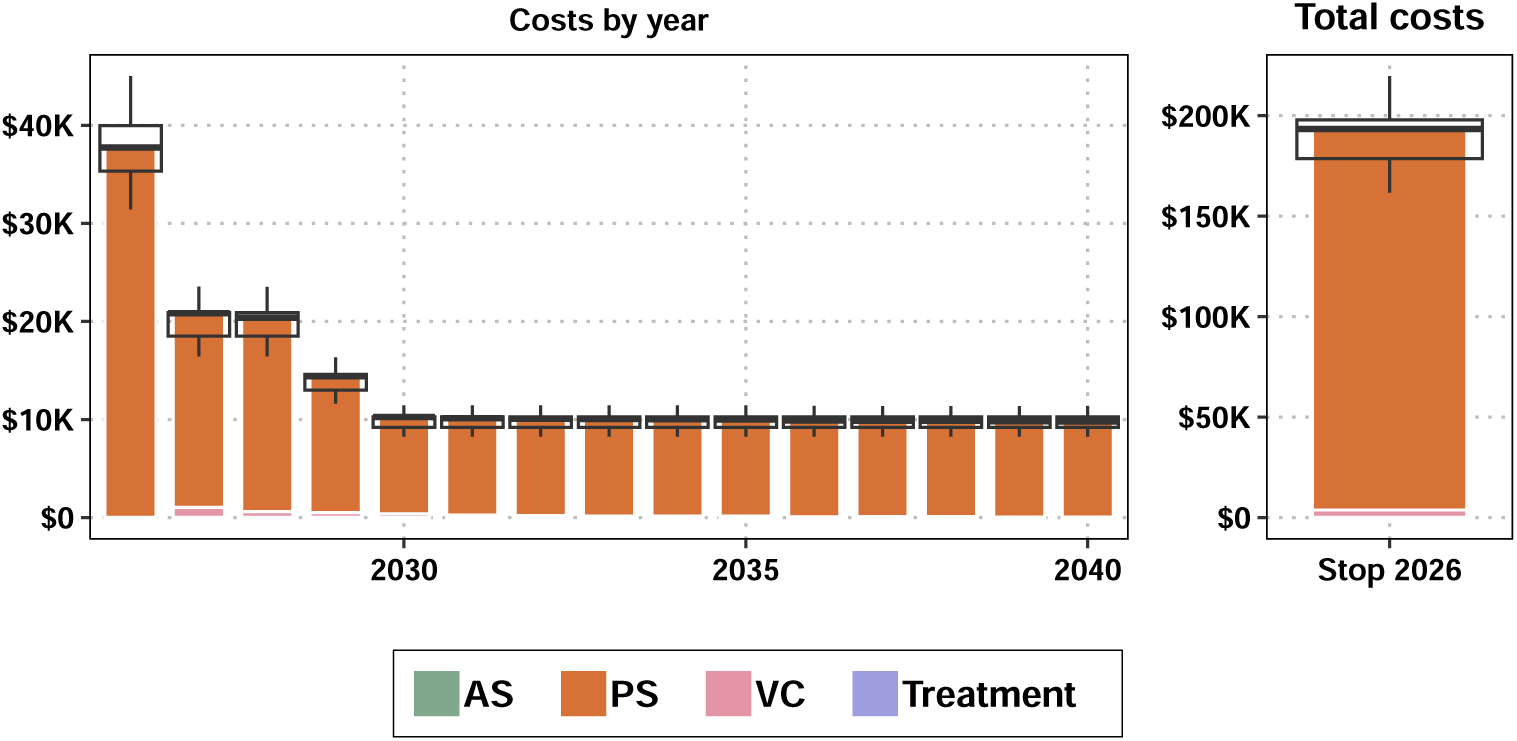
Country-wide costs by year, as well as for the whole period 2026–2040. Only *Stop2026* (the *Status Quo* objective) is shown, as other strategies would not be costreffective at any of the willingness-to-pay (WTP) values considered, nor would they have noticeable disease burden benefit.

**Table 8:** Health and economic outcomes at the national level 2026–2040, showing mean (95% prediction intervals). Only *Stop2026* (the *Status Quo* objective) is shown, as other strategies would not be cost-effective at any of the willingness-to-pay (WTP) values considered, nor would the strategies have a noticeable benefit in terms of disease burden. DALY: disability adjusted life-year.

| Outcome | Year 2026 | 2026–2040 |
| --- | --- | --- |
| Reported Cases | 0 (0-0) | 0 (0-0) |
| Deaths | 0 (0-0) | 0 (0-0) |
| DALYs | 1.54 (0-0.303) | 11.0 (0-0.517) |
| Costs | 37,751 (31,423–45,015) | 193,381 (161,728–219,713) |

When considering the tests needed for screening, no further tests are needed for AS, but slightly fewer than 1000 tests are needed for PS for 2026, followed by a quick decline to 0 by 2030 for PS (Table S34). More information on tests by district are available in the GUI (https://hatmepp.warwick.ac.uk/uganda/v6/).

### Scenario analysis on cross-boundary importation

After matching to case reporting patterns (see the ‘Filtering’ section of the Methods), most of the realisations are close to or have achieved NRI. To focus the analysis on the impact of importation of infections, we further filtered out the results that have not achieved NRI (i.e. possibility of future transmission events, case detections and deaths) in the *Stop 2026* strategy. This second filter only dropped at most 2.16% of the filtered realisations. Table 9 summarises tsetse density at the time of importation, probability of generating non-zero secondary infections, and the median and 95% PIs of time to the LTE when at least one new transmission event happens in the seven districts.

**Table 9:** Summary of case importation simulations. These results show our projections assuming a single importation of an infected stage 1 person in year of importation (*Y_i_*) of 2026 or 2035 and the outcome of this. We show the tsetse density for each district in both years based on the assumed tsetse model bounceback. The middle columns show the probability that the single imported case leads to at least one new infection by district, based on the year of infection importation. The final columns give the predicted time until the last transmission event (LTE) and uncertainty for the simulations, for the simulations where there was at least one new transmission event; 0 indicates that the LTE was in the same year as the importation.

| District | Tsetse density at $Y_i$ as a % of pre-VC population | | Probability of secondary infections | | Median [95% PIs] of time to LTE (years) | |
| --- | --- | --- | --- | --- | --- | --- |
| | $Y_i = 2026$ | $Y_i = 2035$ | $Y_i = 2026$ | $Y_i = 2035$ | $Y_i = 2026$ | $Y_i = 2035$ |
| Adjumani | 92% | 100% | 30% | 39% | 1 [0, 14] | 3 [0, 19] |
| Amuru | 95% | 100% | 29% | 37% | 2 [0, 15] | 2 [0, 16] |
| Arua | 91% | 100% | 32% | 37% | 3 [0, 20] | 3 [0, 19] |
| Koboko | 100% | 100% | 31% | 38% | 3 [0, 24] | 3 [0, 20] |
| Maracha | 100% | 100% | 25% | 36% | 2 [0, 21] | 3 [0, 19] |
| Moyo | 85% | 100% | 30% | 38% | 2 [0, 17] | 2 [0, 18] |
| Yumbe | 95% | 100% | 31% | 40% | 2 [0, 21] | 3 [0, 20] |

We found that, in all districts but Koboko and Maracha, the modelled tsetse population in 2026 was less than the population before VC activities were deployed, whereas by 2035 the modelled tsetse population has resurged to the original population size (see Figure S35 in Supplement 1).

The single stage 1 importation produces at least one secondary infection in 25% to 32% of realisations if the importation happens in 2026, and this increases to 36% to 40% of realisations for the 2035 importation due to a higher vector population and likely scale back in PS simulated following 10 years with no case reporting. In the realisations where secondary cases occur, the median time to the LTE, or to breaking the chain of transmission, is 1–3 years and 2–3 years for the 2026 and 2035 importation scenarios, respectively. With a very small probability (*<* 2.5%), it takes between 14–21 years to reach LTE after importation.

## 4 Discussion

This is the first analysis of the transmission dynamics and the cost-effectiveness of the country-wide elimination of *gambiense* HAT across the seven formerly endemic districts of north-west Uganda, the only country affected by both forms of HAT (*gambiense* and *rhodesiense*) and among a handful of countries struggling with cross-border re-infections. The availability of 25 years of longitudinal case and screening data from the NSSCP of Uganda and the WHO HAT Atlas has enabled us to conduct a robust modelling analysis, and it is noteworthy that such data exist for an neglected tropical disease (NTD). Our modelling analysis finds that transmission has likely halted in all seven districts. Cost-effectiveness finds that all public health activities beyond PS are not economically justified. We expect that economic costs will be less than $250,000 to maintain passive screening until 2040. One of the key requirements of the process of verification of elimination of transmission is to guarantee continued access to diagnosis and treatment post-elimination. A post-verification surveillance strategy integrated within routine health systems has been established to sustain zero-case status [15]. Uganda is one of the first countries to prove that elimination of gHAT can be operationally feasible, despite high prevalence in the 1990s.

Our analysis indicates that a combination of strengthened and more widely available screening in fixed health clinics – enabled by the introduction of serological diagnostics (CATT in 1998 and followed by RDTs in 2013–14) and safer, more accessible treatments for stage 2 disease – alongside VC, have led to the elimination of gHAT transmission. The model indicates that any recent or future case reporting would likely be the result of the disease’s long natural history (historical infections) or importation from South Sudan. However, the sustained reduction in tsetse across endemic districts for many years makes continued transmission less likely.

To our knowledge, only two other transmission modelling studies have been performed, tailored to the Ugandan context; however, these studies used either no or limited case data and cannot readily be used to extrapolate to the present situation nor make a robust quantitative assessment of elimination progress [52, 53]. Davis et al. [52] concluded that sustained tsetse control would be expected to have a large impact on gHAT, in line with many other modelling analyses for gHAT outside of Uganda [9, 19, 54, 55].

Other gHAT modelling studies for north-western Uganda include the ecological capacity to suppress the tsetse population [56], a geospatial model of tsetse abundance to determine where to deploy Tiny Targets based on environmental variables [57], a statistical treatment of VC and cases [30], and the optimal placement of PS health centres to maintain adequate health care access for the population at risk [26]. Vale et al. (2024) used a spatially explicit model to analyse the capacity of Tiny Targets to destabilise and substantially diminish the tsetse population to better control the latter’s population; however, they did not model the transmission of *T.b gambiense*. Lastly, Longbottom et al. (2021) focused on optimal placement of PS diagnostic locations so that the population at risk would have access to testing within a one-hour travel time by motorcycle to a clinic [26].

Moreover, other modelling studies tailored to similar contexts like Mandoul in Chad or low-prevalence regions show the importance of VC on case reduction, even if our current recommendation is to cease VC [9, 19, 55]. Rock et al. showed that in lowrisk locations, although PS is the last intervention to be reduced, it is VC and AS that most likely ensure elimination of transmission by 2030 and which reduce the greatest number of cases across models from three different modelling groups [34]. In Mandoul, which was nearing elimination but had persistent transmission, VC was estimated to have sustainably broken the chain of transmission 15 years ahead of medical-only interventions, although it contributed to only 34% of the case reduction in 2015–19 (see Figure S8, in Rock et al’s 2022 publication) [9, 19]. An additional study in Mandoul by Antillon et al. on the cost-effectiveness of past interventions showed that the addition of VC to 2014 activities was cost-saving, and the addition of both VC and improved PS (what was done) was cost-effective for the Chadian context. The results were robust even if the rollout of fexinidazole, which occured in 2020, had taken place before Chad intensified PS and added VC to its strategy. In short, this analysis confirms previous results showing that VC could play a strong role in achieving elimination, with the cost being either modest or cost-saving [45].

In terms of cost-effectiveness, the only other paper that had treated the total costs of interventions in Uganda came from Sutherland and colleagues, who estimated that the costs of control were $38M spent in 2013–2020, or about $4.75M/year based on costs primarily arising from the DRC and using a transmission model calibrated to a generic “low-risk” setting [58]. In contrast, using locally-sourced information, including the district case data and budgets from the programme, we estimated that the costs were about one-tenth as high. The costs remaining for the period of 2026–2040 will be about $193K ($161K–$220K) in total or about $10,000 by the year 2030, showing how quickly costs may be reduced with elimination campaigns. One more study by Shaw et al. calculated the unit costs for VC, which were input into our analysis, but did not consider the other components of the programme nor its implications on activity efficiencies [59].

Although we did not use any data on infection in animals, the model finding that there is limited evidence of animal transmission aligns with empirical studies which have also failed to find *T. b. gambiense* in livestock despite these animals being an important source of bloodmeals for tsetse [60, 61]. Our model considers a general, non-species-specific population of animals that contribute to transmission. In the absence of animal infection data, when we fit this variant of the model, detection of animal transmission would be dependent upon there being ongoing transmission of gHAT beyond that which can be explained by the human–tsetse–human transmission pathway. For example, prolonged elimination of transmission.

It is, of course, feasible to incorporate available animal infection data into gHAT transmission models. However, an appropriate treatment of these data would require the inclusion of separate transmission paths for each recorded species, as well as the continuing use of a general animal population contributing to transmission (i.e. as we currently have with no surveyed animal populations) to account for all the species that might contribute to transmission that are not being surveyed for gHAT infection. We believe that estimation of the necessary species-specific parameters would pose considerable difficulties, while previous survey results [61] would imply that the impact on the modelling results would likely be minimal.

Our transmission and health economic modelling frameworks can be applied to study gHAT dynamics and analyse the cost-effectiveness of plausible intervention strategies in different endemic settings, as has been done before [44, 45]. Model fitting requires historical case data and local contextual information, such as intervention history, to calibrate local parameters. To study elimination metrics, future intervention strategies (i.e., a combination of AS, PS, and VC) are needed if elimination has not yet been achieved. Local financial and economic costs of operating interventions and treatments should be collected to perform a cost-effectiveness analysis.

### Limitations

The present work is subject to limitations that arise from the structure of the model, the assumptions underpinning it, and the completeness and reliability of the data used.

It is well established that stage 2 patients suffer more severe disease symptoms compared to stage 1 patients. It is, therefore, logical to conclude that stage 2 patients will spend less time conducting activities in tsetse habitats (i.e. fishing, bring animals to water) and consequently receive fewer tsetse bites. In this analysis, we did not consider this behavioural change, i.e. a reduction in transmission probability from stage 2 humans to tsetse, as we found it is not possible to estimate this additional model parameter from the data available. Furthermore, we do not believe having this parameter would have a substantial impact on the results because in Uganda, due to historical VC, has witnessed large declines in transmission regardless of the infectiousness of stage 2 compared to stage 1-infected individuals. However, considering reduced transmission between stage 2 humans and tsetse may be important to consider in higher prevalence locations.

Our assumption that low-and high-risk humans have an equal probability of participating in AS in the *Max AS (d2d)* strategy represents the best-case scenario for door-to-door screening. If we used the same coverage of screening but assumed high-risk people have a lower probability of participating, then case finding would be reduced. If we had data on the age and gender of participants of door-to-door screening compared to traditional screening, we could modify the relative screening probability for different risk groups, however, given our results here are so similar in dynamics for *Max AS (d2d)* compared to *Mean AS* this additional strategy variant was not considered to be valuable to simulate.

The transmission model allows for an improvement in the rate of case detection from PS over time. This is modelled via a logistic curve, requiring the amplitude, mid-point and steepness of the change to be either estimated from the data or assumed to be known. We estimated that the median mid-point for the PS improvement year had a range between 2003 and 2011 across the districts; while the mid-point of the magnitude of the improvement in the stage 2 detection rate was 0.18–8.07 times the stage 2 detection rate estimated in 1998 across districts. We know further improvements were made to the PS system from 2013–4 under the Trypa-NO! project [25], however, it is challenging to detect differences resulting from these changes given the low level of case reporting from that year, and therefore we did not directly include this in the model. We believe this is not a reflection on the quality of the clinics to detect cases, but rather a reflection that the situation is close to the zero infection target and therefore the metric of screening quality cannot be solely cases detected.

Tsetse capture data were only available in three of the seven districts at the time of this analysis (Arua, Koboko, and Maracha), so we assumed a VC effectiveness of 80% or 90% in the other districts. As this is broadly in line with a tsetse habitat suitability analysis performed by a spatiotemporal model recently [39], we believe this is a sensible assumption, and any deviations would have a limited impact on our fits to the historical epidemiological data. Despite the lack of detail, sustained high tsetse reductions of even 60% would be expected to have a big impact on the transmission of gHAT infection and subsequent case reporting compared to medical interventions alone [33]. In the future, we could refine our tsetse model using more entomological data, however, we believe it will have little quantitative impact on our parameter estimates or projections.

Other aspects of VC modelling could be refined; in particular, how we handle case coverage in each district: As the amount and location of Tiny Targets changed over time, using time varying case coverage, rather than one fixed value, would give us better estimates of tsetse populations and the transmission between people and vectors, however, we do not think that the improvement in our tsetse density estimation will noticeably change our findings because of the low number of remaining infected people when VC started in Uganda. Other studies [39, 62] used a different type of modelling approach to evaluate tsetse habitat suitability and abundance using remote sensing data (e.g. Normalised Difference Vegetation Index (NDVI)) in the same region of Northern Uganda. It could be possible to use the results of this spatiotemporal, geostatistical modelling coupled with fitting our tsetse model to Ugandan trap data to have a more accurate representation of tsetse dynamics in our full gHAT transmission model. It is the overall tsetse density trends over time which would most benefit from this additional information. However, since the absolute number of tsetse is not possible to measure through geostatistical modelling nor trapping, so the tsetse to human density would still need to be estimated as a parameter in our transmission model fitting.

The political instability in South Sudan has resulted in the immigration of people, and even animals, from South Sudan to northern Uganda, primarily into the same districts which have reported gHAT [5, 6]. Due to ongoing transmission of gHAT in South Sudan and limited interventions, there have been imported cases detected in Uganda. However, as a scenario analysis, our results in Table 9 show that the likelihood of secondary infections is low, and that the years until transmission is interrupted again are few. It is worth noting that our scenario analysis shows the outcome in the case of a single stage 1 importation, but Uganda receives numerous South Sudanese refugees who cross the border repeatedly for the purpose of trade. Since we cannot know the prevalence of the disease in the places where the refugees come from or visit in South Sudan, our scenario analysis is merely illustrative. In order to tackle the realistic transboundary threats, regular coordination meetings and information sharing take place between HAT control stakeholders in Uganda and South Sudan as part of the gHAT elimination strategy. These measures ensure sustained surveillance and responsiveness.

Likewise, we did not consider the impact of importations of infected animals and/or tsetse on gHAT transmission in humans, but we believe the effect would be small or non-existent. Our ensemble model showed weak support for animal contribution towards gHAT transmission in the seven Ugandan districts modelled in this analysis. As for vector movement, we considered that the “local” remaining tsetse population can resurge following cessation of VC and ignored the possibility of repopulation from nearby locations. While this choice may impact the bounce-back speed, different speeds are unlikely to impact outcomes when we have no human case importation. Our human importation scenario analysis shows that a bigger vector population could lead to a higher chance of secondary infections, and therefore, breaking the chain of transmission will take longer. Although a total re-emergence rate based on tsetse capture data after VC stops could be incorporated, which would more accurately assess the risk of imported human cases, we expect that such results would be interesting but not operationally consequential for the national programme.

## 5 Conclusion

In this analysis, we used three elimination metrics to assess the past and future progress of gHAT transmission. Our modelling results show that all seven districts have a very high chance of having achieved the last transmission event (LTE) already; however, it is possible that there could still be some case reporting after 2025. The model suggests it could take up to 3 years on average to have no remaining infections (NRI) after the LTE at both district and national levels.

As modelling suggested that all districts have already achieved the LTE, the recommended minimal-intervention strategy for achieving NRI is the *Stop 2026* strategy. This consists of continuing PS and only reactive vertical interventions (RS and RVC) if passive cases are found. No resurgence is predicted to happen in our simulations for any district over the next three decades in the absence of parasite importations from other countries, such as South Sudan. Even with sporadic single case importations, the risk of gHAT resurgence in Uganda is extremely low. However, there is a moderate probability of a few new Ugandan infections occurring before transmission stops again, particularly if the tsetse population resurges to pre-vector control levels. Our cost-effectiveness analysis also shows that we have very high confidence that the *Stop 2026* strategy is cost-effective under short- and medium-term projections, and the total economic cost for this strategy is projected to be $193K ($161K–$220K) between 2026–2040.

## Supporting information

Supplement 3: PRIME-NTD Criteria

Supplement 4: CHEER checklist

Supplement 2: Parameter Glossary

Supplement 1: Supplementary Methods and Supplementary Results

## Data Availability

Data cannot be shared publicly because they were aggregated from the World Health Organization’s HAT Atlas which is under the stewardship of the WHO. Data are available from the WHO (contact) for researchers who meet the criteria for access. Model code and outputs produced from this study are available through Open Science Framework.

https://osf.io/kj82q/?view_only=6f36f27746044675805033ef1cd7d02c

## Supplementary Information

Supplement 1: Supplementary Methods and Supplementary Results. Supplement 2: Parameter Glossary. Supplement 3: PRIME-NTD Criteria. Supplement 4: CHEERS checklists.

## Acknowledgements

The authors thank the National Sleeping Sickness Control Programme of Uganda for original data collection and WHO for data access (in the framework of the WHO HAT Atlas [2]).

## Declarations

### Ethics approval and consent to participate

Ethics approval was granted by the University of Warwick Biomedical and Scientific Research Ethics Committee (application numbers BSREC 80/21-22 and 81/21-22). All methods were carried out in accordance with the principles of the Declaration of Helsinki. This study used previously collected Ugandan human African trypanosomiasis (HAT) data, provided through the framework of the WHO HAT Atlas [2] and the Trypa-NO! project, for secondary modelling analysis. No new data were collected as part of this study.

#### Consent to Participate declaration

Not applicable.

### Consent for publication

Not applicable.

### Data availability

Data cannot be shared publicly because they were aggregated from the World Health Organization’s HAT Atlas which is under the stewardship of the WHO. Data are available from the WHO (contact) for researchers who meet the criteria for access.

### Code availability

Model code and outputs produced from this study are available through OpenScienceFramework.

### Author contributions

– Conceptualisation CH KSR

– Methodology CH REC SAS MA BH RS

– Software PEB

– Validation CH

– Formal Analysis CH REC MA BH

– Investigation CH MA

– Data Curation REC CH PRB MA CW ST AH

– Writing - Original Draft CH KSR

– Writing Review and editing all authors

– Visualisation CH MA

– Supervision KSR EHC FT

– Project administration EHC

– Funding acquisition KSR

