## Supplement 3: PRIME-NTD Criteria for "Transmission and cost-effectiveness modelling to estimate the progress towards elimination and future strategy optimisation for *gambiense* human African trypanosomiasis in Uganda"

### **NTD Prime Criteria**:
### Transmission modelling to estimate the progress towards elimination and future predictions for *gambiense* human African trypanosomiasis in Uganda

Ching-I Huang[1,2*], Marina Antillon[1,2,3,4], Ronald E Crump[1,2◑], Samuel A Sutherland[2,4◑], Paul R Bessell[5,6], Albert Mugenyi[7], Richard Selby[8], Paul E Brown[1,2], Brady Hooley[3], Andrew Hope[8], Sophie Dunkley[8], Rob Sunnucks[1,2], Steve J Torr[8], Fabrizio Tediosi[3], Joseph Ndung'u[6], Emily H Crowley[1,2], Eric Kidega[9], Charles Wamboga[9], and Kat S Rock[1,2]

[1]Zeeman Institute for System Biology and Infectious Disease Epidemiology Research, The University of Warwick, Coventry, U.K.
[2]Mathematics Institute, The University of Warwick, Coventry, U.K.
[3]Warwick Medical School, The University of Warwick, Coventry, U.K.
[4]Swiss Tropical and Public Health Institute, Basel, Switzerland
[5]Epi Interventions Ltd, Edinburgh, U.K.
[6]Foundation for Innovative New Diagnostics, Geneva, Switzerland
[7]Ministry of Agriculture Animal Industry and Fisheries, Kampala, Uganda
[8]Department of Vector Biology, Liverpool School of Tropical Medicine, Liverpool, U.K.
[9]Ministry of Health, Kampala, Uganda

◑These authors contributed equally to this work.

Good modelling practices should include the Policy-Relevant Items for Reporting Models in Epidemiology of Neglected Tropical Diseases (PRIME-NTD) table to address the five key principles of communication, quality and relevance of analyses.

| What has been done to satisfy the principle? | Where in the manuscript is this described? |
|---|---|
| **1. Stakeholder engagement**<br>This modelling study has been conducted in conjunction with a range of partners, including the national sleeping sickness control programme in Uganda – co-authors A Mugenyi, C Wamboga, and E Kidega, experts in tsetse control – co-authors P R Bessell, R Selby, A Hope, S Dunkley, and S J Torr, as well as an expert in diagnostics – co-author J Ndung'u. Numerous discussions took place to ensure the modellers produced meaningful outputs with policy relevance. This article reflects a small piece in an ongoing body of work between the modelling group (HAT MEPP) and our partners mentioned above. We have had regular meetings and occasional modelling workshops over several years when we discuss ongoing and planned strategies, historical data, and new data analyses. A draft version of this article was also shared with the WHO HAT team who reviewed all of our modelling work which utilises the WHO HAT Atlas data. | Authorship list and acknowledgements |
| **2. Complete model documentation**<br>Full modelling code and documentation are available through OpenScienceFramework (OSF). The epidemiological model was fully described in the main text and this supplementary information. Previous versions of the model are fully described elsewhere [**? ? ? ? ? ? ? ?** ]. | Methods in the main text, Supplement 1, and our OSF. |
| **3. Complete description of data used**<br>The original human and tsetse data and how we aggregated the human data for fitting were described in the main text and this supplementary information and can be viewed through the corresponding graphical user interface (GUI). | Methods section in the main text, Human case and active screening data & Tsetse data in Sections **??** and **??** in Supplement 1, and our GUI. |
| **4. Communicating uncertainty**<br>*Structural uncertainty*: We considered 8 model variants to incorporate the uncertainty of infection risk and participation in active screenings in humans. The model evidence, or marginal likelihood, for each model, was used to create our ensemble model for projections. | See Methods section as well as Sections **??-??** of Supplement 1. |
| *Parameter uncertainty*: We used estimates from joint ensemble posterior distributions of fitted parameters generated from the model fitting process. | Model fitting procedure in Section **??** of Supplement 1. |
| *Prediction uncertainty*: We represent uncertainty in our results by providing box and whisker plots for predicted outcomes (median, 50% and 95% credible intervals) or probabilities of outcomes. | Figure 2–4 in the main text and additional results from the transmission model in Section **??** of Supplement 1. |
| **5. Testable model outcomes**<br>Epidemiological model outputs are routinely reported metrics of the disease course: detected active and passive case detections. Therefore, these predictions can be compared to future data as long as the data is put into context alongside measures of active screening coverage and the actual coverage of vector control. Our results presented in this article and our open-source code mean this validation exercise should be straightforward to conduct. | Epidemiological projections in 2025–2030 under five strategies are shown in Figures **??–??** for seven endemic districts in Uganda. Full epidemiological outcomes for each year until 2054 can be viewed at our GUI |
