## Supplement 4: CHEER checklist for "Transmission and cost-effectiveness modelling to estimate the progress towards elimination and future strategy optimisation for *gambiense* human African trypanosomiasis in Uganda"

| Section/item | Item No | Recommendation | Reported on page no, line no |
|---|---|---|---|
| **Title** | | | |
| Title | 1 | Identify the study as an economic evaluation or use more specific terms such as "cost-effectiveness analysis", and describe the interventions compared. | Title & Table 2 |
| **Abstract** | | | |
| Abstract | 2 | Provide a structured summary of objectives, perspective, setting, methods (including study design and inputs), results (including base case and uncertainty analyses), and conclusions. | Abstract |
| **Introduction** | | | |
| Background and objectives | 3 | Provide an explicit statement of the broader context for the study. Present the study question and its relevance for health policy or practice decisions. | Introduction section, in particular the last 4 paragraphs and the 'Historical interventions' of the Methods section. |
| **Methods** | | | |
| Health economic analysis plan | 4 | Indicate whether a health economic analysis plan was developed and where available. | No previous protocol was published. |

*(continued)*

| Section/item | Item No | Recommendation | Reported on page no, line no |
|---|---|---|---|
| Study population | 5 | Describe characteristics of the study population (such as age range, demographics, socioeconomic, or clinical characteristics). | The population is described in the Methods section 'Demographic, screening and case data', Section **??** and Table **??** of Supplement 1, and more specific inputs for each district in Table **??** of Supplement 1. |
| Settings and location | 6 | Provide relevant contextual information that may influence findings. | The location is described in the Methods sections 'Study area' and 'Historical interventions' as well as Section **??** of Supplement 1. |
| Comparators | 7 | Describe the interventions or strategies being compared and state why they were chosen. | Methods section 'Projections' and in Table 2. |
| Perspective | 8 | State the perspective(s) adopted by the study and the rationale. | Fourth subsection of the methods, "Costs and cost-effectiveness analysis", second paragraph. |
| Time horizon | 9 | State the time horizon over and why the horizon is appropriate. | Fourth subsection of the methods, "Costs and cost-effectiveness analysis", first paragraph. |
| Discount rate | 10 | Report the discount rate(s) and reason chosen. | Fourth subsection of the methods, "Costs and cost-effectiveness analysis", second paragraph. 3% is the recommended rate by WHO-CHOICE and the Gates Reference Case. |
| Selection of outcomes | 11 | Describe what outcomes were used as the measure(s) of benefit(s) and harm(s). | Fifth paragraph of the methods section called "Costs and cost-effectiveness analysis". Moreover, Sections **??** of Supplement 1 contain detailed explanations of how DALYs are calculated and how the natural history of HAT was considered. |
| Measurement of outcomes | 12 | Describe how outcomes used to capture benefit(s) and harm(s) were measured. | Fifth paragraph of the methods section called "Costs and cost-effectiveness analysis". Moreover, Section **??** of Supplement 1 contains details on how life lived with gHAT was weighted, and treatment outcomes were described in Section **??** of Supplement 1. |
| Valuation of outcomes | 13 | Describe the population and methods used to measure and value outcomes. | DALYs were used, not preference-based outcomes. |
| Measurement and valuation of resources and costs | 14 | Describe how costs were valued. | The construction of programme costs are detailed in Section **??** in Supplement 1. As can be seen, in a publication of this scope, detailing the cost inputs in the main body would be unfeasible. However, the resulting expected costs are in Figure 5 and 6 of the main body of the paper. In the `GUI`, one may find the costs broken down by activity by health zone, coordination, and the country for under a variety of sensitivity analyses. |

*(continued)*

| Section/item | Item No | Recommendation | Reported on page no, line no |
|---|---|---|---|
| Currency, price date, and conversion | 15 | Report the dates of the estimated resource quantities and unit costs, plus the currency and year of conversion. | Second paragraph of the methods, "Costs and cost-effectiveness analysis". Our general approach for costing and conversions across years is in the section "Principles for parameterisation" in Section **??** in Supplement 2, followed by the specific choices for each parameter. |
| Rationale and description of model | 16 | If modelling is used, describe in detail and why used. Report if the model is publicly available and where it can be accessed. | The decision analytic model is illustrated in detail in Supplementary Figure **??**, but the components models feeding into the decision analytic model are described as follows: the dynamic transmission (SEIRS) model is described briefly in the third Methods section "Transmission model" and Section **??** in Supplement 1 and pictured in Supplementary Figure **??**, and the treatment model is described briefly in Section **??** in Supplement 1 and shown in Supplementary Figure **??**. |
| Analytics & assumptions | 17 | Describe any methods for analysing or statistically transforming data, any extrapolation methods, and approaches for validating any model used. | The transmission, treatment, and health economic models are discussed in Sections **??** and **??** of Supplement 1. The treatment model and its parameters are described in detail in Sections **??** in Supplement 1. Assumptions are described in detail in Supplement 1, Section **??** and the Parameter Glossary. |
| Characterising heterogeneity | 18 | Describe any methods used for estimating how the results of the study vary for subgroups. | Heterogeneity across districts was characterised by the clinics, populations screened, and vector control needs, as described in Table **??** in Supplement 1. |
| Characterising distributional effects | 19 | Describe how impacts are distributed across different individuals or adjustments made to reflect priority populations. | Because there were no subgroups, there were no distributional effects. The differential impacts of treatment on poorer or less poor individuals were beyond the scope of this paper. |
| Characterising uncertainty | 20 | Describe methods to characterise any sources of uncertainty in the analysis. | The epidemiological parameters are the posterior distributions of a model fitted to time-series data, and full details are available in the Section **??** of Supplement 1. For the parameters to model health outcomes and costs, assumptions and estimates were parameterised according to conventions in the economic evaluation literature [**?** ], sampling from large distributions for aspects for which we knew very little. Health outcome and cost-effectiveness parameters: see Table **??**-**??** in Supplement 1 and Supplement 2: Parameter Glossary. |

*(continued)*

| Section/item | Item No | Recommendation | Reported on page no, line no |
|---|---|---|---|
| Approach to engagement with patients and others affected by the study | 21 | Describe any approaches to engage patients or service recipients, the general public, communities, or stakeholders (such as clinicians or payers) in the others affected by the study. | Strategy components were determined along with members of the NSSCP, Drs Charles Wamboga, Eric Kidega and Albert Mugenyi (co-authors). Implementation of simulations was aided by discussion with collaborators at LSTM, Drs Andrew Hope, Richard Selby, Sophie Dunkley and Prof Steve Torr (co-authors) who have run vector control field operations in Uganda since 2013. Input on passive and active strategy structures was provided by Dr Paul Bessell and Prof Joseph Ndung'u of FIND. |
| **Results** | | | |
| Study parameters | 22 | Report the values, ranges, references, and, if used, probability distributions for all parameters. Report reasons or sources for distributions used to represent uncertainty where appropriate. Providing a table to show the input values is strongly recommended. | Table **??** in Supplement 1 and described in more detail in Supplement 2: Parameter Glossary. |
| Incremental costs and outcomes | 23 | For each intervention, report mean values for the main categories of estimated costs and outcomes of interest, as well as mean differences between the comparator groups. If applicable, report incremental cost-effectiveness ratios. | Table 7 of the Results section and the detailed results for each district can be found in Tables **??**–**??** in Supplement 1. |
| Effect of uncertainty | 24 | Model-based economic evaluation: Describe the effects on the results of uncertainty for all input parameters, and uncertainty related to the structure of the model and assumptions. | For four sample health zones, the effect of uncertainty on the cost-effectiveness are shown in Tables **??**-**??** in Supplement 1 and in the `GUI`. The effect of uncertainty on the Elimination of Transmission goal is shown in Figure 3. The interpretation of the uncertainty is in the Results section. |
| Effect of engagement with patients and others affected by the study | 25 | Report on any difference patient/service recipient, the general public, community, or stakeholder involvement made to the approach or findings of the study | Our engagement with the stakeholders (co-authors) was iterative throughout the process. |
| **Discussion** | | | |
| Study findings, limitations, generalisability, and current knowledge | 26 | Summarise key study findings and describe how they support the conclusions reached. Discuss limitations and the generalisability of the findings and how the findings fit with current knowledge. | Discussion. |
| **Other relevant information** | | | |

*(continued)*

| Section/item | Item No | Recommendation | Reported on page no, line no |
|---|---|---|---|
| Source of funding | 27 | Describe how the study was funded and the role of the funder in the identification, design, conduct, and reporting of the analysis. Describe other non-monetary sources of support. | Funding statement. |
| Conflicts of interest | 28 | Describe any potential for conflict of interest of study contributors in accordance with journal policy. In the absence of a journal policy, we recommend authors comply with the International Committee of Medical Journal Editors recommendations. | Conflict of interest statement. |
