## Supplement 2: Parameter Glossary for "Transmission and cost-effectiveness modelling to estimate the progress towards elimination and future strategy optimisation for *gambiense* human African trypanosomiasis in Uganda"

### G1 Principles for parameterization

Please note that this Section contains substantial recycled text from the authors' previous publications [G1, G2, G3]. Parameter values and citations have been substantially updated to match information and data on Uganda where appropriate.

Below are the guidelines our team follows to parameterize the treatment and intervention model. The rationale behind the choices in the tables that describe parameters is more straightforward with these guidelines in mind.

- **Transferability of costs across time** Costs from the literature are updated to 2023 USD values by converting to local currency units in the year of the study in the literature, inflated to 2023 values using the consumer price index (CPI) of the country, and then converted to USD using the exchange rate in 2023. It should be noted that the 2003 WHO Guide to Cost-effectiveness recommends that the GDP inflator be used (see 3.2.6 Transferability of costs across time, page 43) but we found that the data on this measure (from the World Bank) were sometimes sparse so we relied on the consumer price index instead (`'NY.GDP.DEFL.KD.ZG'` in the World Bank Development Indicator Database) [G4].
- **Transferability of costs across settings** To 'borrow' data from other countries, we follow the 2003 WHO Guide to Cost-effectiveness recommendations in section 3.2.7 Transferability of costs across settings) [G4]. For non-traded items (i.e. nurse and doctor time) we convert USD or LCU prices into PPP (international dollars) values in the year of the cost study and then turn the value in international dollars to local currency (still in the year of the study) of the country where a cost estimate is needed. Then, we use the CPI to inflate costs to 2022 levels and then use the exchange rate with USD to get 2023 USD values.
- **Combining multiple sources of information** Values from different publications are combined using meta-analytic methods.
- **Choice of probability distributions** Costs and ratios were modelled via gamma distributions and proportions or probability were modelled with beta distributions. These distributions were parameterized using the method of moments (see Briggs 2006 [G5], Chapter 4).
- **Missing information on uncertainty: Gamma distributions.**

1. Option A: Whenever uncertainty was missing for a cost or a ratio in the literature, we assigned a gamma distribution for the parameter that would yield credible intervals between half and double the estimate.
2. Option B: If at least 2 studies listed a cost, then we take the range of the costs to parameterize a gamma distribution in which the range matched the 95 percent confidence interval (e.g. the 2.5th and 97.5th percentile). In these cases, we use a method to parameterize gamma distributions using quantiles rather than using the mean and standard error of a sample (e.g. "method of moments") [G6].

- **Missing information on uncertainty: Beta distributions.**

1. Option A: Usually modelled assuming that 100 trials were observed with the **proportion_estimate x 100** as the alpha parameter and **(1-proportion_estimate) x 100** as a beta parameter.
2. Option B: If at least 2 studies listed a probability or a proportion, then we take the range of the costs to parameterize a beta distribution by assuming the range matches the 95 percent confidence interval (the 2.5th and 97.5th percentile). We use a method to parameterize Beta distributions using quantiles rather than the mean and standard error of a sample (method of moments) [G7].

### G2 Organisation of parameters

In the code repository, found in `https://osf.io/kj82q/`, the parameters are in an Excel sheet.

Additionally, a list named epi_output (read in from Matlab output) holds the output from the dynamic model: stage 1 and stage 2 cases detected by passive and active screening, as well as person-time spent in stage 1 and 2 before detection.

#### G2.1 Summary of health parameters

Below are all the parameters that model health outcomes, as well as a summary of their characteristics. An extended discussion of our choices is featured in the sections annotated in the table.

Table G1: Health parameters

| Variable description | Variable name | Statistical Distribution | Descriptive Summary | Notes |
|---|---|---|---|---|
| **Screening** | | | | |
| PS: RDTs per district (for all facilities) | ps_rdt | Fixed value | Varies by district | See section G3.1.1 |
| PS: number of facilities (RDT only & confirmation) | ps_fac_rdt & ps_fac_conf | Fixed value | Vary by district | See section G3.1.2 |
| AS: coverage | as_redux, as_mean, as_max | Fixed value | Vary by district | See section G3.1.3 |
| RDT algorithm: diagnostic sensitivity | dx_sens_rdt | Beta(230, 1) | 0.996 (0.984, 1.00) | See section G3.1.4 |
| CATT algorithm: wastage during AS | dx_wastage_catt_as | Beta(8, 92) | 0.080 (0.036, 0.140) | See section G3.1.5 |
| RDT algorithm: wastage during PS | dx_wastage_rdt_ps | Beta(1, 99) | 0.010 (<0.001, 0.037) | See section G3.1.6 |
| **Treatment** | | | | |
| Prop. of cases age<6 | treat_prob_under6yo | Beta(153, 2428) | 0.059 (0.051, 0.069) | See section G3.2.1 |
| Prop. of cases weight<35 kg among age>6 | treat_prob_under35kg | Beta(8.30, 360) | 0.023 (0.010, 0.040) | See section G3.2.2 |
| Prop. of S2 cases that are severe | prob_late_stage2 | Beta(76.9, 44.9) | 0.634 (0.546, 0.713) | See section G3.2.3 |
| Duration, treatment: pentamidine (days) | treat_duration_penta | Fixed value | 7 | See section G3.2.4 |
| Duration, treatment: NECT (days) | treat_duration_nect | Fixed value | 10 | See section G3.2.5 |
| Duration, treatment: fexinidazole (days) | treat_duration_fexi | Fixed value | 10 | See section G3.2.6 |
| Pr. of relapse (treatment failure): pentamidine | treat_prob_failure_pent_s1 | Beta(50.3, 665) | 0.070 (0.053, 0.090) | See section G3.2.7 |
| Pr. of relapse (treatment failure): NECT | treat_prob_failure_nect_s2 | Beta(15.9, 379) | 0.040 (0.023, 0.062) | See section G3.2.8 |
| Pr. of relapse: fexinidazole | treat_prob_failure_fexi | Beta(9.49, 497) | 0.018 (0.009, 0.032) | See section G3.2.9 |
| Pr. of SAE: pentamidine treatment | treat-prob-sae-pent-s1 | Beta(1.43, 551) | 0.003 (<0.001, 0.008) | See section G3.2.10 |
| Pr. of SAE: NECT treatment | treat_prob_sae_nect_s2 | Beta(40.9, 368) | 0.100 (0.073, 0.131) | See section G3.2.11 |
| Pr. of SAE: fexinidazole treatment | treat_prob_sae_fexi | Beta(3, 261) | 0.011 (0.002, 0.027) | See section G3.2.12 |
| Duration, SAE (days) | treat_duration_sae | Gamma(1.22, 2.38) | 2.90 (0.130, 9.85) | See section G3.2.13 |
| **Life-years lost (DALY)** | | | | |
| Age of death from infection | age_of_death | Gamma(148, 0.182) | 26.6 (22.4, 31.1) | See section G3.3.1 |
| Life expectancy at age of death | life_expectancy | Interpolated | Varies by age, 45.4 [41.4–49.9] | See section G3.3.2 |
| Disability weights: S1 disease | disability_weighting_s1 | Beta(23.0, 147) | 0.135 (0.088, 0.190) | See section G3.3.3 |
| Disability weights: S2 disease | disability_weighting_s2 | Beta(18.4, 15.6) | 0.541 (0.375, 0.703) | See section G3.3.4 |
| Disability weights: SAE | disability_weighting_sae | Uniform(0.037, 0.114) | 0.076 (0.039, 0.112) | See section G3.3.5 |
| **Vector control** | | | | |
| Area ($km^2$) for VC | vc_area | Fixed | Varies by district | See section G3.4.1 |
| Area ($km^2$) for reactive VC | vc_area | Fixed | 50 | See section G3.4.1 |
| Target density for VC | vc_target_density | Fixed | Varies by district | See section G3.4.2 |
| Target density for RVC | vc_target_density | Fixed | 8.5 | See section G3.4.2 |
| Replacement rate of targets per year | vc_deployments_yr | Fixed value | 2 | See section G3.4.3 |

#### G2.2 Summary of cost parameters

Below are all the cost parameters and a summary of their characteristics in three tables for screening, treatment, and vector control costs. An extended discussion of our choices is featured in the sections annotated in the tables.

Table G2: Cost parameters

| Variable description | Variable name | Statistical Distribution | Descriptive Summary | Notes |
|---|---|---|---|---|
| **Screening** | | | | |
| AS: capital & management cost per person screened | `as_cost_team_cap_mgmt` | Gamma(245, 0.004) | $0.99 (0.88, 1.13) | See section G4.1.1 |
| RS: capital & management costs per person screened | `rs_cost_team_cap_mgmt` | Gamma(186, 0.009) | $1.74 (1.50, 2.00) | See section G4.1.2 |
| AS: pre-mission communications & awareness per person screened | `as_cost_comms` | Gamma(8.48, 0.042) | $0.36 (0.16, 0.63) | See section G4.1.3 |
| RS: pre-mission communications & awareness per person screened | `rs_cost_comms` | Gamma(8.48, 0.050) | $0.42 (0.19, 0.75) | See section G4.1.4 |
| CATT algorithm: cost per test | `dx_cost_catt` | Gamma(1139, 0.0007) | $0.79 (0.74, 0.83) | See section G4.1.5 |
| RDT algorithm: cost per test | `dx_cost_rdt` | Gamma(1139, 0.002205) | $2.51 (2.37, 2.66) | See section G4.1.6 |
| PS: capital & management costs of a facility (RDT-only) | `ps_cost_facility_cap_mgmt_rdt` | Gamma(25.3, 13.5) | $341 (221, 486) | See section G4.1.7 |
| PS: capital & management costs of a facility (LAMP & confirmation centers) | `ps_cost_facility_cap_mgmt` | Gamma(137, 8.53) | $1166 (979, 1370) | See section G4.1.8 |
| PS: communications & awareness | `ps_cost_comms` | Gamma(8.48, 33.3) | $282 (126, 502) | See section G4.1.9 |
| National management cost (ISSEP mark-up) | `program_markup` | Fixed value | 0.100 | See section G4.1.10 |
| **Treatment** | | | | |
| Hospital stay: cost per day | `treat_cost_ip_day` | Gamma(5.39, 0.86) | $4.62 (1.58, 9.25) | See section G4.2.1 |
| Outpatient consult: cost | `treat_cost_op_visit` | Gamma(2.48, 0.42) | $1.03 (0.17, 2.66) | See section G4.2.2 |
| Course of pentamidine: cost | `rx_cost_pentamidine` | Fixed value | $54.00 | See section G4.2.3 |
| Course of NECT: cost | `rx_cost_nect` | Fixed value | $460.00 | See section G4.2.4 |
| Course of fexinidazole: cost | `rx_cost_fexinidazole` | Fixed value | $220.00 | See section G4.2.5 |
| Drug delivery mark-up | `rx_delivery_markup` | Beta(45, 55) | 0.450 (0.354, 0.548) | See section G4.2.6 |
| **Vector control** | | | | |
| Operational cost per $km^2$ protected | `vc_cost_management` | Gamma(8.48, 5.25) | 44.50 (19.77, 79.08) | See section G4.3.1 |
| Cost per target deployed | `vc_cost_target` | Gamma(8.48, 0.644) | $5.46 (2.43, 9.70) | See section G4.3.2 |

### G3 Health parameters

#### G3.1 Screening parameters

##### G3.1.1 PS: RDTs per district

↩ Return to the Summary of Health Outcome Parameters.

- Name in the code: `ps_rdt`
- Source: ISSEP 2023
- Country of estimate: Uganda
- Statistical distribution and parameters: Specific per district, see Table **??**.
- Summary statistics (mean and 95% CI or fixed value): Varies by coordination, see Table **??**.

**Notes**

The parameters related to passive screening (PS) coverage, clinics are based on the data from ISSEP 2023 below.

| Item | Adjumani | Amuru | Arua | Koboko | Maracha | Moyo | Yumbe |
|---|---|---|---|---|---|---|---|
| Number of basic test centres (RDT only) in 2023 | 6 | 1 | 10 | 6 | 3 | 4 | 9 |
| Number of test centres with RDT and microscopy | 1 | 1 | 1 | 1 | 1 | 3 | 1 |
| Number of enhanced test centres with RDT, microscopy, and LAMP | 1 | 0 | 1 | 0 | 0 | 1 | 1 |
| Number of RDTs done in 2023 | 83 | 41 | 165 | 366 | 26 | 98 | 259 |
| Number of positive RDTs at facilities with parasitology | 2 | 0 | 4 | 3 | 0 | 1 | 2 |
| Number of positive RDTs across RDT-only centres in each district | 0 | 0 | 1 | 5 | 0 | 0 | 3 |

Table G3: Passive screening resource use in 2023. Per district values were provided by the national programme.

Data came from the records in 2023, shown in Table **??**. We assume that 26 RDTs per year are performed when PS scales back to 1 clinic per district, which is the minimum.

###### G3.1.2 PS: number of facilities

↩ Return to the Summary of Health Outcome Parameters.

- Name in the code: ps_fac_rdt & ps_fac_conf
- Source: ISSEP 2023
- Country of estimate: Uganda
- Statistical distribution and parameters: Specific per district, see Table **??**.
- Summary statistics (mean and 95% CI or fixed value): Varies by coordination, see Table **??**.

**Notes**

Data came from the records in 2023. The majority of facilities are only capable of doing screening with rapid diagnostic tests (RDTs) but a select few are capable of confirming cases and treating them too. These data are available in 2023, shown in Table **??**.

###### G3.1.3 AS: coverage

↩ Return to the Summary of Health Outcome Parameters.

- Name in the code: as_redux, as_mean, as_max
- Source: HAT Atlas data
- Country of estimate: Uganda
- Statistical distribution and parameters: Fixed value
- Summary statistics (mean and 95% CI or fixed value): Varies by health zone

**Notes**

The percent of the population that is screened by mobile teams in their villages each year. These were determined by the average percent of the population in each health zone that was screened over the years 2018-2022 (for *Mean AS*) and the maximum that was screening over the years in 2000-2022 (for *Int. AS*). We also simulated *Redux AS*, which was half the coverage of *Mean AS*.

See Table **??** for the number of AS people screened per district.

###### G3.1.4 RDT algorithm: diagnostic sensitivity

↩ Return to the Summary of Health Outcome Parameters.

- Name in the code: dx_sens_rdt
- Source: [G8]
- Country of estimate: Guinea and Côte d'Ivoire
- Statistical distribution and parameters: Beta(230, 1)
- Summary statistics (mean and 95% CI or fixed value): 0.996 (0.984, 1.00)

**Notes**

Based on a study in Guinea and Côte d'Ivoire [G8], there was 1 sample from a gHAT patient that tested negative out of 231. Therefore, the parameter distribution for specificity is Beta(230, 1).

###### G3.1.5 CATT algorithm: wastage during AS

↩ Return to the Summary of Health Outcome Parameters.

- Name in the code: dx_wastage_catt_as
- Source: [G9]
- Country of estimate: DRC
- Statistical distribution and parameters: Beta(8, 92)
- Summary statistics (mean and 95% CI or fixed value): 0.080 (0.036, 0.140)

**Notes**

CATT tests come in packs of 50, and the list cost is assumed to consider that a pack is used on 50 patients. Once a pack is opened, one test is used as a positive control and one test is used as a negative control, so wastage is at least 4 percent. The shelf life of the test is one week in refrigeration and wastage in active screening activities is relatively low; generally, wastage of CATT tests in the context of active screening occurs at the end of the day when there are tests remaining in an open pack. To be conservative, we doubled the 4-percent lower bound for wastage and assigned the parameter a distribution of Beta(8, 92).

##### G3.1.6 RDT algorithm: wastage during PS

↩ Return to the Summary of Health Outcome Parameters.

- Name in the code: `dx_wastage_rdt_ps`
- Source: [G10]
- Country of estimate: DRC
- Statistical distribution and parameters: Beta(1, 99)
- Summary statistics (mean and 95% CI or fixed value): 0.010 (<0.001, 0.036)

###### Notes

We followed the same assumption as Snijders and colleagues that less than 1 percent of RDT tests would not be used [G10]. Because there was no sense of uncertainty in this parameter, we assumed a Beta(1, 99) distribution.

#### G3.2 Treatment parameters

##### G3.2.1 Prop. of cases age<6

↩ Return to the Summary of Health Outcome Parameters.

- Name in the code: `treat_prob_under6yo`
- Source: [G11, G12]
- Country of estimate: South Sudan
- Statistical distribution and parameters: Beta(153, 2428)
- Summary statistics (mean and 95% CI or fixed value): 0.059 (0.051, 0.069)

###### Notes

There were only two studies where the number of children under 5 or 6 years of age was stated explicitly [G12, G11] (Figure G1).

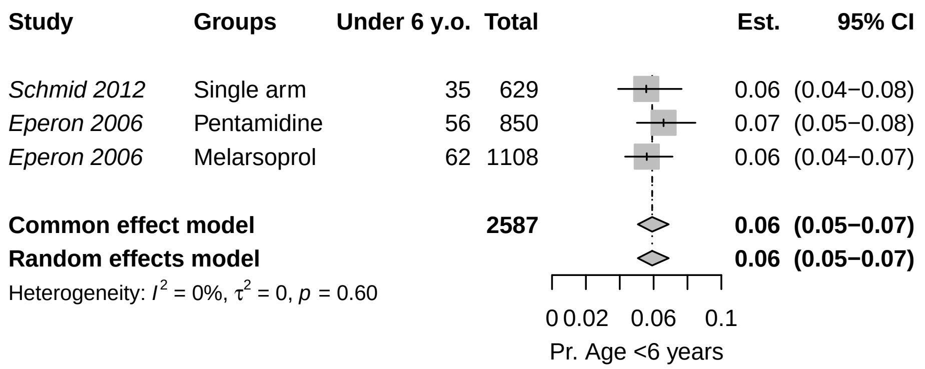

Figure G1: Meta-analysis of two studies with children, and the proportion that were under 6 years old. Reproduced from Antillon *et al* [G1] with permission under a CC-BY licence.

Because the data showed non-significant heterogeneity according to the tau-squared test for heterogeneity, we have chosen to use the fixed-effects combined estimate: 0.059 (0.051, 0.069). The beta parameters of the random-effects estimate Beta(153, 2428) for the probability of that a patient is under 6 years old.

##### G3.2.2 Prop. of cases weight<35 kg among age>6

↩ Return to the Summary of Health Outcome Parameters.

- Name in the code: `treat_prob_under35kg`
- Source: [G12, G13, G14, G15, G16, G17, G18, G19, G20, G21]
- Country of estimate: Various
- Statistical distribution and parameters: Beta(8.30, 360)

- Summary statistics (mean and 95% CI or fixed value): 0.023 (0.010, 0.040)

**Notes**

To determine whether a patient is eligible for fexinidazole treatment, we could not find any studies that would tell us the number of HAT patients who weighed less than 35 kg, but we have estimated the number of people who might weigh less than 35 kg by examining the distribution of weight among patients in the trials in the literature. Furthermore, we have examined how this variable is related to potential selection by age of the study population. We are interested in the proportion of older children and adults that might weigh less than 35 kg, as age under 6 is a contraindication for fexinidazole.

We fit a gamma distribution by the method of moments to the reported mean and standard deviations of each of the studies. For Priotto 2012, no SD was reported, but an interquartile range was reported, so we fit a gamma distribution by the method of Cook [G6].

We then took the expected number of people under 35 kg, and then performed a single-proportion meta-analysis (Fig G2) with the expected number of people in each study under and over the 35 kg threshold.

| Citation | Group | Age Group | Mean weight | Measure of spread | No. of observations | Gamma distr. alpha par. | Gamma distr. beta par. | Prop. <35kg | Simulated No. <35kg |
|---|---|---|---|---|---|---|---|---|---|
| Priotto 2006 | Melarsoprol and nifurtimox | All ages | 49.20 | SD = 14.4 | 18 | 11.67 | 4.21 | 0.16 | 3 |
| Priotto 2006 | Melarsoprol and eflornithine | All ages | 50.00 | SD = 10.3 | 19 | 23.56 | 2.12 | 0.06 | 1 |
| Priotto 2006 | NECT | All ages | 51.40 | SD = 8.4 | 17 | 37.44 | 1.37 | 0.02 | 0 |
| Priotto 2007 | NECT | Over 15 years old | 51.70 | SD = 7.4 | 52 | 48.81 | 1.06 | 0.01 | 0 |
| Priotto 2007 | Eflornithine | Over 15 years old | 53.10 | SD = 7.2 | 51 | 54.39 | 0.98 | 0.00 | 0 |
| Checchi 2007 | NECT | All ages | 44.80 | SD = 15.1 | 31 | 8.80 | 5.09 | 0.28 | 9 |
| Priotto 2009 | NECT | Over 15 years old | 53.00 | SD = 8.7 | 143 | 37.11 | 1.43 | 0.01 | 2 |
| Priotto 2009 | Eflornithine | Over 15 years old | 53.90 | SD = 8.3 | 143 | 42.17 | 1.28 | 0.01 | 1 |
| Ngoyi 2010 | Pentamidine and melarsoprol | Over 12 years old | 56.00 | SD = 10.0 | 360 | 31.36 | 1.79 | 0.01 | 3 |
| Priotto 2012 | Single arm | All ages | 49.00 | IQR: 40-56 | 2190 | 16.37 | 2.96 | 0.12 | 265 |
| Schmid 2012 | Single arm | All ages | 45.00 | SD = 16.0 | 629 | 7.91 | 5.69 | 0.29 | 182 |
| Burri 2016 | Pentamidine | Over 15 years old and ¿35 kg | 48.50 | SD = 7.6 | 40 | 40.83 | 1.19 | 0.03 | 1 |
| Pohlig 2016 | Pentamidine | Over 12 years old and ¿30 kg | 45.70 | SD = 7.8 | 137 | 34.15 | 1.34 | 0.08 | 10 |
| Pohlig 2016 | Pafuramidine | Over 12 years old and ¿30 kg | 44.70 | SD = 7.9 | 136 | 32.02 | 1.40 | 0.10 | 14 |
| Kansiime 2018 | All | Over 15 years old | 51.69 | SD = 9.7 | 109 | 28.22 | 1.83 | 0.03 | 3 |
| Mesu 2018 | NECT | Over 15 years old | 50.70 | SD = 9.6 | 130 | 27.89 | 1.82 | 0.04 | 5 |
| Mesu 2018 | Fexinidazole | Over 15 years old | 50.50 | SD = 8.2 | 264 | 37.93 | 1.33 | 0.02 | 5 |

Table G4: Summary: weight of patients and the resulting proportion who would weigh <35kg. Reproduced from Antillon *et al* [G1] with permission under a CC-BY license.

Because the data showed significant heterogeneity according to the tau-squared test for heterogeneity, we have chosen to use the random-effects estimate: 0.02 (0.01-0.04), represented by probability distribution: Beta(8.30, 360).

| Study | Arm | Events | Total | Est. | 95% CI |
|---|---|---|---|---|---|
| **all_ages_lbl = Adolescents and adults** | | | | | |
| *Priotto 2007* | NECT | 0 | 52 | 0.00 | (0.00−0.07) |
| *Priotto 2007* | Eflornithine | 0 | 51 | 0.00 | (0.00−0.07) |
| *Priotto 2009* | NECT | 2 | 143 | 0.01 | (0.00−0.05) |
| *Priotto 2009* | Eflornithine | 1 | 143 | 0.01 | (0.00−0.04) |
| *Ngoyi 2010* | Pentamidine and Melarsoprol | 3 | 360 | 0.01 | (0.00−0.02) |
| *Burri 2016* | Pentamidine | 1 | 40 | 0.03 | (0.00−0.13) |
| *Pohlig 2016* | Pentamidine | 10 | 137 | 0.07 | (0.04−0.13) |
| *Pohlig 2016* | Pafuramidine | 14 | 136 | 0.10 | (0.06−0.17) |
| *Kansiime 2018* | All | 3 | 109 | 0.03 | (0.01−0.08) |
| *Mesu 2018* | NECT | 5 | 130 | 0.04 | (0.01−0.09) |
| *Mesu 2018* | Fexinidazole | 5 | 264 | 0.02 | (0.01−0.04) |
| **Common effect model** | | | **1565** | **0.03** | **(0.02−0.04)** |
| **Random effects model** | | | | **0.02** | **(0.01−0.04)** |
| Heterogeneity: $I^2 = 70\%$, $\tau^2 = 0.8$, $p < 0.01$ | | | | | |
| **all_ages_lbl = All ages** | | | | | |
| *Priotto 2006* | Melarsoprol and Nifurtimox | 3 | 18 | 0.17 | (0.04−0.41) |
| *Priotto 2006* | Melarsoprol and Eflornithine | 1 | 19 | 0.05 | (0.00−0.26) |
| *Priotto 2006* | NECT | 0 | 17 | 0.00 | (0.00−0.20) |
| *Checchi 2007* | NECT | 9 | 31 | 0.29 | (0.14−0.48) |
| *Priotto 2012* | Single arm | 265 | 2190 | 0.12 | (0.11−0.14) |
| *Schmid 2012* | Single arm | 182 | 629 | 0.29 | (0.25−0.33) |
| **Common effect model** | | | **2904** | **0.16** | **(0.15−0.17)** |
| **Random effects model** | | | | **0.15** | **(0.08−0.27)** |
| Heterogeneity: $I^2 = 95\%$, $\tau^2 = 0.4$, $p < 0.01$ | | | | | |

Test for subgroup differences (fixed effect): $\chi^2_1 = 135.06$, df = 1 ($p < 0.01$)

Test for subgroup differences (random effects): $\chi^2_1 = 17.61$, df = 1 ($p < 0.01$)

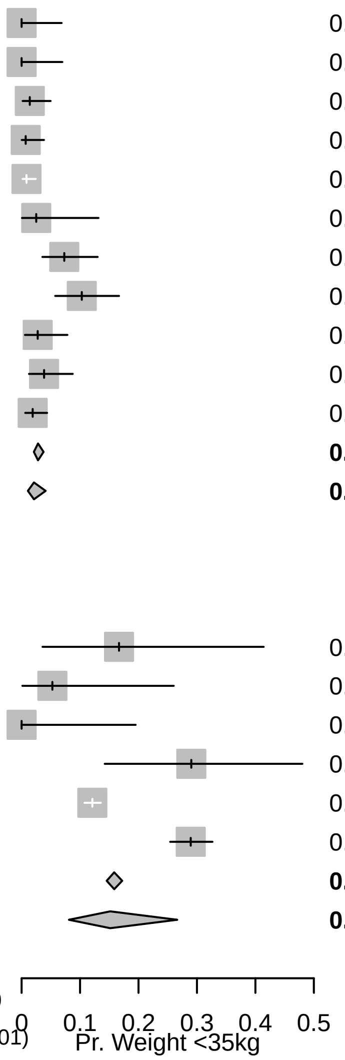

Figure G2: Meta-analysis: simulated number of patients <35kg. Reproduced from Antillon *et al* [G1] with permission under a CC-BY license.

##### G3.2.3 Prop. of S2 cases that are severe

↩ Return to the Summary of Health Outcome Parameters.

- Name in the code: `prob_late_stage2`
- Source: [G11, G12, G13, G14, G15, G16, G17, G22]
- Country of estimate: Various
- Statistical distribution and parameters: Beta(76.9, 44.9)
- Summary statistics (mean and 95% CI or fixed value): 0.634 (0.546, 0.713)

###### Notes

The definition of severe stage 2 gHAT disease by the WHO is when there are more than 100 white blood cells (WBC, leukocytes) per micro-litre in the cerebrospinal fluid. We have searched the clinical trials for the proportion of stage 2 patients that have high concentrations of leukocytes upon admission to treatment.

| Study | Arm | Events | Total | Est. | 95% CI |
|---|---|---|---|---|---|
| *Priotto 2006* | Melarsoprol and Nifurtimox | 9 | 18 | 0.50 | (0.26–0.74) |
| *Priotto 2006* | Melarsoprol and Eflornithine | 12 | 19 | 0.63 | (0.38–0.84) |
| *Priotto 2006* | NECT | 6 | 17 | 0.35 | (0.14–0.62) |
| *Eperon 2006* | Melarsoprol | 407 | 1108 | 0.37 | (0.34–0.40) |
| *Priotto 2007* | NECT | 31 | 52 | 0.60 | (0.45–0.73) |
| *Priotto 2007* | Eflornithine | 39 | 51 | 0.76 | (0.63–0.87) |
| *Checchi 2007* | NECT | 18 | 31 | 0.58 | (0.39–0.75) |
| *Priotto 2009* | NECT | 107 | 143 | 0.75 | (0.67–0.82) |
| *Priotto 2009* | Eflornithine | 115 | 143 | 0.80 | (0.73–0.87) |
| *Ngoyi 2010* | Pentamidine and Melarsoprol | 209 | 272 | 0.77 | (0.71–0.82) |
| *Priotto 2012* | Single arm | 1373 | 2190 | 0.63 | (0.61–0.65) |
| *Schmid 2012* | Single arm | 397 | 629 | 0.63 | (0.59–0.67) |
| **Common effect model** | | | **4673** | **0.58** | **(0.57–0.60)** |
| **Random effects model** | | | | **0.63** | **(0.55–0.71)** |

Heterogeneity: $I^2 = 96\%$, $\tau^2 = 0.3$, $p < 0.01$

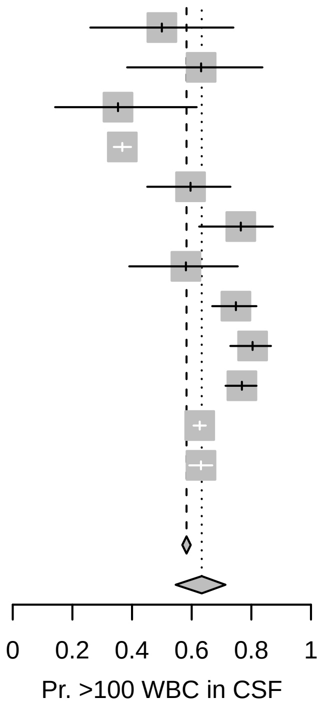

Figure G3: Meta-analysis: proportion of S2 cases that are severe. Reproduced from Antillon *et al* [G1] with permission under a CC-BY license.

Because the data showed significant heterogeneity according to the tau-squared test for heterogeneity, we have chosen to use the random-effects combined estimate: 0.634 (0.546, 0.713), represented by the probability distribution Beta(76.9, 44.9).

##### G3.2.4 Duration, treatment: pentamidine (days)

↩ Return to the Summary of Health Outcome Parameters.

- Name in the code: `treat_duration_penta`
- Source: [G22]
- Country of estimate: Global recommendations
- Statistical distribution and parameters: fixed value
- Summary statistics (mean and 95% CI or fixed value): 7

###### Notes

For NECT treatment, patients must stay in inpatient care for a minimum of 7 days for the eflornithine infusions, and whether they stay for a total of 10 days for nifurtimox administration is unclear. For the most recent clinical trial [G21], NECT patients were released on days 13-18 after admission, but we have assumed that for the most recent trial, the average patient can be released from care after 10 days in the hospital.

##### G3.2.5 Duration, treatment: NECT (days)

↩ Return to the Summary of Health Outcome Parameters.

- Name in the code: `treat_duration_nect`
- Source: [G22, G21]
- Country of estimate: Global recommendations
- Statistical distribution and parameters: fixed value
- Summary statistics (mean and 95% CI or fixed value): 10

**Notes**

For NECT treatment, patients must stay in inpatient care for a minimum of 7 days for the eflornithine infusions, and whether they stay for a total of 10 days for nifurtimox administration is unclear. For the most recent clinical trial [G21], NECT patients were released on days 13-18 after admission, but we have assumed that for the most recent trial, the average patient can be released from care after 10 days in the hospital.

##### G3.2.6 Duration, treatment: fexinidazole (days)

↩ Return to the Summary of Health Outcome Parameters.

- Name in the code: `treat_duration_fexi`
- Source: [G22, G21]
- Country of estimate: global recommendations
- Statistical distribution and parameters: fixed value
- Summary statistics (mean and 95% CI or fixed value): 10

**Notes**

For the only trial that is published [G21], patients were released on days 13-18 after the initiation of treatment, although the treatment only took 10 days, so we have assumed that in routine care the average patient will be in inpatient care for 10 days.

##### G3.2.7 Pr. of relapse (treatment failure): pentamidine

↩ Return to the Summary of Health Outcome Parameters.

- Name in the code: `treat_prob_failure_pent_s1`
- Source: [G11, G17, G18, G19, G23, G24]
- Country of estimate: Various
- Statistical distribution and parameters: Beta(50.3, 665)
- Summary statistics (mean and 95% CI or fixed value): 0.070 (0.053, 0.090)

**Notes**

The WHO guidelines for the treatment of HAT in 2019 [G22] presented existing data on treatment failure of pentamidine treatment. To produce one comprehensive estimate of treatment failure, we performed a meta-analysis on proportions within a single group (Figure G4).

Because the data showed significant heterogeneity according to the tau-squared test for heterogeneity, we have chosen to use the random-effects estimate, to which we assigned a distribution of Beta(50.3, 665).

##### G3.2.8 Pr. of relapse (treatment failure): NECT

↩ Return to the Summary of Health Outcome Parameters.

- Name in the code: `treat_prob_failure_nect_s2`
- Source: [G13, G14, G15, G16, G20, G21]
- Country of estimate: Various
- Statistical distribution and parameters: Beta(15.9, 379)
- Summary statistics (mean and 95% CI or fixed value): 0.040 (0.023, 0.062)

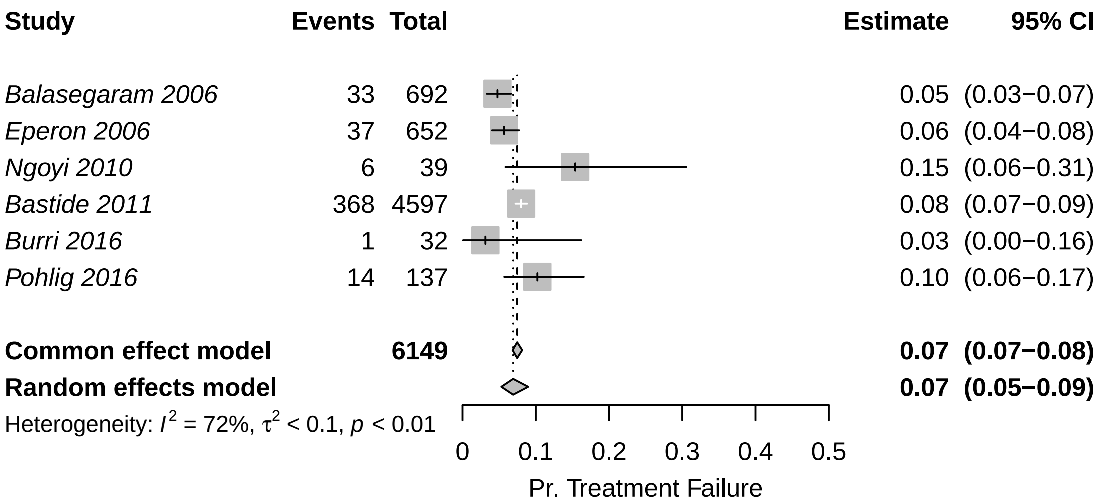

Figure G4: Meta-analysis: probability of treatment failure with pentamidine. Reproduced from Antillon *et al* [G1] with permission under a CC-BY license.

###### Notes

The WHO guidelines for the treatment of HAT in 2019 [G22] presented existing data on treatment failure of NECT. Kansiime and colleagues [G20] also performed a systematic review of studies estimating the outcomes of NECT treatment. To produce one comprehensive estimate of treatment failure, we performed a meta-analysis on proportions within single groups (Figure G5).

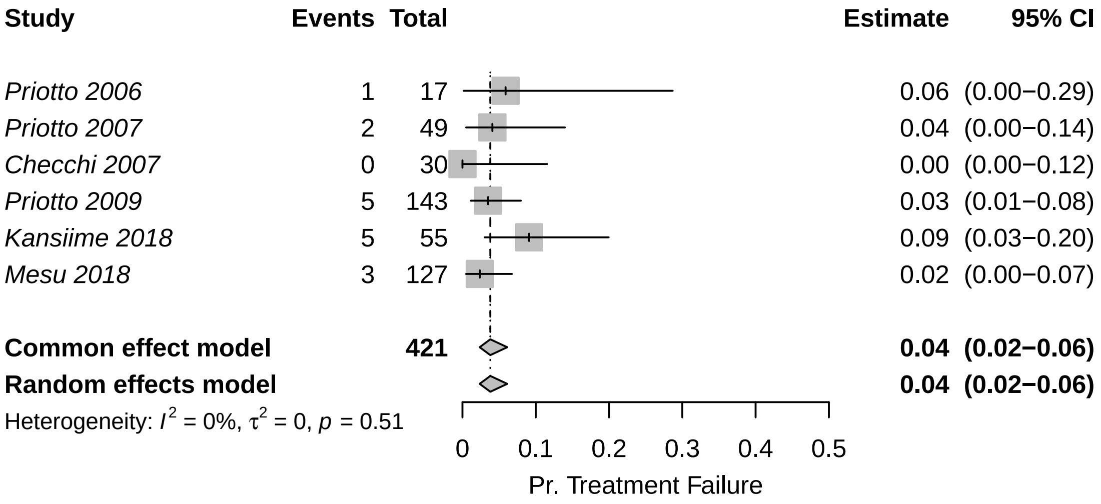

Figure G5: Meta-analysis: probability of treatment failure due to NECT. Reproduced from Antillon *et al* [G1] with permission under a CC-BY license.

Because the data showed non-significant heterogeneity according to the tau-squared test for heterogeneity, we have chosen to use the fixed-effects estimate, to which we assigned a distribution of Beta(15.9, 379).

##### G3.2.9 Pr. of relapse: fexinidazole

↩ Return to the Summary of Health Outcome Parameters.

- Name in the code: `treat_prob_failure_fexi`
- Source: [G22]
- Country of estimate: DRC
- Statistical distribution and parameters: Beta(9.49, 497)
- Summary statistics (mean and 95% CI or fixed value): 0.018 (0.009, 0.032)

###### Notes

Mesu and colleagues [G21] have published the only study on fexinidazole treatment effectiveness in late-stage 2 cases. Moreover, the accompanying meta-analysis for the WHO treatment guidelines released in 2019 shows the outcomes of an additional extension study on stage 1, both early and late-stage 2 disease as well for the data from Mesu et al stratified by the concentration of WBC in the CSF [G22]. We produced a meta-analysis with the same studies (Figure G6).

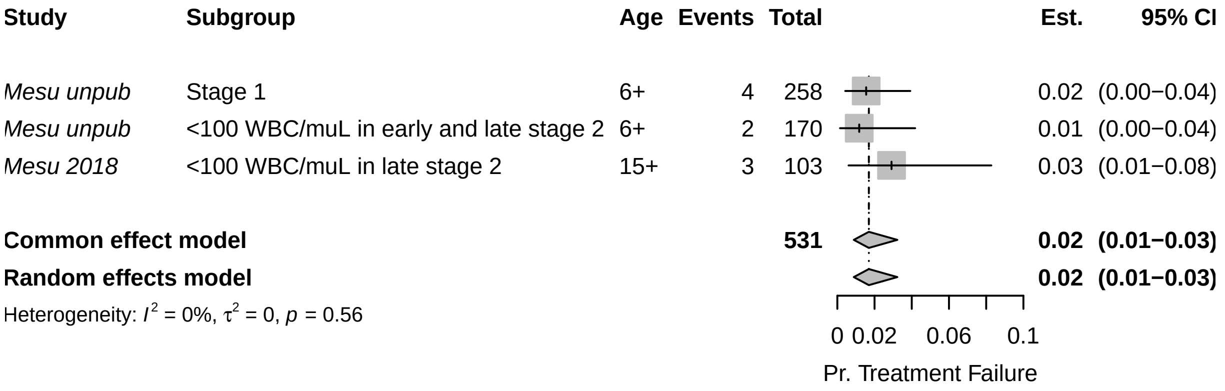

Figure G6: Meta-analysis: probability of treatment failure due to fexinidazole. Reproduced from Antillon *et al* [G1] with permission under a CC-BY license.

Because the data showed non-significant heterogeneity according to the tau-squared test for heterogeneity, we have chosen to use the fixed-effects combined estimate: 0.018 (0.009, 0.032), for which we assigned a distribution of Beta(9.49, 497).

##### G3.2.10 SAE: pentamidine

↩ Return to the Summary of Health Outcome Parameters.

- Name in the code: `treat_prob_sae_pent_s1`
- Source: [G11, G18, G19]
- Country of estimate: DRC and South Sudan
- Statistical distribution and parameters: Beta(1.43, 551)
- Summary statistics (mean and 95% CI or fixed value): 0.003 (<0.001, 0.008)

###### Notes

As part of the WHO guidelines for the treatment of HAT in 2019 [G22], Cochrane performed a systematic review of studies that evaluated the efficacy of NECT compared to fexinidazole studies and presented the probability of serious adverse events. We provide a meta-analysis with the same studies here (Figure G7). Severe or serious adverse events in studies for S1 treatment were defined as "significant hazard, contra-indication, side effect, or precaution" [G11, G18, G19].

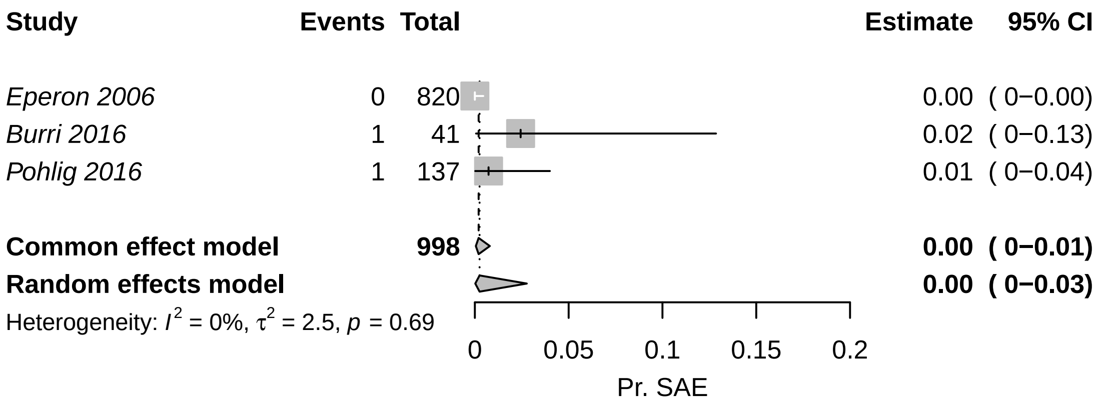

Figure G7: Meta-analysis: probability of severe adverse events (SAE) due to pentamidine treatment. Reproduced from Antillon *et al* [G1] with permission under a CC-BY license.

Because the results do not contain evidence of significant heterogeneity, we have chosen to use the fixed (pooled) estimate of 0 (0-0.01), which would result from a beta distribution of Beta(1.43, 551).

##### G3.2.11 SAE: NECT

↩ Return to the Summary of Health Outcome Parameters.

- Name in the code: `treat_prob_sae_nect_s2`
- Source: [G13, G14, G15, G16, G20, G21]
- Country of estimate: DRC
- Statistical distribution and parameters: Beta(40.9, 368)
- Summary statistics (mean and 95% CI or fixed value): 0.100 (0.073, 0.131)

###### Notes

The WHO guidelines for treatment of gHAT in 2019 [G22], presented all NECT studies to date, as did Kansiime and colleagues [G20]. We searched through these studies for evidence of the probability of severe adverse vents (SAEs), described as events of Grade 3 or higher according to the National Cancer Institute Common Toxicity Criteria for Adverse Events (CTCAE).

To produce one comprehensive estimate of the probability of SAE, we performed a meta-analysis on proportions within single groups (Figure G8).

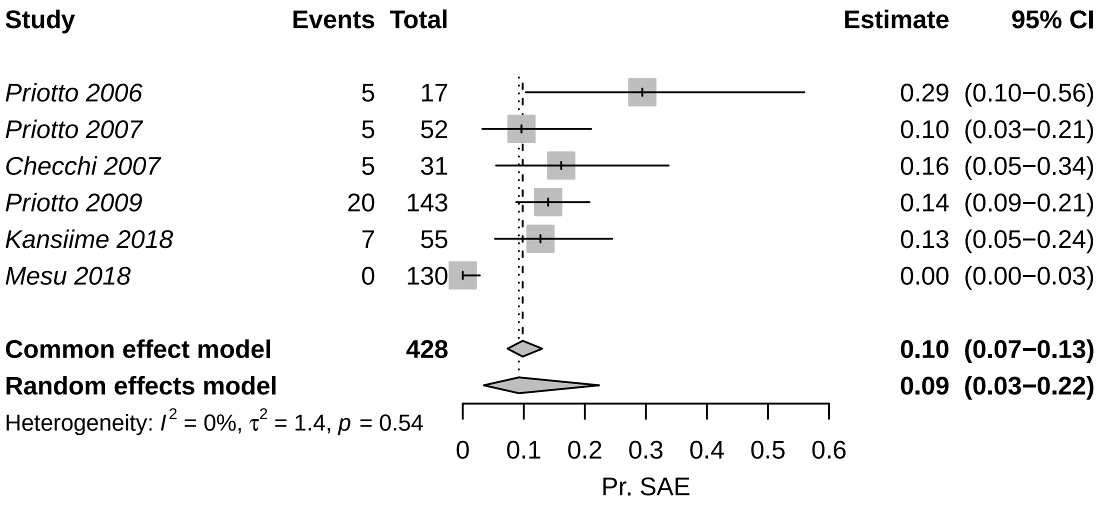

Figure G8: Meta-analysis: probability of severe adverse events (SAE) due to NECT treatment. Reproduced from Antillon *et al* [G1] with permission under a CC-BY license.

Because the data showed non-significant heterogeneity according to the tau-squared test for heterogeneity, we have chosen to use the fixed-effects estimate: 0.098 (0.073, 0.130), which would result from a beta distribution of Beta(40.9, 368).

##### G3.2.12 SAE: fexinidazole

↩ Return to the Summary of Health Outcome Parameters.

- Name in the code: `treat_prob_sae_fexi`
- Source: [G21]
- Country of estimate: DRC
- Statistical distribution and parameters: Beta(3, 261)
- Summary statistics (mean and 95% CI or fixed value): 0.011 (0.002, 0.027)

###### Notes

There is only one published study on fexinidazole, so the probability of serious adverse events will be parameterized with the observations from that study: 4 adverse events attributable to fexinidazole in 3 people among 264 people, so we have assigned a distribution of Beta(3, 261).

There were additional data reported in the appendix to the WHO's interim guidelines ([G22]) related to studies that are ongoing. However, since we do not know details about whether those SAEs were attributable to treatment, we have chosen to omit those data.

###### G3.2.13 Duration, SAE (days)

↩ Return to the Summary of Health Outcome Parameters.

- Name in the code: `treat_duration_sae`
- Source: [G25]
- Country of estimate: DRC
- Statistical distribution and parameters: Gamma(1.22, 2.38)
- Summary statistics (mean and 95% CI or fixed value): 2.90 (0.130, 9.85)

**Notes**

Our only source of information for the duration of severe adverse events (SAEs) is Alirol 2013 [G25], which lists the most common adverse events and the median duration of these events. Most events last a median of 1-2 days (with interquartile ranges reaching up to 4 days).

For simplicity, we have fit a gamma distribution with interquartile range of 1-4 days. Our distribution is therefore Gamma(1.22, 2.38) with a mean and 95% confidence interval of 2.90 (0.130, 9.85), which provides a sufficiently large range of values in light of the scarce information we have.

#### G3.3 Life-years lost (DALY) parameters

↩ Return to the Summary of Health Outcome Parameters.

##### G3.3.1 Age of death from infection

↩ Return to the Summary of Health Outcome Parameters.

- Name in the code: `age_of_death`
- Source: [G11, G12, G13, G14, G15, G16, G17, G18, G19, G20, G21, G23, G26, G25]
- Country of estimate: DRC and South Sudan
- Statistical distribution and parameters: Gamma(148, 0.18)
- Summary statistics (mean and 95% CI or fixed value): 26.6 (22.4, 31.1)

**Notes**

No good registry of the age of infection exists, so we have searched through the literature that we have used to parameterize the model for the average age of HAT patients. Among the studies that we have used to inform other parameters, eight studies reported age information in a sample of patients of all ages, and nine studies reported the age information in a sample of older children or adults (12-15 years and older).

However, the data exists in a state that is difficult to synthesize, as shown in Table G5.

Therefore, we have fit a gamma distribution to the means and medians of the studies that included patients of all ages. We have omitted the median from Alirol et al. 2013 [G25] as this median seems unusually high – even higher than the mean age of patients in studies where only adults (over the age of 15) were recruited. Our distribution is therefore Gamma(148, 0.182) with a mean and 95% confidence interval of 26.6 (22.4, 31.1), which provides a sufficiently large bound of uncertainty in lieu of the information we have.

| Citation | Group | Age Group | Summary |
|---|---|---|---|
| Priotto 2006 | Melarsoprol and Nifurtimox | All ages | Mean: 29.1 range: 5-56 |
| Priotto 2006 | Melarsoprol and Eflornithine | All ages | Mean: 28.1 range: 11-61 |
| Priotto 2006 | NECT | All ages | Mean: 29.1 range: 9-62 |
| Balasegaram 2006 | Pentamidine | All ages | 148 under 15 and 504 over 15 |
| Eperon 2006 | Pentamidine | All ages | 56 patients 0-5, 226 patients 6-15, 568 patients 15+ |
| Eperon 2006 | Melarsoprol | All ages | 63 patients 0-5, 249 patients 6-15, 796 patients 15+ |
| Priotto 2007 | NECT | Over 15 years old | Mean: 33.1 range: 15-69 |
| Priotto 2007 | Eflornithine | Over 15 years old | Mean: 36.1 range: 15-70 |
| Checchi 2007 | NECT | All ages | Mean: 23.9 range: 4-45 |
| Priotto 2009 | NECT | Over 15 years old | Mean: 32.8 SD: 12.5 |
| Priotto 2009 | Eflornithine | Over 15 years old | Mean: 34.6 SD: 13.5 |
| Ngoyi 2010 | Pentamidine | Over 12 years old | Mean: 35 SD: 13 |
| Ngoyi 2010 | Pentamidine and Melarsoprol | Over 12 years old | Mean: 34 SD: 12 |
| Priotto 2012 | Single arm | All ages | Median: 24 IQR: 15-35 |
| Schmid 2012 | Single arm | All ages | 35 patients 0-4 yo, 65 patients 5-11 yo, and 529 patients 12 yo or more. |
| Hasker 2012 | All | All ages | Median: 27 IQR: 16-40 |
| Alirol 2013 | Single arm | All ages | Median: 36 IQR: 20-50 |
| Burri 2016 | Pentamidine | Over 15 years old and >35 kg | Median: 31 range: 15-50 |
| Pohlig 2016 | Pentamidine | Over 12 years old and >30 kg | Median: 31 range: 13-75 |
| Pohlig 2016 | Pafuramidine | Over 12 years old and >30 kg | Median: 30 range: 12-64 |
| Kansiime 2018 | NECT | Over 15 years old | Mean: 27.23 SD: 12.07 |
| Kansiime 2018 | Eflornithine | Over 15 years old | Mean: 27.33 SD: 8.59 |
| Mesu 2018 | NECT | Over 15 years old | Mean: 35.2 SD: 13.2 |
| Mesu 2018 | Fexinidazole | Over 15 years old | Mean: 34.5 SD: 12.6 |

Table G5: Summary: age of death from infection. Reproduced from Antillon *et al* [G1] with permission under a CC-BY license.

##### G3.3.2 Life expectancy

↩ Return to the Summary of Health Outcome Parameters.

- Name in the code: `life_expectancy`
- Source: [G27]
- Country of estimate: UGA
- Statistical distribution and parameters: Interpolation
- Summary statistics (mean and 95% CI or fixed value): See below for age. Life expectancy at birth is 68.25 in 2023.

**Notes**

We took age-specific life expectancy at around the time when people die of HAT in Uganda for 2023, the last year for which there are estimates.

We see here that the expected years of life left at each of the ages is shown in Table G6.

| Age | Years left |
|---|---|
| 15-19 | 56.46 |
| 20-24 | 51.79 |
| 25-29 | 47.25 |
| 30-34 | 42.78 |
| 35-39 | 38.39 |
| 40-44 | 34.14 |
| 45-49 | 30.03 |
| 50-54 | 26.12 |

Table G6: Life-years left for each age group. Source: https://population.un.org/wpp/downloads?folder=Standard%20Projections&group=Mortality. Excel file called "Abridged Life Table - Both Sexes".

Using these data, we made an interpolating function in R (function: `approxfun`) that would calculate the life years left for each age of death (see previous parameter, `age_of_death`). The sample of life expectancy at death yields 45.4 [41.4–49.9].

##### G3.3.3 Disability weights: S1 disease

↩ Return to the Summary of Health Outcome Parameters.

- Name in the code: `disability_weighting_s1`
- Source: [G28]
- Country of estimate: GBD
- Statistical distribution and parameters: Beta(23.0, 147)
- Summary statistics (mean and 95% CI or fixed value): 0.135 (0.088, 0.190)

**Notes**

The Global Burden of Disease listed the impact of sleeping sickness as equivalent to the health state labelled "Motor plus cognitive impairments, severe" and their estimate for a disability weight is 0.542 (0.374-0.702), using the 2013 weight values. No distinction was made between stage 1 and 2 of the disease.

While this seems appropriate for the second stage of sleeping sickness, for stage 1 disability we chose to use the disability weights for equivalent to "infectious disease, acute episode, severe", which is described as "has a high fever and pain, and feels very weak, which causes great difficulty with daily activities" and has a much lower disability weight equivalent to 0.133 (0.088-0.190). The distribution for this parameter is therefore Beta(23.0, 147).

It should be noted that other cost-effectiveness analyses have used different values for disability weights [G29, G30, G31]. These values arise from the 1994 Global Burden of Disease Study but we prefer to consider updated values. Since most of the disability is due to deaths rather than illness during life, we do not believe that this difference is cause for concern.

##### G3.3.4 Disability weights: S2 disease

↩ Return to the Summary of Health Outcome Parameters.

- Name in the code: `disability_weighting_s2`
- Source: [G28]
- Country of estimate: GBD
- Statistical distribution and parameters: Beta(18.4, 15.6)
- Summary statistics (mean and 95% CI or fixed value): 0.541 (0.375, 0.703)

**Notes**

The Global Burden of Disease listed the impact of sleeping sickness as equivalent to the health state labeled "Motor plus cognitive impairments, severe" and their estimate for a disability weight is 0.542 (0.374-0.702), using the 2013 weight values. No distinction was made between stage 1 and 2 of the disease. The distribution for the parameter is Beta(18.4, 15.6).

It should be noted that other cost-effectiveness analyses have used different values for disability weights [G29, G30, G31]. These values arise from the 1994 Global Burden of Disease Study but we prefer to consider updated values. Since most of the disability is due to deaths rather than illness during life, we do not believe that this difference is cause for concern.

##### G3.3.5 Disability weights: SAE

↩ Return to the Summary of Health Outcome Parameters.

- Name in the code: `disability_weighting_sae`
- Source: [G28]
- Country of estimate: GBD
- Statistical distribution and parameters: Uniform(0.037, 0.114)
- Summary statistics (mean and 95% CI or fixed value): 0.076 (0.039, 0.112)

**Notes**

As far as we are aware, no one has considered the disability due to severe adverse events attributable to gHAT treatment, but the most common adverse events are gastrointestinal problems and headaches.

We consulted the Global Burden of Disease for disability weights. The health state labeled "symptomatic tension-type headache" was described as "moderate headache that also affects the neck, which causes difficulty in daily activities" and was estimated to have a disability weight equal to 0.037 (0.022–0.057). The health state labeled "moderate symptomatic gastritis and duodenitis without anaemia" was described as "abdominopelvic problem, moderate has pain in the belly and feels nauseous; the person has difficulties with daily activities" and was estimated to have a disability weight equal to 0.114 (0.078–0.159).

Our distribution is therefore Uniform 0.037-0.114. Since most of the disability is due to death rather than illness during life, we do not believe that the uncertainty in this parameter is cause for concern for the purpose of the conclusions of this analysis.

#### G3.4 Vector control parameters

↩ Return to the Summary of Health Outcome Parameters.

| District | Total area, sq-km | Watersheds covered, sq-km | Targets |
|---|---|---|---|
| Adjumani | 3101 | 323 | 1373 |
| Amuru | 3642 | 229 | 1576 |
| Arua | 4385 | 645 | 4872 |
| Koboko | 764 | 236 | 1688 |
| Maracha | 445 | 297 | 3277 |
| Moyo | 1865 | 416 | 3275 |
| Yumbe | 2332 | 837 | 9435 |

Table G7: Vector control areas and targets in 2017-18, when VC was the most ubiquitous. These parameters are also listed in **??**.

##### G3.4.1 Square-kilometres of vector control

↩ Return to the Summary of Health Outcome Parameters.

- Name in the code: `vc_area`
- Source: [G32]
- Country of estimate: Uganda
- Statistical distribution and parameters: Fixed value
- Summary statistics (mean and 95% CI or fixed value): Varies

**Notes**

Defined by the historical maximum for each district, shown by the watersheds covered in Table G7.

For reactive vector control, we will assume 50 $km^2$ and multiply by the units necessary to cover the villages of the cases simulated.

##### G3.4.2 VC target density

↩ Return to the Summary of Health Outcome Parameters.

- Name in the code: `vc_area`
- Source: [G32]
- Country of estimate: Uganda
- Statistical distribution and parameters: Fixed value
- Summary statistics (mean and 95% CI or fixed value): Varies.

**Notes**

Defined by the historical maximum for each district, shown by the Targets column in Table G7.

For reactive vector control, we will assume that 8.5 targets per square kilometre, as this was the average in all districts.

##### G3.4.3 Replacement rate of targets per year

↩ Return to the Summary of Health Outcome Parameters.

- Name in the code: `vc_deployments_yr`
- Source: [G33, G34]
- Country of estimate: Uganda
- Statistical distribution and parameters: Fixed value
- Summary statistics (mean and 95% CI or fixed value): 2

**Notes**

We have set this parameter as a fixed number, as this is the number of times that one must replace a set of targets in order to provide continuous protection throughout the year [G35, G34].

### G4 Cost parameters

#### G4.1 Screening cost parameters

##### G4.1.1 AS: capital & management costs per person screened

↩ Return to the Summary of Cost Parameters.

- Name in the code: `as_cost_team_cap_mgmt`
- Source: ISSEP 2023
- Country of estimate: Uganda
- Distribution and parameters: Gamma(245, 0.004)
- Summary statistics (mean and 95% CI or fixed value): $0.99 (0.88, 1.13)

**Notes**

Capital and management costs are denominated in 2023 US dollars.

To our knowledge, no active surveillance costs have been estimated via a detailed costing study [G36]. In 2023, the program has budgeted 8,000 USD in 2023 for two missions of 4,000 screens each to do active screening among refugees from South Sudan, which comes down to $1 per person. In 2022, the program had budgeted 10,500 USD to screen 12,000 people, which comes down to $0.875. We took the lower price as the lower confidence interval, and the current price as the mean, with $1.125 as the upper confidence interval. Therefore, we chose a Gamma distribution of Gamma(245, 0.004) which yielded a 95% confidence interval of: 0.995 (0.875, 1.125), or $0.99 (0.88, 1.13) in USD.

##### G4.1.2 RS: capital & management costs per person screened

↩ Return to the Summary of Cost Parameters.

- Name in the code: `rs_cost_team_cap_mgmt`
- Source: ISSEP 2023
- Country of estimate: Uganda
- Distribution and parameters: Gamma(186, 0.009)
- Summary statistics (mean and 95% CI or fixed value): $1.74 (1.50, 2.00)

**Notes**

Capital and management costs are denominated in 2023 US dollars.

To our knowledge, no active surveillance costs have been estimated via a detailed costing study [G36]. In 2023, the program has budgeted 7,000 USD in 2023 for two missions to do reactive screening among 4,000 community members where there are case-positives, which comes down to $1.75 per person. In 2022, the program had budgeted 9,000 USD for three missions that would target 10,000 people, coming to $1.50. We took the lower price as the lower confidence interval, and the current price as the mean, with $2.00 as the upper confidence interval. Therefore, we chose a Gamma distribution of Gamma(186, 0.009) which yielded a 95% confidence interval of: $1.74 (1.50, 2.00).

##### G4.1.3 AS: pre-mission communications and awareness

↩ Return to the Summary of Cost Parameters.

- Name in the code: `as_cost_comms`
- Source: ISSEP 2019 budget
- Country of estimate: Uganda
- Distribution and parameters: Gamma(8.48, 0.042)
- Summary statistics (mean and 95% CI or fixed value): $0.36 (0.16, 0.63)

**Notes**

Communication costs are denominated in 2023 US dollars. This parameter refers to the communications activities that are undertaken before active screening missions to refugee camps. Raising awareness constitutes printing and producing brochures and t-shirts, discussions with community mobilization and sensitization before a mission, discussion with health workers, and broadcast of radio content before a mission. We got the estimates of these costs from the 2019 budget for ISSEP had costs for raising awareness.

Altogether, that year it cost about 0.27 USD per person screened to undertake communications activities (equivalent to 0.32 USD in 2023).

We have parameterized a Gamma distribution with 95% confidence intervals that stretch from half and double the value of 0.32: Gamma(8.475, 0.042). This yields a distribution of 0.357 (0.158, 0.633). Although it has a slightly higher mean, it characterizes the uncertainty well.

###### G4.1.4 RS: pre-mission communications and awareness

↩ Return to the Summary of Cost Parameters.

- Name in the code: `rs_cost_comms`
- Source: ISSEP 2019 budget
- Country of estimate: Uganda
- Distribution and parameters: Gamma(8.48, 0.050)
- Summary statistics (mean and 95% CI or fixed value): $0.42 (0.19, 0.75)

**Notes**

Communication costs are denominated in 2023 US dollars. This parameter refers to the communications activities that are undertaken before reactive screening missions to villages that have witnessed a HAT case. Raising awareness constitutes printing and producing brochures and t-shirts, discussions with community mobilization and sensitization before a mission, discussion with health workers, and broadcast of radio content before a mission. We got the estimates of these costs from the 2019 budget for ISSEP had costs for raising awareness.

Altogether, that year it cost about 0.32 USD per person screened to undertake communications activities (equivalent to 0.37 USD in 2023).

We have parameterized a Gamma distribution with 95% confidence intervals that stretch from half and double the value of 0.37: Gamma(8.475, 0.0421). This yields a distribution of 0.422 (0.188, 0.751). Although it has a slightly higher mean, it characterizes the uncertainty well.

###### G4.1.5 CATT algorithm: cost per test used

↩ Return to the Summary of Cost Parameters.

- Name in the code: `dx_cost_catt`
- Source: [G37]
- Country of estimate: international market (Belgium)
- Distribution and parameters: Gamma(1140, 0.0006897)
- Summary statistics (mean and 95% CI or fixed value): $0.79 (0.74, 0.83)

**Notes**

The CATT test is sold in the international market by Institute of Tropical Medicine in Antwerp.

A kit of reagents is 280.18€ for 500 tests and accessories are 35.82€ for 250 tests, which yields a total of 0.70€ per person [G37]. The capital necessary to carry out the test (the rotator field kit) is considered under the parameter for capital costs.

The price of the CATT varies due to uncertainty in the exchange with the USD. Judging by variation in the five years previous to and including 2024 (0.85-0.95), the range of the cost of the test can be between US$0.74-0.83, which is the 95% confidence interval of a distribution given by Gamma(1140, 0.0006897). The final distribution is given by $0.79 (0.74, 0.83) in USD.

Although delivery costs (described in G4.2.6) are considered to be about 45% of the cost, after conversations with ITM colleagues, we decided to assume that the delivery cost for this is equal to a 10% markup per year. The markup is higher than the RDT tests because the CATT test shipping may need a cold-chain, whereas the RDT shipping would not (CATT reagents must be stored in temperatures of 2-8C; see G4.1.6). The 10% estimate was also the assumption taken in a previous costing paper by Snijders and colleagues [**?** ].

###### G4.1.6 RDT: costs per test used

↩ Return to the Summary of Cost Parameters.

- Name in the code: `dx_cost_rdt`
- Source: Co-authors from ITM
- Country of estimate: international market (Belgium)
- Distribution and parameters: Gamma(1140, 0.002205)
- Summary statistics (mean and 95% CI or fixed value): $2.51 (2.37, 2.66)

**Notes**

If one takes the Snijders et al estimate from the micro-costing analysis the cost is between 0.85 and 1.97 in 2018 USD, from Abbott and Coris, respectively. The Abbott RDT is no longer available, and therefore, one can only purchage tests from Coris. Those tests costs would be equivalent to 1.50€. However, as of 2025, Coris Sero-K-Set RDTs 2.25€ per test.

The price of the RDT varies due to uncertainty in the exchange with the USD. Judging by variation in the five years previous to and including 2024 (0.85-0.95), the range of the cost of the test can be between US$1.904-2.128, which is the 95% confidence interval of a distribution given by Gamma(1139.58093, 0.002205). The final distribution is given by $2.51 (2.37, 2.66).

Although delivery costs (described in G4.2.6) are considered to be about 45% of the cost, after conversations with ITM colleagues, we decided to assume that the delivery cost for this is equal to a 5% markup per year. The markup is lower than the CATT tests because the CATT test shipping may need a cold-chain, whereas the RDT shipping would not (CATT reagents must be stored in temperatures of 2-8C; see G4.1.5).

##### G4.1.7 PS: capital costs of a facility (RDT only)

↩ Return to the Summary of Cost Parameters.

- Name in the code: `ps_cost_facility_cap_mgmt_rdt`
- Source: ISSEP 2023 budget
- Country of estimate: Uganda
- Distribution and parameters: Gamma(25.3, 13.5)
- Summary statistics (mean and 95% CI or fixed value): 341 (221, 486)

**Notes**

Capital costs are denominated in 2023 USD and apply to each health centre or hospital that is capable of HAT screening with RDT tests. The components are shown in Table G8.

Supervision and transportation of samples for confirmation cost 136.54 USD per facility per year, but this was an increase in 2023 from 117.30 USD in 2022.

Additionally, it costs 174.36 per year to provide for transport and coordination costs of confirming RDT positive individuals in these clinics at other clinics or through outreach.

| Item | Data | Distribution | Sample summary |
|---|---|---|---|
| Supervision & monitoring | 117.30 (2022) 136.54 (2023) | Gamma(191, 0.710) | 136 (117 156) |
| Transport cost of samples RDT to confirmation clinics | 174.36 | Gamma(8.47, 23.2) | 196 (87.20, 349) |
| Total | | Gamma(25.3, 13.5) | 341 (221, 486) |

Table G8: Items to make up the costs of managing an RDT-only clinic.

To parameterize the model, we assign a distribution with 95% confidence intervals equal to half and double the costs of the transport costs, and where the lower confidence interval and the mean are equal to the costs in 2022 and 2023 of supervision and monitoring. We simulate two vectors with those gamma distributions and fit the resulting vector to a gamma distribution which yields the distribution listed above under "total": a Gamma(25.3, 13.5), which yields a distribution with mean and confidence intervals of 341 (221, 486).

##### G4.1.8 PS: capital costs of a facility (LAMP & confirmation centers)

↩ Return to the Summary of Cost Parameters.

- Name in the code: `ps_cost_facility_cap_mgmt`

- Source: ISSEP 2023 budget
- Country of estimate: Uganda
- Distribution and parameters: Gamma(137, 8.53)
- Summary statistics (mean and 95% CI or fixed value): 1166 (979, 1370)

**Notes**

Capital costs are denominated in 2023 USD and apply to each health centre or hospital that is capable of HAT screening with RDT tests. The components are shown in Table G9.

Supervision and transportation of samples for confirmation cost 136.54 USD per facility per year, but this was an increase in 2023 from 117.30 USD in 2022.

Additionally, it costs 174.36 per year to provide for transport and coordination costs of confirming RDT positive individuals in these clinics at other clinics or through outreach.

| Item | Data | Distribution | Sample summary |
|---|---|---|---|
| Supervision & monitoring | 117.30 (2022) 136.54 (2023) | Gamma(191, 0.710) | 136 (117 156) |
| Lab consumables for facilities: rotator, hemocrit centrifuge | 748.62 | Gamma(1530, 0.488) | 748 (711, 786) |
| Laboratory reporting | 185 | Gamma(8.475 24.6) | 208 (92.5 370) |
| Total | | Gamma(137, 8.53) | 1166 (979, 1370) |

Table G9: Items to make up the costs of managing an clinic that can perform confirmation tests.

To parameterize the model, we assign a distribution with 95% confidence intervals equal to half and double the costs of the laboratory reporting costs, and where the lower confidence interval and the mean are equal to the costs in 2022 and 2023 of supervision and monitoring. The lab consumables only vary in terms of the exchange rate between the Euro and the USD (like CATT tests described in G4.1.5). We simulate three vectors with those gamma distributions and fit the resulting vector to a gamma distribution which yields the distribution listed above under "total": a Gamma(137, 8.53), which yields a distribution with mean and confidence intervals of 1166 (979, 1370).

##### G4.1.9 PS: communications and awareness in clinics

↩ Return to the Summary of Cost Parameters.

- Name in the code: `ps_cost_comms`
- Source: ISSEP 2019 budget
- Country of estimate: Uganda
- Distribution and parameters: Gamma(8.475, 33.3)
- Summary statistics (mean and 95% CI or fixed value): 282 (126, 502)

**Notes**

Communication costs are denominated in 2023 US dollars. This parameter refers to the communications activities that are undertaken in clinics. Raising awareness constitutes discussions with community mobilization and sensitization. We got the estimates of these costs from the 2019 budget for ISSEP had costs for raising awareness.

Altogether, that year it cost about 1500 USD for all districts to undertake communications activities (equivalent to 1759 USD in 2023). Per district, this is a cost of 251 USD.

We have parameterized a Gamma distribution with 95% confidence intervals that stretch from half and double the value of 251: Gamma(8.475, 33.3). This yields a distribution of 282 (126, 502). Although it has a slightly higher mean, it characterizes the uncertainty well.

##### G4.1.10 National markup

↩ Return to the Summary of Cost Parameters.

- Name in the code: `program_markup`
- Source: ISSEP budgets

- Country of estimate: Uganda
- Distribution and parameters: Fixed
- Summary statistics (mean and 95% CI or fixed value): 0.10

**Notes**

ISSEP budgets indicated a 10% markup. We assume the same markup even for the portions of the budget that are probably not paid by ISSEP since these funds come from non-profits which also have indirect costs associated to them.

#### G4.2 Treatment cost parameters

##### G4.2.1 Hospital stay: cost per day

↩ Return to the Summary of Cost Parameters.

- Name in the code: `treat_cost_ip_day`
- Source: [G4, G38, G39]
- Country of estimate: Uganda
- Distribution and parameters: Gamma(5.39, 0.86)
- Summary statistics (mean and 95% CI or fixed value): 4.62 (1.58, 9.25)

**Notes**

We got the estimates of inpatient treatment costs from the 2010 WHO CHOICE cost estimates (most recently updated by [G38] and [G39]). In 2010, a consult at a primary hospital in Uganda would be 10.25 (4.08, 6.86) in international dollars. After converting to 2023 USD, the estimates are 3.98 (1.58, 9.25) per day at a primary hospital.

To parameterize the model, we assign a gamma distribution with 95% confidence intervals equal to those reported by WHO CHOICE [G4]. The distribution is Gamma(5.39, 0.86), which yields a distribution with mean and confidence intervals of 4.62 (1.58, 9.25). Although this yields a higher mean, the uncertainty is adequately characterized.

We checked the literature to assess the adequacy of this estimate, and we could only find at the moment an estimate specifically on a per bed-day basis in one study in a private, non-profit hospital which reported a cost of 15.70 USD (2013) per bed day, which is equivalent to 16.77 in 2023 values [G40]. This is a bit elevated, potentially for being a private hospital, but it indicated that our estimate was not too high.

##### G4.2.2 Outpatient consultation: cost

↩ Return to the Summary of Cost Parameters.

- Name in the code: `treat_cost_op_visit`
- Source: [G4, G38, G39]
- Country of estimate: Uganda
- Distribution and parameters: Gamma(2.48, 0.42)
- Summary statistics (mean and 95% CI or fixed value): 1.03 (0.17, 2.66)

**Notes**

We got WHO-CHOICE estimates, which showed that an outpatient visit cost 2.32 (0.46, 6.86) in 2010 in International dollars, which is equivalent to 0.90 (0.17, 2.66) USD in 2023 values. WHO-CHOICE estimates were updated most recently by [G38] and [G39].

The distribution to represent the random-effects estimate is Gamma(2.48, 0.42), which yields a distribution with mean and confidence intervals of 1.03 (0.17, 2.66). Although this yields a higher mean, the uncertainty is adequately characterized.

To assess the adequacy of these estimates, we checked with a study that collected specific information directly from a mix of private, non-profit and government facilities in rural areas in 2012-14 [G41]. They reported costs for outpatient consultations of pneumonia, diarrhoea, malaria and general fevers at around 0.30-0.60 USD (reported in 2021 values) using an approach that looks at marginal costs. By comparison using a total-costing approach (incl. overheads and indirect costs) the costs were closer to 2.60-12.90 USD (reported in 2021 values). We decided to use the WHO-CHOICE estimates, which sat in between the marginal and full-costing approach.

###### G4.2.3 Course of pentamidine: cost

↩ Return to the Summary of Cost Parameters.

- Name in the code: `rx_cost_pentamidine`
- Source: [G36]
- Country of estimate: WHO
- Distribution and parameters: Fixed value
- Summary statistics (mean and 95% CI or fixed value): 54

**Notes**

The cost of pentamidine, for stage 1 disease. Because it is available on the international market, where is it sold in USD, and not subject to the inflationary pressures of any particular country, we have not inflated the cost or converted them to any other currency.

In the future pentamidine treatment it may be replaced with fexinidozole treatment, which would circumvent the need for a lumbar puncture.

###### G4.2.4 Course of NECT: cost

↩ Return to the Summary of Cost Parameters.

- Name in the code: `rx_cost_nect`
- Source: [G42]
- Country of estimate: WHO
- Distribution and parameters: Fixed value
- Summary statistics (mean and 95% CI or fixed value): 360

**Notes**

This represents the cost of NECT to the capital for stage 2 disease. Simarro and colleagues listed a cost of 1440 USD for the treatment of four patients.

Because it is available on the international market, where is it sold in USD, and not subject to the inflationary pressures of any particular country, we have not inflated the cost or converted them to any other currency.

In the future, it may be replaced with fexinidozole and this would be the drug for treatment failures or very severe patients.

###### G4.2.5 Course of fexinidazole: cost

↩ Return to the Summary of Cost Parameters.

- Name in the code: `rx_cost_fexinidazole`
- Source: [G30]
- Country of estimate: WHO
- Distribution and parameters: Fixed value
- Summary statistics (mean and 95% CI or fixed value): 50

**Notes**

The cost of fexanidozole, for stage 1 and 2 disease. In the near future this will be the drug of choice for first-line treatment for both stages of disease. It may require hospitalization, but eventually, it should be taken on an outpatient basis.

###### G4.2.6 Drug delivery mark-up

↩ Return to the Summary of Cost Parameters.

- Name in the code: `rx_delivery_markup`
- Source: [G38, G39]
- Country of estimate: Uganda

- Distribution and parameters: Beta(45, 55)
- Summary statistics (mean and 95% CI or fixed value): 0.45 (0.35, 0.55)

###### Notes

Because we do not know the delivery price of drugs for each country, we have applied the standard value for the mark-up of traded goods recommended by the WHO CHOICE programme for AFRO E: https://www.who.int/teams/health-financing-and-economics/economic-analysis/costing-and-technical-efficiency/quantities-and-unit-prices-(cost-inputs) [G43].

#### G4.3 Vector control cost parameters

##### G4.3.1 Operational cost per kilometer of riverbank covered

↫ Return to the Summary of Cost Parameters.

- Name in the code: `vc_cost_management`
- Source: [G44]
- Country of estimate: Uganda
- Distribution and parameters: Gamma(8.47, 5.25)
- Summary statistics (mean and 95% CI or fixed value): 44.48 (19.76, 79.05).

###### Notes

Vector control operational costs are denominated in 2023 US dollars on a per-square-kilometre basis.

To our knowledge, only one vector control micro-costing study has been performed in Uganda by Shaw and colleagues [G44]. In that study, centred in Arua, targets were laid out across 250 km$^2$ for a total cost of 21,337 USD or 85.40 USD per km$^2$ in 2014. The equivalent cost is 87.23 per km$^2$ in 2023 USD. However, about 4,290 USD of 17.20 USD per sq-km were used for maintenance activities, so a single deployment costed 68.19 USD. Further, the target deployment itself cost 7370 USD. Therefore, the cost of the operation excluding target deployment was 9,677 USD per year, or \$38.70/km$^2$ in 2014 USD in Uganda.

After adjusting for inflation and transferring to Ugandan values in 2023, the costs would be equivalent to 39.53 USD. This underscores the level of uncertainty present. Because we do not want to bias the costs downward, we will take into account the Ugandan cost only and assigned a distribution with confidence intervals equal to half and double the costs. The distribution is Gamma(8.47, 5.25), which yields a distribution with mean and confidence intervals of 44.48 (19.76, 79.05), which is a bit higher in its mean but adequately characterizes the uncertainty.

There have been other studies in other countries, all summarised in a study by Snijders et al [G45]. There, it says that the cost per square kilometre in Yasa Bonga, DRC and Mandoul, Chad is 62 and 67 USD in 2016 values, compared to 71.63 in Uganda in 2016 values - demonstrating that the values are comparable to other locations in West or Central Africa. Another study, from Cote d'Ivoire, was set aside because the vector control is done differently in a forested area.

##### G4.3.2 Cost per target deployed

↫ Return to the Summary of Cost Parameters.

- Name in the code: `vc_cost_target`
- Source: [G44]
- Country of estimate: Uganda
- Distribution and parameters: Gamma(8.48, 0.644)
- Summary statistics (mean and 95% CI or fixed value): 5.46 (2.43, 9.70).

###### Notes

To our knowledge, only one vector control micro-costing study has been performed in Uganda by Shaw and colleagues [G44]. In that study, centred in Arua, Uganda, 1,551 targets were laid out. The target deployment activities cost 7,370 USD, or per target 4.75 USD (in 2014 values).

After adjusting for inflation and transferring to Ugandan values in 2023, the costs would be equivalent to 4.85 per target. The distribution is Gamma(8.47, 0.64), which yields a distribution with mean and confidence intervals of 5.46 (2.43, 9.70), which is a bit higher in its mean but adequately characterizes the uncertainty.

[G5] Briggs A, Claxton K, Sculpher M. Decision Modelling for Health Economic Evaluation. 1st ed. Oxford, UK: Oxford University Press; 2006.

[G6] Cook JD. Determining distribution parameters from quantiles; 2010. Available from: https://www.johndcook.com/quantiles_parameters.pdf.

[G7] Joseph L, Bélisle P. beta.parms.from.quantiles: [R] Computing Beta distribution parameters; 2017. Available from: http://www.medicine.mcgill.ca/epidemiology/joseph/pbelisle/BetaParmsFromQuantiles.html.

[G8] Jamonneau V, Camara O, Ilboudo H, Peylhard M, Koffi M, Sakande H, et al. Accuracy of Individual Rapid Tests for Serodiagnosis of Gambiense Sleeping Sickness in West Africa. PLoS Neglected Tropical Diseases. 2015;9(2):1–9.

[G9] Snijders R, Fukinsia A, Claeys Y, Mpanya A, Hasker E, Meheus F, et al. Cost of a new method of active screening for human African trypanosomiasis in the Democratic Republic of the Congo. PLoS Neglected Tropical Diseases. 2020;14(12):e0008832.

[G10] Snijders R, Fukinsia A, Claeys Y, Hasker E, Mpanya A, Miaka E, et al. Costs and outcomes of integrated human African trypanosomiasis surveillance system using rapid diagnostic tests, Democratic Republic of the Congo. Emerging Infectious Diseases. 2021;27(8):2144.

[G11] Eperon G, Schmid C, Loutan L, Chappuis F. Clinical presentation and treatment outcome of sleeping sickness in Sudanese pre-school children. Acta Tropica. 2006 1;101(1):31–39. Available from: https://linkinghub.elsevier.com/retrieve/pii/S0001706X06002439.

[G12] Schmid C, Kuemmerle A, Blum J, Ghabri S, Kande V, Mutombo W, et al. In-Hospital Safety in Field Conditions of Nifurtimox Eflornithine Combination Therapy (NECT) for T. b. gambiense Sleeping Sickness. PLoS Neglected Tropical Diseases. 2012;6(11).

[G13] Priotto G, Fogg C, Balasegaram M, Erphas O, Louga A, Checchi F, et al. Three Drug Combinations for Late-Stage Trypanosoma brucei gambiense Sleeping Sickness: A Randomized Clinical Trial in Uganda. PLo S Clinical Trials. 2006;1(8):e39.

[G14] Priotto G, Kasparian S, Ngouama D, Ghorashian S, Arnold U, Ghabri S, et al. Nifurtimox-Eflornithine Combination Therapy for Second-Stage Trypanosoma brucei gambiense Sleeping Sickness: A Randomized Clinical Trial in Congo. Clinical Infectious Diseases. 2007;45(11):1435–1442.

[G15] Checchi F, Piola P, Ayikoru H, Thomas F, Legros D, Priotto G. Nifurtimox plus eflornithine for late-stage sleeping sickness in Uganda: A case series. PLoS Neglected Tropical Diseases. 2007;1(2):1–6.

[G16] Priotto G, Kasparian S, Mutombo W, Ngouama D, Ghorashian S, Arnold U, et al. Nifurtimox-eflornithine combination therapy for second-stage African Trypanosoma brucei gambiense trypanosomiasis: a multicentre, randomised, phase III, non-inferiority trial. The Lancet. 2009;374(9683):56–64. Available from: http://dx.doi.org/10.1016/S0140-6736(09)61117-X.

[G17] Ngoyi DM, Lejon V, Pyana P, Boelaert M, Ilunga M, Menten J, et al. How to Shorten Patient Follow-Up after Treatment for Trypanosoma brucei gambiense Sleeping Sickness. The Journal of Infectious Diseases. 2010;201(3):453–463.

[G18] Burri C, Yeramian PD, Allen JL, Merolle A, Serge KK, Mpanya A, et al. Efficacy, Safety, and Dose of Pafuramidine, a New Oral Drug for Treatment of First Stage Sleeping Sickness, in a Phase 2a Clinical Study and Phase 2b Randomized Clinical Studies. PLoS Neglected Tropical Diseases. 2016;10(2):1–18.

[G19] Pohlig G, Bernhard SC, Blum J, Burri C, Mpanya A, Lubaki JPF, et al. Efficacy and Safety of Pafuramidine versus Pentamidine Maleate for Treatment of First Stage Sleeping Sickness in a Randomized, Comparator-Controlled, International Phase 3 Clinical Trial. PLoS Neglected Tropical Diseases. 2016;10(2):1–17.

[G20] Kansiime F, Adibaku S, Wamboga C, Idi F, Kato CD, Yamuah L, et al. A multicentre, randomised, non-inferiority clinical trial comparing a nifurtimox-eflornithine combination to standard eflornithine monotherapy for late stage Trypanosoma brucei gambiense human African trypanosomiasis in Uganda. Parasites and Vectors. 2018;11(1):1–11.

[G21] Mesu VKBK, Kalonji WM, Bardonneau C, Mordt OV, Blesson S, Simon F, et al. Oral fexinidazole for late-stage African *Trypanosoma brucei gambiense* trypanosomiasis: a pivotal multicentre, randomised, non-inferiority trial. The Lancet. 2018;391(10116):144–154.

[G22] World Health Organization. WHO interim guidelines for the treatment of *gambiense* human African trypanosomiasis. World Health Organization; 2019.

[G23] Balasegaram M, Harris S, Checchi F, Hamel C, Karunakara U. Treatment outcomes and risk factors for relapse in patients with early-stage human African trypanosomiasis (HAT) in the Republic of the Congo. Bulletin of the World Health Organization. 2006;84(10):777–782.

[G24] Bastide S, Priotto G, Ecochard R, Etard JF. Effectiveness of short vs. long treatment schedules with pentamidine in first-stage HAT: a large field cohort study. Tropical Medicine & International Health. 2011;16(S1):68–69. Available from: https://onlinelibrary.wiley.com/toc/13653156/2011/16/s1.

[G25] Alirol E, Schrumpf D, Amici Heradi J, Riedel A, De Patoul C, Quere M, et al. Nifurtimox-eflornithine combination therapy for second-stage gambiense human African trypanosomiasis: Médecins Sans Frontières experience in the Democratic Republic of the Congo. Clinical Infectious Diseases. 2013;56(2):195–203.

[G26] Hasker E, Lutumba P, Chappuis F, Kande V, Potet J, De Weggheleire A, et al. Human African Trypanosomiasis in the Democratic Republic of the Congo: A Looming Emergency? PLoS Neglected Tropical Diseases. 2012;6(12):e1950.

[G27] World Bank. Life expectancy at birth, total (years). Washington, DC.: World Bank; 2020. Available from: https://data.worldbank.org/indicator/SP.DYN.LE00.IN?locations=CD.

[G28] GBD 2017 DALYs and HALE Collaborators. Global, regional, and national disability-adjusted life-years (DALYs) for 359 diseases and injuries and healthy life expectancy (HALE) for 195 countries and territories, 1990-2017: A systematic analysis for the Global Burden of Disease Study 2017. The Lancet. 2018;392(10159):1859–1922.

[G29] Hackett F, Berrang Ford L, Fèvre E, Simarro P. Incorporating Scale Dependence in Disease Burden Estimates: The Case of Human African Trypanosomiasis in Uganda. PLoS Neglected Tropical Diseases. 2014;8(2).

[G30] Sutherland CS, Stone CM, Steinmann P, Tanner M, Tediosi F. Seeing beyond 2020: an economic evaluation of contemporary and emerging strategies for elimination of *Trypanosoma brucei gambiense*. The Lancet Global Health. 2017;5(1):e69–e79.

[G31] Bessell PR, Lumbala C, Lutumba P, Baloji S, Biéler S, Ndung'u JM. Cost-effectiveness of using a rapid diagnostic test to screen for human African trypanosomiasis in the Democratic Republic of the Congo. PLoS ONE. 2018 9;13(9):e0204335. Available from: https://dx.plos.org/10.1371/journal.pone.0204335.

[G32] Bessell PR, Esterhuizen J, Lehane MJ, Longbottom J, Mugenyi A, Selby R, et al. Estimating the impact of Tiny Targets in reducing the incidence of Gambian sleeping sickness in the North-west Uganda focus. Parasites & Vectors. 2021;14:1–13.

[G33] Tirados I, Hope A, Selby R, Mpembele F, Miaka EM, Boelaert M, et al. Impact of tiny targets on Glossina fuscipes quanzensis, the primary vector of human African trypanosomiasis in the Democratic

Republic of the Congo. PLOS Neglected Tropical Diseases. 2020;14:e0008270. Available from: https://dx.plos.org/10.1371/journal.pntd.0008270.

[G34] Liverpool School of Tropical Medicine. Use of Tiny Targets to control tsetse flies in Gambian HAT foci: standard operating procedures; 2016. October.

[G35] Tirados I, Esterhuizen J, Kovacic V, Mangwiro TNC, Vale GA, Hastings I, et al. Tsetse Control and Gambian Sleeping Sickness; Implications for Control Strategy. PLoS Neglected Tropical Diseases. 2015;9(8):e0003822.

[G36] Keating J, Yukich JO, Sutherland CS, Woods G, Tediosi F. Human African trypanosomiasis prevention, treatment and control costs: A systematic review. Acta Tropica. 2015;150:4–13.

[G37] CATT/T.b.g. REAGENTS (KIT R500 tests) - ITM Webshop for in vitro diagnostics;. Available from: https://diagnostics.itg.be/product/catt-t-b-g-reagents-kit-r500-tests/.

[G38] Bertram MY, Stenberg K, Brindley C, Li J, Serje J, Watts R, et al. Disease control programme support costs: an update of WHO-CHOICE methodology, price databases and quantity assumptions. Cost Effectiveness and Resource Allocation. 2017 12;15(1):21. Available from: http://resource-allocation.biomedcentral.com/articles/10.1186/s12962-017-0083-6.

[G39] Stenberg K, Lauer JA, Gkountouras G, Fitzpatrick C, Stanciole A. Econometric estimation of WHO-CHOICE country-specific costs for inpatient and outpatient health service delivery. Cost Effectiveness and Resource Allocation. 2018;16(1):1–15. Available from: https://doi.org/10.1186/s12962-018-0095-x.

[G40] Okiria JC, Kamara Ronald. Cost of Providing Health Care Services in the Private Not for Profit Health Sector in Uganda: A Case Study of Nyenga Hospital (FY 2012/2013). International Journal of Science and Research (IJSR). 2016 6;5(6):1333–1338. Available from: https://www.ijsr.net/archive/v5i6/NOV163618.pdf.

[G41] Batura N, Kasteng F, Condoane J, Bagorogosa B, Castel-Branco AC, Kertho E, et al. Costs of treating childhood malaria, diarrhoea and pneumonia in rural Mozambique and Uganda. Malaria Journal. 2022 12;21(1).

[G42] Simarro PP, Diarra A, Ruiz Postigo JA, Franco JR, Jannin JG. The human African trypanosomiasis control and surveillance programme of the World Health Organization 2000-2009: the way forward. PLoS Neglected tropical diseases. 2011 2;5(2):e1007.

[G43] Health Financing and Economics;. Available from: https://www.who.int/teams/health-financing-and-economics/economic-analysis/costing-and-technical-efficiency/quantities-and-unit-prices-(cost-inputs).

[G44] Shaw AP, Tirados I, Mangwiro CT, Esterhuizen J, Lehane MJ, Torr SJ, et al. Costs of using "tiny targets" to control Glossina fuscipes fuscipes, a vector of gambiense sleeping sickness in Arua District of Uganda. PLoS Neglected Tropical Diseases. 2015;9(3):e0003624.

[G45] Snijders R, Shaw APM, Selby R, Tirados I, Bessell PR, Fukinsia A, et al. The cost of sleeping sickness vector control in Yasa Bonga, a health district in the Democratic Republic of the Congo. PLoS Neglected Tropical Diseases. 2024 11;18(11):e0011959. Available from: https://dx.plos.org/10.1371/journal.pntd.0011959.
