## Supplement 1: Supplementary Methods and Supplementary Results for "Transmission and cost-effectiveness modelling to estimate the progress towards elimination and future strategy optimisation for *gambiense* human African trypanosomiasis in Uganda"

### **Supplementary Methods and Supplementary Results**: Transmission modelling to estimate the progress towards elimination and future predictions for *gambiense* human African trypanosomiasis in Uganda

Ching-I Huang[1,2*], Marina Antillon[1,2,3,4], Ronald E Crump[1,2], Samuel A Sutherland[2,4], Paul R Bessell[5,6], Albert Mugenyi[7], Richard Selby[8], Paul E Brown[1,2], Brady Hooley[3], Andrew Hope[8], Sophie Dunkley[8], Rob Sunnucks[1,2], Steve J Torr[8], Fabrizio Tediosi[3], Joseph Ndung'u[6], Emily H Crowley[1,2], Eric Kidega[9], Charles Wamboga[9], and Kat S Rock[1,2]

[1]Zeeman Institute for System Biology and Infectious Disease Epidemiology Research, The University of Warwick, Coventry, U.K.
[2]Mathematics Institute, The University of Warwick, Coventry, U.K.
[3]Warwick Medical School, The University of Warwick, Coventry, U.K.
[4]Swiss Tropical and Public Health Institute, Basel, Switzerland
[5]Epi Interventions Ltd, Edinburgh, U.K.
[6]Foundation for Innovative New Diagnostics, Geneva, Switzerland
[7]Ministry of Agriculture Animal Industry and Fisheries, Kampala, Uganda
[8]Department of Vector Biology, Liverpool School of Tropical Medicine, Liverpool, U.K.
[9]Ministry of Health, Kampala, Uganda

These authors contributed equally to this work.

S21 Results in Adjumani. Summary of effects, costs, elimination of transmission (EoT) by 2030, and cost-effectiveness with and without uncertainty. Means are given along with 95% prediction intervals (PIs). YLL: years of life lost (to fatal disease), YLD: years of life lost to disability, DALYs: disability-adjusted life-years, PS: passive screening, AS: active screening, VC: vector control, ICER: incremental cost-effectiveness ratio, WTP: willingness to pay (USD per DALY averted), EoT: elimination of transmission. . . . 68

S22 Results in Amuru. Summary of effects, costs, elimination of transmission (EoT) by 2030, and cost-effectiveness with and without uncertainty. Means are given along with 95% prediction intervals (PIs). YLL: years of life lost (to fatal disease), YLD: years of life lost to disability, DALYs: disability-adjusted life-years, PS: passive screening, AS: active screening, VC: vector control, ICER: incremental cost-effectiveness ratio, WTP: willingness to pay (USD per DALY averted), EoT: elimination of transmission. . . . . . 69
S23 Results in Arua. Summary of effects, costs, elimination of transmission (EoT) by 2030, and cost-effectiveness with and without uncertainty. Means are given along with 95% prediction intervals (PIs). YLL: years of life lost (to fatal disease), YLD: years of life lost to disability, DALYs: disability-adjusted life-years, PS: passive screening, AS: active screening, VC: vector control, ICER: incremental cost-effectiveness ratio, WTP: willingness to pay (USD per DALY averted), EoT: elimination of transmission. . . . . . 70
S24 Results in Koboko. Summary of effects, costs, elimination of transmission (EoT) by 2030, and cost-effectiveness with and without uncertainty. Means are given along with 95% prediction intervals (PIs). YLL: years of life lost (to fatal disease), YLD: years of life lost to disability, DALYs: disability-adjusted life-years, PS: passive screening, AS: active screening, VC: vector control, ICER: incremental cost-effectiveness ratio, WTP: willingness to pay (USD per DALY averted), EoT: elimination of transmission. . . . . . 71
S25 Results in Maracha. Summary of effects, costs, elimination of transmission (EoT) by 2030, and cost-effectiveness with and without uncertainty. Means are given along with 95% prediction intervals (PIs). YLL: years of life lost (to fatal disease), YLD: years of life lost to disability, DALYs: disability-adjusted life-years, PS: passive screening, AS: active screening, VC: vector control, ICER: incremental cost-effectiveness ratio, WTP: willingness to pay (USD per DALY averted), EoT: elimination of transmission. . . . . . 72
S26 Results in Moyo. Summary of effects, costs, elimination of transmission (EoT) by 2030, and cost-effectiveness with and without uncertainty. Means are given along with 95% prediction intervals (PIs). YLL: years of life lost (to fatal disease), YLD: years of life lost to disability, DALYs: disability-adjusted life-years, PS: passive screening, AS: active screening, VC: vector control, ICER: incremental cost-effectiveness ratio, WTP: willingness to pay (USD per DALY averted), EoT: elimination of transmission. . . . . . 73
S27 Results in Yumbe. Summary of effects, costs, elimination of transmission (EoT) by 2030, and cost-effectiveness with and without uncertainty. Means are given along with 95% prediction intervals (PIs). YLL: years of life lost (to fatal disease), YLD: years of life lost to disability, DALYs: disability-adjusted life-years, PS: passive screening, AS: active screening, VC: vector control, ICER: incremental cost-effectiveness ratio, WTP: willingness to pay (USD per DALY averted), EoT: elimination of transmission. . . . . . 74

### S1 Relationship to Previous Modelling Work

The modelling group that performed this work has published a series of modelling analyses on eliminating the transmission of *gambiense* human African trypanosomiasis (gHAT). The series of studies includes 14 fitting and projections papers [S1, S2, S3, S4, S5, S6, S7, S8, S9, S10, S11, S12, S13, S14] and 4 cost-effectiveness analysis papers [S15, S16, S17, S18]. There are minimal differences in the model structures themselves (see S2). The key difference from previous publications is that projections were generated based on Uganda-specific strategies and costs estimated based on how health facilities and intervention teams operate in Uganda. For transparency, much of the same model information that has been published previously is given here to provide consistent descriptions of our methods and to aid the reader with the present study. Previous studies are all published under a CC-BY license.

Because our methods are similar to other papers some of the supplement text is recycled. The text in the main paper is not recycled, however.

### S2 Summary of updates in models

#### S2.1 Epidemiological model

- The model was fitted to gHAT epidemiological data from districts in Uganda for the first time.
- The algorithm used to infer missing numbers of people screened in active screening has been updated from that reported in Crump *et al* (2021) [S7], see Section S7.2.
- To align our modelling results with the observed data, filtering is applied to the stochastic simulation results for the period with observations, retaining realisations that matched the observed presence/absence of reported cases in 2020–2024, the most recent 5 years of data.
- Where possible, the tsetse model was fitted to the tsetse capture data to estimate the regional tsetse birth rates and the death probabilities in the presence of VC in Uganda (see Section S8.6).
  - The watershed coverage data evaluated via geolocations of target deployments were used to infer the case coverage of VC and estimate the overall effectiveness in each district (see Section S8.4).
  - The information on VC scaled back in all districts was included in the model fitting process (see Table S6).
- A hash-based matching, pseudo-random number generator [S19] is introduced to our stochastic code to perform fair comparison among different counterfactual scenarios and predict impacts of different strategies in the cost-effectiveness analysis (see Section S9).

#### S2.2 Health impact and cost model

Generally, the health impact and economic analysis was carried out in a structurally similar manner as previous publications while adapting as appropriate for the Ugandan context [S16, S17, S18].

- Parameters for costs are different and adapted for Uganda
  - except for the cost of the medication and physical tests that are shipped to Uganda, these have remained the same.
- Parameters for screening and vector control (VC) activities are specific for Uganda
  - parameters for treatment duration or treatment success are the same as previous papers for DRC and Chad.
- The cost function of screening has changed according to information from the national programme. Whereas previous functions did not have awareness-raising activities factored in, the costs for passive, active, and reactive screening (PS, AS, and RS, respectively) account for awareness-raising, since this was included in the program's yearly budgets. Within passive screening (PS) confirmation is construed as a cost of the clinics or the mission, rather than a function of the number of false positives of either activity, as was done in the DRC and Chad, because the 2023 NSSEP budget gave items for confirmation (materials, transport) as a function of the number of clinics rather than as a function of the RDTs performed and the RDT-positives that resulted.

- Separately, we assumed the AS and RS mission budget included confirmation, instead of calculating it as a function of the expected false-positive CATT screens.
- Whereas the previous VC cost functions were scaled by linear kilometres of river, these functions are now scaled by square kilometres.
- The structure of the cost function of the treatment is the same, except that the confirmation and staging (microscopy and lumbar puncture) is not included in the price because confirmation and staging costs are included in the PS and AS costs separately.
  - Health effects (to calculate DALYs) are modelled with the same function as previously.

### S3 Study area

#### S3.1 Administrative boundary

The gHAT-endemic area in northern Uganda consists of a number of counties. Over time, the definition of districts in this area has changed as one or more counties have been moved into new districts, see Figure S1. Figure S1 was drawn assuming that the GADM 3.6 county definitions were constant during the period reviewed [S20] and hence that modifications to districts are purely the result of the allocation of counties to districts as described online [S21, S22, S23, S24, S25].

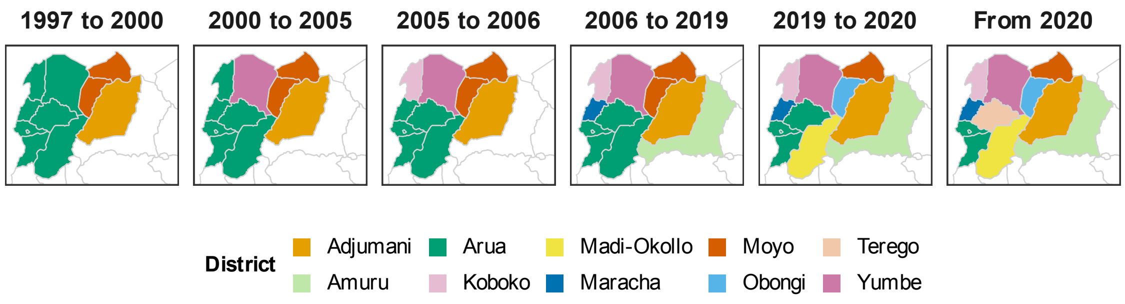

Figure S1: **Evolution of the districts of northern Uganda.** Counties are coloured based on membership of one of the named seven endemic districts. Shapefiles used to produce these maps are available under an academic publishing permitting license at https://gadm.org/download_country_v3.html

The following changes to district definitions took place in the gHAT-endemic area:

**1997** Eastern Moyo County formed Adjumani District [S20, S22].

**2000** Arunga County was given district status as Yumbe District [S21].

**N.B.** Another source gave 2005 as the creation year of Yumbe District [S24], but that was not supported by a map of districts as at the 2002 census [S26].

**2005** Koboko County, from Arua District, was given district status [S24].

**2006** Maracha County, formerly part of Arua District, given district status [S24].

**N.B.** The proposal had been for Maracha and Terego counties to form Maracha (AKA Maracha-Terego or Nyadri) District, however following political disagreement Terego County eventually decided to remain as part of Arua District – hence Maracha-Terego has been ignored in preparing Figure S1.

Kiluk County, formerly part of Gulu District, was given district status as Amuru District [S23].

**2019** Madi-Okollo, formerly part of Arua District, and Obongi, formerly part of Moyo District, became districts themselves[S24].

**2020** From July 2020 Terego (east of Maracha), formerly part of Arua District, became a districts[S24].

The fourth panel of Figure S1, spanning 2006 to 2019, covers the majority of the period for which we have gHAT data (i.e. 2000–2022) - either from the WHO HAT Atlas or from the Trypa-NO! project (see Section S5 below). Therefore, we will use this district definition as the basis of our district level analysis. Geolocations of case records are used to allocate district independent of the district names which may have been recorded at the time of diagnosis or treatment.

##### S3.2 Demography

District population sizes and their growth rates used in this study are provided in Table S1.

Table S1: **Population size and growth rate used in each district.**

| District | Population size* | Population growth rate** |
|---|---|---|
| Adjumani | 225,251 | 1.009 |
| Amuru | 186,696 | 1.027 |
| Arua | 782,077 | 1.027 |
| Koboko | 206,495 | 1.04 |
| Maracha | 186,134 | 1.021 |
| Moyo | 139,012 | 1.025 |
| Yumbe | 484,822 | 1.056 |

* Population numbers were estimated from the 2014 census [S27, S28].

** Population growth rates per year were estimated from 2002 to 2014 (1991 to 2014 for Moyo district) using a combination of citypopulation.de, and the 2002 and 2014 Uganda census results [S26, S27, S28].

#### S4 Study period

The history of gHAT in Uganda has been detailed elsewhere, but here we provide a brief sketch, in Figure 1, of the major events that pertain to our analysis.

In Uganda, gHAT screening resumed after the colonial and civil war periods in 1986 with the help of MSF [S29]. By 2000, screening operations were handled by the national control program.

In Section S5 we will detail the human case data, spanning from 2000-22, which were used for our fitting analysis. In Section S8 we detail the vector control (VC) activities and how we take those activities into account in our model. More resources were put in place for PS in 2012-13, as detailed in Figure 1 in the paper and in [S30]. Our CEA spans from 2026-2040, though our main analysis will have an economic horizon until 2040, and alternative horizons will be available in the `GUI`.

#### S5 Human case and active screening data

##### S5.1 WHO HAT Atlas data

Data from the WHO HAT Atlas for 2000–2017 were provided in an Excel spreadsheet. There were $3\,956$ rows in this file, in which 3927 entries have geolocations and 1712 entries have complete residence names (district, county, subcounty, parish). To aggregate these data by district, we assumed that the geolocations recorded in the WHO HAT Atlas were correct and dropped 26 records that were not in the seven endemic districts in Uganda. For 31 records without geolocations, we used residence names to assign their districts in the aggregated data, which applied to 17 records. Of these 3918 records, 34 are identified as originating in South Sudan epidemiological data files and 2 are from a file of diagnoses outside the endemic districts. Because there was limited information on these imported and exported cases, we kept them in our analysis to incorporate the contribution to local transmission while they were present in the endemic regions in Uganda.

Table S2 shows the count of rows remaining in the WHO HAT Atlas by surveillance type and whether the number screened is known, by year. The number screened for active surveillance was absent for much of the early data period, however, these values have been supplied by the Ugandan National Sleeping Sickness Programme, as far as possible. Two classes of rows will be deleted:

- the single row that purports to be a Passive screening record, but has 23 people screened but no cases detected; and
- the 124 rows where the surveillance type is unknown, these rows report 197, 1 and 55 gHAT cases in 2000, 2007 and 2010, respectively.

Table S2: **Number of records (rows) in the Uganda HAT Atlas data.** Records with or without a recorded number screened by year. N.B. 2016 and 2017 HAT Atlas data were not used in our model fitting.

| **Surveillance:** | | Active | | Passive | | Unknown |
|---|---|---|---|---|---|---|
| **# Screened:** | | No | Yes | No | Yes | Unknown |
| **Year:** | 2000 | 68 | | 261 | | 68 |
| | 2001 | 44 | | 397 | | |
| | 2002 | 120 | | 362 | | |
| | 2003 | 67 | | 423 | | |
| | 2004 | 42 | | 318 | | |
| | 2005 | 45 | | 298 | | |
| | 2006 | 75 | | 153 | 1 | |
| | 2007 | 43 | | 103 | | 1 |
| | 2008 | 81 | 202 | 108 | | |
| | 2009 | 20 | 8 | 78 | | |
| | 2010 | 3 | 174 | 32 | | 55 |
| | 2011 | 4 | 129 | 26 | | |
| | 2012 | | | 21 | | |
| | 2013 | | | 8 | | |
| | 2014 | | | 9 | | |
| | 2015 | | | 4 | | |
| | 2016 | | 25 | 4 | | |
| | 2017 | | 38 | | | |

There are then 3 793 rows remaining for aggregation by district, surveillance type and year. Figure S2 shows the number of geolocated active and passive cases aggregated across years and Figure S3 shows geolocated new cases (both active and passive) within year.

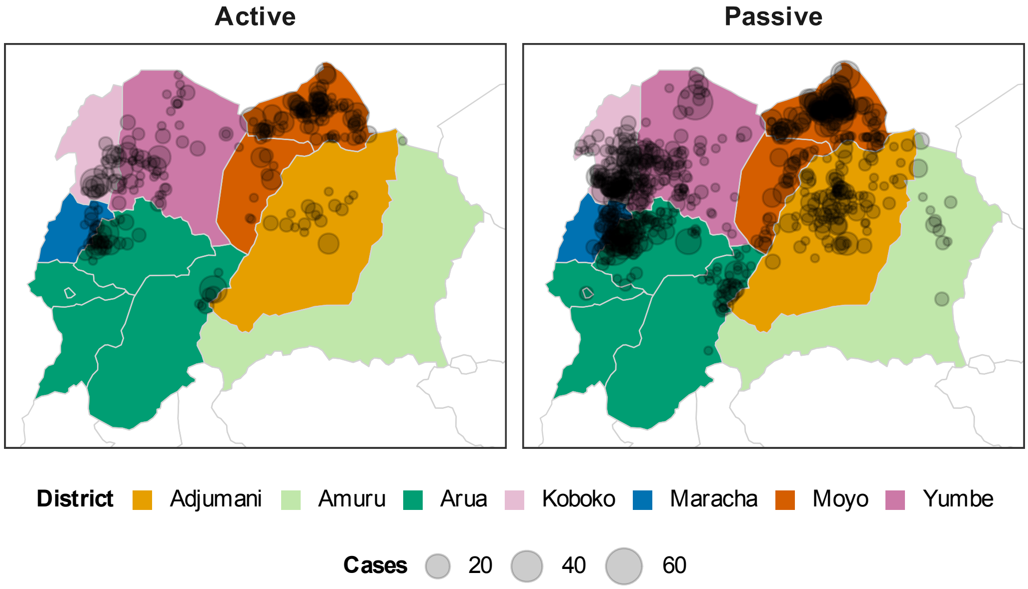

Figure S2: **Cases detected by geolocation for active and passive screening 2000–2015.**

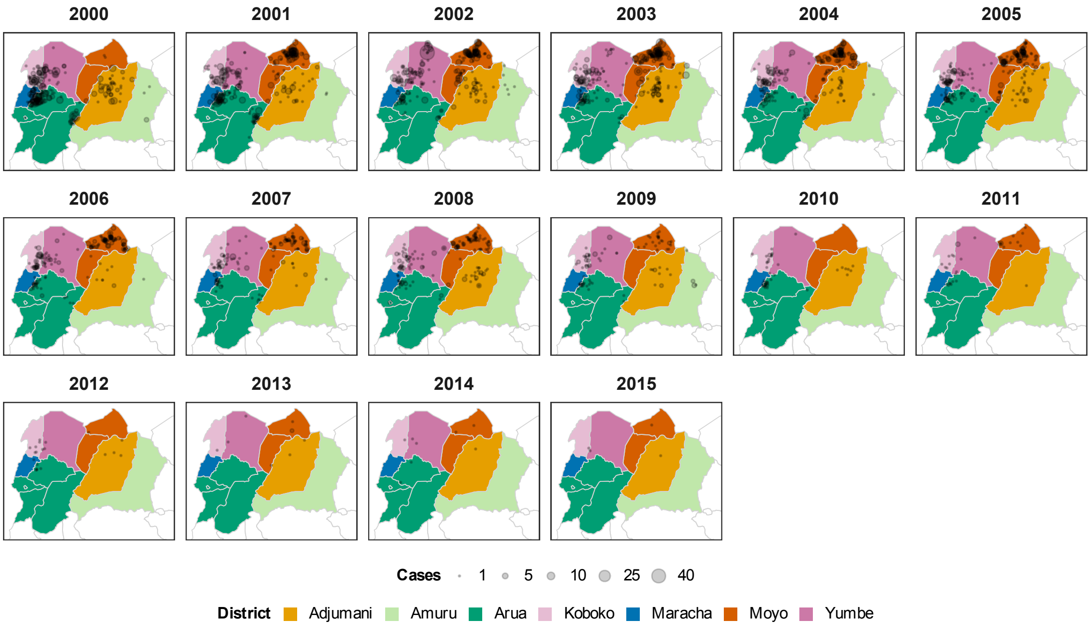

Figure S3: **New cases detected by geolocation within the year.**

##### S5.2 Missing screening data and updated case data

Missing screening information in 2000–2011 in the WHO HAT Atlas data (summarised in Table S2) caught our attention after the data processing procedure described above (see Section S5.1). The National Sleeping Sickness Programme (NSSCP) in Uganda, provided aggregated missing screening data by district and year from the original paper records and updated corresponding case data. There were 40 district-year screening data and 16 active or passive case data were updated. Followed by NSSCP, the Trypa-NO! project also updated 5 screening and 2 case aggregated data.

##### S5.3 Supplemental data from Ugandan National Sleeping Sickness Programme (NSSCP) via Trypa-NO! project

The WHO HAT Atlas data ends in 2017. The Trypa-NO! project provided completed screening and case data (both active and passive detections, including staging information) recorded in 2012–2022. We combined both data sets, 2000–2015 from the WHO HAT Atlas data and 2016–2022 from the Trypa-NO! data, to form the 2000–2022 data for our analysis.

##### S5.4 Exclude imported cases after VC started

We believe the imported cases from South Sudan acquired their infections in South Sudan but then could create local transmissions in Uganda after settling down in Uganda and before diagnosis. In this paper, we are interested in the future dynamics and predictions of local transmission and elimination. The proportion of imported cases has gone up substantially from 4.3% before VC started (2000–2012) to 24.2% after VC. Unlike old imported cases, recent imported cases are less likely to create local transmission because VC reduced the number of tsetse and therefore suppressed transmission. For this reason, we excluded imported cases detected after VC rolled out in each district to improve the accuracy of future projections. To be specific, we removed 3 cases from Moyo reported in 2015, 2018, and 2020 respectively and 2 from Yumbe reported in 2017 and 2020.

#### S6 Epidemiological model description

##### S6.1 Model variants

The compartmental gHAT infection model (Figure S4) and its equations (Eqn S6.1) have been presented elsewhere previously [S5, S9, S14] for the model variants. Descriptions of model parameters can be found in

Tables S3 and S4.

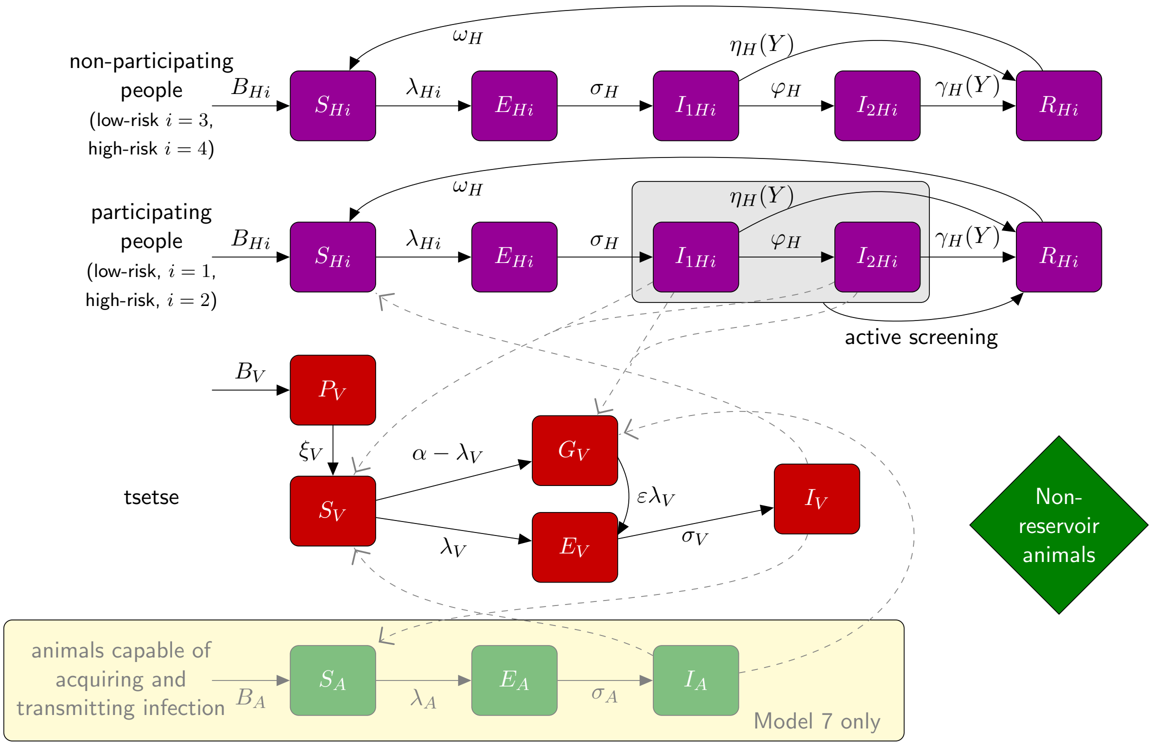

Figure S4: **Illustration of compartmental gHAT model.** Compartmental model schematic using Greek notation for rates and capital Roman characters for different host and vector infection states. Purple and red components form the non-animal baseline models and are also included in the two animal model variants. The light yellow boxes and arrows are only found in the animal model variants. Transmission pathways are shown as dashed grey lines. Births and deaths are included but are not shown here to aid readability. The grey oval and dashed black lines indicate infection classes assumed to be detectable using a traditional screen-confirm-treat approach in active screening (although some infections may still be missed due to imperfect diagnostic sensitivity). Adapted from Crump et al. [S9] under a CC-BY license.

$$
\text{Humans}\begin{cases}
\frac{\mathrm{d}S_{Hi}}{\mathrm{d}t} &= \mu_H N_{Hi} + \omega_H R_{Hi} - \alpha m_{\text{eff}} f_{Hi}\frac{S_{Hi}}{N_{Hi}} I_V - \mu_H S_{Hi}\\
\frac{\mathrm{d}E_{Hi}}{\mathrm{d}t} &= \alpha m_{\text{eff}} f_{Hi}\frac{S_{Hi}}{N_{Hi}} I_V - (\sigma_H + \mu_H) E_{Hi}\\
\frac{\mathrm{d}I_{1Hi}}{\mathrm{d}t} &= \sigma_H E_{Hi} - (\varphi_H + \eta_H(Y) + \mu_H) I_{1Hi}\\
\frac{\mathrm{d}I_{2Hi}}{\mathrm{d}t} &= \varphi_H I_{1Hi} - (\gamma_H(Y) + \mu_H) I_{2Hi}\\
\frac{\mathrm{d}R_{Hi}}{\mathrm{d}t} &= \eta_H(Y) I_{1Hi} + \gamma_H(Y) I_{2Hi} - (\omega_H + \mu_H) R_{Hi}
\end{cases}
$$

$$
\text{Animals}\begin{cases}
\frac{\mathrm{d}S_{A}}{\mathrm{d}t} &= \mu_A N_A - \alpha m_{\text{eff}} f_A \frac{S_A}{N_A} I_V - \mu_A S_A\\
\frac{\mathrm{d}E_{A}}{\mathrm{d}t} &= \alpha m_{\text{eff}} f_A \frac{S_A}{N_A} I_V - (\sigma_A + \mu_A) E_A\\
\frac{\mathrm{d}I_{A}}{\mathrm{d}t} &= \sigma_A E_A - \mu_A I_A
\end{cases}
$$

$$
\text{Tsetse}\begin{cases}
\frac{\mathrm{d}P_{V}}{\mathrm{d}t} &= B_V N_H - (\xi_V + \frac{P_V}{K}) P_V\\
\frac{\mathrm{d}S_{V}}{\mathrm{d}t} &= \xi_V \mathbb{P}(\text{pupating}) P_V - \alpha S_V - \mu_V S_V\\
\frac{\mathrm{d}E_{1V}}{\mathrm{d}t} &= \alpha\big(1 - h_{TT}(t)\big) p_V \big( \sum_i f_{Hi} \frac{(I^b_{1Hi} + x I^s_{1Hi} + I_{2Hi})}{N_{Hi}} + f_A \frac{I_A}{N_A}\big)(S_V + \varepsilon G_V)\\
&\quad -(3\sigma_V + \mu_V + \alpha h_{TT}(t)) E_{1V}\\
\frac{\mathrm{d}E_{2V}}{\mathrm{d}t} &= 3\sigma_V E_{1V} - (3\sigma_V + \mu_V + \alpha h_{TT}(t)) E_{2V}\\
\frac{\mathrm{d}E_{3V}}{\mathrm{d}t} &= 3\sigma_V E_{2V} - (3\sigma_V + \mu_V + \alpha h_{TT}(t)) E_{3V}\\
\frac{\mathrm{d}I_{V}}{\mathrm{d}t} &= 3\sigma_V E_{3V} - (\mu_V + \alpha h_{TT}(t)) I_V\\
\frac{\mathrm{d}G_{V}}{\mathrm{d}t} &= \alpha(1 - h_{TT}(t))\Big(1 - p_V\big( \sum_i f_{Hi} \frac{(I^b_{1Hi} + x I^s_{1Hi} + I_{2Hi})}{N_{Hi}} + f_A \frac{I_A}{N_A}\big)\Big) S_V\\
&\quad -\alpha\Big(h_{TT}(t) + \big(1 - h_{TT}(t)\big) p_V \varepsilon \big( \sum_i f_{Hi} \frac{(I^b_{1Hi} + x I^s_{1Hi} + I_{2Hi})}{N_{Hi}} + f_A \frac{I_A}{N_A}\big)\Big) G_V\\
&\quad -\mu_V G_V
\end{cases}
\tag{S6.1}
$$

where $i$ represents the four different possible human risk/active screening participation groups: $i = 1$ is low-risk, randomly participating group, $i = 2$ is the high-risk, randomly participating group, $i = 3$ is the low-risk, never participating group, and $i = 4$ is the high-risk, never participating group. Blood feeding on each group $f_{Hi}$ is determined by the proportion in the group and the relative risk, which is $r$-times higher for high-risk people.

Table S3: **Model parameterisation (fixed parameters).** Notation, a brief description, and the values used for fixed parameters.

| Model[§] | Notation | Description | Value | |
|---|---|---|---|---|
| All | $N_H$ | Total human population size in 2014 | Fixed for each district | [S27, S28] |
| All | $\mu_H$ | Natural human mortality rate | $4.5136\times10^{-5}$ days$^{-1}$ | [S31] |
| All | $B_H$ | Total human birth rate | $= \mu_H N_H$ | |
| All | $\sigma_H$ | Human latency rate | 0.0833 days$^{-1}$ | [S32] |
| All | $\varphi_H$ | Stage 1 to 2 progression rate | 0.0019 days$^{-1}$ | [S33, S34] |
| All | $\omega_H$ | Recovery rate or waning-immunity rate | 0.006 days$^{-1}$ | [S35] |
| All | Sens(AS) | Active screening algorithm diagnostic sensitivity | 0.91 | [S36] |
| All | $B_V$ | Tsetse birth rate (per capita rate of depositing new pupae) | Adapted for each district | See Section S8.6 |
| All | $p_{\text{targetdie}}$ | Max probability of a tsetse contacting a Tiny Target and dying per blood meal to yield the assumed/observed population reduction after one year | Adapted for each district | See Section S8.6 |
| All | $\xi_V$ | Rate of pupal development to adult flies | 0.037 days $^{-1}$ | [S37] |
| All | $K$ | Pupal carrying capacity | $= 111.09 N_H$ | Assumed[**] |
| All | $\mathbb{P}$(pupating) | Probability of a pupa surviving to emerge as an adult fly | 0.75 | [S37][***] |
| All | $\mu_V$ | Tsetse mortality rate | 0.03 days$^{-1}$ | [S32] |
| All | $\sigma_V$ | Tsetse incubation rate | 0.034 days$^{-1}$ | [S38, S39] |
| All | $\alpha$ | Tsetse bite rate | 0.333 days$^{-1}$ | [S40] |
| All | $p_V$ | Probability of tsetse infection per single infective bite | 0.065 | [S32] |
| All | $\varepsilon$ | Reduced susceptibility factor for non-teneral (previously fed) flies | 0.05 | [S41][†] |
| All | $f_H$ | Proportion of blood-meals on humans (total), $\sum_{i=1}^{4} f_{Hi}$ | 0.09 | [S42] |
| All | disp$_{\text{act}}$ | Overdispersion parameter for active detection | $4\times10^{-4}$ | Assumed[‡] |
| All | disp$_{\text{pass}}$ | Overdispersion parameter for passive detection | $2.8\times10^{-5}$ | Assumed[‡] |
| Model 6,7,8 | $\mu_A$ | Natural animal mortality rate | 0.0014 days$^{-1}$ | Assumed |
| Model 6,7,8 | $\sigma_A$ | Animal latency rate | 0.0833 days$^{-1}$ | [S32] |

[**] The value of $K$ was chosen to reflect a plausible bounce-back rate as discussed in [S43].
[***] Pupal survival computed based on a 1% per day mortality rate of pupae over 27 days.
[†] Whilst the teneral phenomenon is well known, the exact value for the reduction in susceptibility is unknown and likely depends on the age and nutritional status of fed flies [S41]. Previous modelling demonstrates that this parameter is non-identifiable in model fitting and will be highly correlated with $R_0^2$ which is fitted [S1].
[‡] Overdispersion for both active and passive screening was selected to match the typical variance in case detections at the focus level as discussed in [S7].

Table S4: **Model parameterisation (fitted parameters).** Notation, brief description, and information on the prior distributions for fitted parameters.

| **Model**§ | **Notation** | **Description** | **Prior distribution*** | **Percentiles of prior distribution** [2.5, 50 & 97.5%] | **Unit** |
|---|---|---|---|---|---|
| All | $R_0$ | Basic reproduction number (NGM approach) | $1 + \mathsf{Exp}(10)$ | [1.003, 1.069, 1.369] | - |
| Model 2,4,5,7,8 | $r$ | Relative bites taken on high-risk humans | $1 + \Gamma(3.68, 1.09)$ | [2.015, 4.654, 10.028] | - |
| Model 2,3,4,7 | $k_1$ | Proportion of low-risk people | $\mathsf{B}(16.97, 3.23)$ | [0.6564, 0.8514, 0.9609] | - |
| Model 5,8 | $[k_1, \ldots, k_4]$ | Human population classificiation† | $\mathsf{Dir}(16.97, 1.08, 1.08, 1.08)$ | $k_1 : [0.656, 0.851, 0.961]$<br>$k_{j,j\neq 1} : [0.002, 0.039, 0.182]$‡ | - |
| All | $\eta_H^{\text{post}}$ | Treatment rate from stage 1, 1998 onwards | $\Gamma(3.54, 5.32 \times 10^{-5})$ | [4.59, 17.1, 42.9]$\times 10^{-5}$ | days$^{-1}$ |
| All | $\gamma_H^{\text{post}}$ | Combined treatment and disease-induced death rate from stage 2, 1998 onwards | $\Gamma(6.21, 0.00101)$ | [2.35,5.94,12.08]$\times 10^{-3}$ | days$^{-1}$ |
| All | $\gamma_H^{\text{pre}}$ | Combined treatment and disease-induced death rate from stage 2, pre-1998 | $\Gamma(6.21, 0.00101)$ | [2.35,5.94,12.08]$\times 10^{-3}$ | days$^{-1}$ |
| All | Spec(AS) | Active screening diagnostic specificity | $0.998 + (1 - 0.998)\,\mathsf{B}(7.23, 2.41)$ | [0.9989, 0.9995, 0.9999] | - |
| All | $u$ | Proportion of stage 2 passive cases reported | $\mathsf{B}(20, 40)$ | [0.2208, 0.3315, 0.4564] | - |
| All | $d_{\text{change}}$ | Midpoint year for passive improvement | $2000 + (2020 - 2000)\,\mathsf{B}(5, 6)$ | [2003.2, 2007.7, 2012.5] | Year |
| All | $\eta_{H_{\text{amp}}}$ | Relative improvement in passive stage 1 detection rate | $\Gamma(1, 5.77)$ | [0.146, 4.00, 21.3] | - |
| All | $\gamma_{H_{\text{amp}}}$ | Relative improvement in passive stage 2 detection rate | $\Gamma(1, 11.24)$ | [0.285, 7.79, 41.5] | - |
| All | $d_{\text{steep}}$ | Speed of improvement in passive detection rate | $\Gamma(1, 0.62)$ | [0.0157, 0.430, 2.29] | years$^{-1}$ |
| Model 6,7,8 | $f_A$ | Proportion of blood meals on animals able to acquire and transmit the parasite | $\mathsf{B}(1.3, 1.3)$ | [0.046, 0.500, 0.954] | - |
| Model 6,7,8 | $k_A$ | Relative size of animal population able to acquire and transmit the parasite | $\Gamma(1.26, 19.3)$ | [1.18, 18.3, 19.3] | - |

§ See Table 1 in the main text for the definitions of eight model variants.

* Where Exp(.), $\Gamma$(.), B(.) and Dir(.)are the exponential, gamma (parameterised with shape and scale), beta and Dirichlet distributions, respectively.

† Human population classification: $k_1$ is the proportion of humans at low risk of infection that participate at random in AS; $k_2$ is the proportion of high-risk humans participating at random in AS; $k_3$ is the proportion of low-risk humans that do not participate in AS, and $k_4$ is the proportion of high-risk humans that do not participate in AS.

‡ Percentiles of marginal (beta) distributions.

#### S6.2 Improvements to passive screening

We assumed an improvement in passive case detection rates in all seven endemic districts in Uganda.

$$\eta_H(Y) = \eta_H^{\text{post}} \left[ 1 + \frac{\eta_{H_{\text{amp}}}}{1 + \exp\left(-d_{\mathsf{steep}}(Y - d_{\mathsf{change}})\right)} \right] \tag{S6.2}$$

$$\gamma_H(Y) = \gamma_H^{\text{post}} \left[ 1 + \frac{\gamma_{H_{\text{amp}}}}{1 + \exp\left(-d_{\mathsf{steep}}(Y - d_{\mathsf{change}})\right)} \right] \tag{S6.3}$$

We assume that all stage 1 cases are reported, but that some of the exits from stage 2 are due to death from gHAT disease outside healthcare. In 1998 the reporting probability for an exit from stage 2 is given by $u$, however, as the exit rate from stage 2 increases this reporting probability does not stay constant, but increases (proportionally more people would be detected and treated with higher exit rates). When we compute reporting rates from stage 2 we therefore use the following:

$$\text{Death rate} = (1 - u)\gamma_H^{\text{post}} \tag{S6.4}$$

$$\text{Stage 2 reporting incidence} = (\gamma_H(Y) - \text{Death rate})I_2 \tag{S6.5}$$

where $I_2$ is the total population in stage 2.

The parameters associated with recent improvements to passive screening in Uganda were difficult to estimate. There are few non-stage 2 passive cases in recent years with case reporting. In geographies with ongoing transmission at relatively high levels of cases, the staging ratio is a good indicator of improvement in PS.

#### S6.3 Improvements to active screening diagnostic specificity

During the fitting procedure, we estimated the district-specific active screening specificity based on historical case reporting between 2000–2022. Whilst specificity is assumed to be very high (>99.8%) the high number of historical active screening in some districts means that it is very likely that some false positives were incorrectly confirmed. When case numbers decrease, each individual serological positive sample receives more attention in the diagnosis process to ensure treatments are only given to true positives. According to our case and screening data, the positivity in AS dropped from 0.001 to 0.0001 in 2011. As a result, we increased our modelled active screening algorithm specificity to 100% in all districts in Uganda in 2011.

### S7 Model fitting procedure

An adaptive Metropolis-Hastings Markov chain Monte Carlo (MCMC) algorithm was used to fit deterministic variants of the Warwick HAT model, a compartmental transmission model, to epidemiological data as in previous modelling studies outside Uganda [S7, S9, S5]. The model variants differ in the classification of the human population and the presence or absence of a population of animals contributing to transmission. Model fitting was carried out independently within each district.

For any given model parameterisation, we can analytically compute the model's endemic equilibrium by setting all of the rates of change to zero, which means no changes for each compartment in time. This is the same as setting the left-hand side of the model equations S6.1 to zero and solving these simultaneous equations. This gives us the number of susceptibles, infected people, flies, etc. at endemic equilibrium in terms of the model parameters. See file GetEndemicEq.m on `OSF`. Our ODE models were then run from this endemic equilibrium for each selected parameter set in our MCMC procedure. Two chains were run in the MCMC, and they were initialised using the fixed parameters and by random perturbations around supplied, individually valid, initial values of each parameter being fitted, rejecting those parameter sets that do not produce a valid posterior probability.

Continuing with a standard adaptive MCMC algorithm, we keep picking new parameter sets and assess the goodness-of-fit of each using the log-likelihood function (described below in Section S7.1). We keep parameter sets which produce results which have a higher probability of the model looking like the observed data. We continue this process for a minimum of 100,000 steps in the MCMC chain, checking that we have converged by confirming the Gelman-Rubin statistic is less than 1.05 and that we reach an effective sample size (ESS) of at least 1000. Our final results for each model variant are 2000 district-specific posterior parameter sets generated by us homing in on specific parameters which can best reflect the dynamics observed for each district in Northern Uganda.

#### S7.1 Likelihood

The parameters fitted for each model are described in Table S4 and the model variant-specific subset of these are estimated independently in each district and model variant.

For fitting the model to case data, we transform model ODE solutions (Eqn S6.1) into annual case reporting denoted $A_{M1}$, $A_{M2}$, for active stage 1 and stage 2 and $P_{M1}$, $P_{M2}$, for passive stage 1 and 2. Since we always know the stage (1 or 2) in the model simulations, there is no requirement for a "U" (unknown stage) category for the model. These are computed using solutions to the ODEs for the given set of parameters aggregated across a year.

Detections relate to the transfer from infectious categories to the recovered category – the new annual reported case incidence. This is either by passive detection from stage 1 for year $Y$:

$$P_{M1}(Y) = \int_{Y}^{Y+1} \eta_H(Y)\big(I_{1H1}(t) + I_{1H4}(t)\big)\,\mathrm{d}t,$$

passive detection from stage 2

$$P_{M2}(Y) = \int_{Y}^{Y+1} (\gamma_H(Y) - \text{Death rate})\big((I_{2H1}(t) + I_{2H4}(t)\big)\,\mathrm{d}t,$$

or by AS from the low-risk ($H1$) group in year $Y$

$$A_{M1}(Y) = z(Y)\text{Sens}I_{1H1}(Y) + z(Y)(1 - \text{Spec})\big(k_1 N_H - I_{1H1}(Y) - I_{2H1}(Y)\big)$$

and

$$A_{M2}(Y) = z(Y) \times \text{Sens} \times I_{2H1}(Y)$$

with variable AS coverage by year, $z(Y)$ and fixed diagnostic sensitivity. $A_{M1}$ also contains any false positives that may have been incorrectly identified from non-infected people based on the high but imperfect specificity of the AS algorithm. We assume in Uganda that all false positives would be assigned to be stage 1 and treated, however, in the model false positives stay in the susceptible category, unlike true positives which move to the recovered category.

The log-likelihood function used in the adaptive Metropolis-Hastings MCMC contained two terms in each year for which reported case numbers were available for each source of reported cases (active or passive screening). These were:

- a beta-binomial probability that the total number of cases reported in that year for that source came from the available population (either the reported number of people actively screened for AS or the focus population for PS) with probability calculated from solving the ODE for the current set of parameters, and
- a binomial probability that the reported stage 1 cases come from the total number of reported staged cases, where the probability parameter again comes from the solution of the ODE. In many years, staging is unknown, so this part of the log-likelihood will return zero and not contribute to our calculation. In some years, we only partially know staging information.

This formulation allowed over-dispersion in the observed cases to be included, via the beta-binomial distribution, and any proportion of cases with reported disease stage to be appropriately accounted for (assuming that the reporting of staging information is independent of the disease stage). The log-likelihood function was as follows:

$$
\begin{aligned}
LL(\theta|x) = &\ \log(P(x|\theta)) \\
\propto &\sum_{i=2000}^{2016}\Bigg(\log\left[\mathrm{BetaBin}\left(A_{D1}(i)+A_{D2}(i)+A_{DU}(i);z(i),\frac{A_{M1}(i)+A_{M2}(i)}{z(i)},\mathrm{disp}_{\mathrm{act}}\right)\right] \\
&+\log\left[\mathrm{Bin}\left(A_{D1}(i);A_{D1}(i)+A_{D2}(i),\frac{A_{M1}(i)}{A_{M1}(i)+A_{M2}(i)}\right)\right] \\
&+\ \log\left[\mathrm{BetaBin}\left(P_{D1}(i)+P_{D2}(i)+P_{DU}(i);N_H,\frac{P_{M1}(i)+P_{M2}(i)}{N_H},\mathrm{disp}_{\mathrm{pass}}\right)\right] \\
&+\log\left[\mathrm{Bin}\left(P_{D1}(i);P_{D1}(i)+P_{D2}(i),\frac{P_{M1}(i)}{P_{M1}(i)+P_{M2}(i)}\right)\right]\Bigg)
\end{aligned}
$$

The model takes parameterisation $\theta$, $x$ is the data, $P_{Dj}(i)$ and $A_{Dj}(i)$ are the number of cases detected by passive or AS (of stage $j$, which may be 1, 2 or unknown, $U$) in year $i$ of the data. $P_{Mj}(i)$ and $A_{Mj}(i)$ are the number of cases detected by passive or AS (of stage $j$) in year $i$ of the model, and $z(i)$ is the number of people actively screened in year $i$. $\mathrm{BetaBin}(m;n,p,\rho)$ gives the probability of obtaining $m$ successes out of $n$ trials with probability $p$ and overdispersion parameter $\rho$. The overdispersion accounts for a larger variance than under the binomial. The probability density function of this distribution is given by:

$$
\mathrm{BetaBin}(m;n,p,\rho)=\frac{\Gamma(n+1)\Gamma(m+a)\Gamma(n-m+b)\Gamma(a+b)}{\Gamma(n-m+1)\Gamma(n+a+b)\Gamma(a)\Gamma(b)}
$$

where $a=p(1/\rho-1)$ and $b=a(1-p)/p$.

#### S7.2 Missing active screening numbers

Despite considerable effort by the national control programme, the number of people screened is not present in the data for some district-year combinations. Imputation of the number of negative active screening outcomes takes place within the MCMC fitting procedure, and the number screened is generated by adding the observed number of active screening cases.

We use a Geometric prior for the number of negative screening tests in year $t$, $A_D^-(t) \sim \mathrm{Geom}(\lambda_t)$, where $\lambda_t=\frac{1}{1+\bar{N}_t}$ and $\bar{N}_t=\frac{\sum_{j=2000,j\neq t}^{2020} N_j e^{-|t-j|}}{\sum_{j=2000,j\neq t}^{2020} e^{-|t-j|}}$, a weighted mean of the number of people screened in years other than $t$. The proposal distribution for $A_D^-(t)$ was a negative binomial distribution:

$$
A_D^-(t)|A_D(t),p(t) \sim \mathrm{NB}(A_D(t)+1, 1-(1-p(t))\,(1-\lambda_t))
$$

where the probability of active case detection, $p(t)$, was sampled from the following Beta distribution:

$$
p(t)|\theta \sim \mathrm{Beta}\left(\hat{p}(t)\left(\frac{1}{\mathrm{disp}_{\mathrm{act}}(t)}-1\right),(1-\hat{p}(t))\left(\frac{1}{\mathrm{disp}_{\mathrm{act}}(t)}-1\right)\right)
$$

and $\hat{p}(t)$ is the probability of active case detection in year $t$ calculated from the ODE outputs.

#### S7.3 Dirichlet priors for human population group proportions

The human population in the Warwick HAT model is split into between 1 and 4 groups depending on the model variant being considered. These groups consist of individuals at low or high risk of gHAT infection who either participate in active screening at random or not at all. The proportions of the population in these groups are therefore determined by up to 4 parameters ($\mathbf{k}=[k_1,k_2,k_3,k_4]$).

Previously, we have only used an informative prior for the proportion of the population at low risk of infection and participates in active screening at random, $k_1 \sim \mathrm{Beta}(16.97, 3.23)$. In Models 5 and 8 for $k_i$, where $i=2,3,4$, we had been using $k_i \sim \mathrm{Beta}(1,1)$ (i.e. uniform over the parameter space), this did not account for the relationship between the 4 parameters.

##### S7.3.1 The Dirichlet distribution

The Dirichlet distribution is a multivariate version of the Beta distribution. For $K \geq 2$ categories, the probability density function of the Dirichlet distribution with parameters $\alpha_1, \ldots, \alpha_K$ $(\alpha_i > 0)$ is defined as:

$$f(x_1, \ldots, x_K) = \frac{\Gamma\left(\sum_{i=1}^{K} \alpha_i\right)}{\prod_{i=1}^{K} \Gamma(\alpha_i)} \prod_{i=1}^{K} x_i^{\alpha_i - 1}$$

The Dirichlet distribution is supported where $x_i \in [0,1]$ and $\sum_{i=1}^{K} x_i = 1$. It is a commonly used prior distribution.

##### S7.3.2 Choice of prior parameters for k

Table S5 shows the different combinations of $k_1, \ldots, k_4$ present in each variant of the Warwick HAT model.

Other than for Models 5 and 8, combination 5 in Table S5, we retained the beta prior for $k_1$ used in our previous studies. This would be equivalent to a two-parameter Dirichlet.

For Models 5 and 8, we chose a four-parameter Dirichlet prior.

Table S5: **Combinations of population proportion parameters in each model variant.**

| Combination | Models | $k_1$ | $k_2$ | $k_3$ | $k_4$ | Note |
|---|---|---|---|---|---|---|
| 1 | M1, M6 | ✓ | | | | Fixed $k_1 = 1$ |
| 2 | M2 | ✓ | ✓ | | | |
| 3 | M3 | ✓ | | ✓ | | |
| 4 | M4, M7, M9 | ✓ | | | ✓ | |
| 5 | M5, M8 | ✓ | ✓ | ✓ | ✓ | |

The marginal distribution of $X_i$ is $X_i \sim \mathrm{Beta}\left(\alpha_i, \sum_{j=1}^{K} \alpha_j - \alpha_i\right)$. Given that we only had a univariate informative prior for $k_1$, ensuring that our Dirichlet's marginal distribution matched the previous Beta prior distribution was straightforward and appealing. This was achieved for combination 5 by setting $\alpha_1 = 16.97$ and $\sum_{i=2}^{4} \alpha_i = 3.23$. As we have no information on any of the other parameters, we may assume that $\alpha_2 = \alpha_3 = \alpha_4 = 3.23/3$, i.e. the prior chosen for $\mathbf{k} = [k_1, k_2, k_3, k_4]$ was $\mathrm{Dir}(16.97, 1.08, 1.08, 1.08)$.

#### S7.4 Model evidence and ensemble

This section has been adapted from the Supplementary Information of Antillon et al. [S18].

The samples from the joint posterior distributions for the eight model variants, the results of the MCMC fitting to historical data, were combined into an ensemble model joint posterior, with the proportion taken from each model based on the relative model evidence ($R$). The number of samples taken from each joint posterior was a random sample from the multinomial distribution $\mathrm{Multinomial}(2000, R)$. For each of the $2\,000$ samples in the ensemble joint posterior, 10 realisations of a stochastic version of our models were projected into the future.

The model evidence, or marginal likelihood, for each model was estimated using an importance-sampled estimator [S44]. The relative model evidence for the $i^{\text{th}}$ model was then the model evidence for model $i$ divided by the sum of the individual model evidence values of the eight models.

The joint distribution of $(\boldsymbol{\theta}_m, \mathbf{x})$, for parameters $\boldsymbol{\theta}_m = \left(\theta_1, \theta_2, \ldots, \theta_{d_m}\right)$ of model $m$ and data $\mathbf{x} = (x_1, x_2, \ldots, x_n)$ satisfies

$$\pi(\boldsymbol{\theta}_m|\mathbf{x})\, \pi(\mathbf{x}|\mathbf{m}) = \pi(\mathbf{x}|\boldsymbol{\theta}_m)\, \pi(\boldsymbol{\theta}_m)\,, \qquad \text{(S7.1)}$$

where $\pi(\boldsymbol{\theta}_m|\mathbf{x})$ is the joint posterior distribution of parameters $1 \ldots d$, $\pi(\mathbf{x}|\mathbf{m})$ is the marginal likelihood or *evidence*; $\pi(\mathbf{x}|\boldsymbol{\theta}_m)$ is the likelihood, and $\pi(\boldsymbol{\theta}_m)$ is the prior distribution.

By use of MCMC methods to investigate the posterior distribution of the parameters, calculation of $\pi(\mathbf{x}|\mathbf{m})$ is avoided. Calculation of the evidence for use in model comparison requires computing the integral:

$$\pi(\mathbf{x}|\mathbf{m}) = \int \pi(\mathbf{x}|\boldsymbol{\theta}_m)\,\pi(\boldsymbol{\theta}_m)\;\mathrm{d}\boldsymbol{\theta}_m \tag{S7.2}$$

$$= \int \pi(\mathbf{x}|\boldsymbol{\theta}_m)\,\frac{\pi(\boldsymbol{\theta}_m)}{q(\boldsymbol{\theta}_m)}q(\boldsymbol{\theta}_m)\;\mathrm{d}\boldsymbol{\theta}_m \tag{S7.3}$$

Equation S7.2 cannot be calculated analytically except for some small set of tractable models. It can, however, be rewritten as equation S7.3, where $q(\boldsymbol{\theta}_m)$ is a $d_m$-dimensional probability density function. From this, an importance sampled estimator of $\pi(\mathbf{x}|\mathbf{m})$ is:

$$\hat{P}_q = \frac{1}{N}\sum_{i=1}^{N}\pi(\mathbf{x}|\boldsymbol{\theta}_{m,i})\,\frac{\pi(\boldsymbol{\theta}_{m,i})}{q(\boldsymbol{\theta}_{m,i})}, \tag{S7.4}$$

where the $\boldsymbol{\theta}_{m,i}$ are $N$ samples drawn from $q$.

A defence mixture [S45] was used for $q(\boldsymbol{\theta}_m)$:

$$q(\boldsymbol{\theta}_m) = p\phi(\boldsymbol{\theta}^*_m; n, \boldsymbol{\mu}_1 \dots \boldsymbol{\mu}_n, \mathbf{C}_1 \dots \mathbf{C}_n)\left|\frac{\boldsymbol{\theta}^*_m}{\boldsymbol{\theta}_m}\right| + (1-p)\,\pi(\boldsymbol{\theta}_m) \tag{S7.5}$$

where $\phi(\cdot)$ is a mixture of $n$ multivariate Gaussian distributions with vectors of means $\boldsymbol{\mu}_j$ ($j = \{1 \dots n\}$), and covariance matrices $\mathbf{C}_j$, $\left|\frac{\boldsymbol{\theta}^*_m}{\boldsymbol{\theta}_m}\right|$ is the Jacobian transformation relating probability on transformed and original scales, and $p$ is a mixing proportion ($p = 0.95$ was chosen for use, being a typical value [S44]).

In each of our district-level MCMC analyses of each of the models $2\,000$ samples from the joint posterior distribution were generated and stored, and $\phi(\boldsymbol{\theta}_m; n, \boldsymbol{\mu}_1 \dots \boldsymbol{\mu}_n, \mathbf{C}_1 \dots \mathbf{C}_n)$ for each district and model was chosen using the Matlab `fitgmdist` function, selecting $n$ based on Akaike's Information Criterion (AIC). To account for the high correlations between some of our model parameters, regularisation was applied to ensure that the covariance matrices, $\mathbf{C}_k$, would be positive semi-definite. Before passing to `fitgmdist`, transformations were applied to the posterior samples to put them in the range $(-\infty, \infty)$ – appropriate for Gaussian distributions – followed by scaling and centring to keep the regularisation consistent across analyses, at least at the simple, single overall covariance matrix level.

Having defined $\phi(\cdot)$ for a given analysis (district-model combination), $\hat{P}_q$ was calculated (equation S7.4) using $N = 20\,000$ samples drawn from $q(\boldsymbol{\theta}_m)$. Note that this is an increase from the $2\,000$ samples used in a previous study [S9], and is expected to reduce the possible influence of sampling on the model evidence results.

### S8 Tsetse population modelling

As detailed in section S6, our transmission model includes explicit modelling of the tsetse population. This allows us to directly quantify the effect of VC interventions by modelling their effect on the fly population and including this into our transmission model.

#### S8.1 History of vector control in Uganda

In Uganda, VC was put in place in the form of Tiny Targets, which were deployed over all seven endemic districts we are modelling. VC started with small pilot areas in Arua and Maracha from 2011–2012, before expanding to cover a large contiguous area covering most of Maracha and western Arua in early 2013, then expanding further to cover the majority of the Arua, Maracha, Moyo, Koboko, and Yumbe health districts through 2015–2016 [S46]. The precise start and end dates of target deployment activity and the assumptions we have used in our modelling are detailed in Table S6. We have used historical case data from 2011–2021 to estimate the coverage of VC at each point in time. We use this to adjust our fitted parametrisation of VC effectiveness, as detailed below in Section S8.4.

#### S8.2 Entomological data

We have data on tsetse population taken from surveys in three of the seven districts we are modelling. We used this to more precisely parameterise our VC model in the zones where this data was available, and we were able to fit. The data were provided by LSTM, and were the raw data from Hope et al. [S46]. The data were separated by the three phases of VC deployment detailed in the paper. The data in the first phase were divided and allocated to districts based on which block they belonged to. The Koboko block was allocated to Koboko district, the Arua and Aiivu blocks were allocated to Arua district and the Inve and Ayi blocks were allocated to Maracha district. The Kubala block spanned between Arua and Maracha, so was not used in the fitting of either district. For the data in the second phase, we filtered the data points to sites that were also present in the first phase, and matched them to the same district as in phase 1. For the data in the third phase, we used all data points that were marked in the data as "in" the intervention area. The data for phase 3 came pre-tagged with their district.

For Maracha, we used data from all 3 phases. For Arua and Koboko, there were no sites in the data for phase 2 that were also in phase 1, so only data from phases 1 and 3 were used. Data were also available for Phase 3 only in Yumbe but these were insufficient for robust model fitting.

#### S8.3 Timing of VC activities across districts

Table S6 shows the history of VC in seven gHAT endemic districts. Differences between the trial start dates (not mentioned in the table) and larger-scale start dates (See the second column in Table S6) for VC should be noted for Arua, Koboko, and Maracha due to initial small-scale trial deployments. Start dates for VC modelling (See sixth column in Table S6) were selected based on the date VC deployment reached a larger scale (larger scale start date) before further expansion. Because of limited capture data during the scaling-down phase, we assumed the first scale-down did not reduce the case coverage of VC and used one scale-down date instead of considering multiple scaling-downs.

Table S6: **Timing and modelling assumptions of vector control (VC) interventions across each district.**

(a) Deployment history

| Deployment | Start date | Affected districts |
|---|---|---|
| Phase 1 (five 7x7km blocks) | Nov 2011 | Arua, Maracha |
| Phase 2 (500km$^2$ area around phase 1 blocks) | Nov 2012 | Arua, Koboko, Maracha |
| Phase 3 (2500km$^2$ area over NW Uganda) | Nov 2014 | Arua, Koboko, Maracha, Moyo, Yumbe |
| Phase 4 (extended to Adjumani & Amuru and some expension in the five original districts) | June 2017 | All seven districts |

(b) Modelling assumptions

| District | Data | | | | Modelling assumption | |
|---|---|---|---|---|---|---|
| | Start date | Scale up date | First scale down date | Full scale down date | Actual Start date | Scale down date |
| Adjumani | June 2017 | | | 2023 | 2017.5 | 2023 |
| Amuru | June 2017 | | | 2023 | 2017.5 | 2023 |
| Arua | Dec 2011 | Nov 2014 & June 2017 | mid-2019 | 2023 | 2012 | 2023 |
| Koboko | Dec 2012 | Nov 2014 & June 2017 | mid-2020 | 2023 | 2015 | 2023 |
| Maracha | Dec 2011 | Dec 2012 & June 2017 | mid-2019 | mid-2020 | 2012 | 2020.5 |
| Moyo | Jan 2014 | June 2017 | mid-2021 | 2023 | 2014 | 2023 |
| Yumbe | Nov 2014 | For refugee: Nov 2017 (two deployments) & June 2020 (single deployment) | | 2023 | 2015 | 2023 |

#### S8.4 Case coverage of VC activities across districts

The Trypa-NO! project provided yearly watershed coverage data for historical VC, which is the percentage of water bodies covered by VC implementations each year. In Uganda, VC had been planned out to cover areas where infections of reported cases might take place. Thus, we assumed VC with the maximum watershed coverage in each district covered 100% reported cases, i.e. reached 100% case coverage. We then used the average watershed coverage across all VC years to estimate the average case coverage of VC in each district. The range of watershed coverage and average case coverage in each district are provided in Table S7.

Table S7: **Watershed and average case coverage of historical VC in each district.**

| District | Range of watershed coverage [min, max] | Average case coverage (value used in the model) |
|---|---|---|
| Adjumani | [9.6%, 19.65%] | 72% |
| Amuru | [2.35%, 7.63%] | 65% |
| Arua | [6.96%, 19.61%] | 63% |
| Koboko | [19.94%,44.17 %] | 74% |
| Maracha | [24.87%, 66.75%] | 69% |
| Moyo | [8.26%, 30.23%] | 69% |
| Yumbe | [13.81%, 35.93%] | 57% |

#### S8.5 Tsetse dynamic model

In our transmission model, the tsetse model is a continuous compartmental model, with compartments for pupae, susceptible teneral adults, susceptible non-teneral adults, and then three compartments for exposed adults, and one for infectious adults. Broadly, the model is based on a logistic population model with birth rate proportional to the adult population, fixed carrying capacity for pupae, and constant natural death rate. Full equations are specified in equation S6.1. In order to model the tsetse-specific parameters, the tsetse model can be simplified by combining the exposed and infectious tsetse with the adults, giving the following equations:

$$\begin{aligned}
\frac{\mathrm{d}P_V}{\mathrm{d}t} &= B_V\left(S_V + G_V\right) - \left(\xi_V + \frac{P_V}{K}\right)P_V \\
\frac{\mathrm{d}S_V}{\mathrm{d}t} &= \xi_V \mathbb{P}(\text{pupating})P_V - \alpha S_V - \mu_V S_V \\
\frac{\mathrm{d}G_V}{\mathrm{d}t} &= \alpha(1 - h_{TT}(t))S_V - \alpha h_{TT}(t) G_V - \mu_V G_V
\end{aligned} \tag{S8.1}$$

The function which describes the probability of a host-seeking tsetse both hitting a Tiny Target and dying as a result, $h_{TT}$, is time-dependent ($t$, in days) from when the targets were first deployed:

$$h_{TT}(t) = p_{\text{targetdie}}\left(1 - \frac{1}{1 + \exp(-0.068(\text{mod}(t, 182.5) - 127.75))}\right) \tag{S8.2}$$

and $p_{\text{targetdie}}$ – the maximum daily probability of contacting a Tiny Target and dying as a result. $p_{\text{targetdie}}$ modifies all the bite rates $\alpha$ in our tsetse equations to produce an additional Tiny-Target-induced mortality for tsetse. The value 182.5 reflects twice-yearly deployments of Tiny Targets, as used in Uganda [S46].

#### S8.6 Tsetse model fitting to data

The values of $p_{\text{targetdie}}$ and $B_V$ (tsetse birth rate) we used in this analysis were estimated in Arua, Koboko, and Maracha by fitting the tsetse model in Eqn S8.1 to tsetse capture data shared by LSTM as detailed in Section S8.2. Table S8 summarises the values of $p_{\text{targetdie}}$, $B_V$, and the reduction of tsetse population after one year under full coverage of VC that we used in the seven districts in Uganda that we modelled. This fitting was done by performing a maximum-likelihood estimation on these parameters using a Poisson likelihood function, summing the Poisson likelihood over all the data points for each district. This fitting was performed independently of the epidemiological fitting, and the results from this were used to inform the parameters used in the epidemiological model during the fitting process. The fitted tsetse control parameters consisted of the values $B_V$ and $p_{\text{targetdie}}$ as given above, and additionally a scale parameter which was not used in the epidemiological model to match the scale of the uncontrolled tsetse population to the trap density. The fits are shown in Figures S5, S6, and S7

In our simulations, the overall effectiveness of VC in the *MeanAS+VC* strategy and reactive VC (RVC) in all strategies in each district was weighted by its case coverage. This was done by scaling the reduction after one year by the coverage and back-calculating the necessary value of $p_{\mathrm{targetdie}}$ to give this reduction.

Table S8: **Values of VC parameters in the model in each district.**

| District | Assumed/Fitted | $p_{\mathrm{targetdie}}$ | $B_V$ | Reduction after one year |
|---|---|---|---|---|
| Adjumani | Assumed | 0.0525 | 0.0505 | 80% |
| Amuru | Assumed | 0.0525 | 0.0505 | 80% |
| Arua | Fitted | 0.0316 | 0.0478 | 64.7% |
| Koboko | Fitted | 0.0575 | 0.0764 | 63.8% |
| Maracha | Fitted | 0.0537 | 0.0563 | 75.6% |
| Moyo | Assumed | 0.0750 | 0.0505 | 90% |
| Yumbe | Assumed | 0.0750 | 0.0505 | 90% |

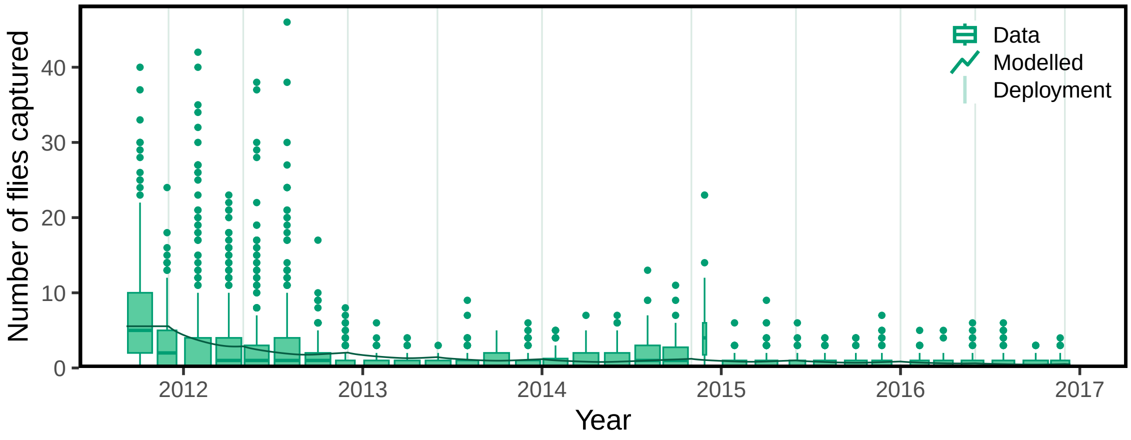

Figure S5: **Vector control fitting plot for Arua district.** Vertical lines show each deployment time. For plotting purposes only, the catch data are binned by 2-month intervals and displayed as a box plot.

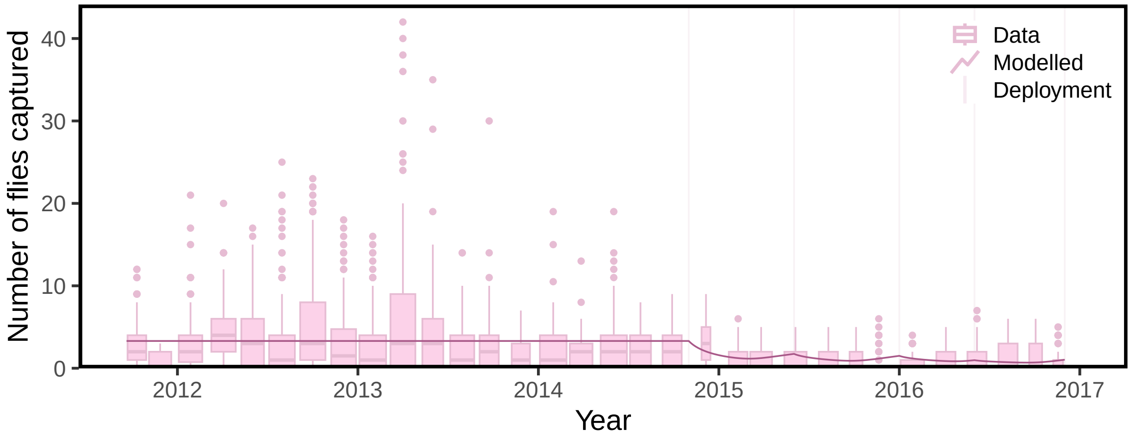

Figure S6: **Vector control fitting plot for Koboko district.** Vertical lines show each deployment time. For plotting purposes only, the catch data are binned by 2-month intervals and displayed as a box plot.

#### S8.7 Assumptions about vector control in districts without data

In the four districts without sufficient tsetse capture data to estimate the VC effectiveness and the birth rate of tsetse, we used the assumed values shown in Table S8. The capture data on the survival of Tiny Targets in the field after deployment shows differences in the proportion of functioning targets between districts. Therefore,

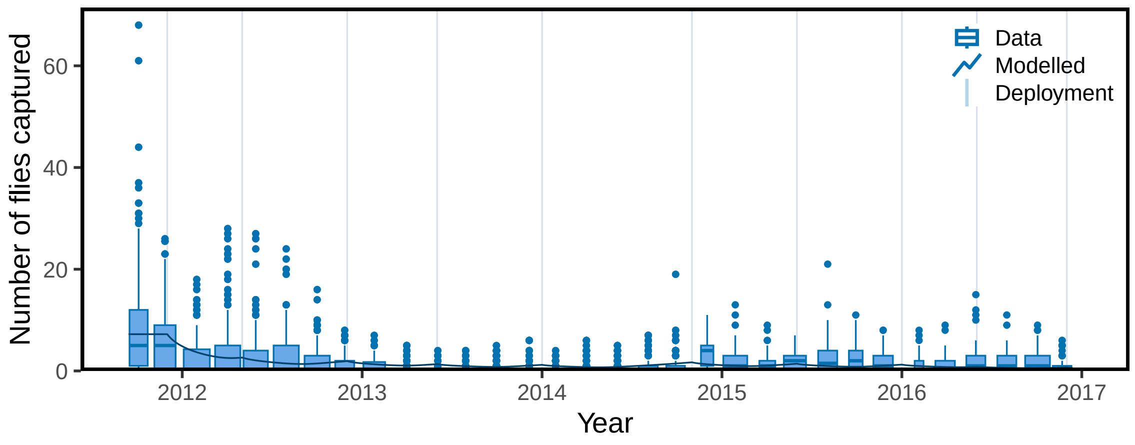

Figure S7: **Vector control fitting plot for Maracha district.** Vertical lines show each deployment time. For plotting purposes only, the catch data are binned by 2-month intervals and displayed as a box plot.

we applied the standard assumptions that we used in our previous publications [S8] in Adjumani and Amuru, and a higher VC effectiveness in Moyo and Yumbe.

#### S9 Counterfactual analysis

Counterfactual analysis is a standard modelling technique often used to compare the outcomes of the existing interventions with the outcomes that would have been achieved if the interventions had not been implemented. In this analysis, we considered one counterfactual scenario (CFS) by simulating scenarios without VC. A hash-based matching, pseudo-random number generator [S19] is introduced to our stochastic code to perform fair pairwise comparison of results between the actual strategy that occurred (with VC) and a strategy without VC. Table 6 in the main text shows the reduction in transmission and deaths attributable to VC in each district, calculated by using the following equations:

Let $I_A$ be the cumulative number of new infections since VC began in our simulations which modelled what actually occurred, and $I_P$ be the cumulative number of new infections since that same time, in our counterfactual simulations which had no VC. Then,

$$\text{\% reduction in new infections attributable to VC} = 100(1 - \frac{I_A}{I_P}).$$

The median and 95% PI can then be calculated from all of these samples (which were preserved in pairwise comparison).

This value can be calculated for each pair of matched counterfactual scenarios, as long as $I_P > 0$. If $I_P = 0$ that implies that, regardless of if VC had taken place, there would not have been any new infections. We remove these results from the list of simulations, and report the % of simulations that were excluded as there was no new infection in CFS or actual scenario.

We also compute the % reduction in mean transmission from VC using

$$\text{\% reduction in mean new infections attributable to VC} = 100\left(1 - \frac{\text{Mean}(I_A)}{\text{Mean}(I_P)}\right).$$

This is notably a different statistic than the mean % reduction that could be taken across all pairwise percentage reduction values, as the pairwise comparison is not preserved.

To aggregate our results for the whole country, we take the following steps:

1. For a district, compute a vector of $I_A$ and $I_P$.
2. For a district, compute the % reduction attributable for each realisation. Order the $I_A$ and $I_P$ vectors from most to least reduction. If percentage reductions match, sort secondly by $I_P$.

3. For all the cases where $I_P = I_A = 0$, randomly disperse these realisations amongst the sorted reductions.
4. Repeat for all districts
5. Sum the ordered $I_A$ and $I_P$ vectors to obtain new vectors for the entire country and recompute the % reduction attributable to VC. Compute median, and 95% PIs of the percentage reduction in new infections attributable to VC. Additionally, compute the percentage reduction in mean new infections attributable to VC.

#### S10 Health economic modelling

The decision analysis is diagrammed in Figure S8. The SEIRS model referred to in Figure S4 is described in Section S6 and the probability tree model for treatment is described in Section S10.3 and Fig S9.

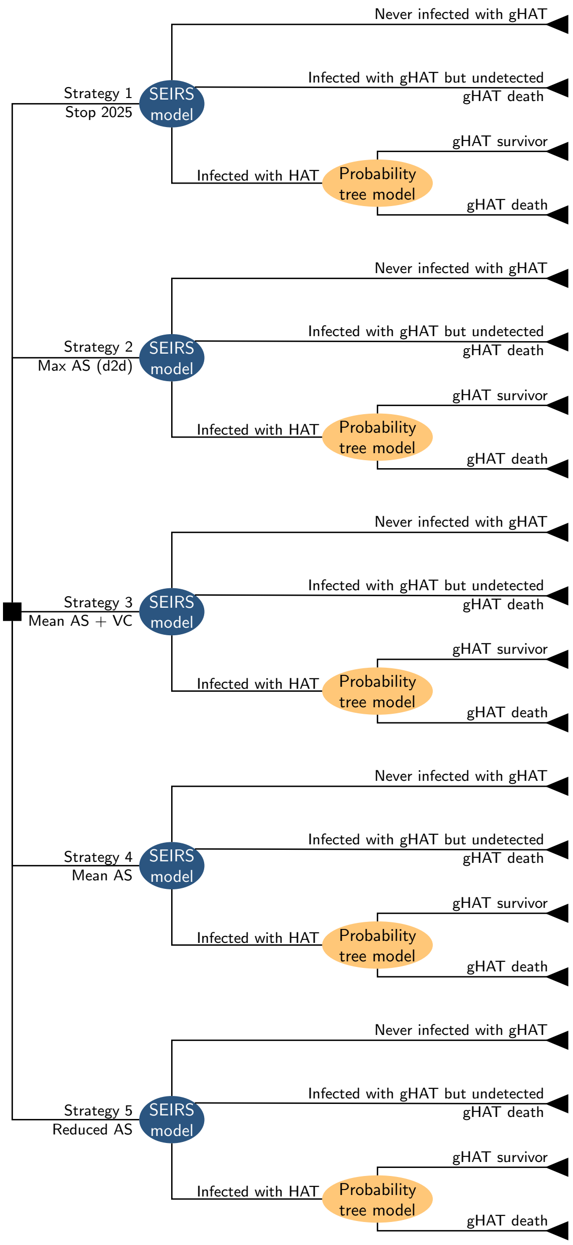

Figure S8: Decision tree of disease suppression and prevention strategies. Decision nodes are square, and probabilistic nodes are ellipses. The transmission model is depicted in Figure S4, and the probability tree model is depicted in Figure S9. Strategies against gHAT include active screening (AS) by mobile teams, passive screening (PS) in fixed health facilities, and vector control (VC). In two strategies (*MeanAS*, *MeanAS+VC*), the proportion screened equalled the mean number screened during 2018–2022 and assumed only low-risk humans would attend active screening. In one other strategy (*MaxAS(d2d)*), the coverage is the maximum number screened during 2000–2022 and assumes that humans, both at low and high risk of transmission, attend active screening campaigns. In *MeanAS+VC*, VC is simulated assuming a reduction as described in Section S8 and in the future as described in Table S9. PS is in place under all strategies. *ReducedAS* is defined as half the screening coverage of *MeanAS* and assumes that only humans at low risk of infection attend AS activities. Abbreviations: SEIRS: susceptible-exposed-infected-recovered-susceptible dynamic model of transmission, AS: active screening, PS: passive screening, VC: vector control.

#### S10.1 Parameters

Parameters are shown in Tables S9-S10 and are parameterized according to standard practice in the literature [S47]. Further details are found in Supplement 2: Parameter Glossary.

##### S10.1.1 Parameters consistent across districts

Table S9: Cost model parameters. For further details and sources, see SI Text 2: Parameter Glossary. CIs: confidence intervals. All values given to 3 significant figures. AS & PS: active and passive screening, respectively, VC: vector control, PNLTHA: Programme de Lutte contre la Trypanosomie Humaine Africaine, NECT: nifurtimox-eflornithine combination therapy, CATT: card agglutination test for trypanosomiasis, S1 & S2: stage 1 & 2 disease, DALYs: disability-adjusted life-years, SAE: severe adverse events, RDT: Rapid Diagnostic Test

| Parameter Description | Statistical Distribution | Summary Mean [95% CIs] | Sources | Section in SI Text 2 |
|---|---|---|---|---|
| **Screening parameters** | | | | |
| RDT algorithm: diagnostic sensitivity | Beta(230, 1) | 0.996 [0.948–1.000] | [S48] | ?? |
| CATT: wastage during AS | Beta(8, 92) | 0.080 [0.036–0.140] | [S49] | ?? |
| RDT: wastage during PS | Beta(1, 99) | 0.010 [<0.001–0.037] | [S50] | ?? |
| **Screening cost parameters** | | | | |
| AS capital & management costs per person screened | Gamma(245, 0.004) | $0.99 [0.88, 1.13] | ISSEP 2023 | ?? |
| RS: capital & management costs per person screened | Gamma(186, 0.009) | $1.74 [1.50, 2.00] | ISSEP 2023 | ?? |
| AS: pre-mission communications & awareness per person screened | Gamma(8.48, 0.042) | $0.36 [0.16, 0.63] | ISSEP 2019 | ?? |
| RS: pre-mission communications & awareness per person screened | Gamma(8.48, 0.050) | $0.42 [0.19, 0.75] | ISSEP 2019 | ?? |
| CATT algorithm: cost per test | Gamma(1139, 0.0007) | $0.79 [0.74, 0.83] | [S51] | ?? |
| RDT algorithm: cost per test | Gamma(1139, 0.002) | $2.51 [2.37–2.66] | | ?? |
| PS: capital & management costs of a facility (RDT only) | Gamma(25.3, 13.5) | $341 [221–486] | ISSEP 2023 | ?? |
| PS: capital & management costs of a facility (LAMP & confirmation centers) | Gamma(137, 8.53) | $1166 [979–1370] | ISSEP 2023 | ?? |
| PS: communications & awareness | Gamma(8.475, 33.3) | $282 [126, 502] | ISSEP 2019 | ?? |
| National management cost (ISSEP mark-up) | Fixed | 0.100 | ISSEP 2023 | ?? |
| **Treatment parameters** | | | | |
| Prop. of cases age<6 | Beta(153, 2430) | 0.059 [0.051–0.069] | [S52, S53] | ?? |
| Prop. of cases weight<35 kg & age>6 | Beta(8.30, 360) | 0.023 [0.010–0.040] | [S53, S54, S55, S56, S57, S58, S59, S60, S61, S62] | ?? |

| | | | | |
|---|---|---|---|---|
| Prop. of S2 cases that are severe | Beta(76.9, 44.9) | 0.634 [0.546–0.713) | [S52, S53, S54, S55, S56, S57, S58, S63] | ?? |
| Duration, treatment: pentamidine (days) | Fixed | 7 | [S63] | ?? |
| Duration, treatment: NECT (days) | Fixed | 10 | [S63, S62] | ?? |
| Duration, treatment: fexinidazole (days) | Fixed | 10 | [S63, S62] | ?? |
| Pr. of relapse: pentamidine | Beta(50.3, 665) | 0.070 [0.053–0.090] | [S52, S58, S59, S60, S64, S65] | ?? |
| Pr. of relapse: NECT | Beta(15.9, 379) | 0.050 [0.023–0.062] | [S54, S55, S56, S57, S61, S62] | ?? |
| Pr. of relapse: fexinidazole | Beta(9.49, 497) | 0.018 [0.009–0.032] | [S62, S63] | ?? |
| Pr. SAE: pentamidine treatment | Beta(1.43, 551) | 0.003 [<0.001–0.008] | [S52, S59, S60] | ?? |
| Pr. SAE: NECT treatment | Beta(40.9, 368) | 0.100 [0.073–0.131] | [S54, S55, S56, S57, S61, S62] | ?? |
| Pr. SAE: fexinidazole treatment | Beta(3.00, 261) | 0.011 [0.002–0.027] | [S62] | ?? |
| Duration, SAE (days) | Gamma (1.22, 2.38) | 2.90 [0.130–9.85] | [S66] | ?? |
| **Treatment cost parameters** | | | | |
| Hospital stay: cost per day | Gamma(5.39, 0.86) | $4.62 [1.58–9.25] | [S67, S68, S69] | ?? |
| Outpatient consult: cost | Gamma(2.48, 0.42) | $1.03 [0.17–2.66] | [S67, S68, S69] | ?? |
| Course of pentamidine: cost | Fixed | $54 | [S70] | ?? |
| Course of NECT: cost | Fixed | $360 | [S71] | ?? |
| Course of fexinidazole: cost | Fixed | $220 | | ?? |
| Drug delivery mark-up | Beta(45, 55) | 0.450 [0.354–0.548] | [S68, S69] | ?? |
| **Vector control parameters** | | | | |
| Area ($km^2$) | Fixed | 50 | Strategy definition | ?? |
| Replacement rate of targets per year | Fixed | 2 | [S72] | ?? |
| Targets per $km^2$ | Fixed | 8.5 | [S72, S73] | ?? |
| **Vector cost parameters** | | | | |
| Operational cost per $km^2$ protected | Gamma(8.48, 5.25) | 44.48 [19.76–79.05] | [S72] | ?? |
| Cost per target deployed | Gamma(8.48, 0.644) | 5.46 [2.43–9.70] | [S72] | ?? |
| **DALY parameters** | | | | |
| Age of death from infection | Gamma(148, 0.182) | 26.6 [22.4–31.1] | [S52, S53, S54, S55, S56, S57, S58, S59, S60, S61, S62, S64, S74, S66] | ?? |

| | | | | |
|---|---|---|---|---|
| Life expectancy at age of death | Interpolated | Varies by age, 45.4 [41.4–49.9] | [S75] | ?? |
| Disability weights: S1 disease | Beta(23.0, 147) | 0.135 [0.088–0.190] | [S76] | ?? |
| Disability weights: S2 disease | Beta(18.4, 15.6) | 0.541 [0.375–0.703] | [S76] | ?? |
| Disability weights: SAE | Uniform(0.037, 0.114) | 0.076 [0.039, 0.112] | [S76] | ?? |

###### S10.1.2 District-specific parameters

Table S10: District-specific cost parameters. Sources: ISSEP records. Abbreviations: AS: active screening, RDT: rapid diagnostic tests, VC: vector control

| Parameter description | Adjumani | Amuru | Arua | Koboko | Maracha | Moyo | Yumbe | Section in SI Text 2 |
|---|---|---|---|---|---|---|---|---|
| AS coverage (Reduced AS) | 1153 | 0 | 857 | 5105 | 0 | 2799 | 474 | ?? |
| AS coverage (Mean AS) | 2206 | 0 | 1714 | 10211 | 0 | 5598 | 948 | ?? |
| AS coverage (Max AS (d2d)) | 5751 | 1305 | 261118 | 30442 | 5600 | 34755 | 22188 | ?? |
| PS: number of facilities (RDT only) | 6 | 1 | 10 | 6 | 3 | 4 | 9 | ?? |
| PS: number of facilities (confirmation centers) | 2 | 1 | 2 | 1 | 1 | 4 | 2 | ?? |
| RDT per district (for all facilities) | 83 | 41 | 165 | 366 | 26 | 98 | 259 | ?? |
| VC area | 323 | 229 | 645 | 236 | 297 | 416 | 837 | ?? |
| VC targets | 1373 | 1576 | 4872 | 1688 | 3277 | 3275 | 9435 | ?? |

#### S10.2 Health burden in DALYs

The following text is largely recycled from our previous publications [S16, S17, S18], because the calculation of DALYs in this analysis remains unchanged to that in other analyses, except for the assumed life-expectancy in Uganda.

For incremental cost-effectiveness calculations, health effects are defined in terms of disability-adjusted life-years (DALYs) [S77, S67], in line with the recommendations of the Bill and Melinda Gates Foundation's reference case and WHO's guidelines for the conduct of cost-effectiveness analyses [S78, S77, S67]. DALYs allow policy-makers to compare the impact of different disease programs with one common metric.

DALYs are conventionally discounted at a rate of 3% per year [S78, S67]. We follow established conventions to calculate DALYs and evaluate the estimates in present-day terms (after applying discounting) [S79, S78, S67]. A more general discussion of this is found in [S16] (Supplementary Methods, pages 10-11), but we provide a brief description here for convenience.

Disability-adjusted life-years (DALY) = Years of life lost to disability (YLD)+Years of life lost due to death (YLL)

where,

$$\text{YLD} = (\text{YLDs before detection} + \text{YLDs during treatment} + \text{YLDs due to side effects}) \times \text{DALY weight}$$

where the DALY weight is a metric to measure the relative severity of gHAT compared to living with other diseases.

Lastly,

$$\text{YLL} = \text{life-expectancy at age of death} - \text{age of death from gHAT}.$$

We use the life-table method to calculate the life-years lost by a case, in line with our previous practice in [S18]. Although the life-expectancy at birth in Uganda is expected to be 66, the life-expectancy at the average age of death from gHAT – which is 26.62 – is 72 (See Section **??** and **??** of this supplement). Usually, life expectancy is higher at older ages than at birth, as survival to old age, conditional on survival past infancy, is higher.

Most DALYs are accrued from unidentified cases, despite the severity of symptoms of gHAT. Although few cases die of gHAT once identified (<0.01 according to Table S12), we believe ~30%-~50% cases were unidentified (see Figure S11 Section S10.3.1), and therefore those still represent the largest portion of DALYs.

#### S10.3 Treatment

The proportions of stage 1 and 2 patients that are eligible for each treatment are shown in Table S11. The outcome of treatment was modelled as a branching process, displayed in Figure S9, for which the outcome parameters are listed in Table S9. The estimates of treatment outcomes are shown in Table S12.

The following text is largely recycled from our previous publications [S16, S17, S18], because the treatment assumptions in the model remain unchanged.

Traditionally, the stage of disease is determined by microscopic examination of the cerebrospinal fluid (CSF), which is extracted via lumbar puncture for cases that have been confirmed by visualisation of the trypanosome (e.g. cases in which trypanosomes are present in the blood). If trypanosomes are present in the CSF, the patient is considered to have stage 2 disease; if not, they are considered to have stage 1 disease. According to the stage of disease, patients are referred to the appropriate health centre or health district hospital for treatment.

In the context of fexinidazole treatment, which has been present in Uganda since 2021, lumbar punctures will not be performed by the active screening team, but once patients are referred to a health centre, the health centre will determine eligibility for fexinidazole treatment and will perform lumbar punctures if fexinidazole is contraindicated.

We assumed the treatment algorithm based on the WHO interim recommendations of 2019 [S63].

- **Step 1, Group A. Patients without clinical symptoms of severe gHAT.** These patients would be eligible for fexinidazole treatment if their presentation fulfils the following criteria:

– **Patient age < 6 years or weight < 20 kg.** These patients would be ineligible for fexinidazole treatment. For simplicity, we assumed that all patients over 6 years old were also over 20 kg due to scant data on patient characteristics. See Step 2, Group A.

– **Patient age > 6 years old and weight > 20 kg.** These patients would be eligible for fexinidazole treatment. See Step 2, Group B. WHO recommendations stipulated that a doctor ought to be certain of the adherence on the part of the patient in order to prescribe fexinidazole on an outpatient basis. For simplicity, we have assumed that this is not an issue because it doesn't make a substantial difference in the total costs or effects of this particular analysis.

- **Step 1, Group B. Patients with clinical symptoms of severe HAT.** Patients whose clinical assessment would be consistent with severe gHAT (see Annex 1 of the WHO Interim guidelines [S63]) would undergo a lumbar puncture to determine the concentration of white blood cells (WBCs) in the cerebrospinal fluid. For a concentration < 100 WBC per microlitre ($\mu$L), the patient would be considered eligible for fexinidazole treatment, depending on age and weight, as detailed in Step 1, Group A. In our model, we assumed that no stage 1 patient would show more than 100 WBC/$\mu$L of CSF as no trypanosomes should be present in the CSF. Moreover, we assumed that some proportion of stage 2 patients will be in late-stage disease (see Table S11).

- **Step 2, Group A. Patients ineligible for Fexinidazole treatment.** Patients age < 6 years old or weight < 20 kg are assumed to submit to a lumbar puncture to determine disease stage with 100% adherence. In our treatment model, we consider the cost of a lumbar puncture in these patients (see Table S19) but we take the outcome of the lumbar puncture (stage 1 or stage 2) from the transmission model, the stage of disease is a critical output of the transmission model.

  – WHO recommendations stipulate the following criteria: if there are no trypanosomes in the CSF, then the patient undergoes pentamidine treatment on an inpatient basis. If there are more than 5 leukocytes (or WBC)/$\mu$L then the patient undergoes NECT treatment on an inpatient basis.

  – Pentamidine treatment would consist of intramuscular infections for 7 days, administered on an inpatient basis.

  – NECT (Nifurtimox-eflornithine combination therapy): Nifurtimox is administered orally for 10 days, while Eflornithine is administered intravenously for 7 days [S63].

- **Step 2, Group B. Patients eligible for Fexinidazole treatment.** We assumed that patients would be treated on an inpatient basis if age >6 years old and 20 kg< weight <35 kg, otherwise, they would be treated on an outpatient basis as directly observed therapy.

Table S11: Eligibility for treatment

| Eligibility | Rationale | Summary |
|---|---|---|
| **Stage 1** | | |
| Pentamidine | Under 6 years old (1) | 0.06 (0.05-0.07) |
| Fexinidazole-inpatient | Over 6 years old but under 35 kg of weight | 0.02 (<0.01-0.04) |
| Fexinidazole-outpatient | Over 6 years old and over 35 kg of weight | 0.92 (0.90-0.93) |
| **Stage 2** | | |
| NECT | Under 6 years old or late-stage disease | 0.65 (0.57-0.73) |
| Fexinidazole-inpatient | Over 6 years old but under 35 kg of weight and early stage-2 disease | <0.01 (<0.01-0.01) |
| Fexinidazole-outpatient | Over 6 years old, over 35 kg of weight, and early stage-2 disease | 0.34 (0.26-0.42) |

[1] For simplicity, all patients over 6 years old were assumed to be over 20 kg in weight.

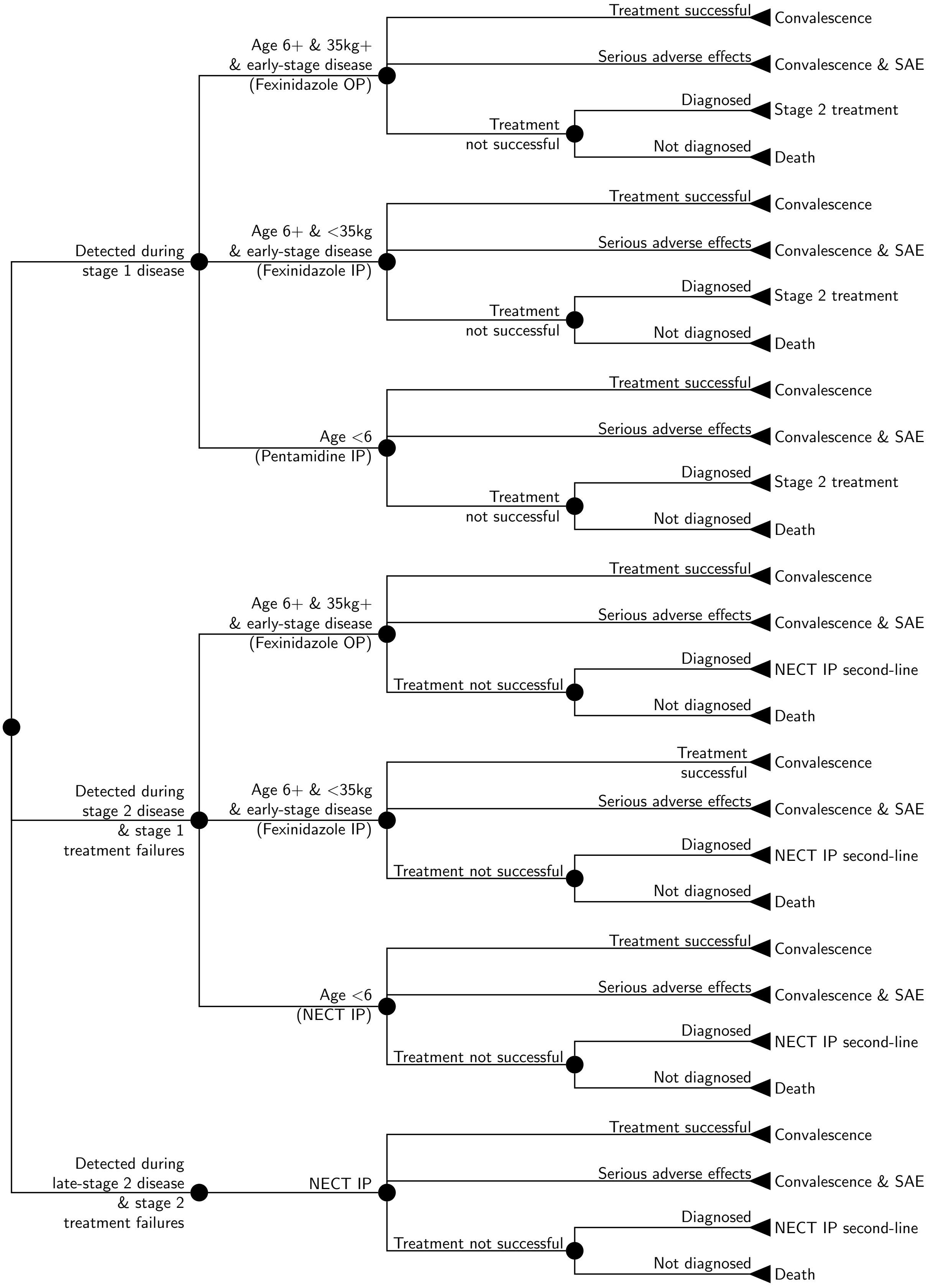

Figure S9: Treatment model. Treatment for diagnosed gHAT patients is modelled as a branching tree process of possible health outcomes, including eligibility for novel Fexinidazole since 2021. Abbreviations: SAE: Serious adverse events, IP: inpatient care, OP: outpatient care, NECT: nifurtimox-eflornithine combination therapy.

Table S12: Treatment outcomes

| Treatment | Outcomes | Estimate |
|---|---|---|
| **Stage 1** | | |
| Pentamidine | Cured | 0.05 (0.05-0.06) |
| | Cured with SAEs | <0.01 (<0.01-<0.01) |
| | Rescue treatment | <0.01 (<0.01-<0.01) |
| | Death | <0.01 (<0.01-<0.01) |
| Fexinidazole - inpatient | Cured | 0.02 (<0.01-0.04) |
| | Cured with SAEs | <0.01 (<0.01-<0.01) |
| | Rescue treatment | <0.01 (<0.01-<0.01) |
| | Death | <0.01 (<0.01-<0.01) |
| Fexinidazole - outpatient | Cured | 0.89 (0.87-0.91) |
| | Cured with SAEs | 0.01 (<0.01-0.02) |
| | Rescue treatment | 0.02 (<0.01-0.03) |
| | Death | <0.01 (<0.01-<0.01) |
| **All treatments** | **Cured** | **0.97 (0.95-0.98)** |
| | **Cured with SAEs** | **0.01 (<0.01-0.03)** |
| | **Rescue treatment** | **0.02 (0.01-0.03)** |
| | **Death** | **<0.01 (<0.01-<0.01)** |
| **Stage 2** | | |
| NECT | Cured | 0.56 (0.49-0.64) |
| | Cured with SAEs | 0.06 (0.04-0.08) |
| | Rescue treatment | 0.03 (0.01-0.04) |
| | Death | <0.01 (<0.01-<0.01) |
| Fexinidazole - inpatient | Cured | <0.01 (<0.01-0.01) |
| | Cured with SAEs | <0.01 (<0.01-<0.01) |
| | Rescue treatment | <0.01 (<0.01-<0.01) |
| | Death | <0.01 (<0.01-<0.01) |
| Fexinidazole - outpatient | Cured | 0.33 (0.25-0.41) |
| | Cured with SAEs | <0.01 (<0.01-<0.01) |
| | Rescue treatment | <0.01 (<0.01-0.01) |
| | Death | <0.01 (<0.01-<0.01) |
| **All treatments** | **Cured** | **0.90 (0.88-0.92)** |
| | **Cured with SAEs** | **0.07 (0.05-0.09)** |
| | **Rescue treatment** | **0.03 (0.02-0.05)** |
| | **Death** | **<0.01 (<0.01-<0.01)** |

##### S10.3.1 Deaths before 2026

To calculate the DALYs and deaths from 2000-25, which are featured in GUI as well as in one selection of our supplemental results, we made certain assumptions about the ubiquity of certain treatments during that period. The following is a description of the data and assumptions we took.

###### Untreated

- Before 2013, we have no records of how many people were untreated, so we assume 1% of people were untreated. Since then, we suspect that there was not 100% treatment adherence in any year, so for those years that indicate 100% treatment, we assume that 1% of cases remain untreated.

###### Stage 1

- From 2000-2012, there was no development for stage 1 drugs, so we put in that 99% of all cases were treated by pentamidine, the standard of care since the 1940s [S40]. In the 2000s there were developments for pafuramidine and in the 2010s there were trials for fexinidazole, but trials for neither drug were in Uganda. Uganda registered fexinidazole for use against both stages of gHAT in 2021, but there have been no cases og gHAT since then, so Stage 1 gHAT has only been treated with pentamidine.

###### Stage 2

- From 1948 until 2003, only melarsoprol was available for the treatment of stage 2. In the 1990's a short course was developed, so for 2000-02, 99% of cases were treated with the short-course of melarsoprol. Rescue treatment consisted of a long course of melarsoprol [S40].
- In 2003-2006, Eflornithine monotherapy was approved, but it was rarely used due to need in training and complicated protocols. Therefore, by 2006, only 20% of cases (anywhere) were treated by Eflornithine monotherapy, so we assumed that the prevalence of use was 5%, 10%, 15%, and 20% in 2003-06. The rest would be treated with the melarsoprol short-course protocol [S40].
- In 2006, the WHO prepared a kit to administer eflornithine monotherapy more easily, so its use increased to 64% by 2009 [S40], and the rest were assumed to be treated by melarsoprol. Therefore, from 2007-2009, we assumed that the percent of cases treated by eflornithine monotherapy would be 33%, 49%, 64% (essentially a linear interpolation from 20-64%).
- In 2009, nifurtimox-eflornithine combination therapy (NECT) was endorsed for use by the WHO, and they developed a kit for delivery, making NECT the dominant treatment for stage 2 starting in 2010, such that 88% of cases in that year were treated by NECT and the remaining 12% by melarsoprol [S40]. We will assume 88% of cases treated with NECT, 11% with melarsoprol, and 1% remained untreated.

###### Coverage of cases 2013-2022

- We assumed that anyone who was treated from 2012-2021 was treated by NECT for stage 2 gHAT. Treatment coverage were acquired from pg 18 of the Third Stakeholders' Meeting Report (assuming coverage for South Sudan and Uganda was equal, since it was reported for both together), page 8 of the Fourth Stakeholders' Meeting Report (assuming Uganda had the same coverage as the rest of Central Africa), and page 11 of the Fifth Stakeholders' Meeting Report (assuming Uganda had the same coverage as the rest of Central Africa). Overall, the Third report gave us numbers for 2013-2017, the Fourth Report gave us numbers for 2016-2020, and the Fifth report gave us numbers for 2018-2022 [S80, S81, S82].

Although fexinidazole was approved in Uganda in December 2021, it has not been used for any gHAT cases, since none have been reported in the country since then [S83]. Therefore, it was not assumed for any simulations before 2025.

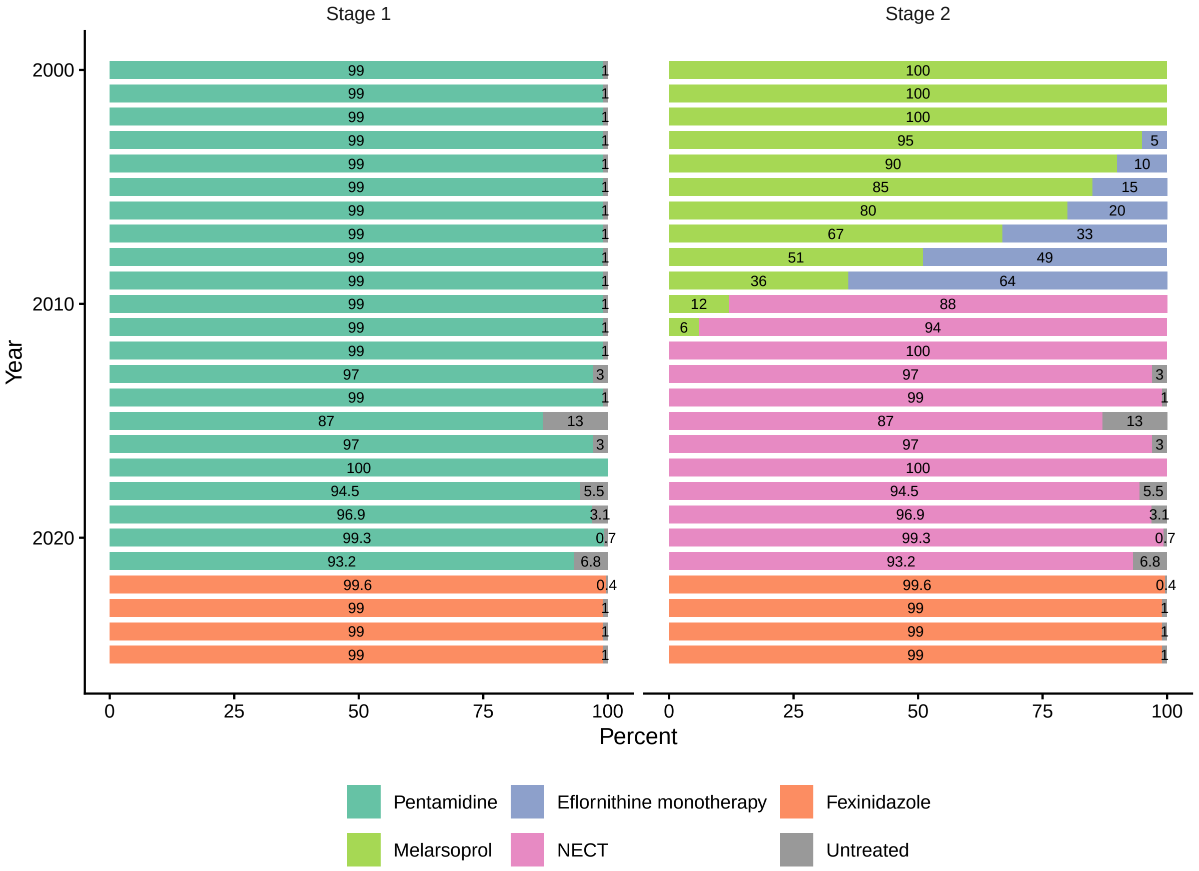

Figure S10: Treatments 2000-25 in Uganda. Abbreviations: NECT: nifurtimox-eflornithine combination therapy.

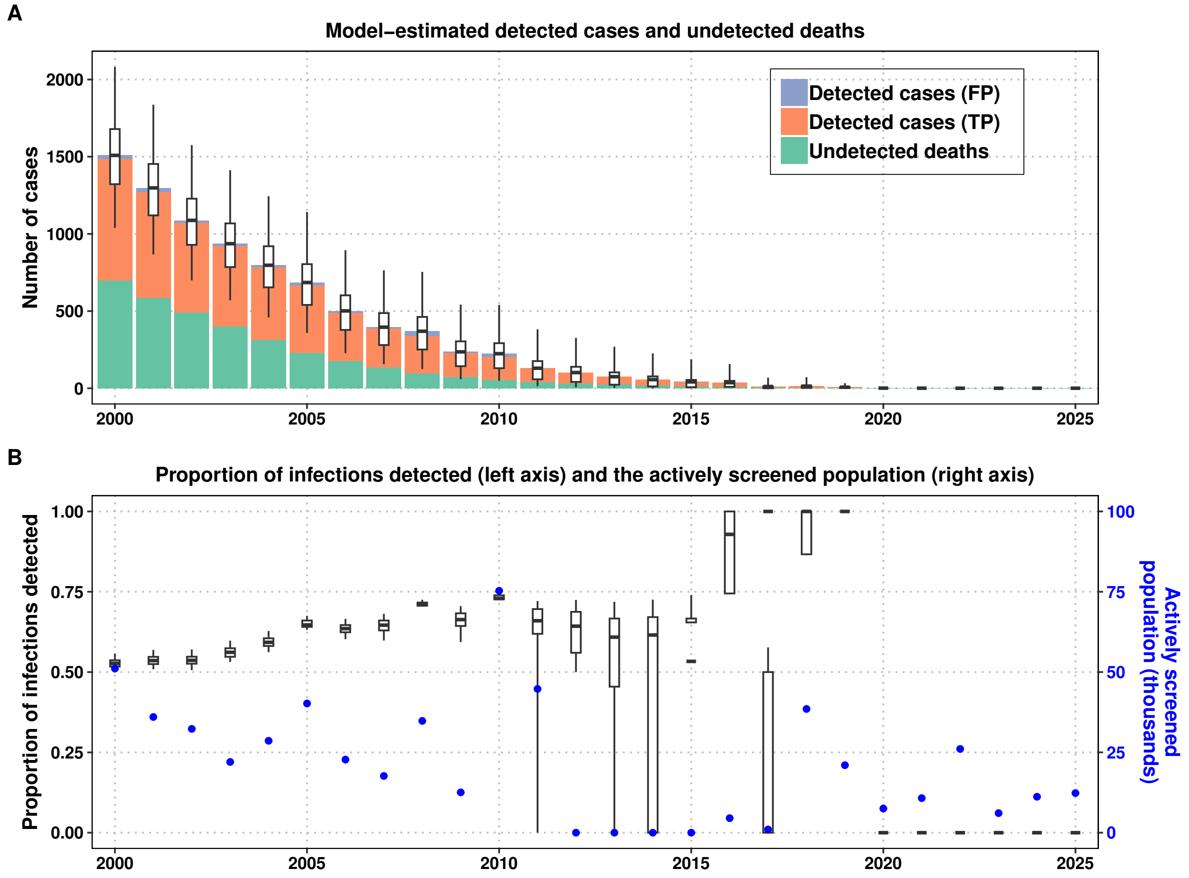

Figure S11: Modelled cases, undetected deaths, and false positives 2000–25 in Uganda's northeast districts endemic for gHAT.

#### S10.4 Cost functions

##### S10.4.1 Passive screening

For each district, the costs for passive screening for year $Y$ and district $D$ are computed as follows, using the parameters from Table S9:

$$\begin{aligned}
\text{Cost}_{\text{PS}}(Y,D) &= \text{RDT costs} + \text{confirmation costs} \\
\text{where} & \\
\text{RDT costs} &= \Big[\Big(\text{Number of RDT-only test centers in district } D \text{ in year } Y \\
&\quad \times \text{PS: capital \& management of an RDT-testing facility}\Big) \\
&+ \Big(\text{Number of RDTs done per district in year } Y \\
&\quad \times (\text{RDT cost} \times (1 + \text{RDT wastage factor during PS}))\Big)\Big] \qquad \text{(S10.1)} \\
&\times (1 + \text{National management cost, ISSEP costs}) \\
\text{Confirmation costs} &= \Big[\Big(\text{Number of confirmation test centers in district } D \text{ in year } Y \\
&\quad \times \text{PS: capital \& management of a confirmation facilities}\Big) \\
&+ \Big(\text{PS: commmunication \& awareness}\Big)\Big] \\
&\times (1 + \text{National management cost, ISSEP costs})
\end{aligned}$$

While cost data was based on few observations of the resource use for 2023 and the budgets from 2019-2023, we characterized uncertainty by assuming that cost data would fit a gamma distribution and parameterized the distribution as if the 95% confidence interval ranged from half to double the observed resource cost – our usual practice (see the Principles for Parameterization for the Parameter Glossary in section **??**). Capital and management costs consist of medical equipment, training, and management. Unlike in other analyses, we did not take into account the consult cost, as the RDT would not take substantial amounts of time and the patients are coming to their consults to address symptoms that they have, even if not caused by gHAT.

The annual costs are summarised in the following subtotals and totals for the districts, as detailed in Table S13 and district totals and the national total is shown in Table S14.

Table S13: Summary of resources, unit costs, and costs by strategy, district, and item

| Item | Resources | Unit Costs | Costs |
|---|---|---|---|
| **Adjumani RDT** | | | |
| Facility | 6.00 | 341 (221-485) | 2,046 (1,324-2,908) |
| Tests | 62.87 (62.27-64.52) | 2.51 (2.37-2.66) | 158 (149-168) |
| Program Markup | | 10% | 220 (148-306) |
| **Subtotal** | | | **2,424 (1,630-3,371)** |
| **Adjumani Confirmation** | | | |
| Facility | 2.00 | 1168 (981-1369) | 2,335 (1,961-2,738) |
| Communications | 1.00 | 283 (127-504) | 283 (127-504) |
| Program Markup | | 10% | 262 (220-307) |
| **Subtotal** | | | **2,880 (2,423-3,375)** |
| **Amuru RDT** | | | |
| Facility | 1.00 | 341 (221-485) | 341 (221-485) |
| Tests | 20.70 (20.51-21.25) | 2.51 (2.37-2.66) | 52.03 (48.95-55.26) |
| Program Markup | | 10% | 39.30 (27.27-53.65) |
| **Subtotal** | | | **432 (300-590)** |
| **Amuru Confirmation** | | | |
| Facility | 1.00 | 1168 (981-1369) | 1168 (981-1369) |
| Communications | 1.00 | 283 (127-504) | 283 (127-504) |
| Program Markup | | 10% | 145 (120-174) |
| **Subtotal** | | | **1,596 (1,317-1,914)** |
| **Arua RDT** | | | |
| Facility | 10.00 | 341 (221-485) | 3,410 (2,207-4,847) |

Table S13: Summary of resources, unit costs, and costs by strategy, district, and item *(continued)*

| Item | Resources | Unit Costs | Costs |
|---|---|---|---|
| Tests | 139 (138-143) | 2.51 (2.37-2.66) | 349 (328-371) |
| Program Markup | | 10% | 376 (256-519) |
| **Subtotal** | | | **4,135 (2,811-5,714)** |
| **Arua Confirmation** | | | |
| Facility | 2.00 | 1168 (981-1369) | 2,335 (1,961-2,738) |
| Communications | 1.00 | 283 (127-504) | 283 (127-504) |
| Program Markup | | 10% | 262 (220-307) |
| **Subtotal** | | | **2,880 (2,423-3,375)** |
| **Koboko RDT** | | | |
| Facility | 6.00 | 341 (221-485) | 2,046 (1,324-2,908) |
| Tests | 317 (314-325) | 2.51 (2.37-2.66) | 796 (749-846) |
| Program Markup | | 10% | 284 (212-371) |
| **Subtotal** | | | **3,126 (2,330-4,080)** |
| **Koboko Confirmation** | | | |
| Facility | 1.00 | 1168 (981-1369) | 1168 (981-1369) |
| Communications | 1.00 | 283 (127-504) | 283 (127-504) |
| Program Markup | | 10% | 145 (120-174) |
| **Subtotal** | | | **1,596 (1,317-1,914)** |
| **Maracha RDT** | | | |
| Facility | 3.00 | 341 (221-485) | 1023 (662-1454) |
| Tests | 19.69 (19.50-20.21) | 2.51 (2.37-2.66) | 49.49 (46.56-52.56) |
| Program Markup | | 10% | 107 (71-150) |
| **Subtotal** | | | **1180 (783-1653)** |
| **Maracha Confirmation** | | | |
| Facility | 1.00 | 1168 (981-1369) | 1168 (981-1369) |
| Communications | 1.00 | 283 (127-504) | 283 (127-504) |
| Program Markup | | 10% | 145 (120-174) |
| **Subtotal** | | | **1,596 (1,317-1,914)** |
| **Moyo RDT** | | | |
| Facility | 4.00 | 341 (221-485) | 1364 (883-1939) |
| Tests | 49.49 (49.01-50.78) | 2.51 (2.37-2.66) | 124 (117-132) |
| Program Markup | | 10% | 149 (101-206) |
| **Subtotal** | | | **1,637 (1,108-2,269)** |
| **Moyo Confirmation** | | | |
| Facility | 4.00 | 1168 (981-1369) | 4,671 (3,922-5,477) |
| Communications | 1.00 | 283 (127-504) | 283 (127-504) |
| Program Markup | | 10% | 495 (418-579) |
| **Subtotal** | | | **5,449 (4,597-6,366)** |
| **Yumbe RDT** | | | |
| Facility | 9.00 | 341 (221-485) | 3,069 (1,986-4,363) |
| Tests | 214 (212-220) | 2.51 (2.37-2.66) | 538 (506-571) |
| Program Markup | | 10% | 361 (252-490) |
| **Subtotal** | | | **3,967 (2,776-5,386)** |
| **Yumbe Confirmation** | | | |
| Facility | 2.00 | 1168 (981-1369) | 2,335 (1,961-2,738) |
| Communications | 1.00 | 283 (127-504) | 283 (127-504) |
| Program Markup | | 10% | 262 (220-307) |
| **Subtotal** | | | **2,880 (2,423-3,375)** |

Table S14: Estimated annual costs (USD) by district

| District | Cost |
|---|---|
| Adjumani | 5,304 (4,378-6,355) |
| Amuru | 2,028 (1,715-2,382) |
| Arua | 7,015 (5,605-8,653) |
| Koboko | 4,722 (3,871-5,706) |
| Maracha | 2,775 (2,276-3,336) |
| Moyo | 7,086 (6,064-8,176) |
| Yumbe | 6,847 (5,560-8,334) |
| **Total** | **35,778 (29,468-42,940)** |

##### S10.4.2 Active screening

The transmission model outputs the number of active screening activities and the number of people screened in each year. The function below is the cost per district:

$$\begin{aligned}\text{Cost}_{\text{AS}}(Y,D) \quad = \quad & \text{Number of screening tests performed in year } Y \\ & \times\Big(\text{AS capital \& management costs} \\ & +\text{CATT costs} \times (1+\text{Wastage for CATT for AS}) \times (1+\text{Delivery mark up}) \\ & +\text{AS: pre-mission communications and awareness}\Big) \\ & \times(1+\text{National management cost, ISSEP costs})\end{aligned} \quad (S10.2)$$

While cost data was based on the budgets from 2019-2023, we characterised uncertainty according to our Principles for Parameterisation, as listed in Supplement 2 (the Parameter Glossary) in section **??**.

The annual costs are summarised in the following subtotals and totals for the districts, as detailed in Table S15. The **Teams** costs are calculated according to the number of individuals screened and the capital & management costs (fuel, equipment, training, and management), the **Tests** costs are per person screened, the **Communications** the costs are also on a per-person basis, and the **Program Markup** are a markup of all the other costs.

Table S15: Summary of resources, unit costs, and costs by strategy, district, and item for an 8,000-person active screening event

| Item | Resources | Unit Costs | Costs |
|---|---|---|---|
| **Adjumani Reduced AS** | | | |
| Teams | 786 | 1.00 (0.88-1.13) | 787 (690-889) |
| Tests | 849 (814-897) | 0.79 (0.74-0.83) | 667 (620-722) |
| Communications | 786 | 0.36 (0.16-0.63) | 280 (126-497) |
| Program Markup | | 10% | 173 (154-198) |
| **Subtotal** | | | **1,908 (1,695-2,177)** |
| **Adjumani Mean AS** | | | |
| Teams | 1,572 | 1.00 (0.88-1.13) | 1,574 (1,380-1,778) |
| Tests | 1,698 (1,627-1,793) | 0.79 (0.74-0.83) | 1,335 (1,239-1,443) |
| Communications | 1,572 | 0.36 (0.16-0.63) | 560 (251-993) |
| Program Markup | | 10% | 347 (308-396) |
| **Subtotal** | | | **3,816 (3,389-4,354)** |
| **Adjumani Max AS** | | | |
| Teams | 5,751 | 1.00 (0.88-1.13) | 5,758 (5,050-6,504) |
| Tests | 6,211 (5,954-6,560) | 0.79 (0.74-0.83) | 4,883 (4,533-5,279) |
| Communications | 5,751 | 0.36 (0.16-0.63) | 2049 (919-3635) |
| Program Markup | | 10% | 1,269 (1,127-1,448) |
| **Subtotal** | | | **13,959 (12,399-15,929)** |
| **Amuru Max AS** | | | |
| Teams | 1,305 | 1.00 (0.88-1.13) | 1,307 (1,146-1,476) |
| Tests | 1,409 (1,351-1,488) | 0.79 (0.74-0.83) | 1,108 (1,029-1,198) |
| Communications | 1,305 | 0.36 (0.16-0.63) | 465 (209-825) |
| Program Markup | | 10% | 288 (256-329) |
| **Subtotal** | | | **3,168 (2,814-3,615)** |
| **Arua Reduced AS** | | | |
| Teams | 1,162 | 1.00 (0.88-1.13) | 1,163 (1,020-1,314) |
| Tests | 1,255 (1,203-1,325) | 0.79 (0.74-0.83) | 987 (916-1067) |
| Communications | 1,162 | 0.36 (0.16-0.63) | 414 (186-734) |
| Program Markup | | 10% | 256 (228-293) |
| **Subtotal** | | | **2,820 (2,505-3,218)** |
| **Arua Mean AS** | | | |
| Teams | 2,323 | 1.00 (0.88-1.13) | 2,326 (2,040-2,627) |
| Tests | 2,509 (2,405-2,650) | 0.79 (0.74-0.83) | 1,973 (1,831-2,132) |

Table S15: Summary of resources, unit costs, and costs by strategy, district, and item for an 8,000-person active screening event *(continued)*

| Item | Resources | Unit Costs | Costs |
|---|---|---|---|
| Communications | 2,323 | 0.36 (0.16-0.63) | 828 (371-1468) |
| Program Markup | | 10% | 513 (455-585) |
| **Subtotal** | | | **5,639 (5,008-6,434)** |
| **Arua Max AS** | | | |
| Teams | 26,118 | 1.00 (0.88-1.13) | 26,150 (22,936-29,539) |
| Tests | 28,205 (27,039-29,790) | 0.79 (0.74-0.83) | 22,178 (20,586-23,975) |
| Communications | 26,118 | 0.36 (0.16-0.63) | 9,305 (4,175-16,506) |
| Program Markup | | 10% | 5,763 (5,119-6,576) |
| **Subtotal** | | | **63,395 (56,311-72,341)** |
| **Koboko Reduced AS** | | | |
| Teams | 1,814 | 1.00 (0.88-1.13) | 1,816 (1,593-2,052) |
| Tests | 1,959 (1,878-2,069) | 0.79 (0.74-0.83) | 1,540 (1,430-1,665) |
| Communications | 1,814 | 0.36 (0.16-0.63) | 646 (290-1146) |
| Program Markup | | 10% | 400 (356-457) |
| **Subtotal** | | | **4,403 (3,911-5,024)** |
| **Koboko Mean AS** | | | |
| Teams | 3,628 | 1.00 (0.88-1.13) | 3,632 (3,186-4,103) |
| Tests | 3,918 (3,756-4,138) | 0.79 (0.74-0.83) | 3,081 (2,860-3,330) |
| Communications | 3,628 | 0.36 (0.16-0.63) | 1292 (580-2293) |
| Program Markup | | 10% | 801 (711-914) |
| **Subtotal** | | | **8,806 (7,822-10,049)** |
| **Koboko Max AS** | | | |
| Teams | 30,442 | 1.00 (0.88-1.13) | 30,479 (26,733-34,430) |
| Tests | 32,874 (31,516-34,722) | 0.79 (0.74-0.83) | 25,849 (23,994-27,944) |
| Communications | 30,442 | 0.36 (0.16-0.63) | 10,845 (4,866-19,239) |
| Program Markup | | 10% | 6,717 (5,967-7,665) |
| **Subtotal** | | | **73,891 (65,633-84,317)** |
| **Maracha Reduced AS** | | | |
| Teams | 300 | 1.00 (0.88-1.13) | 300 (263-339) |
| Tests | 324 (311-342) | 0.79 (0.74-0.83) | 255 (236-275) |
| Communications | 300 | 0.36 (0.16-0.63) | 107 (48-190) |
| Program Markup | | 10% | 66.20 (58.80-75.54) |
| **Subtotal** | | | **728 (647-831)** |
| **Maracha Mean AS** | | | |
| Teams | 601 | 1.00 (0.88-1.13) | 602 (528-680) |
| Tests | 649 (622-686) | 0.79 (0.74-0.83) | 510 (474-552) |
| Communications | 601 | 0.36 (0.16-0.63) | 214 (96-380) |
| Program Markup | | 10% | 133 (118-151) |
| **Subtotal** | | | **1,459 (1,296-1,665)** |
| **Maracha Max AS** | | | |
| Teams | 5,680 | 1.00 (0.88-1.13) | 5,687 (4,988-6,424) |
| Tests | 6,134 (5,880-6,479) | 0.79 (0.74-0.83) | 4,823 (4,477-5,214) |
| Communications | 5,680 | 0.36 (0.16-0.63) | 2024 (908-3590) |
| Program Markup | | 10% | 1,253 (1,113-1,430) |
| **Subtotal** | | | **13,787 (12,246-15,732)** |
| **Moyo Reduced AS** | | | |
| Teams | 1,777 | 1.00 (0.88-1.13) | 1,779 (1,560-2,010) |
| Tests | 1,919 (1,840-2,027) | 0.79 (0.74-0.83) | 1,509 (1,401-1,631) |
| Communications | 1,777 | 0.36 (0.16-0.63) | 633 (284-1123) |
| Program Markup | | 10% | 392 (348-447) |
| **Subtotal** | | | **4,313 (3,831-4,922)** |
| **Moyo Mean AS** | | | |

Table S15: Summary of resources, unit costs, and costs by strategy, district, and item for an 8,000-person active screening event *(continued)*

| Item | Resources | Unit Costs | Costs |
|---|---|---|---|
| Teams | 3,555 | 1.00 (0.88-1.13) | 3,559 (3,122-4,021) |
| Tests | 3,839 (3,680-4,055) | 0.79 (0.74-0.83) | 3,019 (2,802-3,263) |
| Communications | 3,555 | 0.36 (0.16-0.63) | 1266 (568-2247) |
| Program Markup | | 10% | 784 (697-895) |
| **Subtotal** | | | **8,629 (7,665-9,846)** |
| **Moyo Max AS** | | | |
| Teams | 34,755 | 1.00 (0.88-1.13) | 34,797 (30,520-39,307) |
| Tests | 37,532 (35,981-39,642) | 0.79 (0.74-0.83) | 29,512 (27,394-31,904) |
| Communications | 34,755 | 0.36 (0.16-0.63) | 12,382 (5,555-21,965) |
| Program Markup | | 10% | 7,669 (6,812-8,751) |
| **Subtotal** | | | **84,360 (74,932-96,263)** |
| **Yumbe Reduced AS** | | | |
| Teams | 327 | 1.00 (0.88-1.13) | 327 (287-370) |
| Tests | 353 (339-373) | 0.79 (0.74-0.83) | 278 (258-300) |
| Communications | 327 | 0.36 (0.16-0.63) | 116 (52-207) |
| Program Markup | | 10% | 72.16 (64.09-82.34) |
| **Subtotal** | | | **794 (705-906)** |
| **Yumbe Mean AS** | | | |
| Teams | 652 | 1.00 (0.88-1.13) | 653 (573-737) |
| Tests | 704 (675-744) | 0.79 (0.74-0.83) | 554 (514-599) |
| Communications | 652 | 0.36 (0.16-0.63) | 232 (104-412) |
| Program Markup | | 10% | 144 (128-164) |
| **Subtotal** | | | **1,583 (1,406-1,806)** |
| **Yumbe Max AS** | | | |
| Teams | 22,189 | 1.00 (0.88-1.13) | 22,216 (19,485-25,095) |
| Tests | 23,962 (22,971-25,309) | 0.79 (0.74-0.83) | 18,841 (17,489-20,368) |
| Communications | 22,189 | 0.36 (0.16-0.63) | 7,905 (3,547-14,023) |
| Program Markup | | 10% | 4,896 (4,349-5,587) |
| **Subtotal** | | | **53,859 (47,840-61,458)** |

##### S10.4.3 Reactive screening

The transmission model outputs the number of one-off reactive screening (RS) activities and the number of people screened annually. The function below is the cost per district:

$$\begin{aligned}\text{Cost}_{\text{RS}}(Y,D) \quad = \quad & \text{Number of screening tests performed in year } Y \\ & \times\Big(\text{RS capital \& management costs} \\ & +\text{CATT costs} \times (1+\text{Wastage for CATT for AS}) \times (1+\text{Delivery mark up}) \\ & +\text{RS: pre-mission communications and awareness}\Big) \\ & \times(1+\text{National management cost, ISSEP costs})\end{aligned} \tag{S10.3}$$

While cost data was based on the budgets from 2019-2023, we characterised uncertainty according to our Principles for Parameterisation, as listed in Supplement 2 (the Parameter Glossary) in section **??**.

The annual costs are summarised in the following subtotals and totals for the districts, as detailed in Table S16. The **Teams** costs are calculated according to the number of individuals screened and the capital & management costs (fuel, equipment, training, and management), the **Tests** costs are per person screened, the **Communications** the costs are also on a per-person basis, and the **Program Markup** are a markup of all the other costs.

Table S16: Summary of resources, unit costs, and costs by strategy, district, and item for a 4,000-person reactive screening event.

| Item | Resources | Unit Costs | Costs |
|---|---|---|---|
| Teams | 4,000 | 1.67 (1.44-1.92) | 6,693 (5,756-7,687) |
| Tests | 4,320 (4,141-4,562) | 0.79 (0.74-0.83) | 3,397 (3,153-3,672) |
| Communications | 4,000 | 0.42 (0.19-0.75) | 1694 (750-3017) |
| Program Markup | | 10% | 1,178 (1,039-1,345) |
| **Subtotal** | | | **12,962 (11,426-14,792)** |

##### S10.4.4 Vector control and reactive vector control

For alternative strategies of VC, we assume that the extent of VC and the density of targets are as detailed in the district-specific parameter table (Table S10). We split the cost function into two elements: one is the management cost per square kilometre, and the other is the supply and labour cost of each vector target deployed. Costs are detailed in the parameter glossary, sections **??**, **??**, **??**, **??**, **??**.

$$\begin{aligned}\text{Cost}_{\text{VC}}(Y) \quad = \quad & X \text{ km}^2 \text{ per RVC deployment} \\ & \times\Big(\text{Cost of VC management per km}^2 + \text{Targets per km}^2 \text{ in that district} \times 2 \text{ deployments per year}\Big)\end{aligned} \tag{S10.4}$$

For deployments of reactive VC, the transmission model outputs the number of "units" of reactive vector control (RVC) each year. We assume that a single RVC unit would 50 km$^2$ in size to cover and surround the village where the detected case was living; this is believed to be the smallest operational area to have an impact on tsetse populations [S84]. We also assume that for RVC, two deployments would happen in line with the previous VC deployments, which occurred twice yearly in Uganda. The function below is the cost per district, assuming 8.5 targets per km$^2$.

$$\begin{aligned}\text{Cost}_{\text{RVC}}(Y) \quad = \quad & \text{Number of units of RVC in year } Y \times 50\text{km}^2 \text{ per RVC deployment} \\ & \times\Big(\text{ Cost of VC management per km}^2 + \text{Targets per km}^2 \times 2 \text{ deployments per year}\Big)\end{aligned} \tag{S10.5}$$

While cost data was based on the budgets from 2019-2023, we characterised uncertainty according to our Principles for Parameterisation, as listed in Supplement 2 (the Parameter Glossary) in section **??**.

The annual costs are summarised in the following subtotals and totals for the districts, as detailed in Table S17.

Table S17: Summary of resources, unit costs, and costs by strategy, district, and item

| Item | Resources | Unit Costs | Costs |
|---|---|---|---|
| **Adjumani** | | | |
| VC management | 608 sq-km | 44.35 (19.44-78.92) | 26,967 (11,821-47,982) |
| Targets | 2,590 | 5.42 (2.44-9.68) | 14,043 (6,314-25,067) |
| **Subtotal** | | | **41,009 (23,313-63,841)** |
| **Amuru** | | | |
| VC management | 200 sq-km | 44.35 (19.44-78.92) | 8,871 (3,889-15,784) |
| Targets | 1,374 | 5.42 (2.44-9.68) | 7,449 (3,350-13,298) |
| **Subtotal** | | | **16,320 (9,476-25,020)** |
| **Arua** | | | |
| VC management | 631 sq-km | 44.35 (19.44-78.92) | 27,987 (12,268-49,797) |
| Targets | 4,770 | 5.42 (2.44-9.68) | 25,864 (11,630-46,168) |
| **Subtotal** | | | **53,851 (31,291-82,302)** |
| **Koboko** | | | |
| VC management | 285 sq-km | 44.35 (19.44-78.92) | 12,641 (5,541-22,492) |
| Targets | 2,038 | 5.42 (2.44-9.68) | 11,048 (4,968-19,721) |
| **Subtotal** | | | **23,689 (13,751-36,278)** |
| **Maracha** | | | |
| VC management | 151 sq-km | 44.35 (19.44-78.92) | 6,697 (2,936-11,917) |
| Targets | 1,666 | 5.42 (2.44-9.68) | 9,030 (4,060-16,119) |
| **Subtotal** | | | **15,727 (9,102-24,165)** |
| **Moyo** | | | |
| VC management | 554 sq-km | 44.35 (19.44-78.92) | 24,572 (10,771-43,721) |
| Targets | 4,360 | 5.42 (2.44-9.68) | 23,639 (10,629-42,196) |
| **Subtotal** | | | **48,210 (28,016-73,621)** |
| **Yumbe** | | | |
| VC management | 410 sq-km | 44.35 (19.44-78.92) | 18,185 (7,971-32,356) |
| Targets | 4,621 | 5.42 (2.44-9.68) | 25,052 (11,265-44,719) |
| **Subtotal** | | | **43,237 (24,995-66,483)** |

The annual costs are summarised in the following subtotals and totals for the districts, as detailed in Table S18.

Table S18: Summary of resources, unit costs, and costs by strategy, district, and item for a reactive vector control deployment of 50 sq-km.

| Item | Resources | Unit Costs | Costs |
|---|---|---|---|
| VC management | 50.00 sq-km | 44.35 (19.44-78.92) | 2218 (972-3946) |
| Targets | 425 | 5.42 (2.44-9.68) | 2,304 (1,036-4,113) |
| **Subtotal** | | | **4,522 (2,626-6,910)** |

##### S10.4.5 Treatment

We show costs here conditional on the treatment. Treatment eligibility estimates and distributions are described in Table S11.

$$\begin{aligned}\text{Cost}_{\text{TxFexiOP}}(Y,D) \;=\;& \text{Number of cases in year } Y \text{ eligible for Fexinidazole outpatient treatment}\\ &\times\Big(\text{Fexinidazole Rx cost} \times (1+\text{Delivery mark-up})\\ &\quad +10\times\text{Outpatient consultation: cost}\\ &\quad +\text{Pr. SAE}_{\text{Fexi}}\times\text{Cost treat SAE}\Big)\end{aligned} \tag{S10.6}$$

$$\begin{aligned}\text{Cost}_{\text{TxFexiIP}}(Y,D) \;=\;& \Big(\text{Number of cases in year } Y \text{ eligible for Fexinidazole inpatient treatment}\\ &\times\text{Fexinidazole Rx cost} \times (1+\text{Delivery mark-up})\\ &\quad +\text{Outpatient consultation: cost}\\ &\quad +\text{Hospital stay: cost per day}\\ &\quad +\text{Pr. SAE}_{\text{Fexi}}\times\text{Cost treat SAE}\Big)\end{aligned} \tag{S10.7}$$

$$\begin{aligned}\text{Cost}_{\text{TxNECT}}(Y,D) \;=\;& \text{Number of confirmed cases in year } Y \text{ ineligible for Fexi treatment}\\ &\times\Big(\text{NECT Rx Cost} \times (1+\text{Delivery mark-up})\\ &\quad +\text{Outpatient consultation: cost}\\ &\quad +\text{Hospital stay: cost per day}\\ &\quad +\text{Pr. SAE}_{\text{NECT}}\times\text{Cost treat SAE}\Big)\end{aligned} \tag{S10.8}$$

$$\begin{aligned}\text{Cost}_{\text{TxPenta}}(Y,D) \;=\;& \text{Number of confirmed cases in year } Y \text{ ineligible for Fexi treatment}\\ &\times\Big(\text{Course of pentamidine: cost}(1+\text{Delivery mark-up})\\ &\quad +7\times\text{Outpatient consultation: cost}\\ &\quad +\text{Pr. SAE}_{\text{Penta}}\times\text{Cost treat SAE}\Big)\end{aligned} \tag{S10.9}$$

Table S19: Cost per person for different gHAT treatments. Because these are costs averaged over all patients and SAEs are rare, the average cost per patient for SAE is low.

| | Pentamidine | NECT | Fexinidazole - inpatient | Fexinidazole - outpatient |
|---|---|---|---|---|
| Doctor's consult | 7.38 (1.20-18.93) | 1.05 (0.17-2.70) | 1.05 (0.17-2.70) | 10.55 (1.72-27.04) |
| Inpatient care | 0 | 45.96 (15.88-92.43) | 45.96 (15.88-92.43) | 0 |
| Medicine & delivery | 78.44 (63.39-95.35) | 521.65 (416.51-637.64) | 318.60 (255.42-389.16) | 318.60 (255.42-389.16) |
| Treatment for SAE | <0.01 (<0.01-0.01) | 1.46 (0.14-5.52) | 0.17 (<0.01-0.75) | 0.17 (<0.01-0.75) |
| Total | 92.02 (73.05-113.91) | 576.32 (465.10-702.75) | 365.78 (292.03-447.91) | 329.16 (265.16-402.01) |

### S11 Additional results from the epidemiological model

#### S11.1 Model comparison

Figure S12 shows the relative model evidence for each model variant within district – that is, model evidence values scaled such that their within-district sum across the eight models is equal to one. The model variant with the highest evidence value is the favoured variant. The relative model evidence values are the weights applied when creating the ensemble model joint posterior.

#### S11.2 Parameter estimation

Figures S13–S19 show the histogram of each fitted parameter for seven districts.

#### S11.3 Fitting

Figures S20–S26 show the fitting results for seven districts.

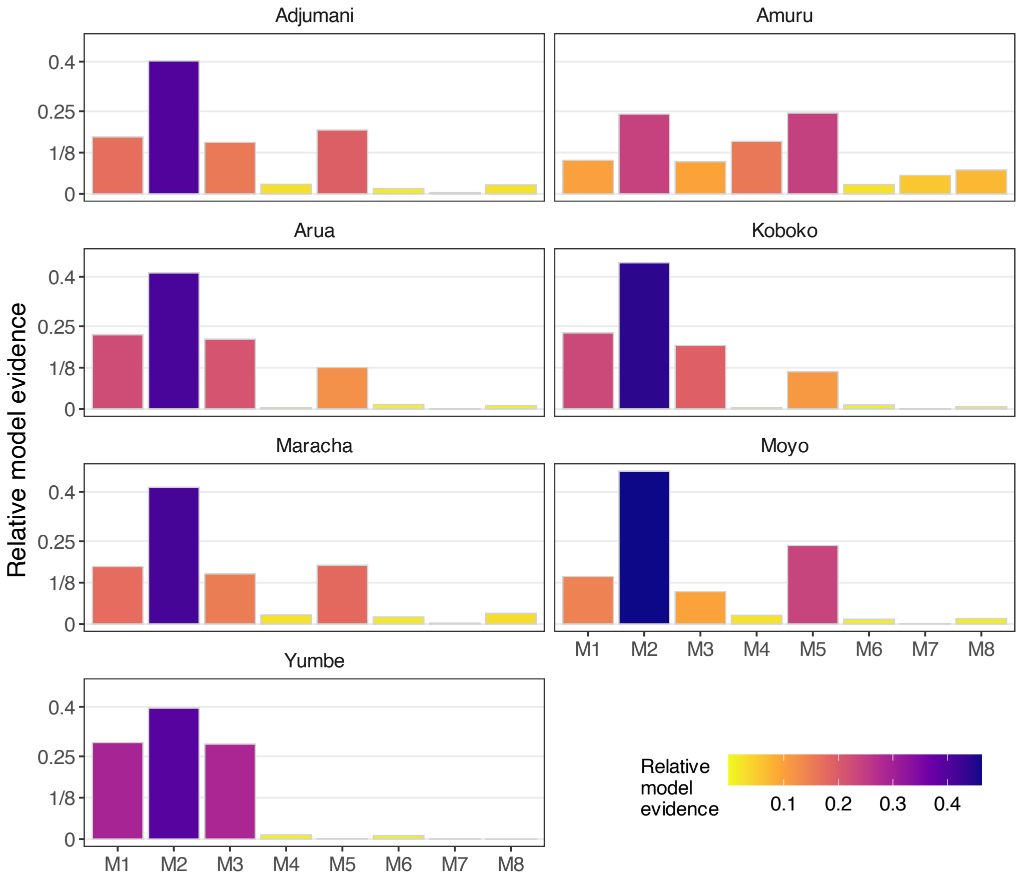

Figure S12: **Relative model evidence.** Evidence (marginal likelihood) for each model, scaled so that the sum across model variants within a district is one. These values are the weights used to create the ensemble model posteriors.

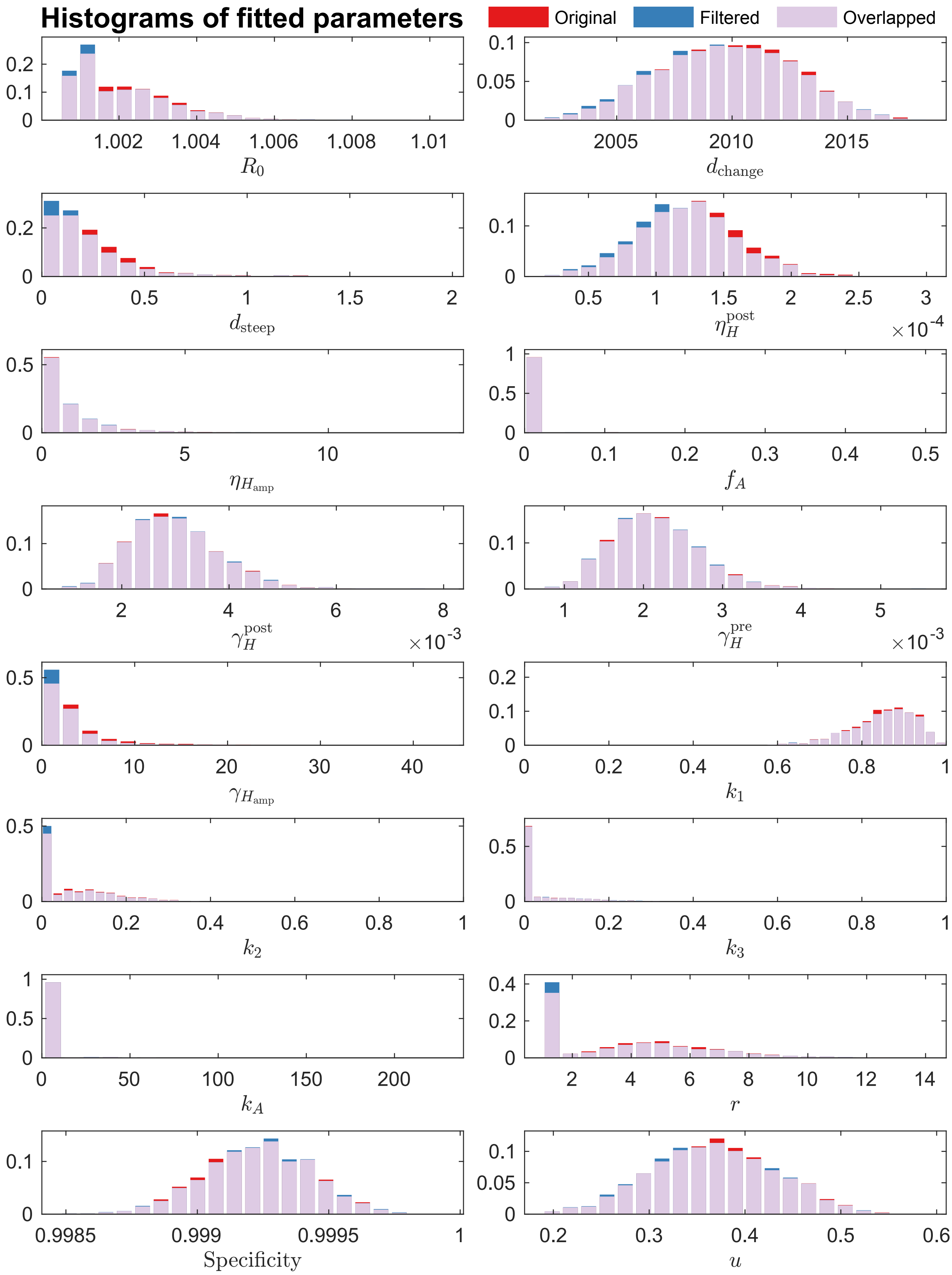

Figure S13: **Histograms of model fitted parameters for Adjumani.** The histograms are normalised to illustrate the probability density of fitted parameters. Two sets of histograms are presented in each figure to highlight the changes in parameter distributions arising from discarding realisations that do not match the observed presence/absence of reported cases in 2020–2024. The original (non-filtered) ensemble posterior results are shown in red, and the filtered results are shown in blue. Their overlaps are coloured in light purple. Of the 16,856 realisations that survived filtering, we randomly select 20,000 samples to form the new ensemble posterior for simulations. See Table S4 for the descriptions of fitted parameters.

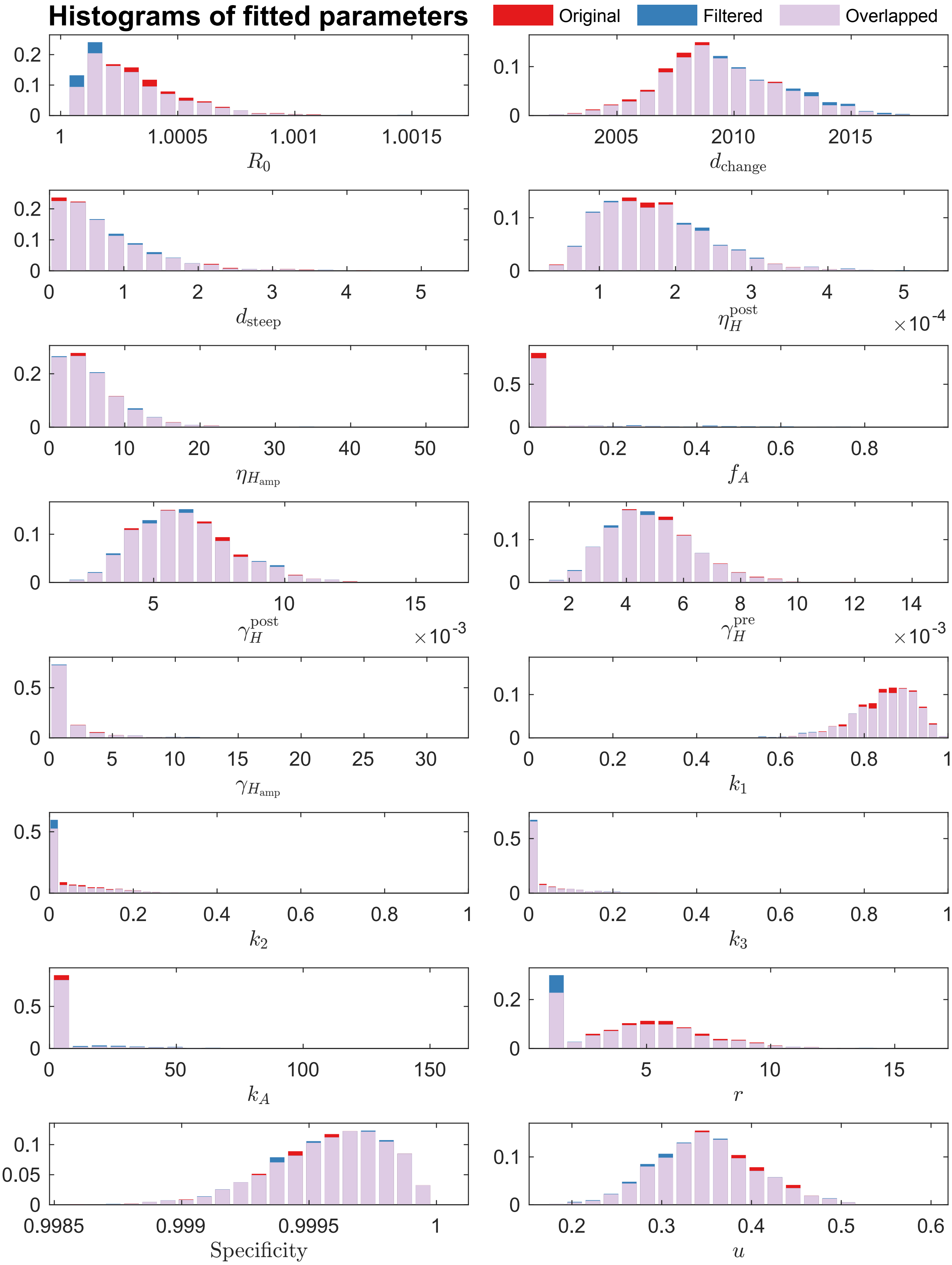

Figure S14: **Histograms of model fitted parameters for Amuru.** The histograms are normalised to illustrate the probability density of fitted parameters. Two sets of histograms are presented in each figure to highlight the changes in parameter distributions arising from discarding realisations that do not match the observed presence/absence of reported cases in 2020–2024. The original (non-filtered) ensemble posterior results are shown in red, and the filtered results are shown in blue. Their overlaps are coloured in light purple. Of the 19,493 realisations that survived filtering, we randomly select 20,000 samples to form the new ensemble posterior for simulations. See Table S4 for the descriptions of fitted parameters.

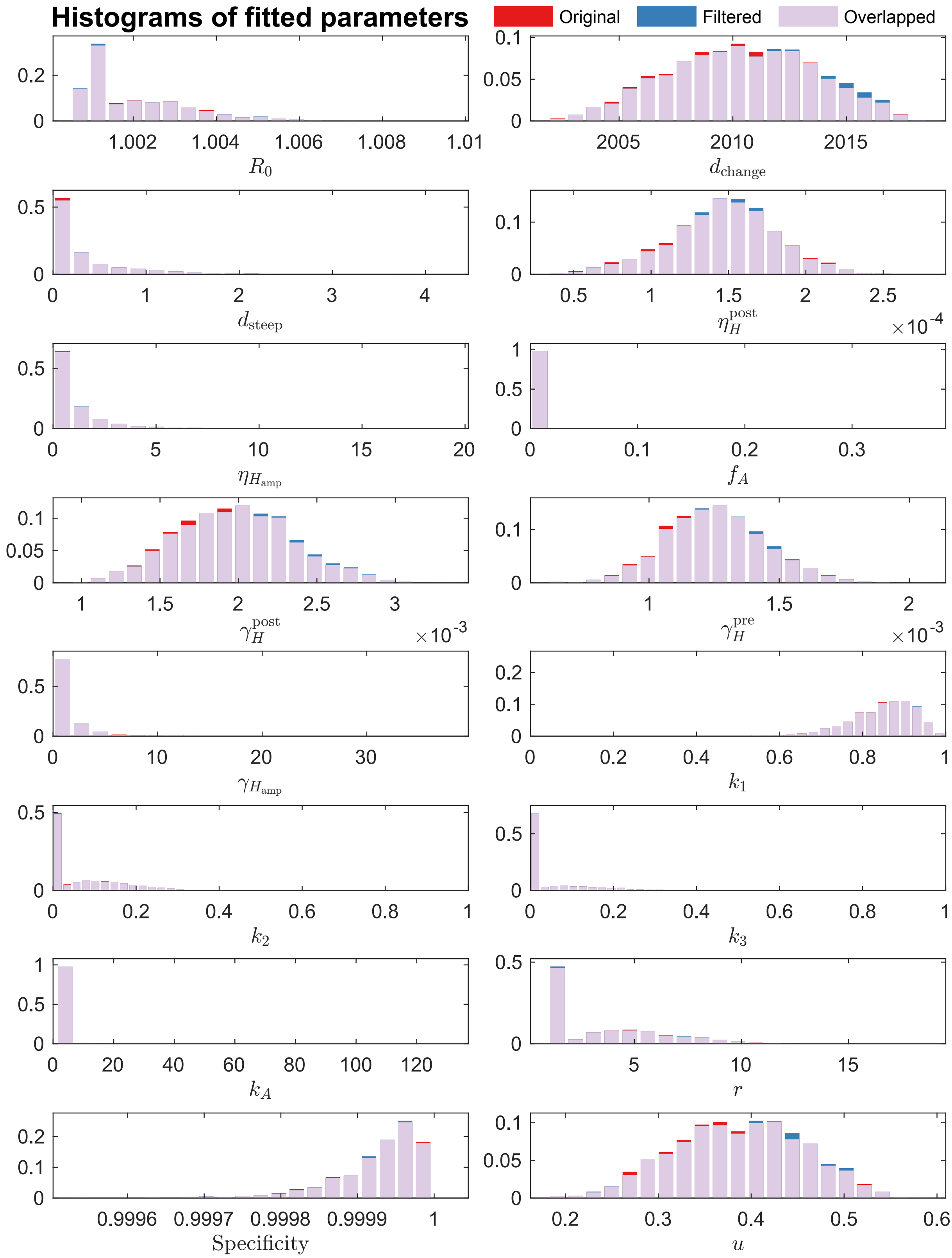

Figure S15: **Histograms of model fitted parameters for Arua.** The histograms are normalised to illustrate the probability density of fitted parameters. Two sets of histograms are presented in each figure to highlight the changes in parameter distributions arising from discarding realisations that do not match the observed presence/absence of reported cases in 2020–2024. The original (non-filtered) ensemble posterior results are shown in red, and the filtered results are shown in blue. Their overlaps are coloured in light purple. Of the 17,769 realisations that survived filtering, we randomly select 20,000 samples to form the new ensemble posterior for simulations. See Table S4 for the descriptions of fitted parameters.

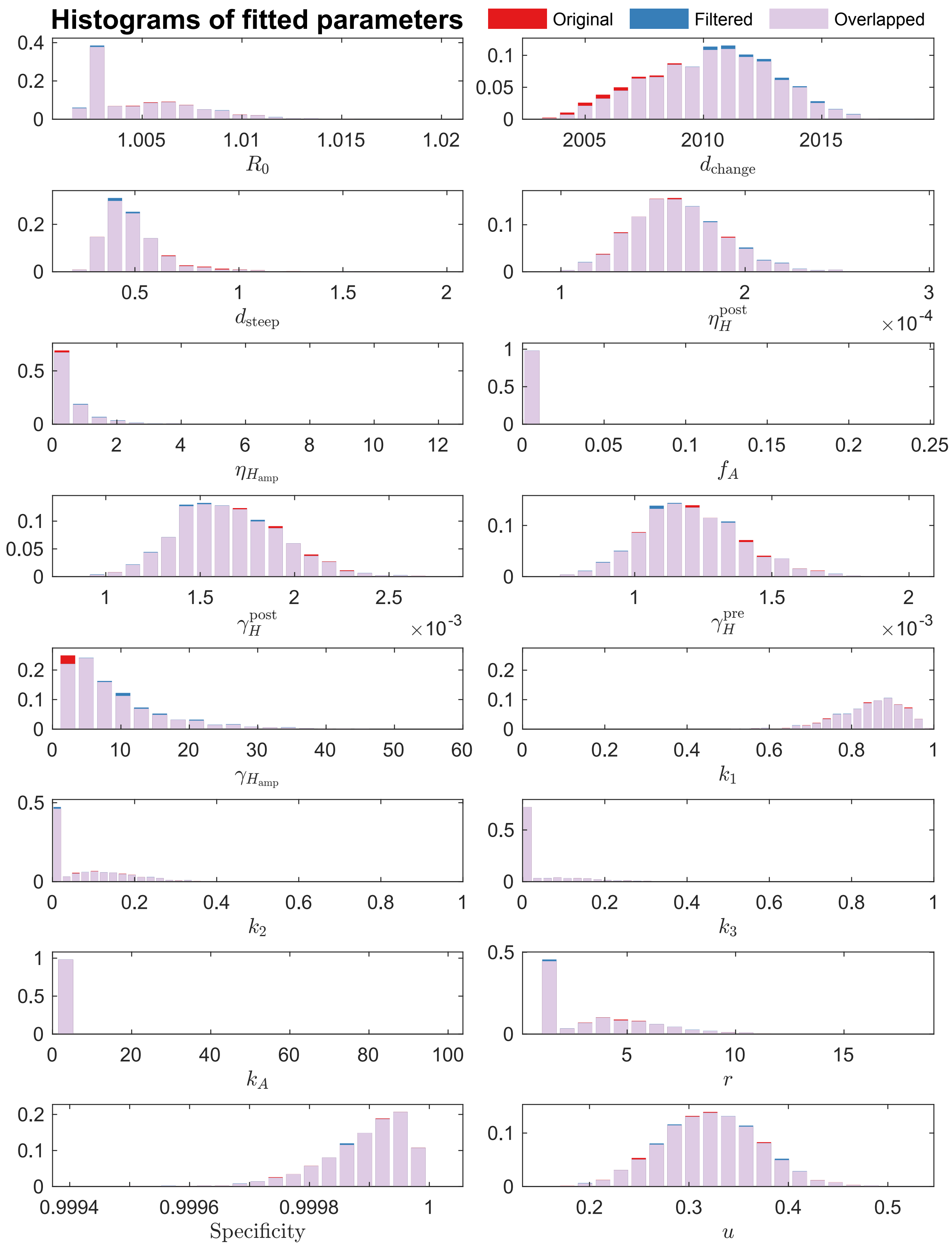

Figure S16: **Histograms of model fitted parameters for Koboko.** The histograms are normalised to illustrate the probability density of fitted parameters. Two sets of histograms are presented in each figure to highlight the changes in parameter distributions arising from discarding realisations that do not match the observed presence/absence of reported cases in 2020–2024. The original (non-filtered) ensemble posterior results are shown in red, and the filtered results are shown in blue. Their overlaps are coloured in light purple. Of the 18,921 realisations that survived filtering, we randomly select 20,000 samples to form the new ensemble posterior for simulations. See Table S4 for the descriptions of fitted parameters.

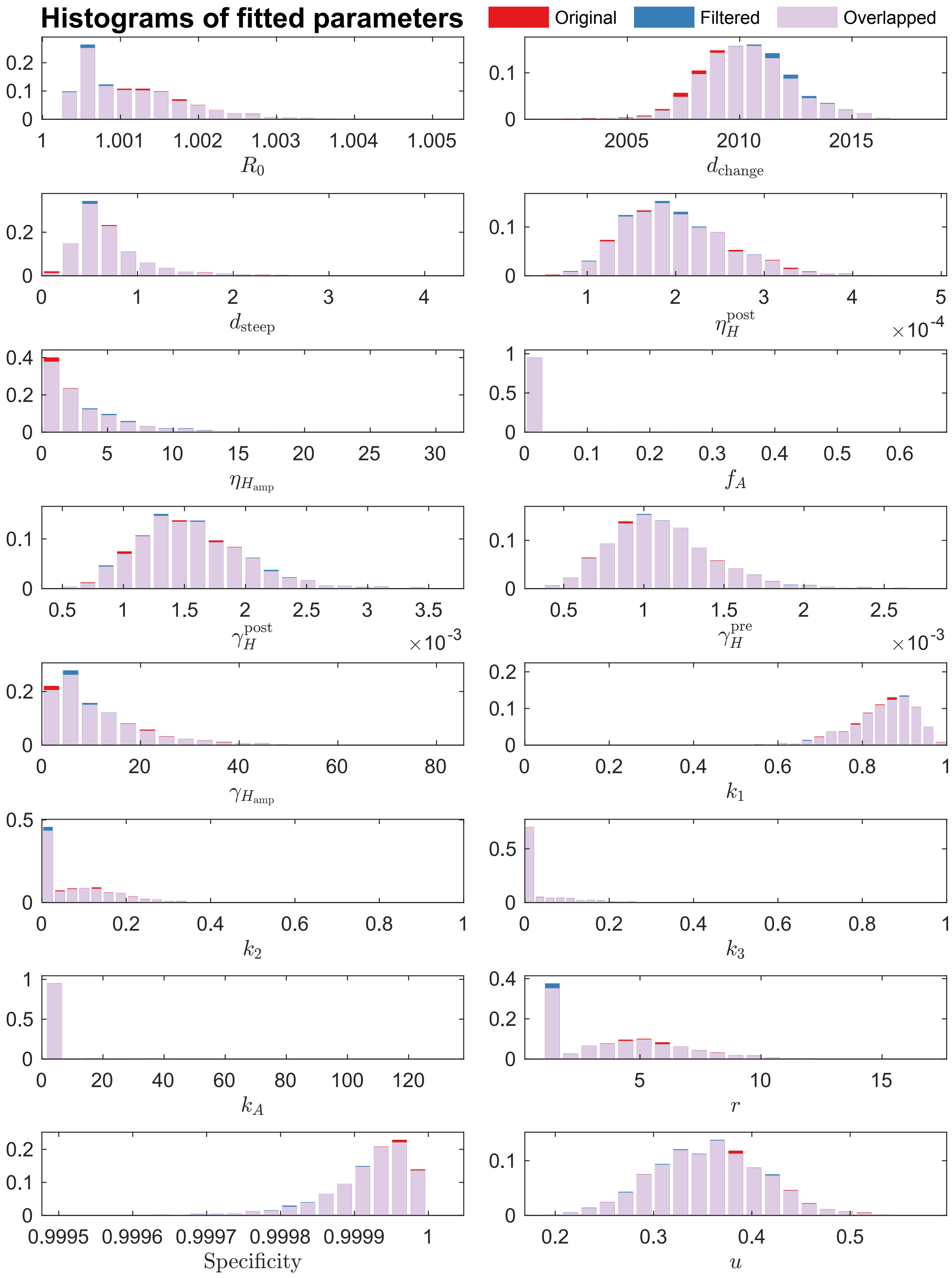

Figure S17: **Histograms of model fitted parameters for Maracha.** The histograms are normalised to illustrate the probability density of fitted parameters. Two sets of histograms are presented in each figure to highlight the changes in parameter distributions arising from discarding realisations that do not match the observed presence/absence of reported cases in 2020–2024. The original (non-filtered) ensemble posterior results are shown in red, and the filtered results are shown in blue. Their overlaps are coloured in light purple. Of the 19,932 realisations that survived filtering, we randomly select 20,000 samples to form the new ensemble posterior for simulations. See Table S4 for the descriptions of fitted parameters.

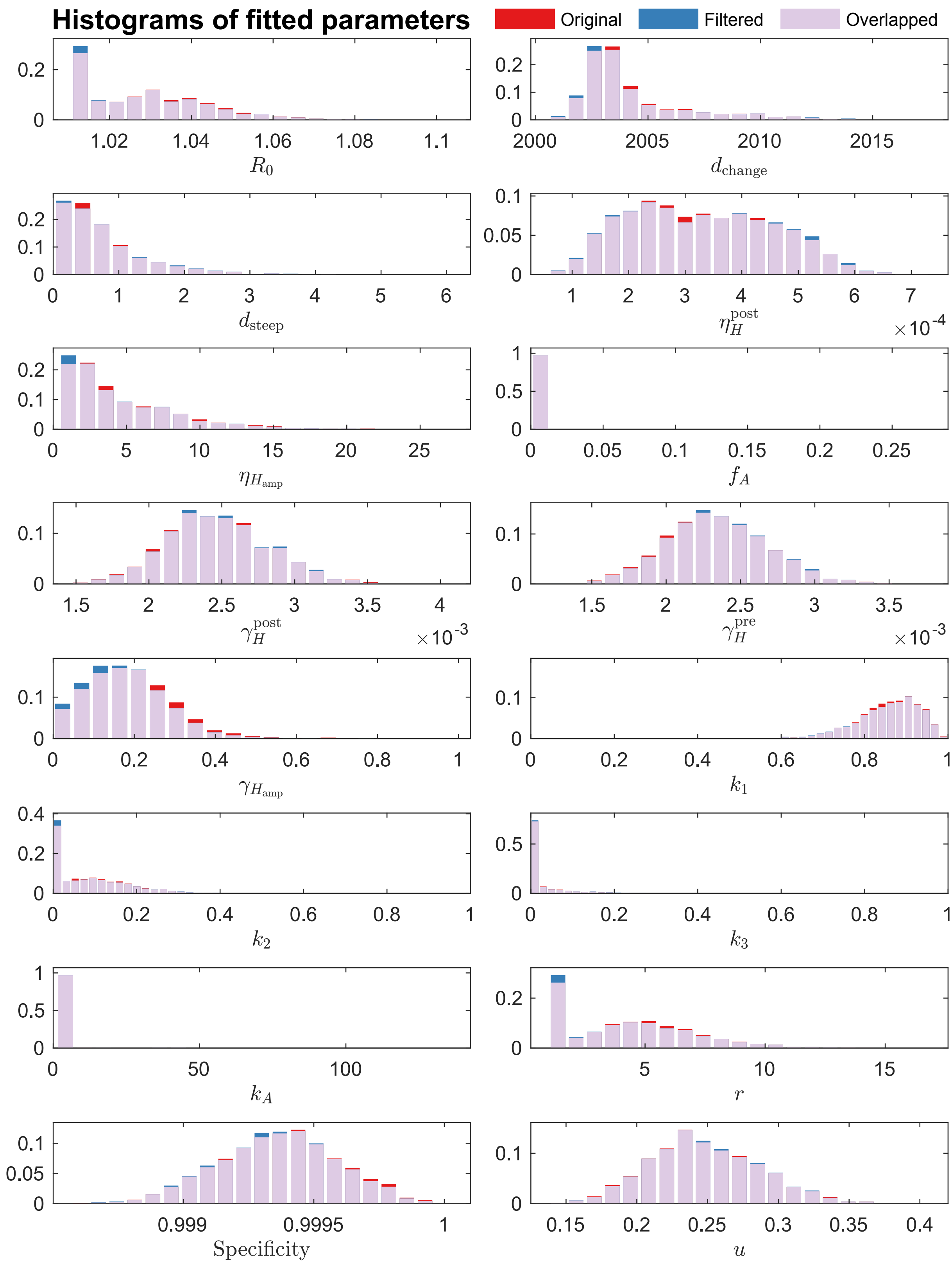

Figure S18: **Histograms of model fitted parameters for Moyo.** The histograms are normalised to illustrate the probability density of fitted parameters. Two sets of histograms are presented in each figure to highlight the changes in parameter distributions arising from discarding realisations that do not match the observed presence/absence of reported cases in 2020–2024. The original (non-filtered) ensemble posterior results are shown in red, and the filtered results are shown in blue. Their overlaps are coloured in light purple. Of the 19,710 realisations that survived filtering, we randomly select 20,000 samples to form the new ensemble posterior for simulations. See Table S4 for the descriptions of fitted parameters.

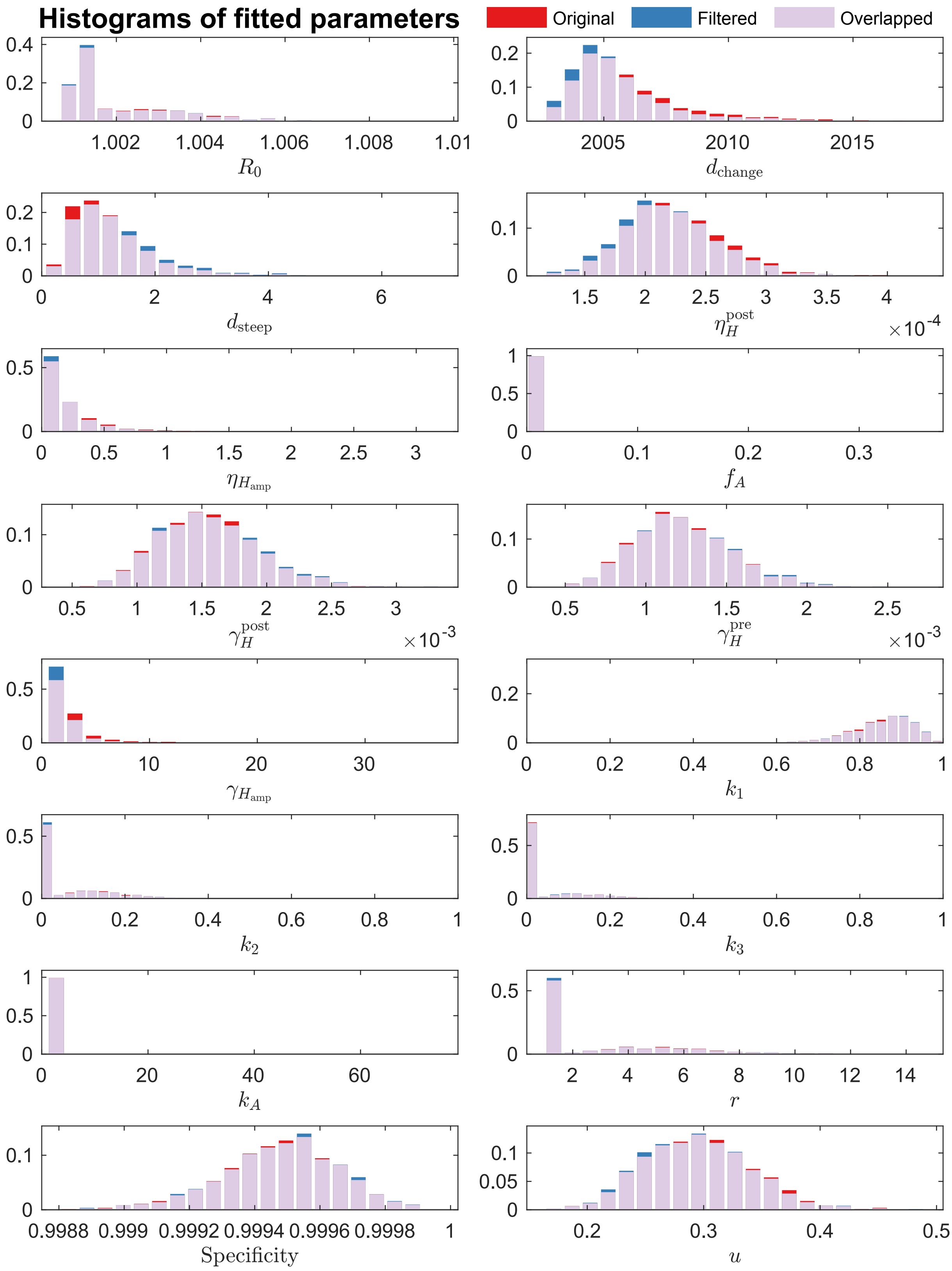

Figure S19: **Histograms of model fitted parameters for Yumbe.** The histograms are normalised to illustrate the probability density of fitted parameters. Two sets of histograms are presented in each figure to highlight the changes in parameter distributions arising from discarding realisations that do not match the observed presence/absence of reported cases in 2020–2024. The original (non-filtered) ensemble posterior results are shown in red, and the filtered results are shown in blue. Their overlaps are coloured in light purple. Of the 18,770 realisations that survived filtering, we randomly select 20,000 samples to form the new ensemble posterior for simulations. See Table S4 for the descriptions of fitted parameters.

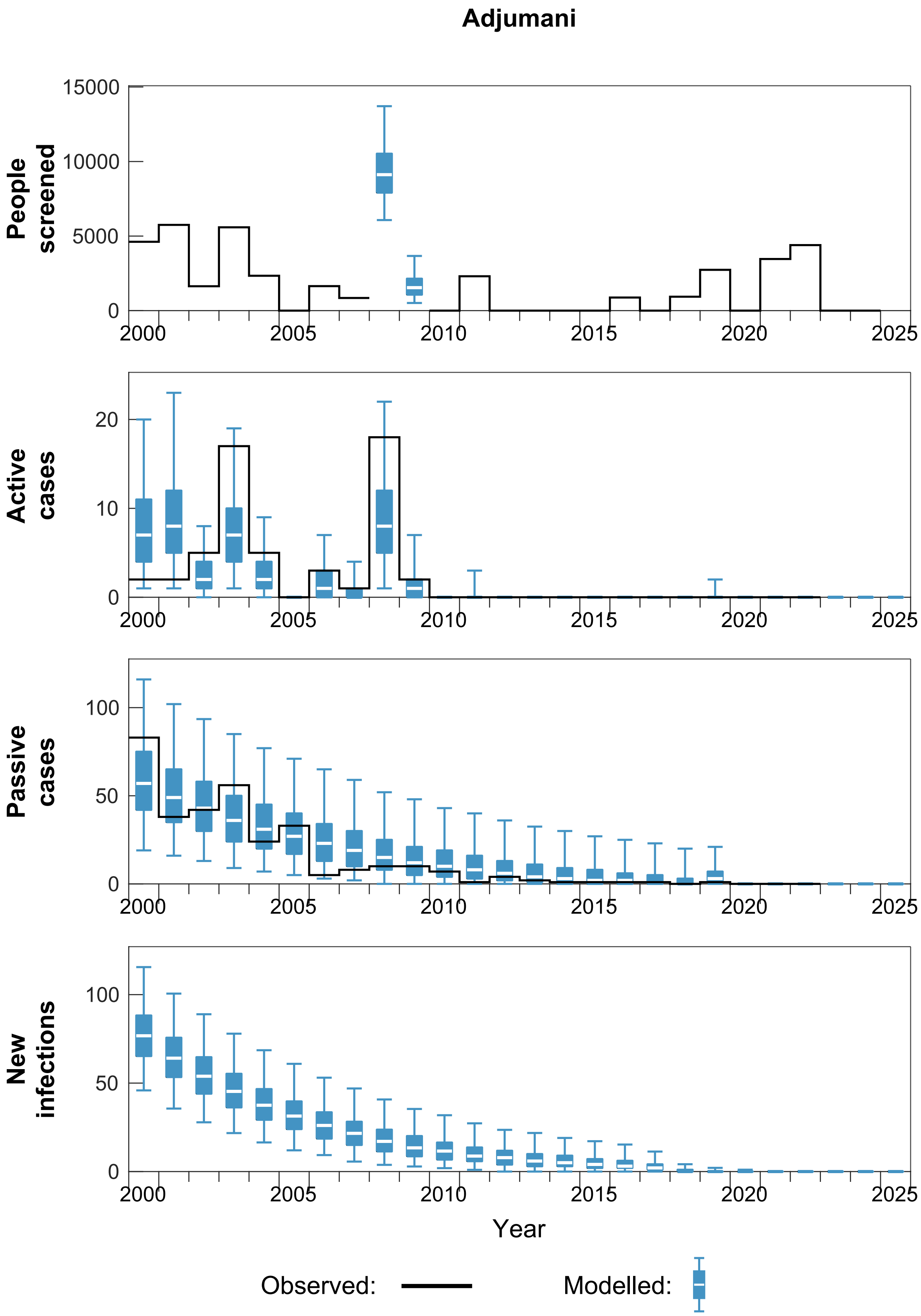

Figure S20: **Ensemble fitting results for Adjumani.** There are n = 20,000 independent realisations, filtered and randomly selected from 250 samples from each of 2,000 independent samples from the joint ensemble posterior distributions of the fitted model parameters. Box plots summarise parameter and observational uncertainty. The lines in the boxes represent the medians of predicted results. The lower and upper bounds of the boxes indicate 25th and 75th percentiles. The minimum and maximum values are 2.5th and 97.5th percentiles and therefore whiskers cover 95% prediction intervals.

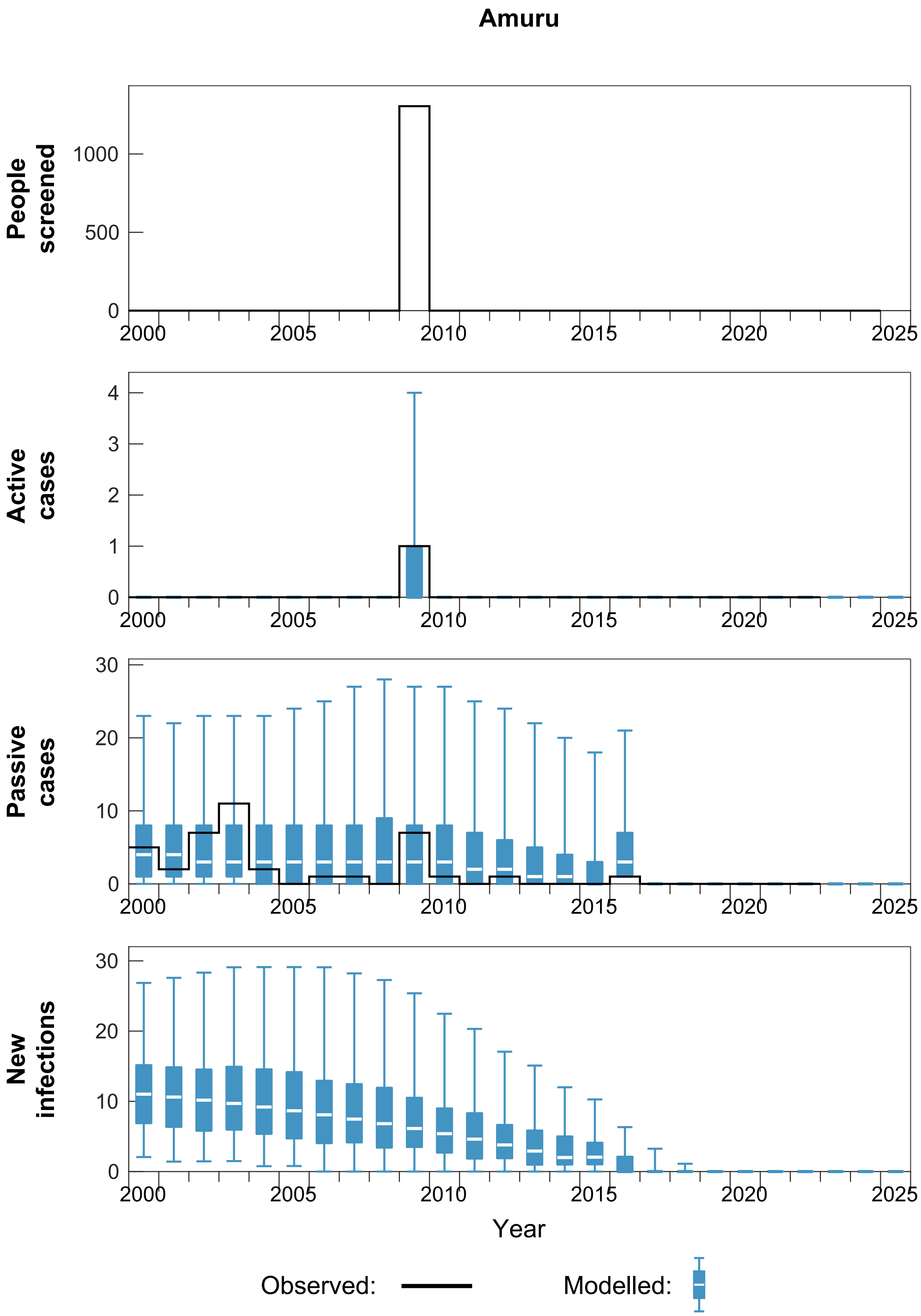

Figure S21: **Ensemble fitting results for Amuru.** There are n = 20,000 independent realisations, filtered and randomly selected from 250 samples from each of 2,000 independent samples from the joint ensemble posterior distributions of the fitted model parameters. Box plots summarise parameter and observational uncertainty. The lines in the boxes represent the medians of predicted results. The lower and upper bounds of the boxes indicate 25th and 75th percentiles. The minimum and maximum values are 2.5th and 97.5th percentiles and therefore whiskers cover 95% prediction intervals.

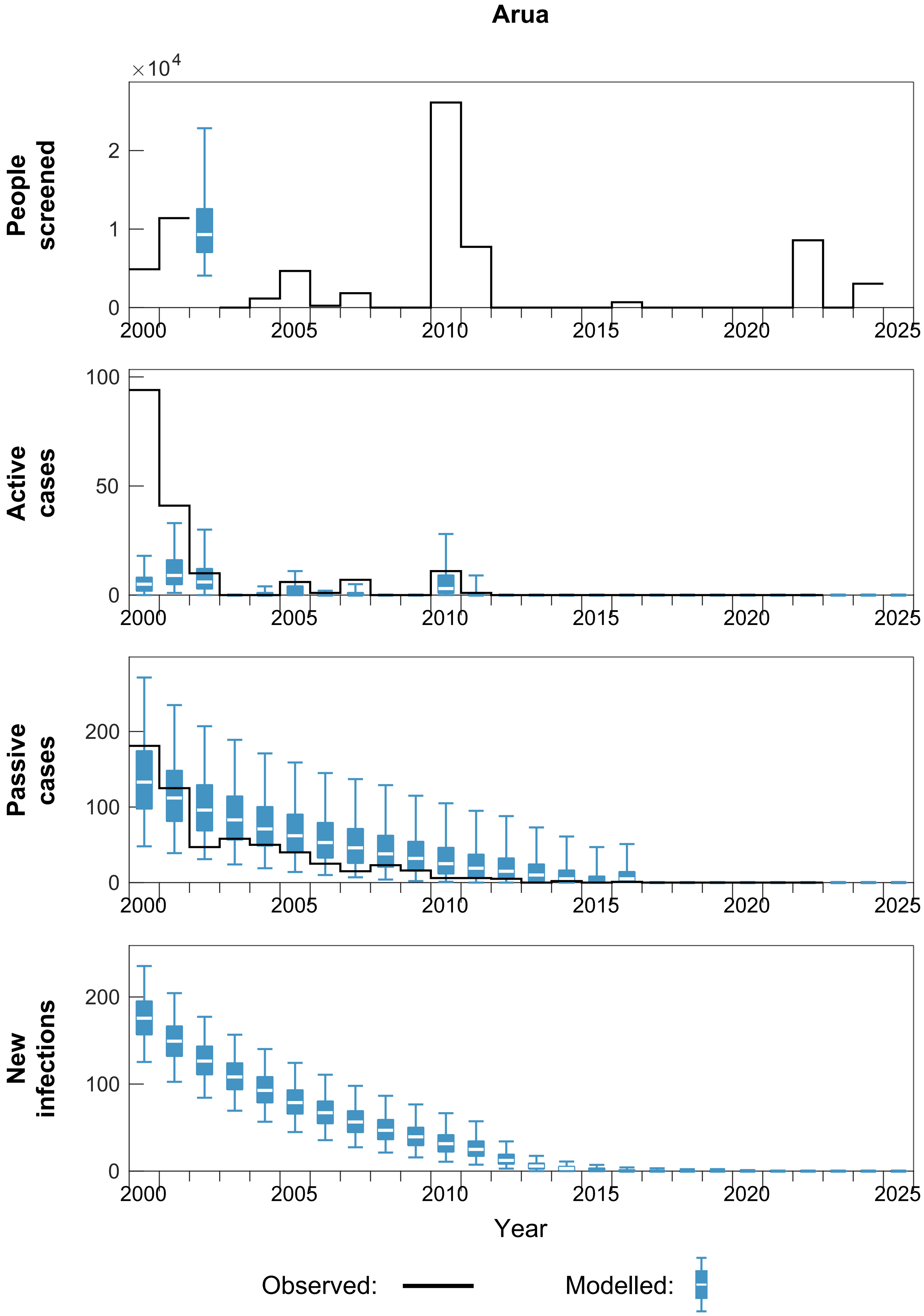

Figure S22: **Ensemble fitting results for Arua.** There are n = 20,000 independent realisations, filtered and randomly selected from 250 samples from each of 2,000 independent samples from the joint ensemble posterior distributions of the fitted model parameters. Box plots summarise parameter and observational uncertainty. The lines in the boxes represent the medians of predicted results. The lower and upper bounds of the boxes indicate 25th and 75th percentiles. The minimum and maximum values are 2.5th and 97.5th percentiles and therefore whiskers cover 95% prediction intervals.

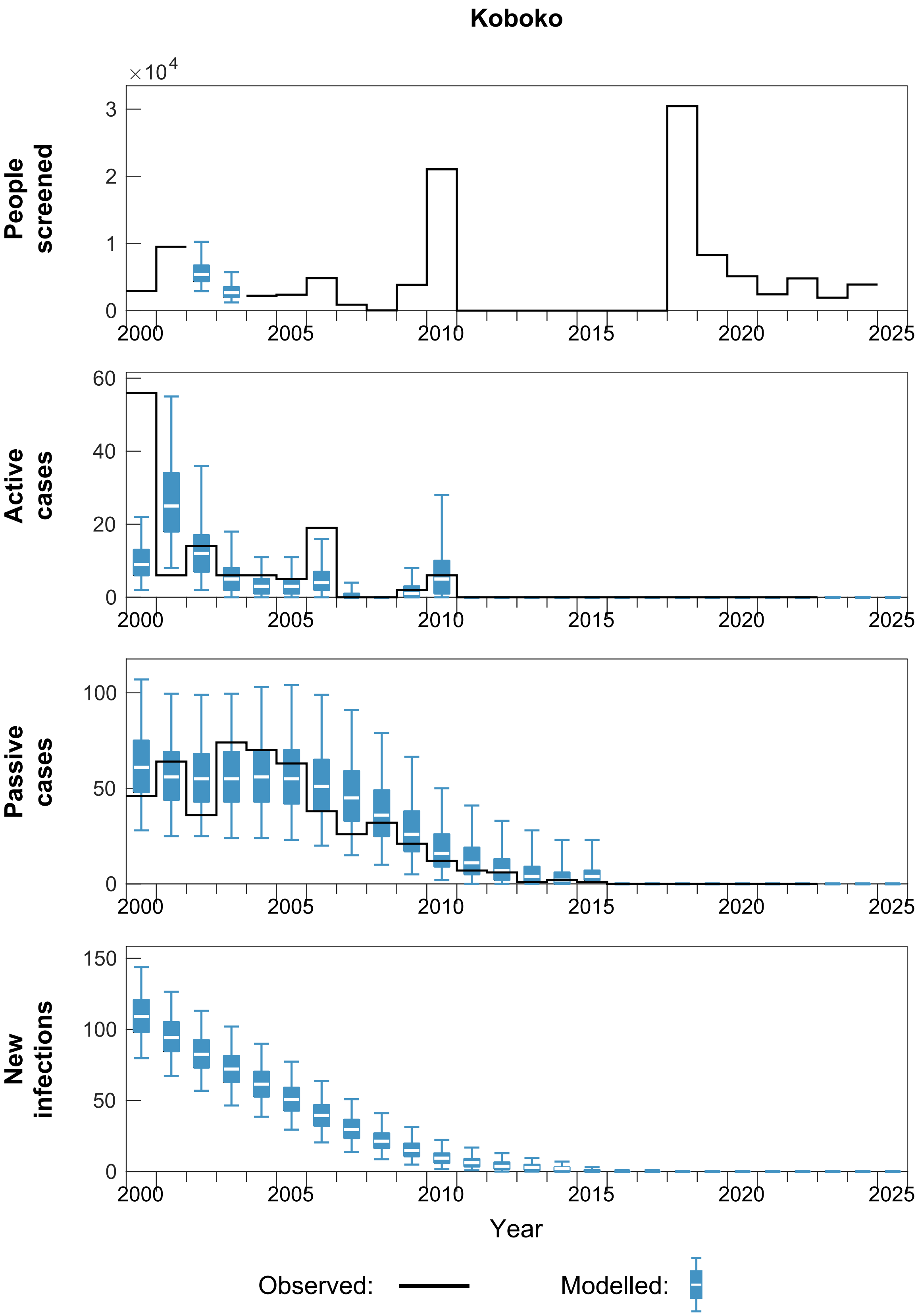

Figure S23: **Ensemble fitting results for Koboko.** There are n = 20,000 independent realisations, filtered and randomly selected from 250 samples from each of 2,000 independent samples from the joint ensemble posterior distributions of the fitted model parameters. Box plots summarise parameter and observational uncertainty. The lines in the boxes represent the medians of predicted results. The lower and upper bounds of the boxes indicate 25th and 75th percentiles. The minimum and maximum values are 2.5th and 97.5th percentiles and therefore whiskers cover 95% prediction intervals.

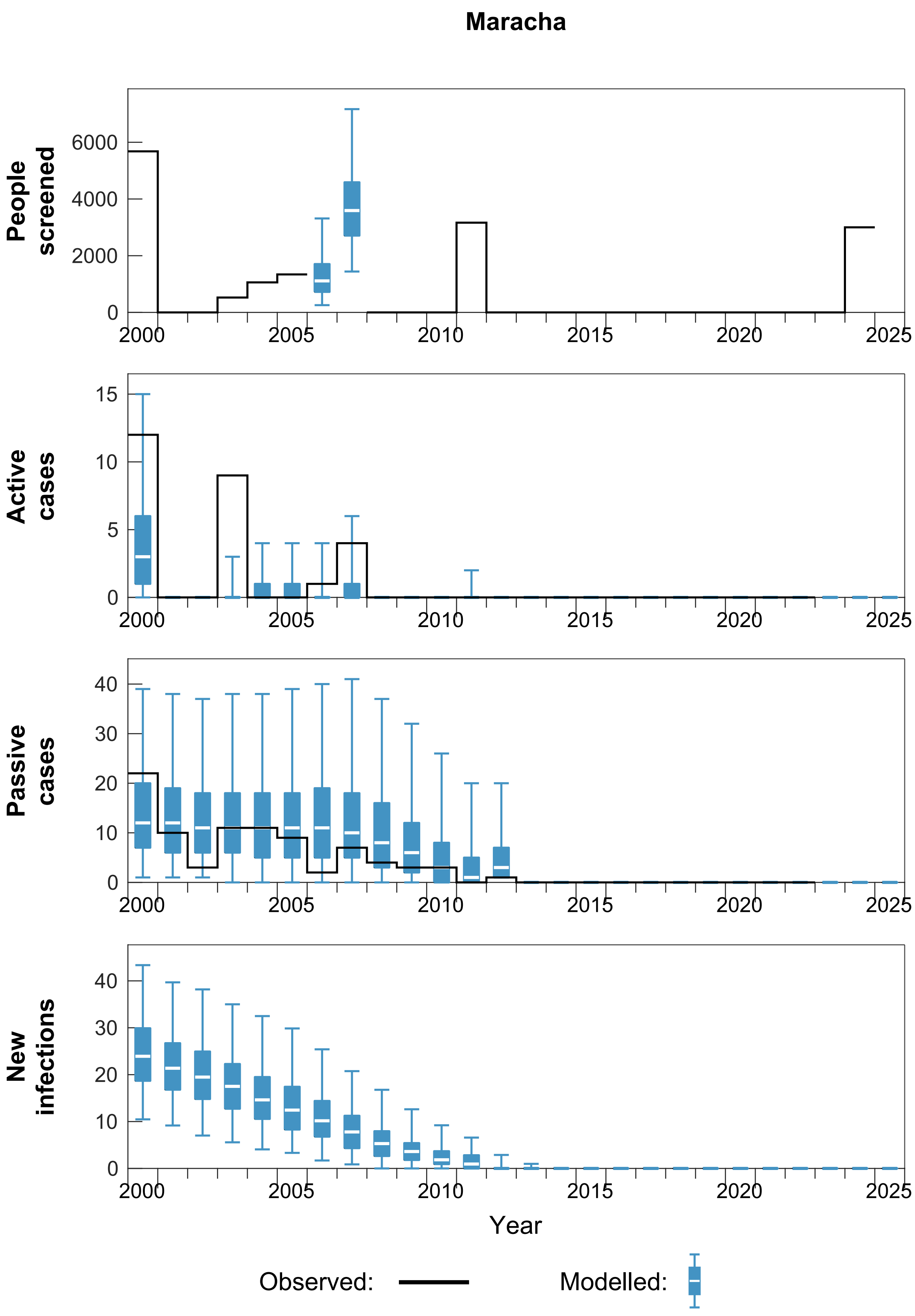

Figure S24: **Ensemble fitting results for Maracha.** There are n = 20,000 independent realisations, filtered and randomly selected from 250 samples from each of 2,000 independent samples from the joint ensemble posterior distributions of the fitted model parameters. Box plots summarise parameter and observational uncertainty. The lines in the boxes represent the medians of predicted results. The lower and upper bounds of the boxes indicate 25th and 75th percentiles. The minimum and maximum values are 2.5th and 97.5th percentiles and therefore whiskers cover 95% prediction intervals.

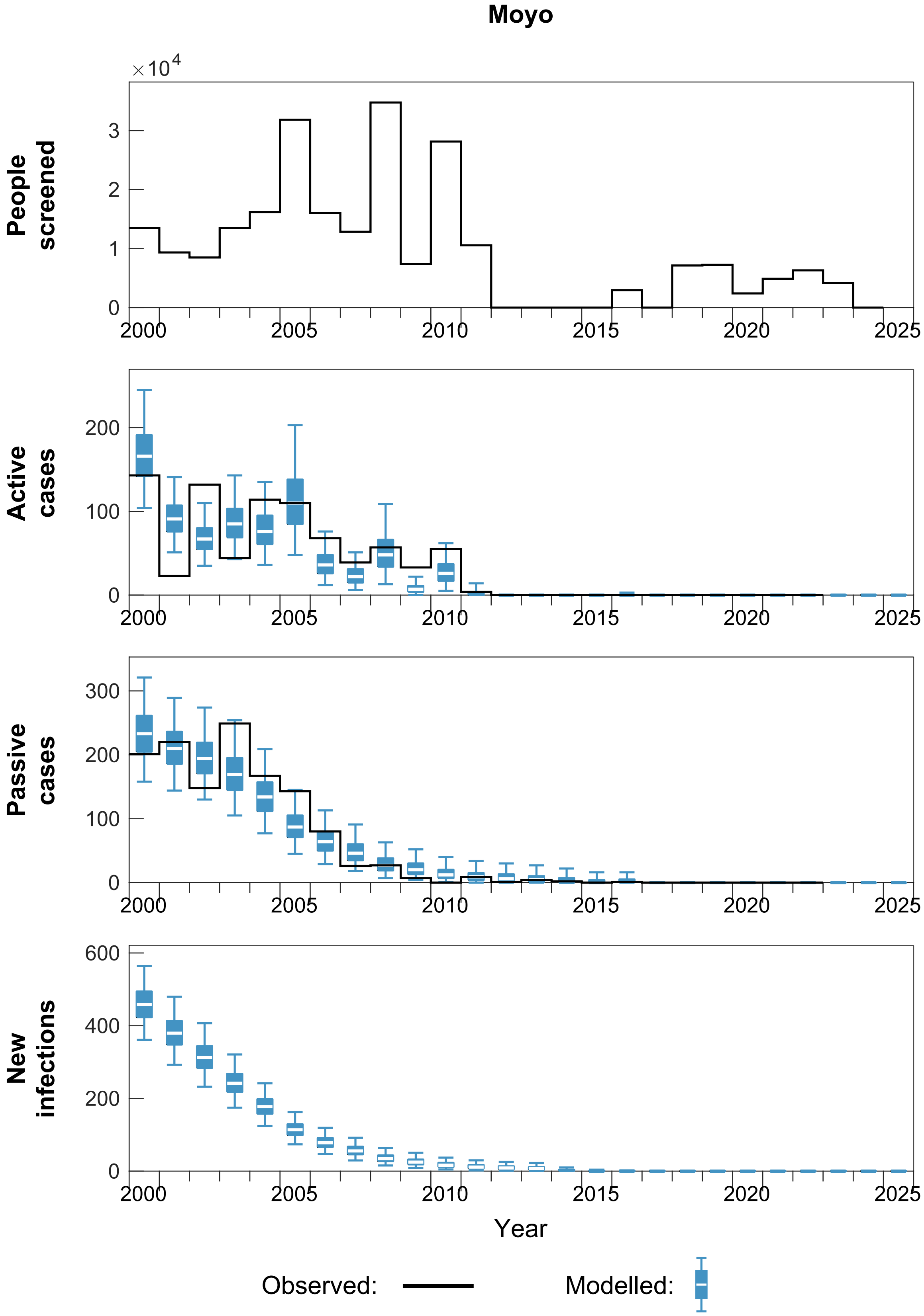

Figure S25: **Ensemble fitting results for Moyo.** There are n = 20,000 independent realisations, filtered and randomly selected from 250 samples from each of 2,000 independent samples from the joint ensemble posterior distributions of the fitted model parameters. Box plots summarise parameter and observational uncertainty. The lines in the boxes represent the medians of predicted results. The lower and upper bounds of the boxes indicate 25th and 75th percentiles. The minimum and maximum values are 2.5th and 97.5th percentiles and therefore whiskers cover 95% prediction intervals.

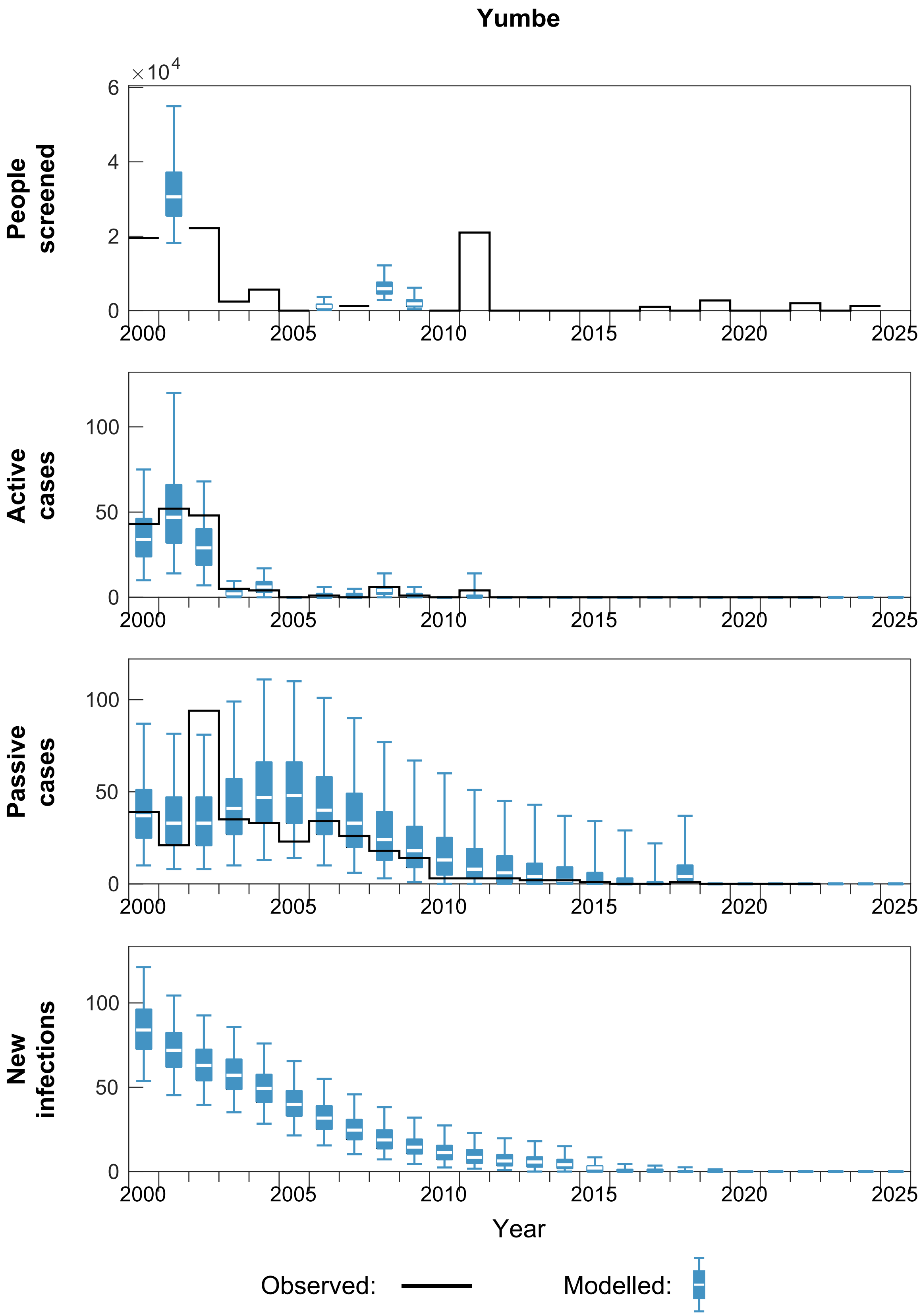

Figure S26: **Ensemble fitting results for Yumbe.** There are n = 20,000 independent realisations, filtered and randomly selected from 250 samples from each of 2,000 independent samples from the joint ensemble posterior distributions of the fitted model parameters. Box plots summarise parameter and observational uncertainty. The lines in the boxes represent the medians of predicted results. The lower and upper bounds of the boxes indicate 25th and 75th percentiles. The minimum and maximum values are 2.5th and 97.5th percentiles and therefore whiskers cover 95% prediction intervals.

#### S11.4 Projections

Figures S27–S33 show the district-level projections results for seven districts. Table S20 summarises ranges of elimination years (LTE and NRI).

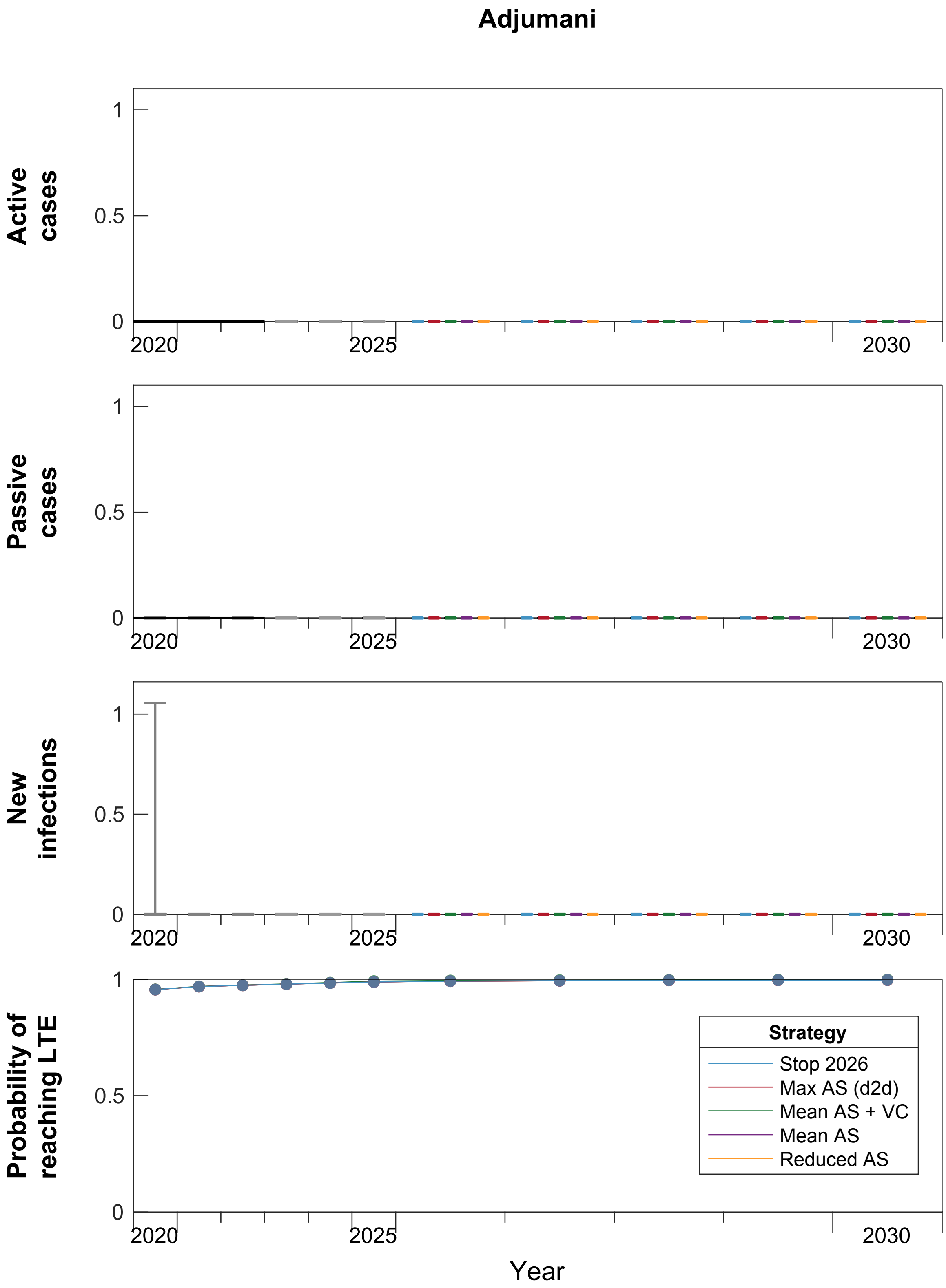

Figure S27: **Ensemble projection results under five strategies for Adjumani.** Passive screening is considered in all strategies. There are n = 20,000 independent realisations, filtered and randomly selected from 250 samples from each of 2,000 independent samples from the joint ensemble posterior distributions of the fitted model parameters. Box plots summarise parameter and observational uncertainty. The lines in the boxes represent the medians of predicted results. The lower and upper bounds of the boxes indicate 25th and 75th percentiles. The minimum and maximum values are 2.5th and 97.5th percentiles and therefore whiskers cover 95% prediction intervals. Abbreviations: AS: active screening; VC: vector control; LTE: Last transmission event

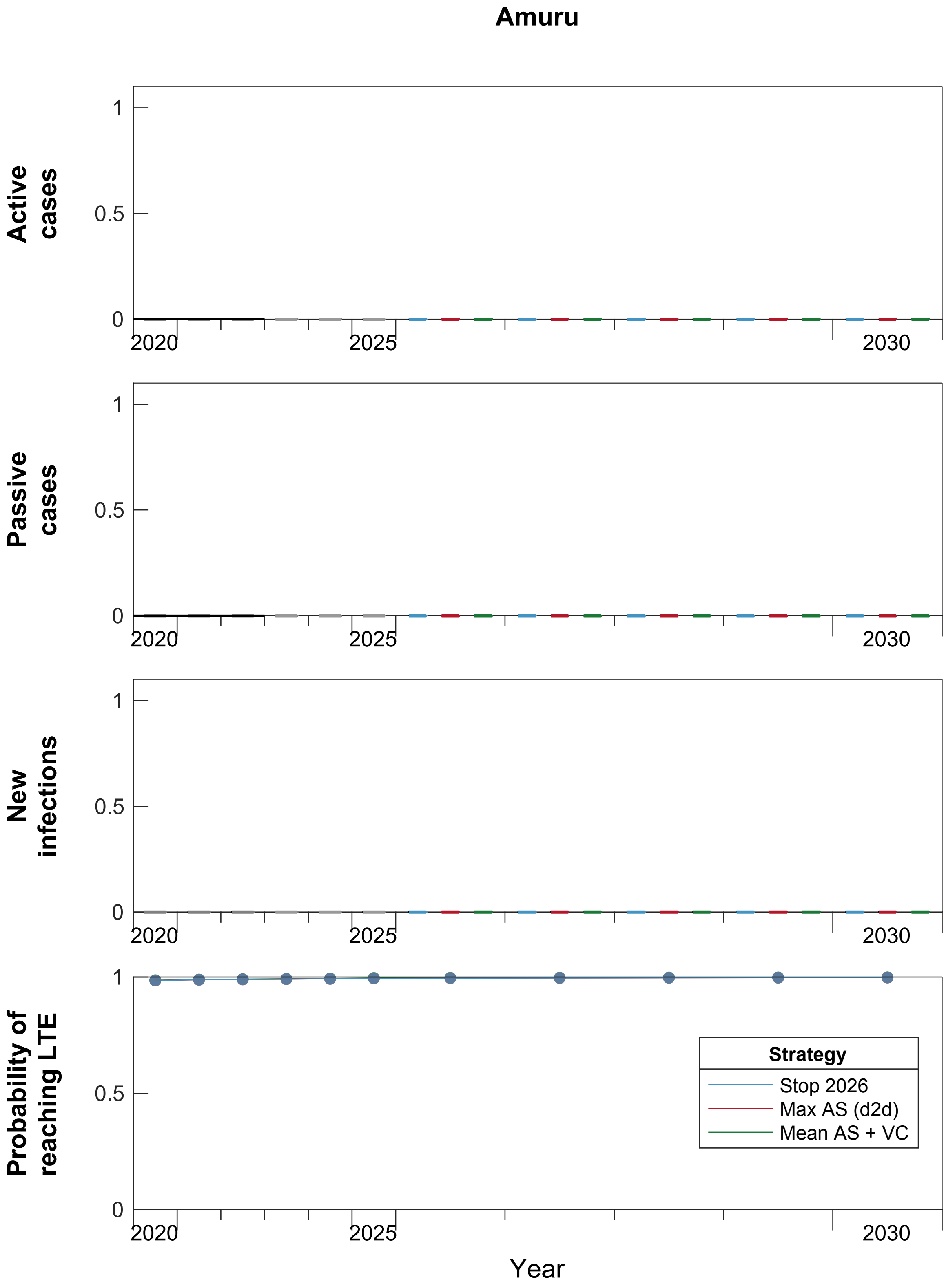

Figure S28: **Ensemble projection results under three strategies for Amuru.** Passive screening is considered in all strategies. There are n = 20,000 independent realisations, filtered and randomly selected from 250 samples from each of 2,000 independent samples from the joint ensemble posterior distributions of the fitted model parameters. Box plots summarise parameter and observational uncertainty. The lines in the boxes represent the medians of predicted results. The lower and upper bounds of the boxes indicate 25th and 75th percentiles. The minimum and maximum values are 2.5th and 97.5th percentiles and therefore whiskers cover 95% prediction intervals. N.B. There have been no screenings in 2020–2024, and therefore only three strategies are considered in Amuru. Abbreviations: AS: active screening; VC: vector control; LTE: Last transmission event

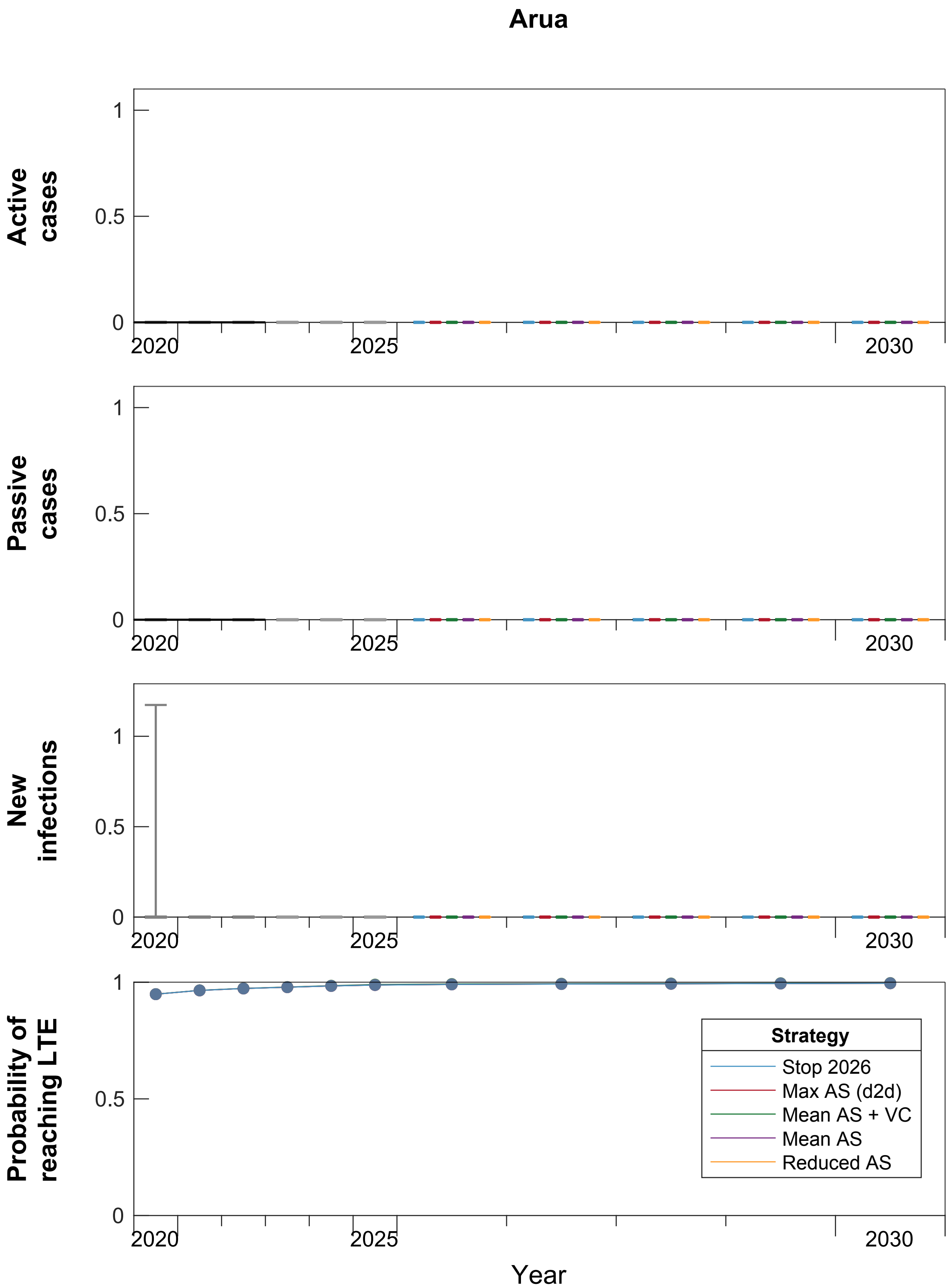

Figure S29: **Ensemble projection results under five strategies for Arua.** Passive screening is considered in all strategies. There are n = 20,000 independent realisations, filtered and randomly selected from 250 samples from each of 2,000 independent samples from the joint ensemble posterior distributions of the fitted model parameters. Box plots summarise parameter and observational uncertainty. The lines in the boxes represent the medians of predicted results. The lower and upper bounds of the boxes indicate 25th and 75th percentiles. The minimum and maximum values are 2.5th and 97.5th percentiles and therefore whiskers cover 95% prediction intervals. Abbreviations: AS: active screening; VC: vector control; LTE: Last transmission event

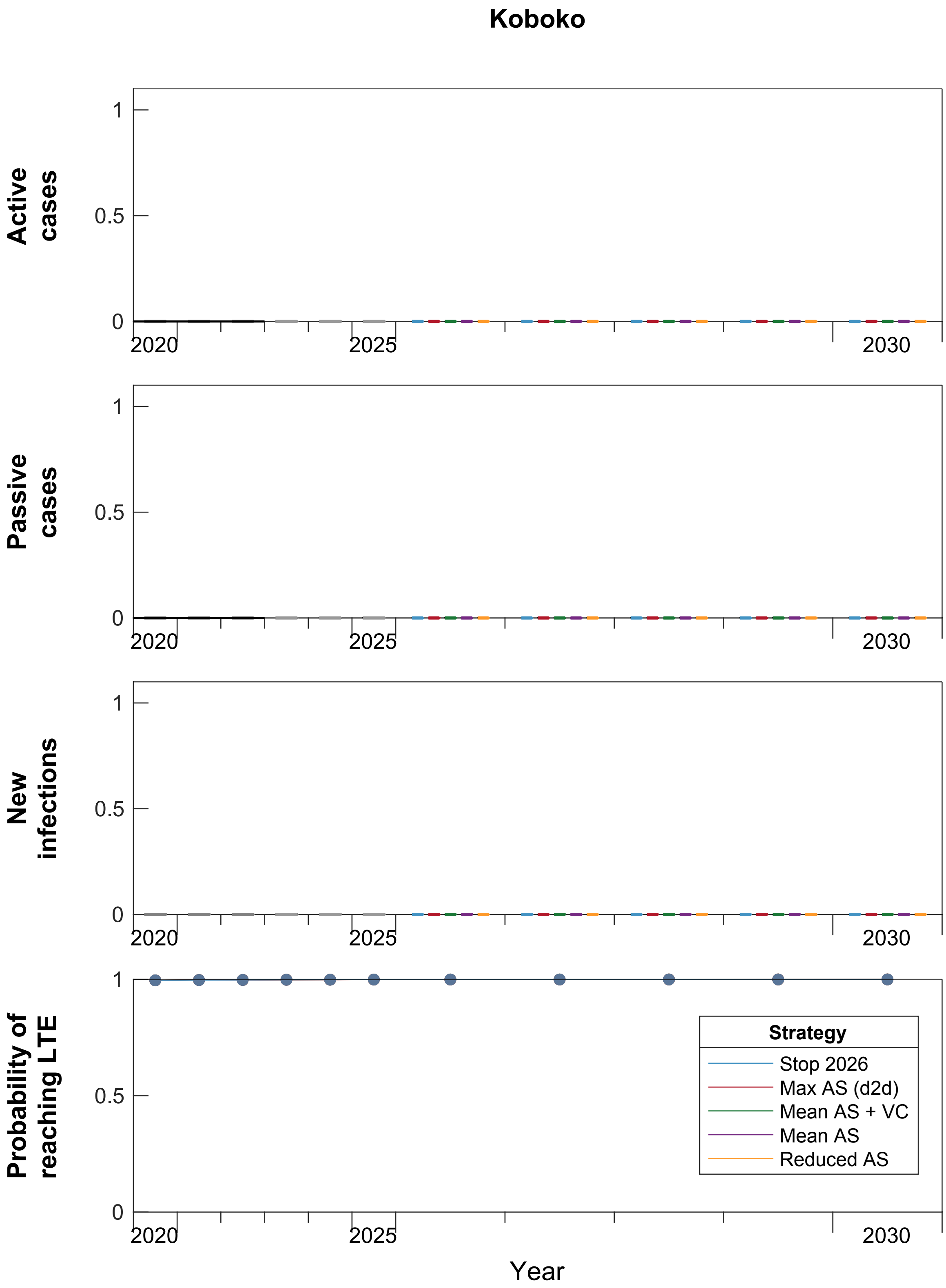

Figure S30: **Ensemble projection results under five strategies for Koboko.** Passive screening is considered in all strategies. There are n = 20,000 independent realisations, filtered and randomly selected from 250 samples from each of 2,000 independent samples from the joint ensemble posterior distributions of the fitted model parameters. Box plots summarise parameter and observational uncertainty. The lines in the boxes represent the medians of predicted results. The lower and upper bounds of the boxes indicate 25th and 75th percentiles. The minimum and maximum values are 2.5th and 97.5th percentiles and therefore whiskers cover 95% prediction intervals. Abbreviations: AS: active screening; VC: vector control; LTE: Last transmission event

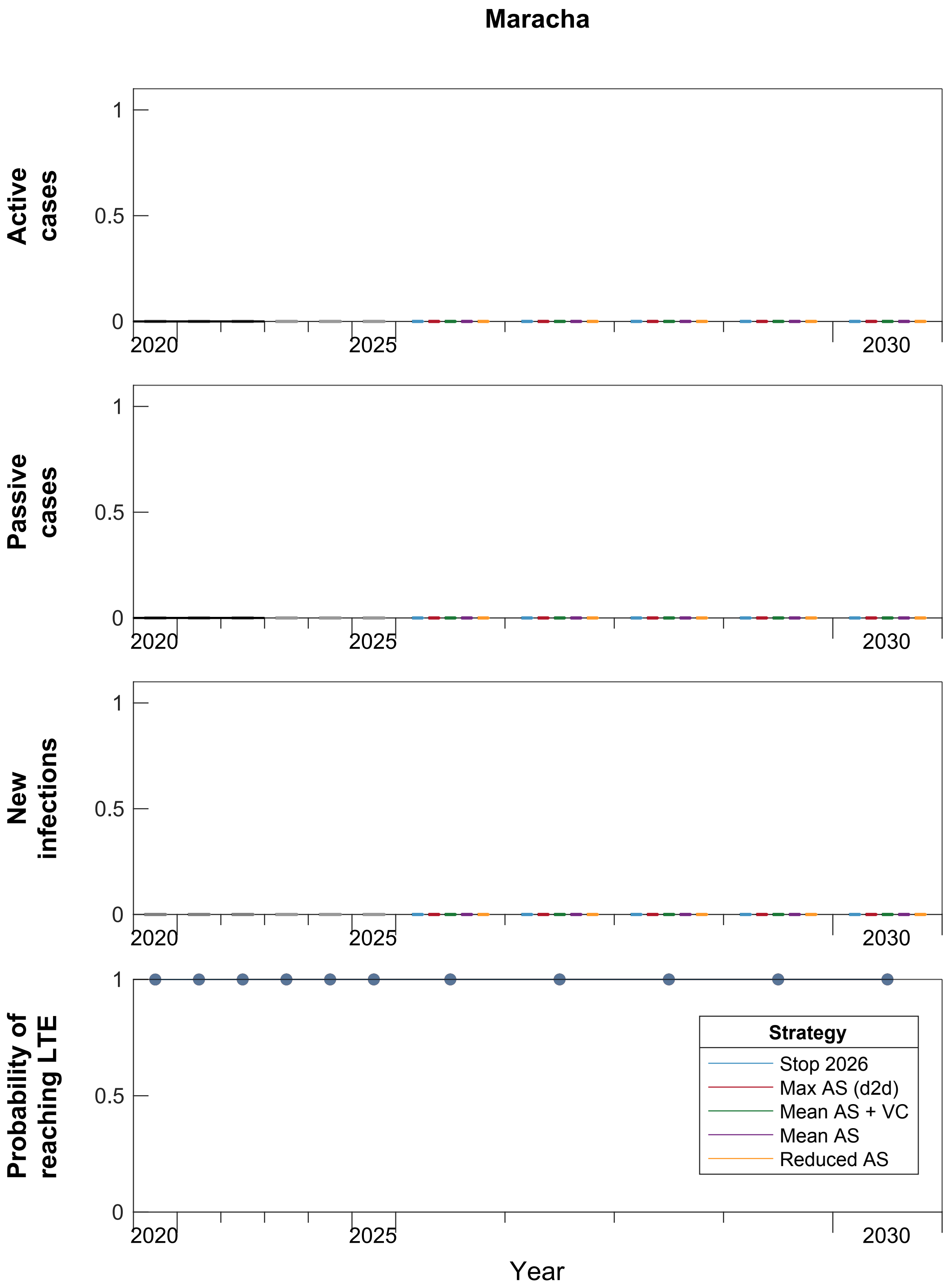

Figure S31: **Ensemble projection results under three strategies for Maracha.** Passive screening is considered in all strategies. There are n = 20,000 independent realisations, filtered and randomly selected from 250 samples from each of 2,000 independent samples from the joint ensemble posterior distributions of the fitted model parameters. Box plots summarise parameter and observational uncertainty. The lines in the boxes represent the medians of predicted results. The lower and upper bounds of the boxes indicate 25th and 75th percentiles. The minimum and maximum values are 2.5th and 97.5th percentiles and therefore whiskers cover 95% prediction intervals. Abbreviations: AS: active screening; VC: vector control; LTE: Last transmission event

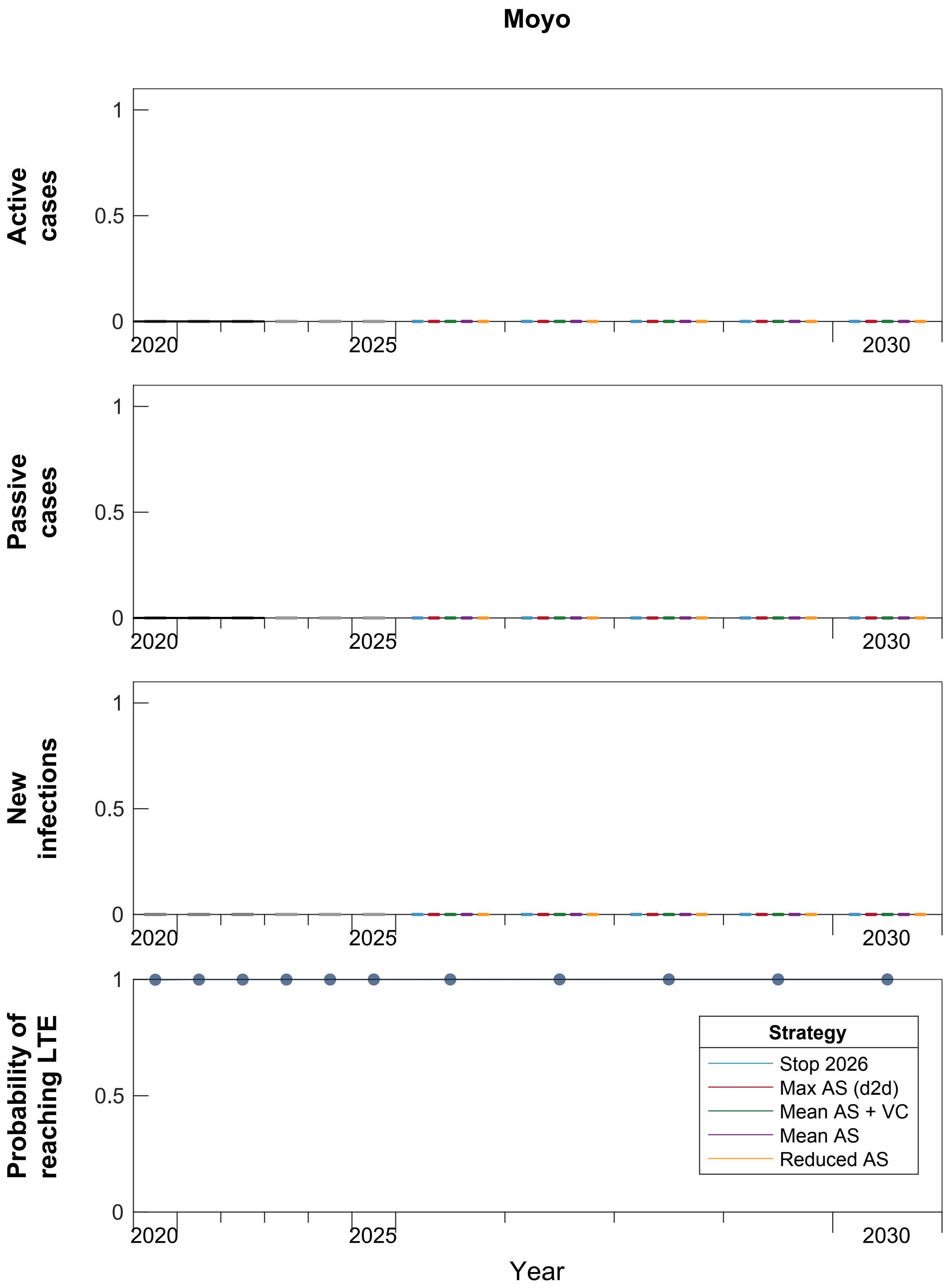

Figure S32: **Ensemble projection results under five strategies for Moyo.** Passive screening is considered in all strategies. There are n = 20,000 independent realisations, filtered and randomly selected from 250 samples from each of 2,000 independent samples from the joint ensemble posterior distributions of the fitted model parameters. Box plots summarise parameter and observational uncertainty. The lines in the boxes represent the medians of predicted results. The lower and upper bounds of the boxes indicate 25th and 75th percentiles. The minimum and maximum values are 2.5th and 97.5th percentiles and therefore whiskers cover 95% prediction intervals. Abbreviations: AS: active screening; VC: vector control; LTE: Last transmission event

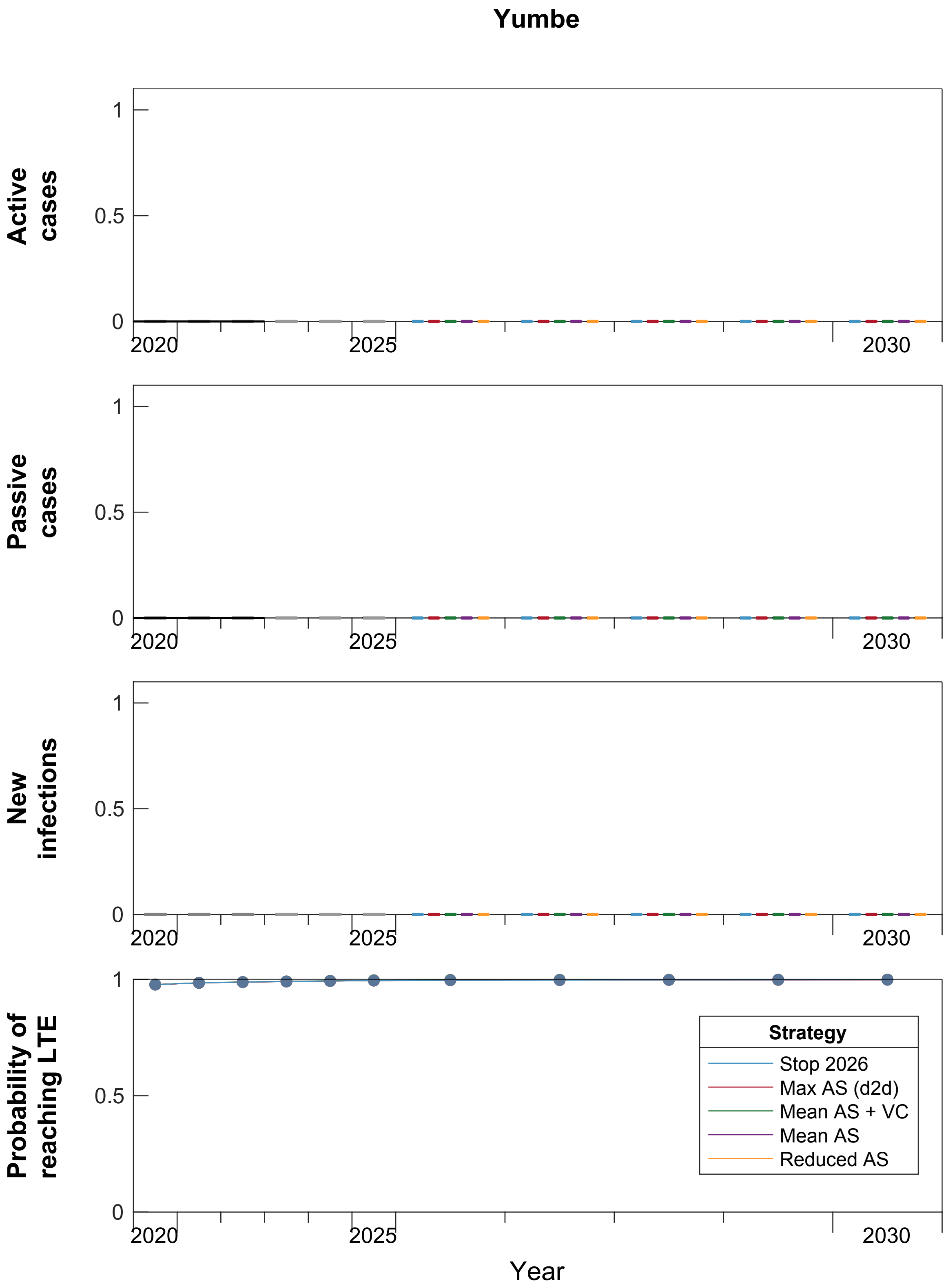

Figure S33: **Ensemble projection results under five strategies for Yumbe.** Passive screening is considered in all strategies. There are n = 20,000 independent realisations, filtered and randomly selected from 250 samples from each of 2,000 independent samples from the joint ensemble posterior distributions of the fitted model parameters. Box plots summarise parameter and observational uncertainty. The lines in the boxes represent the medians of predicted results. The lower and upper bounds of the boxes indicate 25th and 75th percentiles. The minimum and maximum values are 2.5th and 97.5th percentiles and therefore whiskers cover 95% prediction intervals. Abbreviations: AS: active screening; VC: vector control; LTE: Last transmission event

Table S20: **Predicted years ([2.5, 50, & 97.5%] PI) to achieve different elimination indicators.** All districts have had their last transmission event and reported their last case before entering the projection period, and therefore, the prediction intervals of years of last transmission event do not vary under different strategies. Koboko, Maracha, Moyo, and Yumbe have completely got rid of gHAT infections before entering projections. In Adjumani and Arua, the 97.5th percentile of modelling NRI prediction interval is 2026 under all strategies. A small variation in NRI among different strategies (2027 for the *Mean AS + VC* strategy and 2028 for strategies without VC) can be found in Amuru.

| District | Indicator | |
|---|---|---|
| | Last transmission event (LTE) | No remaining infection (NRI) |
| Adjumani | [2015, 2017, 2023] | [2019, 2020, 2026] |
| Amuru | [2013, 2016, 2019] | [2016, 2017, 2028] |
| Arua | [2013, 2016, 2023] | [2016, 2019, 2026] |
| Koboko | [2012, 2014, 2017] | [2015, 2016, 2021] |
| Maracha | [2009, 2011, 2014] | [2012, 2013, 2019] |
| Moyo | [2013, 2014, 2017] | [2016, 2017, 2022] |
| Yumbe | [2014, 2016, 2020] | [2018, 2019, 2024] |

### S12 Additional results from the economic model

Table S21: Results in Adjumani. Summary of effects, costs, elimination of transmission (EoT) by 2030, and cost-effectiveness with and without uncertainty. Means are given along with 95% prediction intervals (PIs). YLL: years of life lost (to fatal disease), YLD: years of life lost to disability, DALYs: disability-adjusted life-years, PS: passive screening, AS: active screening, VC: vector control, ICER: incremental cost-effectiveness ratio, WTP: willingness to pay (USD per DALY averted), EoT: elimination of transmission.

| | Stop 2026 | Max AS (d2d) | Mean AS + VC | Mean AS | Reduced AS |
|---|---|---|---|---|---|
| **Health effects** | | | | | |
| Reported cases | 0 (0-0) | 0 (0-0) | 0 (0-0) | 0 (0-0) | 0 (0-0) |
| Deaths undetected | 0 (0-0) | 0 (0-0) | 0 (0-0) | 0 (0-0) | 0 (0-0) |
| Cases total | 0 (0-0) | 0 (0-0) | 0 (0-0) | 0 (0-0) | 0 (0-0) |
| Deaths detected[a] | 0 (0-0) | 0 (0-0) | 0 (0-0) | 0 (0-0) | 0 (0-0) |
| YLD | 0 (0-0) | 0 (0-0) | 0 (0-0) | 0 (0-0) | 0 (0-0) |
| YLL | 0 (0-2) | 0 (0-2) | 0 (0-1) | 0 (0-2) | 0 (0-2) |
| DALYs | 0.103 (<0.001-1.891) | 0.069 (<0.001-1.902) | 0.090 (<0.001-1.408) | 0.100 (<0.001-1.931) | 0.103 (<0.001-1.929) |
| **Costs, in US$** | | | | | |
| AS costs | 0 (0-54) | 17,789 (14,651-19,016) | 4,683 (4,005-5,183) | 4,706 (4,005-5,191) | 2,363 (2,002-2,596) |
| PS costs | 39,064 (33,141-45,891) | 39,079 (33,141-45,936) | 39,017 (33,141-45,763) | 39,070 (33,141-45,891) | 39,069 (33,141-45,891) |
| VC costs | 0 (0-1,582) | 0 (0-785) | 87,548 (46,682-140,664) | 0 (0-781) | 0 (0-787) |
| Treatment costs | 0 (0-38) | 0 (0-39) | 0 (0-27) | 0 (0-39) | 0 (0-39) |
| Costs total | 40,737 (33,149-45,978) | 57,692 (49,165-62,786) | 131,274 (89,302-184,067) | 44,596 (37,606-50,535) | 42,258 (35,389-48,257) |
| **EoT** | | | | | |
| Year of EoT | 2019 (2016-2024) | 2019 (2016-2024) | 2019 (2016-2023) | 2019 (2016-2024) | 2019 (2016-2024) |
| Prob EoT 2030 | >0.99 | >0.99 | >0.99 | >0.99 | >0.99 |
| **Cost-effectiveness without uncertainty (discounted)[b]** | | | | | |
| DALYs averted | 0.000 | -0.003 | 0.237 | -0.019 | -0.018 |
| Costs averted | 0 | 16,838 | 90,226 | 3,883 | 1,555 |
| ICER | Min Cost | Dominated | 380,790 | Dominated | Dominated |
| **Cost-effectiveness with uncertainty, conditional on WTP[c].** | | | | | |
| WTP: $0 | 0.99(p) | 0 | 0 | 0 | 0.01 |
| WTP: $500 | 0.99(p) | 0 | 0 | 0 | 0.01 |
| WTP: $1000 | 0.99(p) | 0 | 0 | 0 | 0.01 |
| WTP: $2000 | 0.99(p) | 0 | 0 | 0 | 0.01 |
| WTP: $3000 | 0.99(p) | 0 | 0 | 0 | 0.01 |

[a] Detected deaths are those that occur due to treatment failure of loss-to-follow-up.

[b] Cost-effectiveness results are given for discounted DALYs and costs as per convention

[c] (p) is the preferred strategy; the strategy with the highest mean net monetary benefits

Table S22: Results in Amuru. Summary of effects, costs, elimination of transmission (EoT) by 2030, and cost-effectiveness with and without uncertainty. Means are given along with 95% prediction intervals (PIs). YLL: years of life lost (to fatal disease), YLD: years of life lost to disability, DALYs: disability-adjusted life-years, PS: passive screening, AS: active screening, VC: vector control, ICER: incremental cost-effectiveness ratio, WTP: willingness to pay (USD per DALY averted), EoT: elimination of transmission.

| | Stop 2026 | Max AS (d2d) | Mean AS + VC |
|---|---|---|---|
| **Health effects** | | | |
| Reported cases | 0 (0-0) | 0 (0-0) | 0 (0-0) |
| Deaths undetected | 0 (0-0) | 0 (0-0) | 0 (0-0) |
| Cases total | 0 (0-0) | 0 (0-0) | 0 (0-0) |
| Deaths detected[a] | 0 (0-0) | 0 (0-0) | 0 (0-0) |
| YLD | 0 (0-0) | 0 (0-0) | 0 (0-0) |
| YLL | 0 (0-1) | 0 (0-1) | 0 (0-1) |
| DALYs | <0.001 (<0.001-1.230) | <0.001 (<0.001-1.236) | <0.001 (<0.001-0.738) |
| **Costs, in US$** | | | |
| AS costs | 0 (0-15) | 3,788 (3,329-4,199) | 0 (0-7) |
| PS costs | 23,023 (19,578-26,747) | 23,023 (19,578-26,747) | 23,022 (19,578-26,746) |
| VC costs | 0 (0-253) | 0 (0-159) | 33,034 (18,959-50,723) |
| Treatment costs | 0 (0-12) | 0 (0-14) | 0 (0-8) |
| Costs total | 23,302 (19,578-26,856) | 26,983 (23,266-30,589) | 56,072 (41,403-73,892) |
| **EoT** | | | |
| Year of EoT | 2017 (2014-2020) | 2017 (2014-2020) | 2017 (2014-2020) |
| Prob EoT 2030 | >0.99 | >0.99 | >0.99 |
| **Cost-effectiveness without uncertainty (discounted)[b]** | | | |
| DALYs averted | 0.000 | -0.002 | 0.235 |
| Costs averted | 0 | 3,678 | 32,766 |
| ICER | Min Cost | Dominated | 139,312 |
| **Cost-effectiveness with uncertainty, conditional on WTP[c].** | | | |
| WTP: $0 | 1(p) | 0 | 0 |
| WTP: $500 | 1(p) | 0 | 0 |
| WTP: $1000 | 1(p) | 0 | 0 |
| WTP: $2000 | 1(p) | 0 | 0 |
| WTP: $3000 | 1(p) | 0 | 0 |

[a] Detected deaths are those that occur due to treatment failure of loss-to-follow-up.

[b] Cost-effectiveness results are given for discounted DALYs and costs as per convention

[c] (p) is the preferred strategy; the strategy with the highest mean net monetary benefits

Table S23: Results in Arua. Summary of effects, costs, elimination of transmission (EoT) by 2030, and cost-effectiveness with and without uncertainty. Means are given along with 95% prediction intervals (PIs). YLL: years of life lost (to fatal disease), YLD: years of life lost to disability, DALYs: disability-adjusted life-years, PS: passive screening, AS: active screening, VC: vector control, ICER: incremental cost-effectiveness ratio, WTP: willingness to pay (USD per DALY averted), EoT: elimination of transmission.

| | Stop 2026 | Max AS (d2d) | Mean AS + VC | Mean AS | Reduced AS |
|---|---|---|---|---|---|
| **Health effects** | | | | | |
| Reported cases | 0 (0-0) | 0 (0-0) | 0 (0-0) | 0 (0-0) | 0 (0-0) |
| Deaths undetected | 0 (0-0) | 0 (0-0) | 0 (0-0) | 0 (0-0) | 0 (0-0) |
| Cases total | 0 (0-0) | 0 (0-0) | 0 (0-0) | 0 (0-0) | 0 (0-0) |
| Deaths detected[a] | 0 (0-0) | 0 (0-0) | 0 (0-0) | 0 (0-0) | 0 (0-0) |
| YLD | 0 (0-0) | 0 (0-0) | 0 (0-0) | 0 (0-0) | 0 (0-0) |
| YLL | 0 (0-6) | 0 (0-6) | 0 (0-5) | 0 (0-6) | 0 (0-6) |
| DALYs | 0.414 (<0.001-5.898) | 0.339 (<0.001-5.773) | 0.375 (<0.001-4.725) | 0.393 (<0.001-5.874) | 0.414 (<0.001-5.876) |
| **Costs, in US$** | | | | | |
| AS costs | 0 (0-17) | 77,804 (66,622-84,927) | 6,747 (5,926-7,534) | 6,750 (5,926-7,547) | 3,377 (2,962-3,774) |
| PS costs | 28,462 (24,231-32,579) | 28,482 (24,231-32,586) | 28,464 (24,231-32,581) | 28,467 (24,231-32,581) | 28,464 (24,231-32,579) |
| VC costs | 0 (0-1,104) | 0 (0-767) | 110,878 (62,597-170,356) | 0 (0-796) | 0 (0-803) |
| Treatment costs | 0 (0-41) | 0 (0-41) | 0 (0-31) | 0 (0-41) | 0 (0-41) |
| Costs total | 29,623 (24,236-32,806) | 107,094 (93,680-113,852) | 146,120 (97,027-205,804) | 36,055 (30,761-39,468) | 32,684 (27,517-36,151) |
| **EoT** | | | | | |
| Year of EoT | 2017 (2014-2024) | 2017 (2014-2024) | 2017 (2014-2024) | 2017 (2014-2024) | 2017 (2014-2024) |
| Prob EoT 2030 | 0.99 | 0.99 | >0.99 | 0.99 | 0.99 |
| **Cost-effectiveness without uncertainty (discounted)[b]** | | | | | |
| DALYs averted | 0.000 | 0.066 | 0.549 | 0.006 | 0.005 |
| Costs averted | 0 | 77,008 | 116,332 | 6,431 | 3,065 |
| ICER | Min Cost | Weakly Dominated | 211,905 | Weakly Dominated | Weakly Dominated |
| **Cost-effectiveness with uncertainty, conditional on WTP[c].** | | | | | |
| WTP: $0 | 1(p) | 0 | 0 | 0 | 0 |
| WTP: $500 | 1(p) | 0 | 0 | 0 | 0 |
| WTP: $1000 | 1(p) | 0 | 0 | 0 | 0 |
| WTP: $2000 | 0.99(p) | 0 | 0 | 0 | 0 |
| WTP: $3000 | 0.99(p) | 0 | 0 | 0 | 0 |

[a] Detected deaths are those that occur due to treatment failure of loss-to-follow-up.

[b] Cost-effectiveness results are given for discounted DALYs and costs as per convention

[c] (p) is the preferred strategy; the strategy with the highest mean net monetary benefits

Table S24: Results in Koboko. Summary of effects, costs, elimination of transmission (EoT) by 2030, and cost-effectiveness with and without uncertainty. Means are given along with 95% prediction intervals (PIs). YLL: years of life lost (to fatal disease), YLD: years of life lost to disability, DALYs: disability-adjusted life-years, PS: passive screening, AS: active screening, VC: vector control, ICER: incremental cost-effectiveness ratio, WTP: willingness to pay (USD per DALY averted), EoT: elimination of transmission.

| | Stop 2026 | Max AS (d2d) | Mean AS + VC | Mean AS | Reduced AS |
|---|---|---|---|---|---|
| **Health effects** | | | | | |
| Reported cases | 0 (0-0) | 0 (0-0) | 0 (0-0) | 0 (0-0) | 0 (0-0) |
| Deaths undetected | 0 (0-0) | 0 (0-0) | 0 (0-0) | 0 (0-0) | 0 (0-0) |
| Cases total | 0 (0-0) | 0 (0-0) | 0 (0-0) | 0 (0-0) | 0 (0-0) |
| Deaths detected[a] | 0 (0-0) | 0 (0-0) | 0 (0-0) | 0 (0-0) | 0 (0-0) |
| YLD | 0 (0-0) | 0 (0-0) | 0 (0-0) | 0 (0-0) | 0 (0-0) |
| YLL | 0 (0-0) | 0 (0-0) | 0 (0-0) | 0 (0-0) | 0 (0-0) |
| DALYs | <0.001 (<0.001-0.342) | <0.001 (<0.001-0.342) | <0.001 (<0.001-0.215) | <0.001 (<0.001-0.347) | <0.001 (<0.001-0.347) |
| **Costs, in US$** | | | | | |
| AS costs | 0 (0-2) | 87,214 (77,559-97,653) | 10,366 (9,243-11,638) | 10,370 (9,243-11,638) | 5,185 (4,622-5,819) |
| PS costs | 26,242 (22,645-30,061) | 26,244 (22,645-30,065) | 26,241 (22,645-30,061) | 26,243 (22,645-30,061) | 26,243 (22,645-30,061) |
| VC costs | 0 (0-54) | 0 (0-14) | 47,676 (27,747-73,177) | 0 (0-14) | 0 (0-14) |
| Treatment costs | 0 (0-4) | 0 (0-4) | 0 (0-3) | 0 (0-4) | 0 (0-4) |
| Costs total | 26,302 (22,645-30,068) | 113,477 (103,036-124,471) | 84,286 (63,717-110,120) | 36,631 (32,755-40,576) | 31,446 (27,743-35,272) |
| **EoT** | | | | | |
| Year of EoT | 2015 (2013-2018) | 2015 (2013-2018) | 2015 (2013-2018) | 2015 (2013-2018) | 2015 (2013-2018) |
| Prob EoT 2030 | >0.99 | >0.99 | >0.99 | >0.99 | >0.99 |
| **Cost-effectiveness without uncertainty (discounted)[b]** | | | | | |
| DALYs averted | 0.000 | 0.000 | 0.055 | -0.002 | -0.002 |
| Costs averted | 0 | 87,118 | 57,965 | 10,327 | 5,144 |
| ICER | Min Cost | Dominated | 1,048,599 | Dominated | Dominated |
| **Cost-effectiveness with uncertainty, conditional on WTP[c].** | | | | | |
| WTP: $0 | 1(p) | 0 | 0 | 0 | 0 |
| WTP: $500 | 1(p) | 0 | 0 | 0 | 0 |
| WTP: $1000 | 1(p) | 0 | 0 | 0 | 0 |
| WTP: $2000 | 1(p) | 0 | 0 | 0 | 0 |
| WTP: $3000 | 1(p) | 0 | 0 | 0 | 0 |

[a] Detected deaths are those that occur due to treatment failure of loss-to-follow-up.

[b] Cost-effectiveness results are given for discounted DALYs and costs as per convention

[c] (p) is the preferred strategy; the strategy with the highest mean net monetary benefits

Table S25: Results in Maracha. Summary of effects, costs, elimination of transmission (EoT) by 2030, and cost-effectiveness with and without uncertainty. Means are given along with 95% prediction intervals (PIs). YLL: years of life lost (to fatal disease), YLD: years of life lost to disability, DALYs: disability-adjusted life-years, PS: passive screening, AS: active screening, VC: vector control, ICER: incremental cost-effectiveness ratio, WTP: willingness to pay (USD per DALY averted), EoT: elimination of transmission.

| | Stop 2026 | Max AS (d2d) | Mean AS + VC | Mean AS | Reduced AS |
|---|---|---|---|---|---|
| **Health effects** | | | | | |
| Reported cases | 0 (0-0) | 0 (0-0) | 0 (0-0) | 0 (0-0) | 0 (0-0) |
| Deaths undetected | 0 (0-0) | 0 (0-0) | 0 (0-0) | 0 (0-0) | 0 (0-0) |
| Cases total | 0 (0-0) | 0 (0-0) | 0 (0-0) | 0 (0-0) | 0 (0-0) |
| Deaths detected[a] | 0 (0-0) | 0 (0-0) | 0 (0-0) | 0 (0-0) | 0 (0-0) |
| YLD | 0 (0-0) | 0 (0-0) | 0 (0-0) | 0 (0-0) | 0 (0-0) |
| YLL | 0 (0-0) | 0 (0-0) | 0 (0-0) | 0 (0-0) | 0 (0-0) |
| DALYs | <0.001 (<0.001-0.025) | <0.001 (<0.001-0.021) | <0.001 (<0.001-0.016) | <0.001 (<0.001-0.030) | <0.001 (<0.001-0.030) |
| **Costs, in US$** | | | | | |
| AS costs | 0 (0-0) | 16,194 (14,492-18,187) | 1,713 (1,533-1,924) | 1,713 (1,533-1,924) | 855 (765-961) |
| PS costs | 23,702 (20,296-27,412) | 23,702 (20,296-27,412) | 23,702 (20,296-27,412) | 23,702 (20,296-27,412) | 23,702 (20,296-27,412) |
| VC costs | 0 (0-18) | 0 (0-2) | 31,591 (18,415-48,322) | 0 (0-2) | 0 (0-2) |
| Treatment costs | 0 (0-1) | 0 (0-1) | 0 (0-0) | 0 (0-1) | 0 (0-1) |
| Costs total | 23,721 (20,296-27,412) | 39,899 (36,001-44,071) | 57,006 (43,267-74,145) | 25,418 (22,009-29,118) | 24,560 (21,150-28,267) |
| **EoT** | | | | | |
| Year of EoT | 2012 (2010-2015) | 2012 (2010-2015) | 2012 (2010-2015) | 2012 (2010-2015) | 2012 (2010-2015) |
| Prob EoT 2030 | >0.99 | >0.99 | >0.99 | >0.99 | >0.99 |
| **Cost-effectiveness without uncertainty (discounted)[b]** | | | | | |
| DALYs averted | 0.000 | 0.003 | 0.005 | -0.002 | -0.002 |
| Costs averted | 0 | 16,177 | 33,282 | 1,698 | 841 |
| ICER | Min Cost | 5,646,881 | 7,223,158 | Dominated | Dominated |
| **Cost-effectiveness with uncertainty, conditional on WTP[c].** | | | | | |
| WTP: $0 | 1(p) | 0 | 0 | 0 | 0 |
| WTP: $500 | 1(p) | 0 | 0 | 0 | 0 |
| WTP: $1000 | 1(p) | 0 | 0 | 0 | 0 |
| WTP: $2000 | 1(p) | 0 | 0 | 0 | 0 |
| WTP: $3000 | 1(p) | 0 | 0 | 0 | 0 |

[a] Detected deaths are those that occur due to treatment failure of loss-to-follow-up.

[b] Cost-effectiveness results are given for discounted DALYs and costs as per convention

[c] (p) is the preferred strategy; the strategy with the highest mean net monetary benefits

Table S26: Results in Moyo. Summary of effects, costs, elimination of transmission (EoT) by 2030, and cost-effectiveness with and without uncertainty. Means are given along with 95% prediction intervals (PIs). YLL: years of life lost (to fatal disease), YLD: years of life lost to disability, DALYs: disability-adjusted life-years, PS: passive screening, AS: active screening, VC: vector control, ICER: incremental cost-effectiveness ratio, WTP: willingness to pay (USD per DALY averted), EoT: elimination of transmission.

| | Stop 2026 | Max AS (d2d) | Mean AS + VC | Mean AS | Reduced AS |
|---|---|---|---|---|---|
| **Health effects** | | | | | |
| Reported cases | 0 (0-0) | 0 (0-0) | 0 (0-0) | 0 (0-0) | 0 (0-0) |
| Deaths undetected | 0 (0-0) | 0 (0-0) | 0 (0-0) | 0 (0-0) | 0 (0-0) |
| Cases total | 0 (0-0) | 0 (0-0) | 0 (0-0) | 0 (0-0) | 0 (0-0) |
| Deaths detected[a] | 0 (0-0) | 0 (0-0) | 0 (0-0) | 0 (0-0) | 0 (0-0) |
| YLD | 0 (0-0) | 0 (0-0) | 0 (0-0) | 0 (0-0) | 0 (0-0) |
| YLL | 0 (0-0) | 0 (0-0) | 0 (0-0) | 0 (0-0) | 0 (0-0) |
| DALYs | <0.001 (<0.001-0.143) | <0.001 (<0.001-0.136) | <0.001 (<0.001-0.187) | <0.001 (<0.001-0.138) | <0.001 (<0.001-0.143) |
| **Costs, in US$** | | | | | |
| AS costs | 0 (0-1) | 99,111 (88,626-111,480) | 10,132 (9,065-11,403) | 10,132 (9,065-11,403) | 5,065 (4,531-5,700) |
| PS costs | 28,229 (24,157-32,663) | 28,227 (24,156-32,660) | 28,229 (24,157-32,663) | 28,229 (24,157-32,663) | 28,229 (24,157-32,663) |
| VC costs | 0 (0-52) | 0 (0-23) | 96,662 (56,178-147,982) | 0 (0-23) | 0 (0-23) |
| Treatment costs | 0 (0-1) | 0 (0-1) | 0 (0-1) | 0 (0-1) | 0 (0-1) |
| Costs total | 28,283 (24,157-32,667) | 127,362 (115,879-140,229) | 135,024 (94,136-186,765) | 38,385 (34,120-42,901) | 33,318 (29,199-37,748) |
| **EoT** | | | | | |
| Year of EoT | 2016 (2014-2018) | 2016 (2014-2018) | 2016 (2014-2018) | 2016 (2014-2018) | 2016 (2014-2018) |
| Prob EoT 2030 | >0.99 | >0.99 | >0.99 | >0.99 | >0.99 |
| **Cost-effectiveness without uncertainty (discounted)[b]** | | | | | |
| DALYs averted | 0.000 | 0.004 | -0.013 | 0.003 | 0.000 |
| Costs averted | 0 | 99,059 | 106,731 | 10,103 | 5,036 |
| ICER | Min Cost | 62,588,505 | Dominated | 3,814,072 | Dominated |
| **Cost-effectiveness with uncertainty, conditional on WTP[c].** | | | | | |
| WTP: $0 | 1(p) | 0 | 0 | 0 | 0 |
| WTP: $500 | 1(p) | 0 | 0 | 0 | 0 |
| WTP: $1000 | 1(p) | 0 | 0 | 0 | 0 |
| WTP: $2000 | 1(p) | 0 | 0 | 0 | 0 |
| WTP: $3000 | 1(p) | 0 | 0 | 0 | 0 |

[a] Detected deaths are those that occur due to treatment failure of loss-to-follow-up.

[b] Cost-effectiveness results are given for discounted DALYs and costs as per convention

[c] (p) is the preferred strategy; the strategy with the highest mean net monetary benefits

Table S27: Results in Yumbe. Summary of effects, costs, elimination of transmission (EoT) by 2030, and cost-effectiveness with and without uncertainty. Means are given along with 95% prediction intervals (PIs). YLL: years of life lost (to fatal disease), YLD: years of life lost to disability, DALYs: disability-adjusted life-years, PS: passive screening, AS: active screening, VC: vector control, ICER: incremental cost-effectiveness ratio, WTP: willingness to pay (USD per DALY averted), EoT: elimination of transmission.

| | Stop 2026 | Max AS (d2d) | Mean AS + VC | Mean AS | Reduced AS |
|---|---|---|---|---|---|
| **Health effects** | | | | | |
| Reported cases | 0 (0-0) | 0 (0-0) | 0 (0-0) | 0 (0-0) | 0 (0-0) |
| Deaths undetected | 0 (0-0) | 0 (0-0) | 0 (0-0) | 0 (0-0) | 0 (0-0) |
| Cases total | 0 (0-0) | 0 (0-0) | 0 (0-0) | 0 (0-0) | 0 (0-0) |
| Deaths detected[a] | 0 (0-0) | 0 (0-0) | 0 (0-0) | 0 (0-0) | 0 (0-0) |
| YLD | 0 (0-0) | 0 (0-0) | 0 (0-0) | 0 (0-0) | 0 (0-0) |
| YLL | 0 (0-2) | 0 (0-2) | 0 (0-1) | 0 (0-2) | 0 (0-2) |
| DALYs | <0.001 (<0.001-2.206) | <0.001 (<0.001-2.227) | <0.001 (<0.001-1.394) | <0.001 (<0.001-2.236) | <0.001 (<0.001-2.236) |
| **Costs, in US$** | | | | | |
| AS costs | 0 (0-10) | 65,154 (56,557-71,673) | 1,886 (1,662-2,104) | 1,889 (1,662-2,106) | 949 (833-1,056) |
| PS costs | 40,281 (34,407-46,427) | 40,284 (34,407-46,433) | 40,259 (34,407-46,389) | 40,281 (34,407-46,427) | 40,281 (34,407-46,427) |
| VC costs | 0 (0-661) | 0 (0-343) | 88,520 (50,573-136,630) | 0 (0-351) | 0 (0-351) |
| Treatment costs | 0 (0-35) | 0 (0-38) | 0 (0-27) | 0 (0-37) | 0 (0-37) |
| Costs total | 40,986 (34,407-46,482) | 105,818 (94,134-113,730) | 130,692 (91,838-178,662) | 42,558 (36,238-48,345) | 41,618 (35,312-47,414) |
| **EoT** | | | | | |
| Year of EoT | 2017 (2015-2021) | 2017 (2015-2021) | 2017 (2015-2021) | 2017 (2015-2021) | 2017 (2015-2021) |
| Prob EoT 2030 | >0.99 | >0.99 | >0.99 | >0.99 | >0.99 |
| **Cost-effectiveness without uncertainty (discounted)[b]** | | | | | |
| DALYs averted | 0.000 | -0.009 | 0.362 | -0.014 | -0.014 |
| Costs averted | 0 | 64,611 | 89,601 | 1,591 | 652 |
| ICER | Min Cost | Dominated | 247,333 | Dominated | Dominated |
| **Cost-effectiveness with uncertainty, conditional on WTP[c].** | | | | | |
| WTP: $0 | 1(p) | 0 | 0 | 0 | 0 |
| WTP: $500 | 1(p) | 0 | 0 | 0 | 0 |
| WTP: $1000 | 1(p) | 0 | 0 | 0 | 0 |
| WTP: $2000 | 1(p) | 0 | 0 | 0 | 0 |
| WTP: $3000 | 1(p) | 0 | 0 | 0 | 0 |

[a] Detected deaths are those that occur due to treatment failure of loss-to-follow-up.

[b] Cost-effectiveness results are given for discounted DALYs and costs as per convention

[c] (p) is the preferred strategy; the strategy with the highest mean net monetary benefits

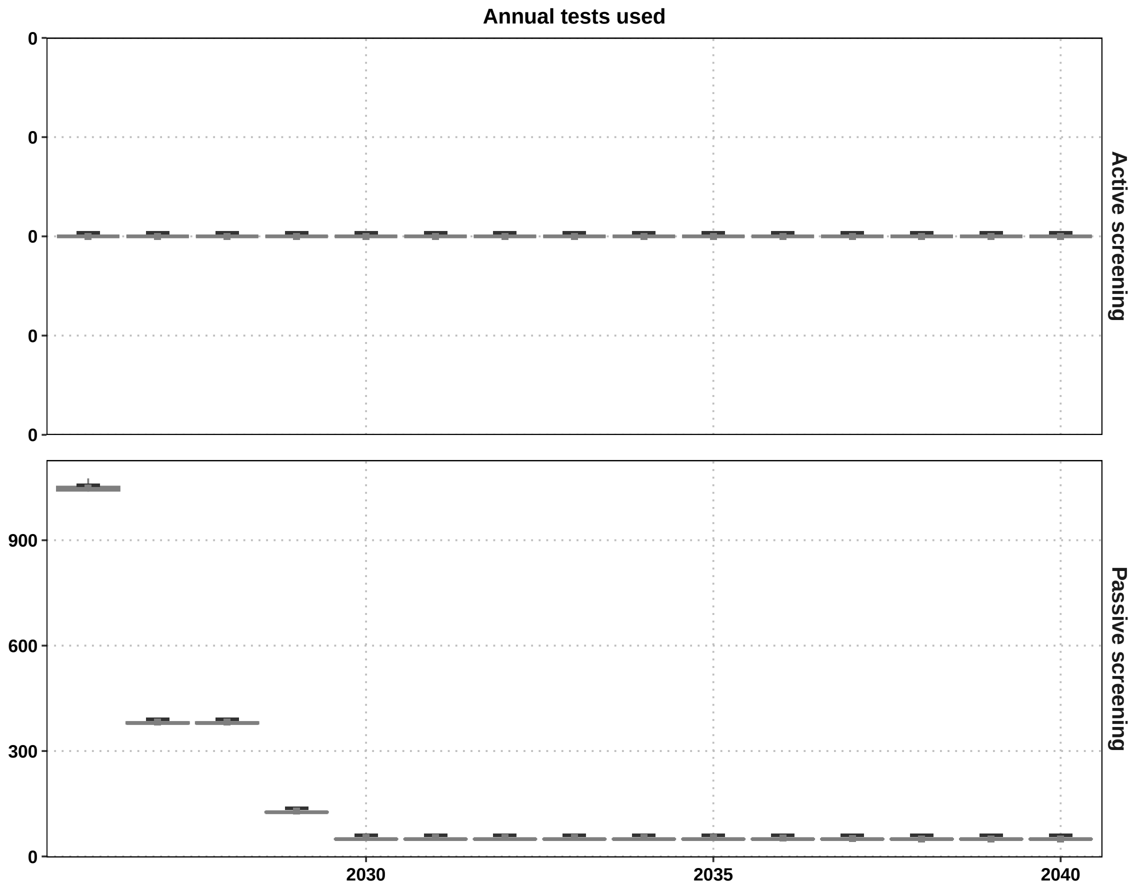

Figure S34: Country-wide resources consumed and needed by year if the Status Quo is maintained or if the Minimum Cost is maintained. VC targets and treatments are not displayed because they are 0 used in $\approx$ 99% iterations.

#### S13 Additional results from scenario analysis on cross-boundary importation

##### S13.1 Tsetse bounceback

Figures S35 show the district-level tsetse dynamics (under VC and also the bounce back after VC stops) for seven districts.

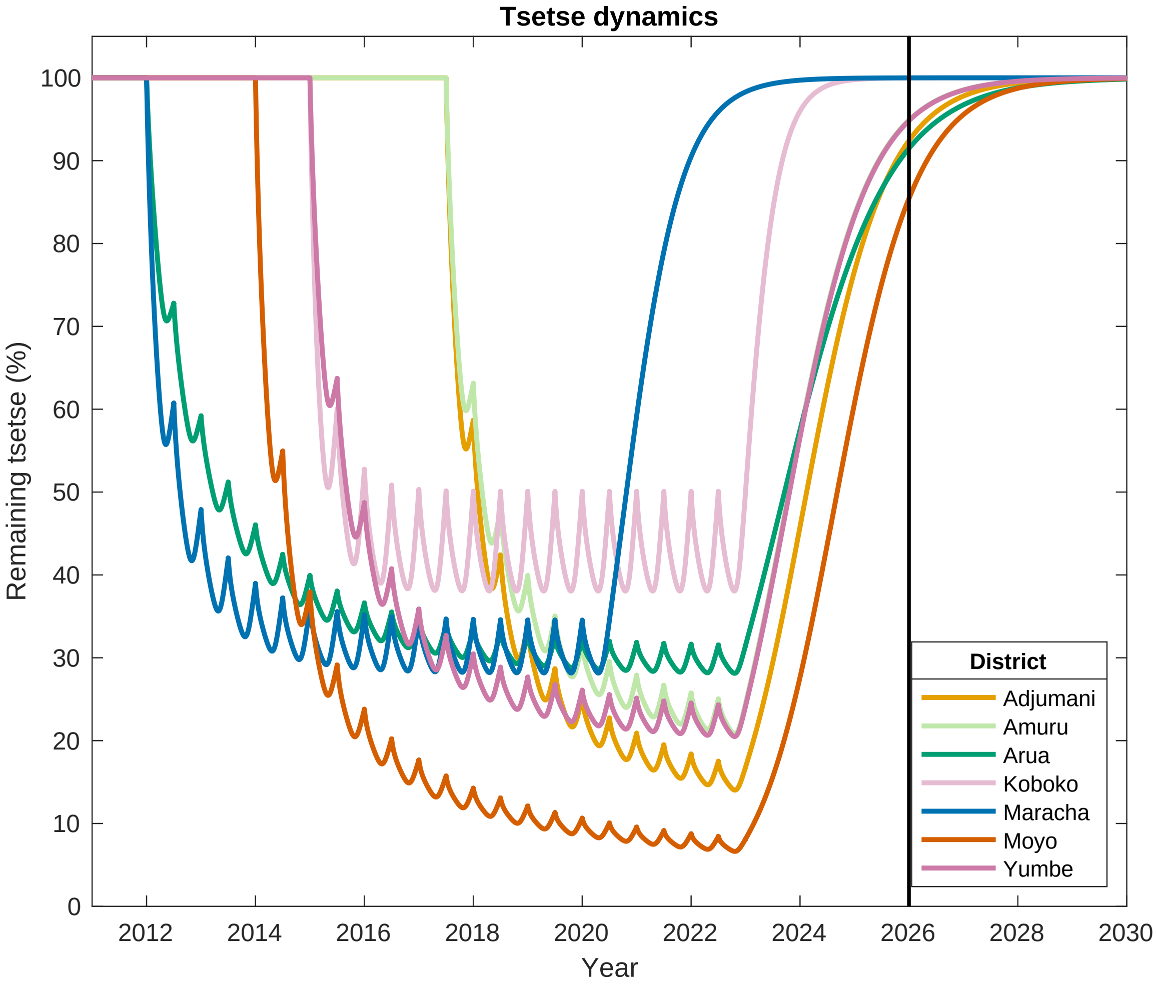

Figure S35: **Tsetse dynamics in seven districts.**

[S26] Uganda Bureau of Statistics. The 2002 Uganda Population and Housing Census, Gender and Special Interest Groups. Kampala, Uganda: Uganda Bureau of Statistics; 2006.

[S27] Brinkhoff, Thomas. City Population; 2021. [Online; accessed 12-January-2022]. http://www.citypopulation.de.

[S28] Uganda Bureau of Statistics. The National Population and Housing Census 2014 – Main Report. Kampala, Uganda: Uganda Bureau of Statistics; 2016.

[S29] Checchi F, Diap G, Karunakara U. Human African Trypanosomiasis, Update on progress and future perspectives. NEGDIS report. 2006;p. 7. Available from: https://msfaccess.org/sites/default/files/MSF_assets/NegDis/Docs/NEGDIS_report_HAT_Update_ENG_2006.pdf.

[S30] Wamboga C, Matovu E, Bessell PR, Picado A, BiÃ©ler S, Ndung'u JM. Enhanced passive screening and diagnosis for *gambiense* human African trypanosomiasis in north-western Uganda – Moving towards elimination. PLoS ONE. 2017 10;12(10):1–19. Available from: https://doi.org/10.1371/journal.pone.0186429.

[S31] The World Bank. Data:Democratic Republic of Congo; 2015. Accessed: 2015. Available from: https://data.worldbank.org/country/congo-dem-rep?view=chart.

[S32] Rogers DJ. A general model for the African trypanosomiases. Parasitology. 1988;97:193–212.

[S33] Checchi F, Filipe JAN, Barrett MP, Chandramohan D. The natural progression of *Gambiense* sleeping sickness: What is the evidence? PLoS Neglected Tropical Diseases. 2008 Dec;2(12):e303.

[S34] Checchi F, Funk S, Chandramohan D, Haydon DT, Chappuis F. Updated estimate of the duration of the meningo-encephalitic stage in *gambiense* human African trypanosomiasis. BMC Res Notes. 2015;8(1):292.

[S35] Mpanya A, Hendrickx D, Vuna M, Kanyinda A, Lumbala C, Tshilombo V, et al. Should I get screened for sleeping sickness? A qualitative study in Kasai province, Democratic Republic of Congo. PLoS Neglected Tropical Diseases. 2012 Jan;6(1):e1467.

[S36] Checchi F, Chappuis F, Karunakara U, Priotto G, Chandramohan D. Accuracy of Five Algorithms to Diagnose *Gambiense* Human African Trypanosomiasis. PLoS Neglected Tropical Diseases. 2011 Jul;5(7):e1233–15.

[S37] Hargrove J. Extinction probabilities and times to extinction for populations of tsetse flies Glossina spp.(Diptera: Glossinidae) subjected to various control measures. Bulletin of Entomological Research. 2005;95(1):13–21.

[S38] Davis S, Aksoy S, Galvani AP. A global sensitivity analysis for African sleeping sickness. Parasitology. 2010;138(04):516–526.

[S39] Ravel S, Grebaut P, Cuisance D, Cuny G. Monitoring the developmental status of *Trypanosoma brucei gambiense* in the tsetse fly by means of PCR analysis of anal and saliva drops. Acta Tropica. 2003;88(2):161–165.

[S40] WHO Expert Committee on human African trypanosomiasis. Control and surveillance of human African trypanosomiasis: report of a WHO expert committee; 2013. Available from: https://www.who.int/trypanosomiasis_african/surveillance/en/.

[S41] Haines LR. Examining the tsetse teneral phenomenon and permissiveness to trypanosome infection. Frontiers in cellular and infection microbiology. 2013;3:84.

[S42] Clausen PH, Adeyemi I, Bauer B, Breloeer M, Salchow F, Staak C. Host preferences of tsetse (Diptera: Glossinidae) based on bloodmeal identifications. Medical and Veterinary Entomology. 1998 Mar;12(2):169–180.

[S43] Rock KS, Torr SJ, Lumbala C, Keeling MJ. Predicting the impact of intervention strategies for sleeping sickness in two high-endemicity health zones of the Democratic Republic of Congo. PLoS Neglected Tropical Diseases. 2017;11(1):e0005162.

[S44] Touloupou P, Alzahrani N, Neal P, Spencer SEF, McKinley TJ. Efficient Model Comparison Techniques for Models Requiring Large Scale Data Augmentation. Bayesian Anal. 2018 06;13(2):437–459. Available from: https://doi.org/10.1214/17-BA1057.

[S45] Hesterberg T. Weighted Average Importance Sampling and Defensive Mixture Distributions. Technometrics. 1995;37(2):185–194. Available from: https://www.jstor.org/stable/1269620.

[S46] Hope A, Mugenyi A, Esterhuizen J, Tirados I, Cunningham L, Garrod G, et al. Scaling up of tsetse control to eliminate Gambian sleeping sickness in northern Uganda. PLoS Neglected Tropical Diseases. 2022 06;16(6):1–23. Available from: https://doi.org/10.1371/journal.pntd.0010222.

[S47] Briggs A, Claxton K, Sculpher M. Decision Modelling for Health Economic Evaluation. 1st ed. Oxford, UK: Oxford University Press; 2006.

[S48] Jamonneau V, Camara O, Ilboudo H, Peylhard M, Koffi M, Sakande H, et al. Accuracy of Individual Rapid Tests for Serodiagnosis of Gambiense Sleeping Sickness in West Africa. PLoS Neglected Tropical Diseases. 2015;9(2):1–9.

[S49] Snijders R, Fukinsia A, Claeys Y, Mpanya A, Hasker E, Meheus F, et al. Cost of a new method of active screening for human African trypanosomiasis in the Democratic Republic of the Congo. PLoS Neglected Tropical Diseases. 2020;14(12):e0008832.

[S50] Snijders R, Fukinsia A, Claeys Y, Hasker E, Mpanya A, Miaka E, et al. Costs and outcomes of integrated human African trypanosomiasis surveillance system using rapid diagnostic tests, Democratic Republic of the Congo. Emerging Infectious Diseases. 2021;27(8):2144.

[S51] CATT/T.b.g. REAGENTS (KIT R500 tests) - ITM Webshop for in vitro diagnostics;. Available from: https://diagnostics.itg.be/product/catt-t-b-g-reagents-kit-r500-tests/.

[S52] Eperon G, Schmid C, Loutan L, Chappuis F. Clinical presentation and treatment outcome of sleeping sickness in Sudanese pre-school children. Acta Tropica. 2006 1;101(1):31–39. Available from: https://linkinghub.elsevier.com/retrieve/pii/S0001706X06002439.

[S53] Schmid C, Kuemmerle A, Blum J, Ghabri S, Kande V, Mutombo W, et al. In-Hospital Safety in Field Conditions of Nifurtimox Eflornithine Combination Therapy (NECT) for T. b. gambiense Sleeping Sickness. PLoS Neglected Tropical Diseases. 2012;6(11).

[S54] Priotto G, Fogg C, Balasegaram M, Erphas O, Louga A, Checchi F, et al. Three Drug Combinations for Late-Stage Trypanosoma brucei gambiense Sleeping Sickness: A Randomized Clinical Trial in Uganda. PLo S Clinical Trials. 2006;1(8):e39.

[S55] Priotto G, Kasparian S, Ngouama D, Ghorashian S, Arnold U, Ghabri S, et al. Nifurtimox-Eflornithine Combination Therapy for Second-Stage Trypanosoma brucei gambiense Sleeping Sickness: A Randomized Clinical Trial in Congo. Clinical Infectious Diseases. 2007;45(11):1435–1442.

[S56] Checchi F, Piola P, Ayikoru H, Thomas F, Legros D, Priotto G. Nifurtimox plus eflornithine for late-stage sleeping sickness in Uganda: A case series. PLoS Neglected Tropical Diseases. 2007;1(2):1–6.

[S57] Priotto G, Kasparian S, Mutombo W, Ngouama D, Ghorashian S, Arnold U, et al. Nifurtimox-eflornithine combination therapy for second-stage African Trypanosoma brucei gambiense trypanosomiasis: a multicentre, randomised, phase III, non-inferiority trial. The Lancet. 2009;374(9683):56–64. Available from: http://dx.doi.org/10.1016/S0140-6736(09)61117-X.

[S58] Ngoyi DM, Lejon V, Pyana P, Boelaert M, Ilunga M, Menten J, et al. How to Shorten Patient Follow-Up after Treatment for Trypanosoma brucei gambiense Sleeping Sickness. The Journal of Infectious Diseases. 2010;201(3):453–463.

[S59] Burri C, Yeramian PD, Allen JL, Merolle A, Serge KK, Mpanya A, et al. Efficacy, Safety, and Dose of Pafuramidine, a New Oral Drug for Treatment of First Stage Sleeping Sickness, in a Phase 2a Clinical Study and Phase 2b Randomized Clinical Studies. PLoS Neglected Tropical Diseases. 2016;10(2):1–18.

[S60] Pohlig G, Bernhard SC, Blum J, Burri C, Mpanya A, Lubaki JPF, et al. Efficacy and Safety of Pafuramidine versus Pentamidine Maleate for Treatment of First Stage Sleeping Sickness in a Randomized, Comparator-Controlled, International Phase 3 Clinical Trial. PLoS Neglected Tropical Diseases. 2016;10(2):1–17.

[S61] Kansiime F, Adibaku S, Wamboga C, Idi F, Kato CD, Yamuah L, et al. A multicentre, randomised, non-inferiority clinical trial comparing a nifurtimox-eflornithine combination to standard eflornithine monotherapy for late stage Trypanosoma brucei gambiense human African trypanosomiasis in Uganda. Parasites and Vectors. 2018;11(1):1–11.

[S62] Mesu VKBK, Kalonji WM, Bardonneau C, Mordt OV, Blesson S, Simon F, et al. Oral fexinidazole for late-stage African *Trypanosoma brucei gambiense* trypanosomiasis: a pivotal multicentre, randomised, non-inferiority trial. The Lancet. 2018;391(10116):144–154.

[S63] World Health Organization. WHO interim guidelines for the treatment of *gambiense* human African trypanosomiasis. World Health Organization; 2019.

[S64] Balasegaram M, Harris S, Checchi F, Hamel C, Karunakara U. Treatment outcomes and risk factors for relapse in patients with early-stage human African trypanosomiasis (HAT) in the Republic of the Congo. Bulletin of the World Health Organization. 2006;84(10):777–782.

[S65] Bastide S, Priotto G, Ecochard R, Etard JF. Effectiveness of short vs. long treatment schedules with pentamidine in first-stage HAT: a large field cohort study. Tropical Medicine & International Health. 2011;16(S1):68–69. Available from: https://onlinelibrary.wiley.com/toc/13653156/2011/16/s1.

[S66] Alirol E, Schrumpf D, Amici Heradi J, Riedel A, De Patoul C, Quere M, et al. Nifurtimox-eflornithine combination therapy for second-stage gambiense human African trypanosomiasis: Médecins Sans Frontières experience in the Democratic Republic of the Congo. Clinical Infectious Diseases. 2013;56(2):195–203.

[S67] Baltussen RM, Adam T, Tan-Torres Edejer T, Hutubessy RC, Acharya A, Evans DB, et al. Making choices in health: WHO guide to cost-effectiveness analysis; 2003.

[S68] Bertram MY, Stenberg K, Brindley C, Li J, Serje J, Watts R, et al. Disease control programme support costs: an update of WHO-CHOICE methodology, price databases and quantity assumptions. Cost Effectiveness and Resource Allocation. 2017 12;15(1):21. Available from: http://resource-allocation.biomedcentral.com/articles/10.1186/s12962-017-0083-6.

[S69] Stenberg K, Lauer JA, Gkountouras G, Fitzpatrick C, Stanciole A. Econometric estimation of WHO-CHOICE country-specific costs for inpatient and outpatient health service delivery. Cost Effectiveness and Resource Allocation. 2018;16(1):1–15. Available from: https://doi.org/10.1186/s12962-018-0095-x.

[S70] Keating J, Yukich JO, Sutherland CS, Woods G, Tediosi F. Human African trypanosomiasis prevention, treatment and control costs: A systematic review. Acta Tropica. 2015;150:4–13.

[S71] Simarro PP, Diarra A, Ruiz Postigo JA, Franco JR, Jannin JG. The human African trypanosomiasis control and surveillance programme of the World Health Organization 2000-2009: the way forward. PLoS Neglected tropical diseases. 2011 2;5(2):e1007.

[S72] Shaw AP, Tirados I, Mangwiro CT, Esterhuizen J, Lehane MJ, Torr SJ, et al. Costs of using "tiny targets" to control Glossina fuscipes fuscipes, a vector of gambiense sleeping sickness in Arua District of Uganda. PLoS Neglected Tropical Diseases. 2015;9(3):e0003624.

[S73] Bessell PR, Esterhuizen J, Lehane MJ, Longbottom J, Mugenyi A, Selby R, et al. Estimating the impact of Tiny Targets in reducing the incidence of Gambian sleeping sickness in the North-west Uganda focus. Parasites & Vectors. 2021;14:1–13.

[S74] Hasker E, Lutumba P, Chappuis F, Kande V, Potet J, De Weggheleire A, et al. Human African Trypanosomiasis in the Democratic Republic of the Congo: A Looming Emergency? PLoS Neglected Tropical Diseases. 2012;6(12):e1950.

[S75] United Nations Population Division. World Population Prospects 2019; 2019. Available from: https://population.un.org/wpp/.

[S76] GBD 2017 DALYs and HALE Collaborators. Global, regional, and national disability-adjusted life-years (DALYs) for 359 diseases and injuries and healthy life expectancy (HALE) for 195 countries and territories, 1990-2017: A systematic analysis for the Global Burden of Disease Study 2017. The Lancet. 2018;392(10159):1859–1922.

[S77] Murray C. Quantifying the burden of disease : the technical basis for disability-adjusted life years. Bulletin of the World Health Organization. 1994;72(3):429–445.

[S78] Claxton KP, Revill P, Sculpher M, Wilkinson T, Cairns J, Briggs A. The Gates Reference Case for Economic Evaluation. The Bill and Melinda Gates Foundation; 2014. April. Available from: https://pure.york.ac.uk/portal/en/publications/the-gates-reference-case-for-economic-evaluation(2663ebb2-e102-48c1-af2f-3a482a2eaae0)/export.html.

[S79] Rushby JA, Hanson K. Calculating and presenting disability adjusted life years (DALYs) in cost-effectiveness analysis. Health Policy and Planning. 2001 9;16(3):326–331. Available from: https://academic.oup.com/heapol/article-lookup/doi/10.1093/heapol/16.3.326.

[S80] World Health Organization. Report from the third WHO stakeholders meeting on elimination of gambiense human African trypanosomiasis elimination. Geneva, Switzerland: World Health Organization; 2020. Available from: https://www.who.int/trypanosomiasis_african/resources/9789240002296/en/.

[S81] World Health Organization. Report of the fourth WHO stakeholders meeting on gambiense and rhodesiense human African trypanosomiasis elimination. Geneva: World Health Organization; 2022.

[S82] World Health Organization. Report of the fifth WHO stakeholders meeting on gambiense and rhodesiense human African trypanosomiasis elimination, Geneva, 7–9 June 2023. Geneva: World Health Organization; 2024.

[S83] Fexinidazole for T.b. gambiense — DNDi;. Available from: https://dndi.org/research-development/portfolio/fexinidazole/.

[S84] Tirados I, Esterhuizen J, Kovacic V, Mangwiro TNC, Vale GA, Hastings I, et al. Tsetse Control and Gambian Sleeping Sickness; Implications for Control Strategy. PLoS Neglected Tropical Diseases. 2015;9(8):e0003822.
